# The spread of infectious diseases from a physics perspective

**DOI:** 10.1101/2022.06.01.22275842

**Authors:** J.H.V.J. Brabers

**Affiliations:** Formerly at van der Waals-Zeeman Institute (UvA), Max Planck Institut fuer Metall/Festkoerperforschung

## Abstract

This paper presents a theoretical investigation of the spread of infectious diseases (including Covid-19) in a population network. The central idea is that a population can actually be considered as a network of interlinked nodes. The nodes represent the members of the population, the edges between the nodes the social contacts linking 2 population members. Infections spread throughout the population along these network edges. The actual spread of infections is described within the framework of the SIR compartmental model. Special emphasis is laid on understanding and on the interpretation of phenomena in terms of concepts borrowed from condensed-matter and statistical physics. To obtain a mathematical framework that deals with the influence of the network structure and topology, the original SIR model by Kermack and McKendrick was augmented, leading to a system of differential equations that is in principle exact, but the solution of which appears to be intractable. Therefore, combined algebraic/numerical solutions are presented for simplified (approximative) cases that nevertheless capture the essentials of the effect of the network details on the spread of an infection. Solutions of this kind were successfully tested against the results of direct statistical simulations based on Monte-Carlo methods, indicating the appropriateness of the model. Expressions for the (basic) reproduction numbers in terms of the model parameters are presented, and justify some mild criticisms on the widely spread interpretation of reproduction numbers as being the number of secondary infections due to a single active infection. Throughout the entire paper, special attention is paid to the concept of herd-immunity, its nature and its definition. The model allows for obtaining an exact (algebraic) criterion for the most relevant form of herd-immunity to occur in unvaccinated populations. Analysis of the effects of vaccination leads to an even more general version of this criterion in terms of not only the model parameters but also the effectiveness of the vaccine(s) and the vaccination rate(s). This general criterion is also exact within the context of the SIR model. Furthermore it is shown that the onset of herd-immunity can be considered as a 2nd-order phase transition of the kind that is known from thermodynamics and statistical physics, thus offering a fundamentally new viewpoint on the phenomenon. The role of percolation is highlighted and extensively investigated. It is shown that the herd-immunity transition is actually related to a percolation transition, and marks therewith the transition from a regime where the cumulative infections grow into a large macroscopic cluster that spans a major part of the population, towards a regime were the cumulative infections only occur in smaller secondary clusters of limited size. It appears that percolation phenomena become particularly important in the case of (strict) lock-downs. It is also demonstrated how a system of differential equations can be obtained that accounts for the presence of such percolation phenomena. The analyses presented in this paper also provide insight in how various measures to prevent an epidemic spread of an infection work, how they can be optimised and what potentially deceptive issues have to be considered when such measures are either implemented or scaled down. Herd-immunity appears to be a particularly tricky concept in this respect. Phenomena such as a saturation of the cumulative infection number or a fade-out of the number of active infections may easily be mistaken for a stable case of herd-immunity setting in, whereas in reality such phenomena may be no more than an artefact of protective or contact-reducing measures taken, without any meaning for the vulnerability of a population at large under normal (social) conditions. On the other hand, the paper also highlights and explains the theoretical possibility of “smothering” an epidemic via very restrictive measures that prevent it from developing out of a limited number of initial seed-infections.

## 0. Introduction

The Covid-19 pandemic has spurred an enormous scientific interest in the spread of infective diseases and in the evolution op epidemic outbreaks of such diseases. For a long time these subjects had only appealed to a limited group of researchers, and the overall scientific output dealing with them had been somewhat on the modest side. Possibly responsible for this is (in part) the enormous progress in medicine, especially during the second half of 20th century, which has provided mankind with effective treatments and prophylactics against a wide variety of severe infections. Large-scale epidemic or even pandemic spread of dangerous pathogens, without a cure being available, was considered a thing of the past by many. The Covid-19 outbreak has shattered this wide-spread illusion in a most dramatic way.

Another reason for the somewhat limited interest in the subject of epidemic infection outbreaks is the nature of the subject itself, which, at a closer look, is one of (at least) substantial complexity. This complexity makes it a highly non-trivial exercise to capture even the most elementary features of the phenomenon in simple mathematical formulas that can be dealt with by algebraic methods alone. Even one of the earlier, and still frequently used, models for the spread of infectious diseases, known as the SIR model and proposed by Kermack and McKendrick [1], leads to a set of differential equations that does not allow for an algebraic solution except in the simplest of cases. However, it deals with some essential elements of an epidemic by parting the population in 3 categories of population members: susceptibles (S) that have not been infected but which are vulnerable to infection, active infections (I) spreading the infection via transmission to susceptible, and removed infections (R) representing those members that have been infected but who are no longer infectious (able to spread the infection). In its mathematical form introduced by Kermack and McKendrick, to be referred to as the *standard* SIR-model hereafter, the model is able to reproduce some remarkable features of epidemics like for instance the fact (often observed in real outbreaks) that no matter how easily transmitted the pathogen involved may be, a finite part of a population that falls victim to an epidemic will always remain uninfected (i.e. susceptible). Infection removal turns out to be responsible for this observation. Only when infections are not removed, and each active infection in a population will remain an active infection indefinitely, the entire population gets infected in the end. However, also the fact that not only the standard SIR model but also its extended versions require numerical techniques and, depending on the complexity of a particular model, quite the necessary computational power (CPU-time), is a reason to be considered for the fact that mathematical epidemiology is a field with quite some unexplored territories. Only during the last two decades or so, computational power previously only available from main-frame of super computers has become readily available to a wide community of researchers (and some problems in mathematical epidemiology simply *do* require that power).

Some characteristics of epidemic growth of infections are not dealt with by the standard SIR-model however. When we look closer at the concept of a population from an epidemiological perspective, it is clear that a population actually represents a network of population members (nodes in mathematical terms), with each member being either in contact or involved in some other kind of “interaction” with other members (nodes) in the population. The standard SIR-model does not account for this network structure. It is obvious however, that this network structure is likely to have an influence on the propagation of an infection through a population. In physical systems consisting of networks or showing network-like structures (such as crystal lattices, porous media, electric circuits etc.), effects related specifically to the network characteristics of the system are not only common but, actually, the rule. There is no reason to assume that population networks will make an exception in this respect, especially not in the context of infection propagation. In fact, the spread of an infection through a population lattice can be perceived as a physical process analogous to the flow of a liquid trough a porous medium, or to the (macroscopic) polarisation of spins on a lattice or network under the influence of the (microscopic) interactions between the individual spins (for which the Ising model, which we will encounter more than once throughout this paper, represents the simplest case [2]). The standard SIR model, however, treats the spread of infectious diseases in terms of an analogue of the so-called mean-field approaches used in physics to describe systems with collective interactions. The environment of the population members (nodes) of a specific type (S, I or R) is considered the same for all members of the type and equal to some average over all the members of that type. Local fluctuations and lattice effects are averaged out. This yields a fair approximation in some cases, but generally leads to quantitative and, possibly, even qualitative differences from the exact behaviour of an infection in a given population and for given parameters.

An important phenomenon directly related to physical systems with a network structure is percolation (for a good introduction to the subject see [3]). Percolation becomes relevant when larger numbers of nodes in a network are removed (either randomly or according to some spatial distribution function), or become “inert” in the sense that they can no longer pass-on an interaction of some kind. Examples are the removal of spins on an Ising lattice, the replacement of magnetic atoms or ions by non magnetic ones in real magnetic systems, or the removal of joints in a network of resistors. The essence of percolation is the formation of a single macroscopic cluster of “active” or non-removed nodes that spans throughout the entire population, having a size of the same order of magnitude as the size of the population. Such a cluster is formed when the number of removed or “inert” nodes is sufficiently low. In cases where there are no nodes removed at all, the cluster becomes identical with the population. When a large enough number of nodes is (randomly) removed however, the macroscopic cluster starts breaking up into many smaller isolated clusters, until at some critical value of the removal rate the macroscopic cluster vanishes completely, leaving only so-called secondary clusters of “microscopic” size. The latter is referred to as the percolation transition, which has all the characteristics of a real phase transition from the viewpoint of statistical physics, including universality, scaling laws and critical exponents. Percolation transitions have a huge impact on the properties of real systems. In magnetic systems for instance, they relate to a collapse of the magnetic order (magnetisation), whereas in resistor networks they are accompanied by a notable increase in the equivalent resistance of the network. It may be obvious that, by their very nature, percolation phenomena may also play a role in the spread of infectious diseases. Their is a conceptual similarity for instance between the (random) replacement of magnetic atoms in a solid by non-magnetic ones and (random) vaccination of susceptible members of a population network. Vaccination with a vaccine that provides full immunity against infection turns a susceptible member of a population into an “inert” member that will not only remain uninfected but, inherently, will also not be able to pass on an infection to another member. The relevance of percolation to the problem of epidemic growth of infections is clear therewith. Despite this, the role of percolation in an epidemiological context has not been investigated extensively. Of the first notable research efforts dealing with the issue (like for instance [4]) a significant part has been presented only quite recently. One of the reasons for this must be sought in the fact that it is notoriously difficult to capture the essence as well as the complexity of percolation phenomena in mathematical formulas: easy as they are to visualise, describing them in mathematical terms is an entirely different matter. Therefore, computer simulations are the most widely used tool for investigating percolation. Monte-Carlo techniques provide a very useful (and frequently deployed) method in this respect [5], since these techniques can simulate the actual process of (random) node-removal and, where appropriate, its evolution with time. They also allow for a comprehensive evaluation of results. However, a disadvantage of Monte-Carlo methods is that they require (*very*) substantial computational efforts for achieving meaningful results and, correspondingly, a lot of CPU time compared to many other computational exercises. Hence, it was only after (relatively) fast and powerful computing facilities had become readily available on a thus far unprecedented scale during the last few decades, that the possibility of actually using Monte-Carlo techniques became easily accessible to a wider scientific audience. It is therefore not unsurprising that, until very recently, the role of percolation in the epidemic spread of infections has attracted only a relatively modest level of attention by the scientific community.

Another issue with Monte-Carlo simulations is that the results they generate do not readily provide theoretical insight into the simulated phenomenon. They are experiments in their own right, carried out on a computer instead of in a laboratory, but nevertheless they often require further analysis in the same way data obtained in the real world need to be analysed before they make sense.

The first aim of this paper is to deal with the aforementioned shortcomings of the standard SIR-model, and to incorporate the network structure of the population into a more general model. Such a model should thus account for lattice correlations as well as percolation effects. The partition of the population in susceptibles, active infections and removed infections is maintained. Only a 4th partition is introduced as an option, consisting of vaccinated population members who are (partially) immune to infection. However, the purpose of the model is insight, not numbers^1^. It consist of a mathematical framework that incorporates the influence of the network structure through expansion of key physical quantities as a (Taylor) series in the number of cumulative infections. These series expansions replace their simplified (linear) counterparts in the differential equations of the standard SIR-model. This approach leads to formulas which are exact, but only in a strictly formal sense, since the coefficients in the series expansions cannot be calculated “ab initio” in the majority of the cases of interest. Therefore, the framework is less suitable for making *accurate* predictions on how an existing epidemic in the real world will develop (the results presented in this paper make clear however that such predictions are generally highly problematic anyway). Nevertheless, valuable insights can be obtained on the basis of the model, as well as (qualitative) rules of thumb that can be of use during efforts to bring an actual epidemic under control. The model is extensively tested against data provided by Monte-Carlo simulations for both vaccinated and unvaccinated populations.

Another phenomenon addressed extensively in this paper is herd-immunity. A concept first mentioned in the literature by Potter [6] (following his experience as a veterinarian during an outbreak of the bacterial infection Brucella Abortus among cattle), herd-immunity has been a slippery subject ever since. In fact, the concept of herd-immunity is not even well-defined [7]. Within the scientific community there are many definitions circulating. One of the most prominent ones defines herd-immunity as the stage in the evolution of an epidemic where the number of active infections has reached its peak, after which it is dropping down to eventually fade-out. However, the fact that the active infection rate drops does not imply an immediate return to normality. The transmission of the infection to susceptible population members will still go on for a while and new infections will continue to emerge. Another definition of herd-immunity, one that resonates more with intuition, is in a more literal sense and is obtained when we define herd-immunity as the stage in the evolution of an epidemic where the number of active infections has become almost negligible, leaving a population immune to new major (epidemic) outbreaks of the pathogen involved under all circumstances. It is obvious that these 2 definitions are conceptually related, and therefore probably also relate to the same underlying mechanisms, thus illustrating the confusion that surrounds the concept of herd-immunity. As to the underlying mechanisms there is some confusion as well. In the standard SIR-model, herd-immunity (no matter which definition is used of the 2 definitions mentioned) is a direct result of infection removal. However, the achievement of herd-immunity is often illustrated/explained in a pictorial way on the basis of a (partially immunised) population network in which (fully) immunised members block the routes of the infection towards susceptible members of the population, thus “screening” them from active infections. Obviously the mechanisms leading to herd-immunity are not entirely clear as well, or at least the subject of ambiguity. It does even occur that in the same paper results from the standard SIR-model are quoted (for instance to calculate the herd-immunity threshold), whereas an explanation of herd-immunity is given in terms of the aforementioned pictorial scheme (see [8] for example). An additional aim of this paper is therefore to bring some clarity in these issues. The inclusion, also presented in this paper, of the network-structure of populations and percolation effects into a generalised SIR-model seems to provide an ideal foundation for such an effort.

Another central aspect of mathematical epidemiology are reproduction numbers, among which the *basic* reproduction number *R*_0_ has a special status. Reproduction numbers are usually defined as the total number of secondary infections that a single active infection generates from a given moment in time onwards. The basic reproduction number *R*_0_ is a special case here, and represents the total number of secondary infections generated by a single active infection present at the start of an outbreak (*t* = 0). In practice, estimates for reproduction numbers are calculated in many different ways. However, a closer inspection shows that very often such calculations are in fact approximations based on a rather crude translation of the actual concept of reproduction numbers, as it is defined, into an algebraic expression in terms of the parameters of an underlying epidemiological model (such as the SIR-model). For instance, the expression for *R*_0_ on the basis of the standard SIR-model is very often written as *R*_0_ = *p*_*i*_*/p*_*r*_, where *p*_*i*_ and *p*_*r*_ are the central parameters of the SIR-model, namely the transmission rate (*p*_*i*_) and the rate of removal, or constant of removal (*p*_*r*_). The reasoning thereby is that *τ* = 1*/p*_*r*_ represents the average lifetime of an active infection (*p*_*r*_ has dimension 1/time) and that with a transmission rate *p*_*i*_ (which also has dimension 1/time) the total number of infections generated during the lifetime of an active infection present at *t* = 0 is simply given by *R*_0_ = *τp*_*i*_ = *p*_*i*_*/p*_*r*_. But what is entirely ignored here is that the number of susceptibles is *not* a constant during the lifetime of an active infection and may even vary substantially when *p*_*r*_ is small (i.e. *τ* is large) relative to *p*_*i*_. Since the (temporal) values of reproduction numbers are often considered as indicators for herd-immunity (for instance, the herd-immunity threshold obtained on the basis of the standard SIR-model is often expressed as 1 − 1*/R*_0_) a more detailed discussion about them seems inevitable in the context of this paper. The aim is to clarify how exact, non-approximative, expressions for reproduction numbers can be given on the basis of the generalised SIR-model presented here, as well as to see whether (and how) the network structure of a population has an effect on reproduction numbers generally, and what the precise relation is between reproduction numbers and herd-immunity.

This paper is primarily written with a readership consisting of physicists and (mathematical) epidemiologists in mind. In line with this objective, much emphasis is laid on analogies between epidemiological phenomena and phenomena in physics. Concepts and notions borrowed from statistical and condensed matter physics make their appearance regularly (though this may not be mentioned explicitly on every occasion). The issues dealt with are addressed therewith from a physics perspective, with the aim of providing a wider viewpoint on a the spread of infectious diseases, a topic that not only gained an enormous (instant) relevance when the Covid-19 pandemic broke out, but which will remain of importance in times still to come.

The general outline of the paper is as follows. Chapter 1 presents the generalised SIR-model that accounts for network-effects and local fluctuations. Chapter 2 deals with reproduction numbers and their interpretation. Semi-algebraic solution methods for 3 relevant approximative examples of the basic differential equations of the generalised SIR-model (presented in chapter 1) are discussed in chapter 3. In chapter 4 the generalised SIR-model is tested against to results of Monte-Carlo simulations of epidemic outbreaks in an SIR context. Chapter 5 deals with properties and consequences of the model, and with several important insights obtained from it that lay the basis for the more detailed discussion of herd-immunity presented in the next chapter. Chapter 6 is entirely devoted to herd-immunity. It addresses the general mechanisms responsible for herd-immunity, the definition of herd-immunity and the herd-immunity threshold. As a logical follow up of chapter 6, chapter 7 deals with vaccination and vaccine-acquired herd-immunity. Finally, chapter 8 gives an in-depth account of the role of percolation effects in the spread of infectious diseases and establishes an interesting link between herd-immunity transitions and phase transitions as they are known from statistical physics. It is also shown how the generalised SIR model presented in chapter 1 is able to account for these percolation effects.

## Data Availability

All data produced in the present work are contained in the manuscript

## Glossary of most common symbols

s_s_: fraction/rate of susceptible nodes/population members
s_i_: active-infection rate
s_r_: removed-infection rate
s: cumulative rate of infections
s_0_: initial value of active infection rate
s_0,b_: initial value of active infection rate in bulk the cluster
s_0,c_′: initial value of active infection rate in the secondary clusters
s_e_: final value (asymptotic) of cumulative infection rate
w_i_: transmission probability
p_i_: transmission rate/parameter/constant
p_r_: constant of removal
n_xy_: number of *xy* pairs (*x, y* = *s, i, r*)
ν: number of contacts to a single node/population member
⟨s_yx_⟩: average number of nodes of type *y* surrounding a node of type *x* (*y, x* = *s, i, r*)
f_cn_: contact-frequency: number of contacts made per node per unit time
f_cl_: contact-frequency: number of contacts made per link per unit time
f_cp_: contact-frequency: number if contacts made per unit time throughout entire population
n: number nodes/members in a population
n_0_: number of initial infections
n_e_: final value of the number cumulative infections
n_s_: number of susceptibles
n_i_: number of active infections
n_r_: number of removed infections
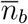: relative (total) size of bulk cluster(s)
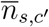: relative total size of secondary clusters
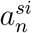: expansion coefficient for *n*th term in series expansion of ⟨*s*_*si*_⟩ (*n* = 0, 1, 2)
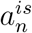: expansion coefficient for *n*th term in series expansion of ⟨*s*_*is*_⟩ (*n* = 0, 1, 2)
R: reproduction number
R_0_: basic reproduction number
s_1,2,3_: roots of 3rd order polynomial approximation of ⟨*s*_*si*_⟩
s_±_: roots of 2nd order polynomial approximation of ⟨*s*_*si*_⟩, or 3rd order of ⟨*s*_*is*_⟩
t: time
N: integer, scales the size of squares representing/enclosing a social bubble (size: 2*N* + 1 × 2*N* + 1)
ξ_0_: relative reduction of social-bubble size at *t* = 0
ξ_e_: relative reduction of social-bubble size for *t* → ∞
ϵ: effectiveness of vaccine
ξ_v_, x_v_: vaccination rate, relative reduction of social-bubble size as a result of vaccination
x_c_: percolation threshold
ξ: correlation length
S_c_, S_c_′: cluster size (number of nodes/members)
S_0_, S_b_: size of the bulk cluster (number of nodes/members)
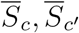: relative cluster size (number of nodes/members)
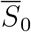: relative size of the bulk cluster (number of nodes/members)
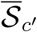: expectation value of relative total size of secondary clusters
A_c_: cluster area

## 1. SIR-model with network correlations and local fluctuations

We represent the population in which the epidemic is spreading as a network (or lattice) of nodes (lattice points) connected by links (lattice bonds) to other nodes in the network (see fig. 1). The nodes represent the individuals belonging to the population, links connecting 2 nodes the social interaction between the individuals represented by the nodes. As such, the multiple of links connecting a single node to other nodes in the network can be seen as the social network of the individual represented by the central node. It is along these links that the infection is transmitted and the epidemic spreads.

**Fig. 1.1:**
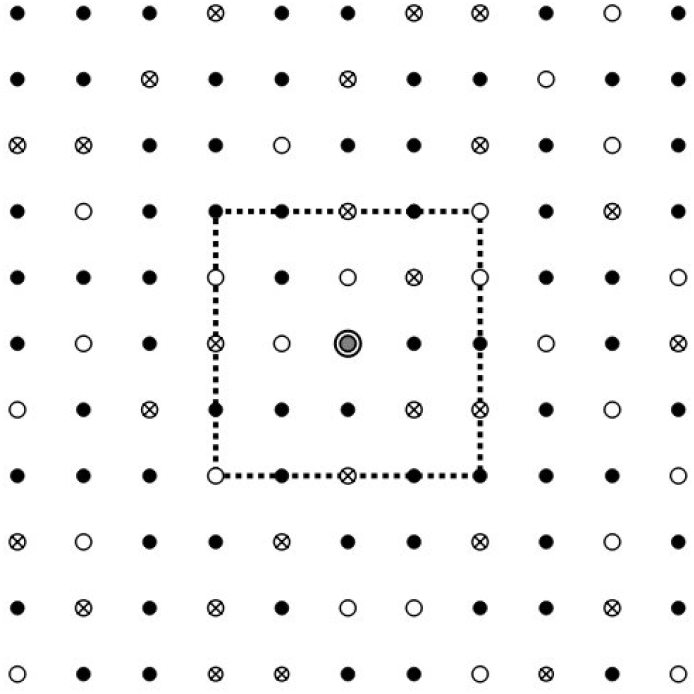
Schematic representation of a population network. There are 3 types of nodes: susceptibles (∘), active infections (•), removed infections (⊗). The dashed square symbolises the social network of the node in the center (central grey dot).

Even the analysis of relatively simple networks and lattices is, in general, a complicated matter however. In most cases, many typical phenomena that may take place in a network, such as cluster-formation and percolation-transitions, defy an exact (algebraic) treatment, and their full analysis requires numerical methods or even rigorous computer-simulations. The purpose of this section however, is to capture some of the essential features of the spread of an epidemic in a phenomenological algebraic model that offers not only the possibility to obtain semi-quantitative results but, above all, more insight into the mechanisms and phenomena involved.

Following the standard SIR-model [1], we assume 3 “types” of individuals or nodes: s) susceptible ones (uninfected but vulnerable to infection via social contacts), i) infectious ones (active, transmissible infections), and r) removed infections, that relate to individuals that have either recovered from an infection and acquired (indefinite) *full* immunity, or individuals that have succumbed to an infection (unlike in real life, there is no difference between these two possibilities from a strictly mathematical viewpoint). To avoid unnecessary complications we assume that each individual keeps contact with the same number *ν* of other individuals in the population. That is, each node is connected via links to an equal number of other nodes in the network.

As soon as the first active infections occur in the population (either by having been “imported” from outside the population or by any other feasible mechanism) and the epidemic starts spreading, the respective population-fractions *s*_*s*_, *s*_*i*_, *s*_*r*_ of respectively susceptible, infected and removed nodes/individuals start to evolve with time. However, it is easy to see that, by definition:

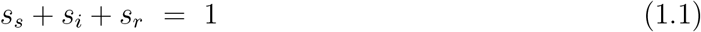

The standard SIR-model is entirely centred around these quantities *s*_*s*_, *s*_*i*_, *s*_*r*_ and is based on viewpoints similar to those underlying the mean-field descriptions of thermodynamic phase transitions. Local fluctuations in the environments (social networks) of the individual nodes are neglected (in fact even ignored). The environment of a node (i.e. the nodes linked to a particular node) is supposed to be homogeneous, with each node in the environment being of type s, i, or r with a probability given by *s*_*s*_, *s*_*i*_, *s*_*r*_ respectively.

With *p*_*i*_ representing the rate of transmission of infection (per active infection and per susceptible individual) and *p*_*r*_ the rate of infection-removal, the following coupled set of (non-) linear differential equations describes the time-evolution of the epidemic in the SIR-model:

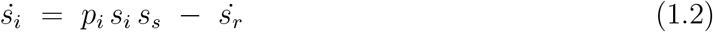

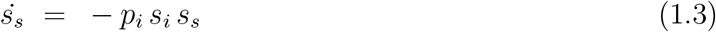

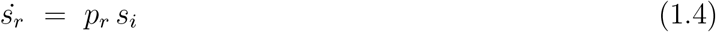

where the dotted symbols represent the time-derivatives. We now introduce the cumulative number of infections at a given time:

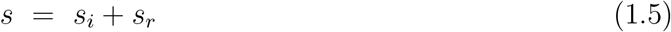

in terms of which *s*_*s*_ can be expressed (via (1.1)) as *s*_*s*_ = 1 − *s*, so that by also using eq. (1.4) we can rewrite eqs. (1.2) and (1.3) as:

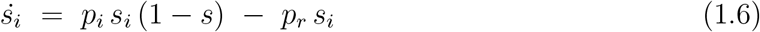

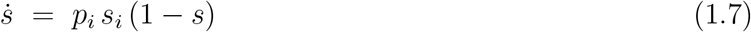

The rationale behind the term *p*_*i*_ *s*_*i*_ *s*_*s*_ = *p*_*i*_ *s*_*i*_ (1−*s*) is that a fraction *s*_*i*_ of the population is infected and that each individual contacted by an infected person is susceptible to transmission with *equal* probability *s*_*s*_ (the fraction of susceptible individuals among the total population). It is mainly in this particular Ansatz that the analogy between mean-field methods in the theory of thermodynamic phase transitions and the SIR-model is rooted (for a brief outline of mean-field methods see [2] for instance). However, it is well known that such an approach does not come without significant shortcomings, even in a qualitative sense. Not only the local fluctuations are ignored, but also the correlations that exist between probabilities of finding individual nodes linked to nodes of a certain type. Such correlations nearly always arise and depend on the particular geometry and topology of the lattice or network (the number of surrounding nodes to which each particular node is linked (*ν*) plays a crucial role in this respect for instance). This is particularly important in the context of percolation-phenomena, which can be seen as a useful paradigm for understanding group- or herd-immunity (as we will see later on).

To obtain an approach that, at least in a formal sense, takes account of local fluctuations and the above-mentioned correlations, we focus on the different type of *links* (or *pairs* of nodes) that can be identified in the network. We have links connecting an active infection (i) with a susceptible (s) node, links connecting an active infection with another active infection, links connecting a susceptible node with a removed infection (r) and so on. When the epidemic spreads, the total numbers of these links or pairs in the network (*n*_*xy*_, *x, y* = *s, i, r*) evolve with time until a stationary (equilibrium) state is reached (marking the end of the epidemic). From simple considerations (adopted from solid-state physics where they are applied to crystal-lattices), the following relations between the numbers of pairs of each type and the number *ν* of social contacts of an individual can be obtained:

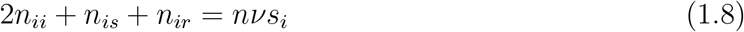

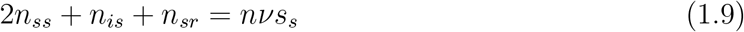

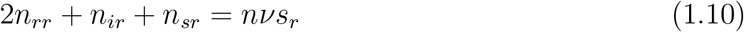

where *n* represents the total number of nodes in the population/network. The idea here is that each node is equally attributed to (divided among) the *ν* links connecting it to the other nodes in the network. As such, a link connecting a node of type *x* (*x* = *s, i, r*) to a node of type *y* (*y* = *s, i, r*) accounts for 1*/ν* of an x-type node and 1*/ν* of an y-type node (and when x=y for 2*/ν* of an x-type node). Summing over all the links (which is equivalent to summing over all the pairs of nodes connected via a (single) link) should yield the total number of s-, i- and r-nodes in the network.

We number the nodes of each particular type *x* = *s, i, r* from 1 to *ns*_*x*_. Let *ν*_*yx*_(*l*_*x*_) be the number of nodes of type *y* = *s, i, r* linked to the *l*_*x*_-th node of type x.

We introduce:

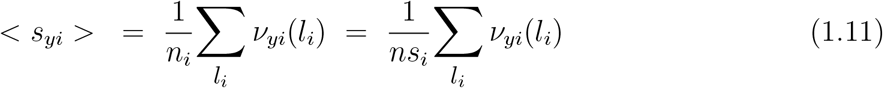

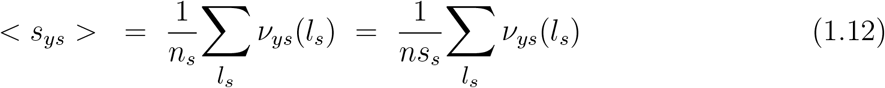

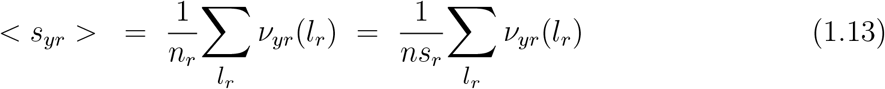

The < *s*_*yx*_ > represent the *average* number of *y*-type nodes linked to an *x*-type node (where the average is taken over *all* the nodes of *x*-type in the network). It is easy to verify that when *x* ≠ *y*, the numbers of *xy*-links or pairs in the network are directly related to the averages < *s*_*yx*_ > via:

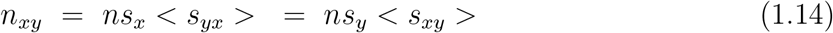

and when *y* = *x* via:

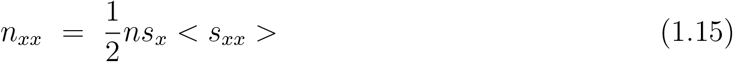

where the division by 2 corrects for double-counting *x*-nodes. By using these identities and dividing out the *ns*_*x*_, eqs. (1.8), (1.9) and (1.10) can be rewritten for those cases where *s*_*i*_; *s*_*s*_; *s*_*r*_ ≠ 0 as:

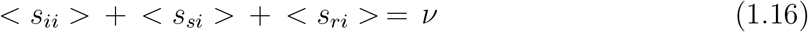

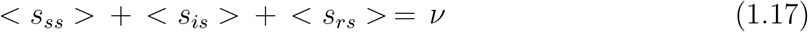

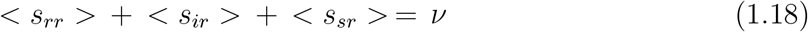

a result not too surprising in itself.

Transmission of infection may take place only upon contact between individuals with an active infection (i) and a susceptible person (s), i.e. between i-type and s-type nodes in the network (forming an i-s pair). The rate of transmission is therefore proportional to the number of i-s pairs *n*_*is*_ and can be expressed as:

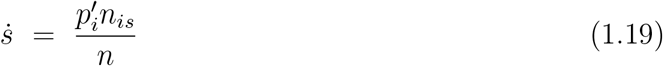

which may serve as a replacement for eq. (1.7), whereas the rate of change of the active infections can be written as:

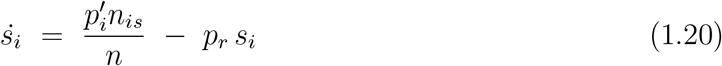

to replace eq. (1.6). The parameter 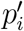 is the rate of transmission per i-s pair. Using (1.11) and (1.14) we can rewrite (1.19) as:

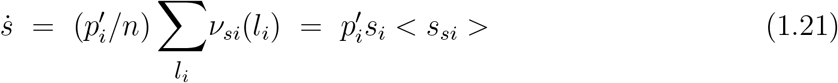

and (1.20) as:

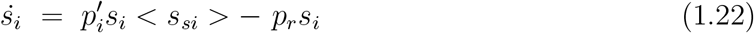

It is worth noticing that (1.21) and (1.22) are in fact *exact* results for infinitely large populations and finite *s*_*i*_ (and as such expected to apply also very well to finite yet sufficiently large populations). They constitute an exact generalisation of the standard SIR-model that accounts, at least in principle, for fluctuations and for correlations arising from to the typical network-structure of the population.

To demonstrate how the master-equations (1.6) and (1.7) of the standard SIR-model relate to their generalisation in the form of (1.21) and (1.22) it is useful to focus more closely on the parameters *p*_*i*_ and 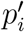 and to express them in terms of other relevant parameters. For that purpose we introduce the frequency *f*_*cn*_, which stands for the number of contacts made per *node* (or individual) per unit of time, as well as the transmission probability *w*_*i*_, which is the probability that the infection is passed on from one person to another upon contact. It is easy to see from its definition implied by (1.6) and (1.7) that:

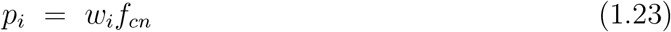

so that the normalised rate of new infections in the standard SIR-model (eq. (1.7)) can be reexpressed as:

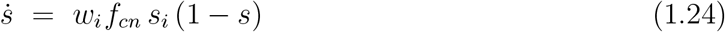

The total number of pairs (i.e. links) in the network is *n*_*p*_ = *νn/*2 (each node is connected via *ν* links, each link is shared by 2 nodes). Now, let *f*_*cl*_ be the number of contacts per unit time made *via a single link*.

The total number of contacts per unit time made throughout the entire *population* can now be written straightforwardly as:

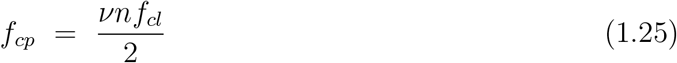

and, since it is easy to see that *f*_*cn*_ = *νf*_*cl*_, alternatively as:

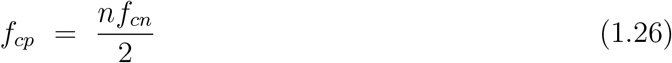

Of all the *n*_*p*_ = *νn/*2 pairs (i.e. links) in the network, a fraction:

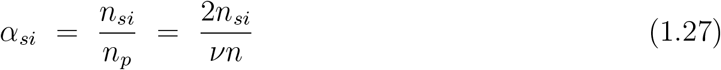

consists of *s* − *i* pairs. For the number of *s* − *i* contacts per unit time we get:

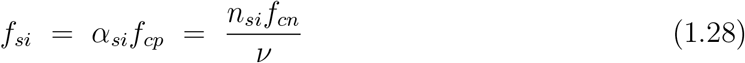

Using this result, the normalised rate of new infections in our generalisation of the SIR-model is now obtained as as:

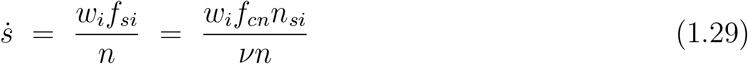

which, since *n*_*si*_*/n* = *s*_*i*_ <*s*_*si*_> (see (1.11)), can be rewritten as:

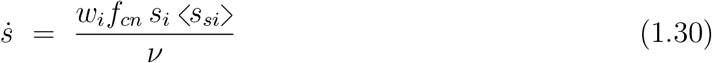

Comparison of (1.21) and (1.30) shows that we can identify 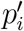 as 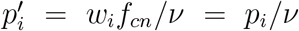 (which is in fact quite a logical result that also follows from the definition of *w*_*i*_ and *p*_*i*_). In addition we have from (1.16):

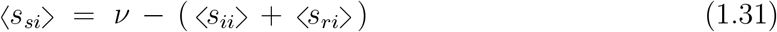

so that (1.30) can be reworked into:

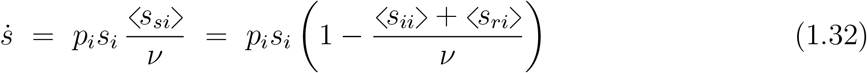

An alternative (but equivalent) expression for 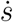 can be obtained by deploying the symmetry relation (see (1.14)):

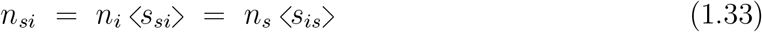

by which we can also write *n*_*si*_*/n* as:

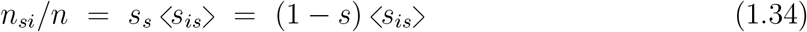

Substitution into (1.29) then yields (with *w*_*i*_*f*_*cn*_*/ν* = *p*_*i*_*/ν*):

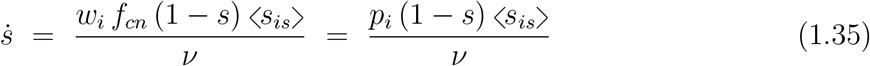

And with <*s*_*is*_> written as (see eq. (1.17)):

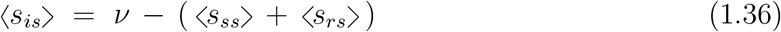

we thus obtain:

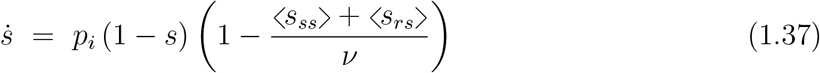

The differential equation relating *s*_*r*_ to *s*_*i*_ remains unchanged in the presence of correlations and is, as before, represented by (1.4). Comparison of (1.32) and eq. (1.7) thus shows that extending the SIR-model by including correlations arising from the typical network structure of the population implies (at least in a mathematical sense) in fact nothing more than replacing the factor (1 − *s*) in (1.7) by 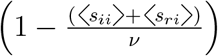, or the factor *s*_*i*_ by 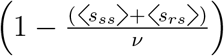. With 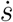 written as in either (1.32) or (1.37), we thus obtain 2 equivalent equations to replace (1.6), respectively given by:

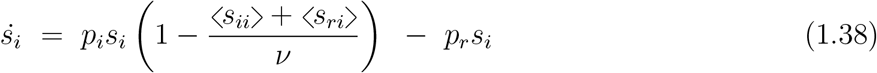

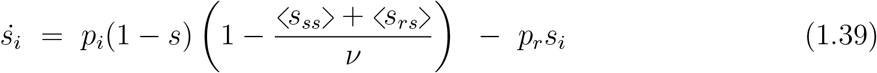

By arbitrarily combining one of the equations (1.32) and (1.37) with one of the equations (1.38) and (1.39), we obtain a system of 2 ordinary differential equations by which (in principle) the variation of *s* and *s*_*i*_ with time is defined (and indirectly, via (1.1) and (1.5), also the variation of *s*_*s*_ and *s*_*r*_). However, an actual solution of such a system requires the explicit algebraic form of either <*s*_*ii*_ > and <*s*_*ri*_> or <*s*_*ss*_> and <*s*_*rs*_> to be known. It is for that purpose that we seek an appropriate parameter in terms of which not only the *s*_*i*_, *s*_*s*_ and *s*_*r*_ but also the averages <*s*_*xy*_> (*x, y* = *i, s, r*) can be expressed. That is, we look for an independent quantity in terms of which the entire problem can be parametrised. Strictly speaking, time (*t*) meets that requirement, but is also an inappropriate/impossible choice since we actually want to solve *s*_*i*_, *s*_*s*_ and *s*_*r*_ for *t*. A proper choice however is *s*. Accounting for the cumulative number of infections, *s* = *s*(*t*) can only *increase* with time. As a result, *s*(*t*) is a bijective function of *t* (i.e. each *t* corresponds to a unique value of *s*). This implies that a parametrisation of individual quantities in terms of *only s* is possible and, moreover, entirely equivalent to a parametrisation of those quantities in terms of *t* (as such, *s* plays a role similar to that of the state-variables in thermodynamic systems).

However, the task of finding a representation of the <*s*_*xy*_> in terms of *s* is a tough problem bedevilled with difficulties that also arise in the analysis of Ising-problems (or lattice-problems in general). Ironically, the root cause of these difficulties is actually the same thing that we seek to incorporate into our present analysis, namely the lattice- or network correlations. They very often prevent an easy systematic enumeration of the states of the system that correspond to a specific value of a relevant state-variable (as a consequence, the exact solution to the 3D Ising-problem is still an open issue for instance). A pragmatic way out of these difficulties is to consider the exact <*s*_*xy*_> as series expansions in *s*:

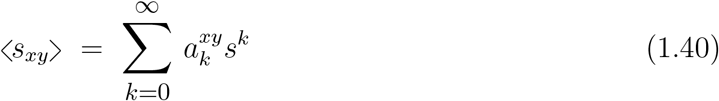

to be truncated at will in practice, thus providing us with approximations for the <*s*_*xy*_> in the form of finite-order polynomials in *s*.

We now assume a scenario where the initial number of active infections at the onset of an epidemic is finite but *almost negligible* at the scale of the total population, so that *s*_*i*_ ≈ 0 at *t* = 0. We furthermore assume that no infections were present before the start of the epidemic, so that there are no recovered and immune individuals at *t* = 0 and therefore *s*_*r*_(0) = 0 (*exactly*). For such a situation, we can easily identify the expansion parameters 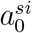 and 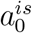 as 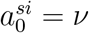 and 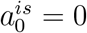, so that (see, respectively, (1.31) and (1.36)):

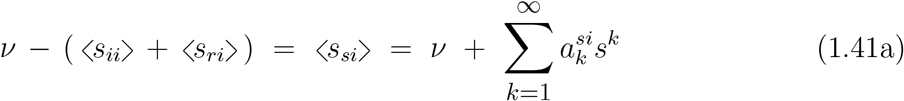

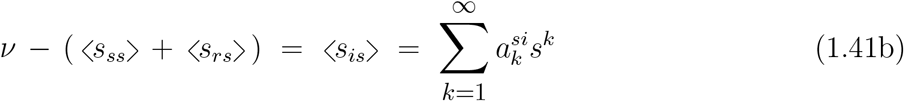

Substituting of (1.41a) for the terms in brackets respectively in (1.38) and (1.32) we get:

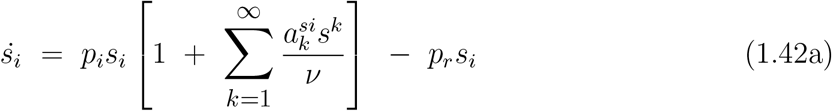

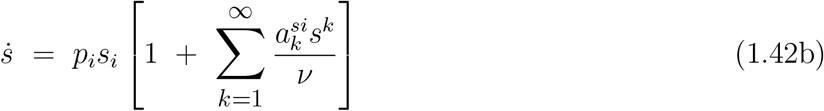

whereas substitution of (1.41b) for the terms in brackets in respectively (1.37) and (1.39) yields:

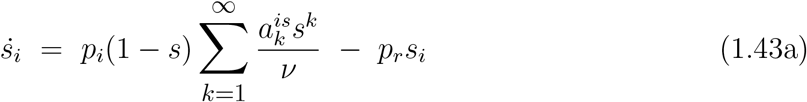

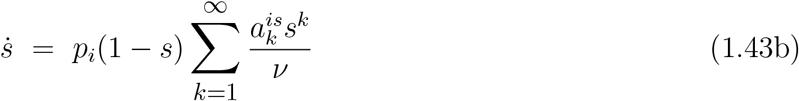

As to the series expansions in (1.42a,b) it should be emphasized that only cases where 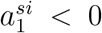 have relevance and “physical” meaning, since generally ⟨*s*_*si*_⟩ must decrease with increasing *s*. This to hold, also when *s* << 1, specifically requires that 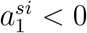.

After truncation of terms of order > 3 in the series expansions in (1.42a,b) we have:

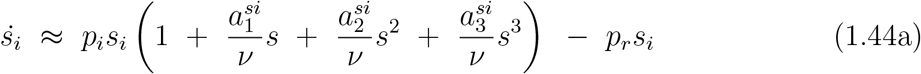

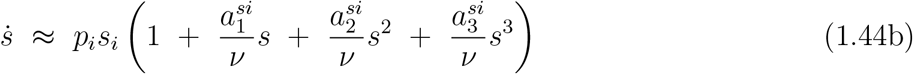

The prime purpose of truncating terms specifically of order > 3 here is that 3 is the largest polynomial order that offers the possibility of a relatively simple analytical solution of (1.42a). Another benefit is that it keeps the number of model-parameters within limits, while still allowing for a fair to very good description of the exact *s*−dependence of <*s*_*si*_> and <*s*_*si*_>.

Eqs. (1.42a,b) and (1.44a,b) provide a most insightful example of how correlations and fluctuations enter the mathematics of the problem and into the basic equations of the SIR-model. Comparison with (1.6) and (1.7) shows that the inclusion of correlations and fluctuations into the SIR-model comes down to nothing more than a replacement of the term (1 − *s*) in (1.6) and (1.7) by a series expansion of the form 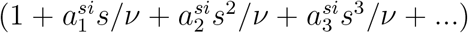. Since the expansion coefficients 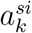 are generally expected to be functions of *ν*, the characteristics of the population network (and more in particular the number *ν* of social contacts of an individual) thereby enter the problem via the quotients 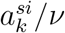. Only if the coefficients 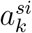 were *proportional* to *ν*, the influence of *ν* would cancel out against the *ν*-dependence of the 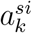. This is generally *not* the case however, and it is therefore already, that the structures of social networks within a population can be considered as a key factor in the evolution of an epidemic. This is in itself not an entirely new or unexpected insight (in fact it provides the epistemic basis for all practical measures limiting social contacts in order to bring outbreaks of infectious diseases under control). The particular merit of eqs. (1.42a,b) and (1.44a,b) however is that they put this already known viewpoint in simple mathematical terms that allow for a better understanding of the phenomena involved. It may be obvious that similar considerations also apply to the expansion coefficients 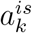 and the quotients 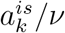, and therefore also to eqs. (1.44a,b) (for which truncation of terms of order > 3 has the same benefits as in the case of (1.42a,b)).

## 2. Reproduction numbers

The main results from the model outlined in this paper primarily evolve around the parameters *p*_*i*_ and *p*_*r*_, with an additional role for the parameters accounting for the network structure of the population. However, the dynamics of the spread of infectious diseases is often described and analysed in terms of different key parameters: the (well-known) *basic reproduction number R*_0_ [1] and *the effective reproduction number R*(*t*). These are defined, respectively, as the total number of new active infections generated by a single active infection already in existence at *t* = 0, and as the total number of new active infections generated by a single active infection counted from a moment in time *t* > 0 onwards. On the basis of the present model, expressions for *R*_0_ and *R* can be given in direct accordance with these definitions.

Consider an ensemble of *n*_*i*_ = *n*_0_ active infections at time *t* = *t*_0_. Due to infection-removal, the ensemble decays over time according to 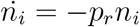 so that for *t* ≥ *t*_0_:

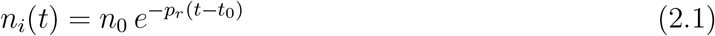

where *n*_*i*_(*t*) represents the remaining number of active infections at an instant *t*. The (average) total number of new infections generated after *t* = *t*_0_ by an infection active at *t* = *t*_0_ ≥ 0 is thus given by:

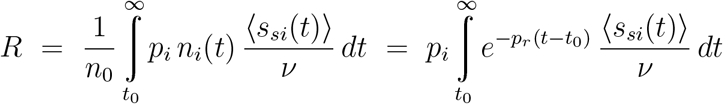

which can be reexpressed as:

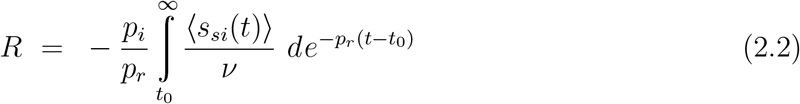

thus establishing the effective reproduction number R. Note that R number is *time dependent* : *R* = *R*(*t*_0_).

Introducing:

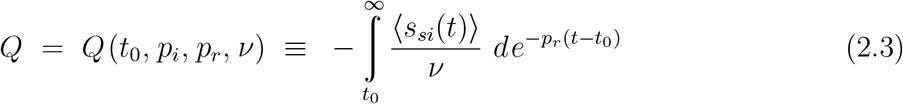

we can write the identity for *R* given by (2.2) as:

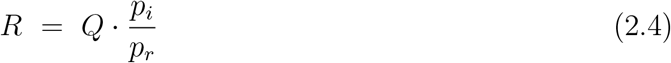

The factor *Q* accounts for the depletion of the reservoir of susceptibles that results from the spread of the infection (i.e. the reduction of *s*_*s*_ and therewith of ⟨*s*_*si*_⟩ with increasing *s*(*t*)).

The *basic* reproduction number *R*_0_ can now be considered as the value for *R* in the special case where *t*_0_ = 0 and *Q* = *Q*(0, *p*_*i*_, *p*_*r*_, *ν*) ≡ *Q*_0_. That is:

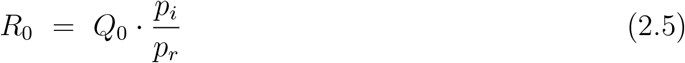

In the literature, reproduction numbers are often linked to criteria for epidemic spreading of an infection. For instance, it is often stated that an (exponential) increase in the number of infections will emerge when *R* > 1 (*R*_0_ > 1), whereas the infection rates will decline and fade-out when *R* < 1 (*R*_0_ < 1). To illustrate such criteria, a pictorial impression of epidemic evolution is often presented, in which active infections pass on their infection to a total of *R* susceptibles which, once infected themselves, pass on their infection to another *R* susceptibles etc. The result is a “tree” of infections, in which each infection forms a node (branch splitting) from which its infection is passed on along a total of *R* outgoing branches, so that after *N* generations (branch splittings) a total of *R*^*N*^ infections can be (indirectly) assigned to a single infection. Although quite illustrative and of considerable educational value, this picture is not entirely correct however. For instance, it ignores the depletion, accounted for by *Q*, of the reservoir of susceptibles upon progression of the epidemic. This depletion continues to progress for *t* > *t*_0_ as well. As such, each generation of new infections will find less susceptibles available to pass the infection on to than the previous generation. Therefore, the frequently drawn image (for the purpose of explaining exponential growth of the number of (cumulative) infections with time) of an infection tree with an equal *R*-related number of branches “growing” out of each infected node does not give a fully correct representation of what actually happens. In chapter 5 we will see that, although often taken for granted, also the condition *R* > 1 (*R*_0_ > 1) for epidemic growth of the number of infections requires serious (re)consideration.

## 3. Solving the differential equations

### a) 3rd-order polynomial approximation of ⟨*s*_*si*_⟩

Truncation of the terms of order > 3 in the series expansions representing < *s*_*si*_ > in (1.42a,b) has the advantage that the resulting set of ODE’s (1.44a,b) can be solved easily via partially analytical methods. With 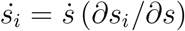 we can rewrite (1.44a) as:

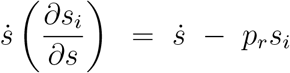

which, upon writing (1.44b) as 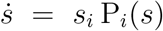, can be reworked into:

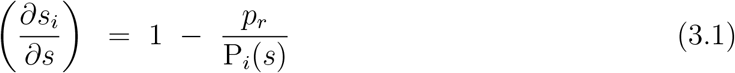

Substitution of the term in brackets in (1.44a) for P_*i*_(*s*) subsequently yields:

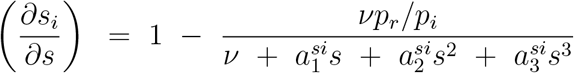

which we rewrite as:

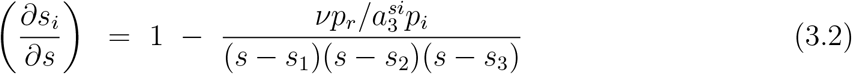

with *s*_1,2,3_ representing the roots (real and complex) of the 3rd-order polynomial 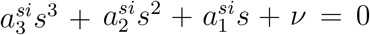, which can be calculated exactly via the somewhat tedious algebraic scheme of Cardano’s method (see, for instance, [1]). This scheme is easily implemented in a computational procedure however. We rewrite the fraction on the right-hand side of (3.2) via decomposition by parts:

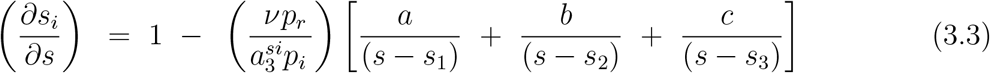

where the *a, b, c* are readily obtained as:

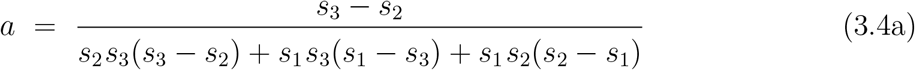

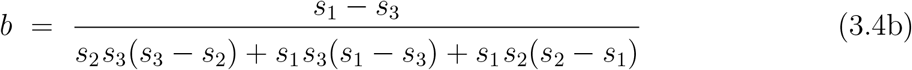

and:

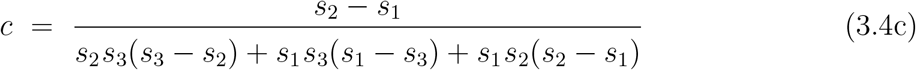

Integration of (3.3) is straightforward:

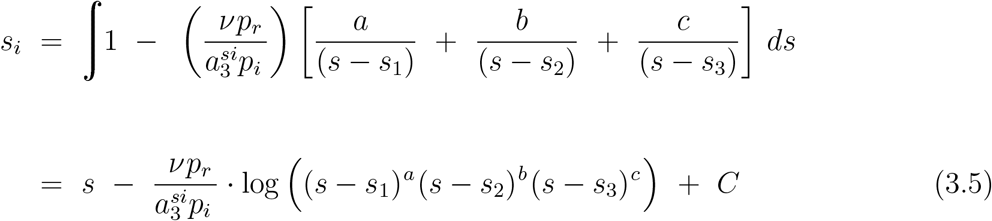

where the complex logarithm function is implicated, and *C* represents the constant of integration. The latter follows from the (initial) condition that *s*_*i*_ = *s* = *s*_0_ when *t* = 0:

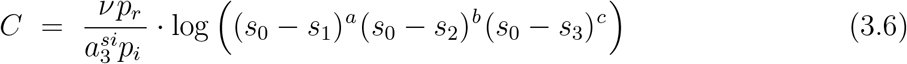

so that (3.5) can be rewritten as:

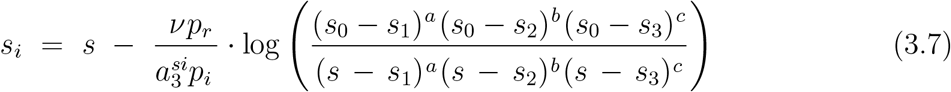

Via substitution of this result into (1.44b), the following non-linear ordinary differential equation for *s* as a function of *t* is obtained at last:

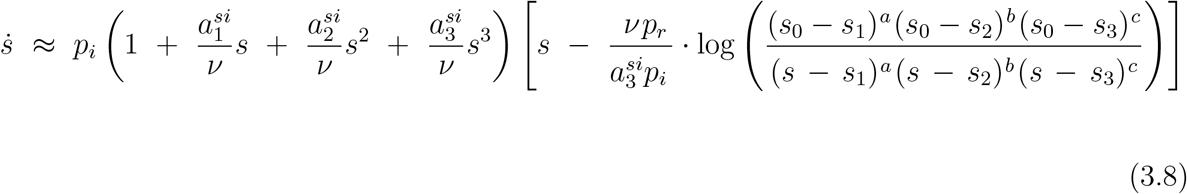

which can only be solved numerically, thus finalising the solution of the system of ODE’s given by (1.44a) and (1.44b).

### b) 3rd-order polynomial approximation of ⟨*s*_*is*_⟩

When approximating ⟨*s*_*is*_⟩ via a series expansion up to 3rd order in *s*, we can obtain the solution of the system of ODE’s that (1.43a) and (1.43b) therewith become along somewhat similar lines.

We have:

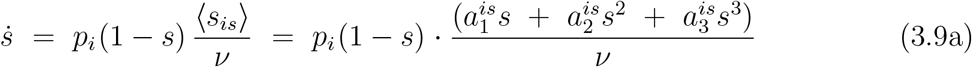

or:

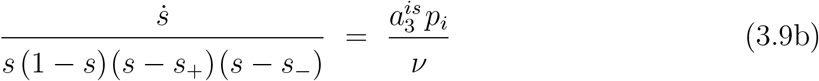

where the *s*_±_ represent the roots of 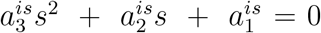. Via separation by parts and some algebra we can write:

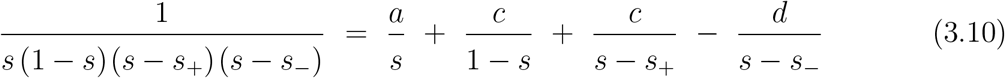

where:

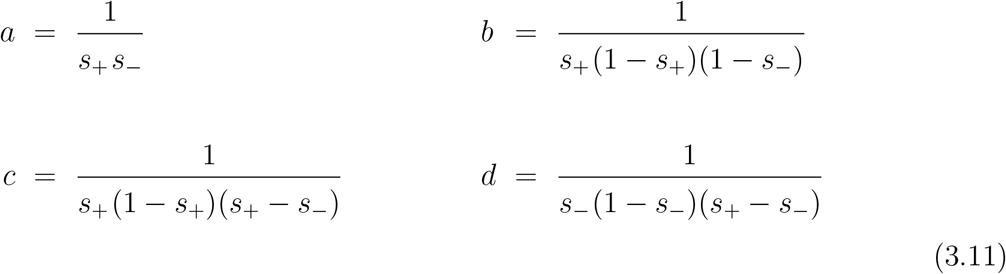

Remember that 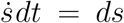 since *s* is a “state variable”. The solutions of the differential equations (3.9a,b) are now easily obtained by substitution of (3.10) into:

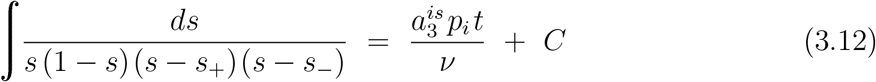

yielding, apart from a constant of integration:

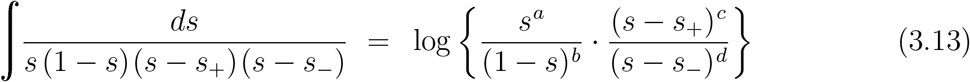

where, as in subsection **a)**, the complex logarithm function is applied. Via substitution of (3.11) for *a, b, c, d* into (3.13) we get, after some algebra:

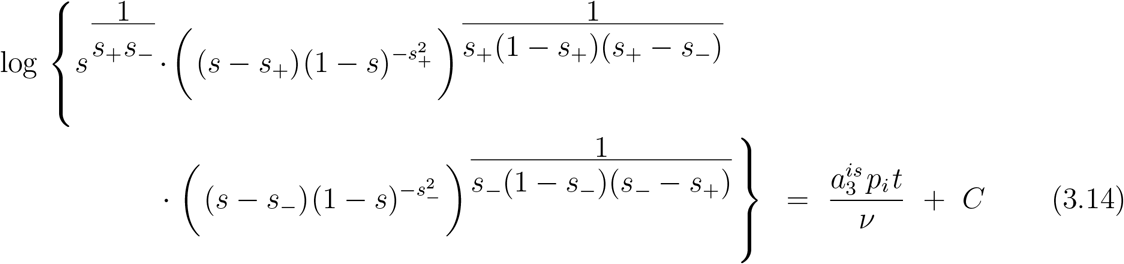

The constant of integration *C* is determined by value *s*′ of *s* at some arbitrary time *t* = *t*′ > 0 :^2^

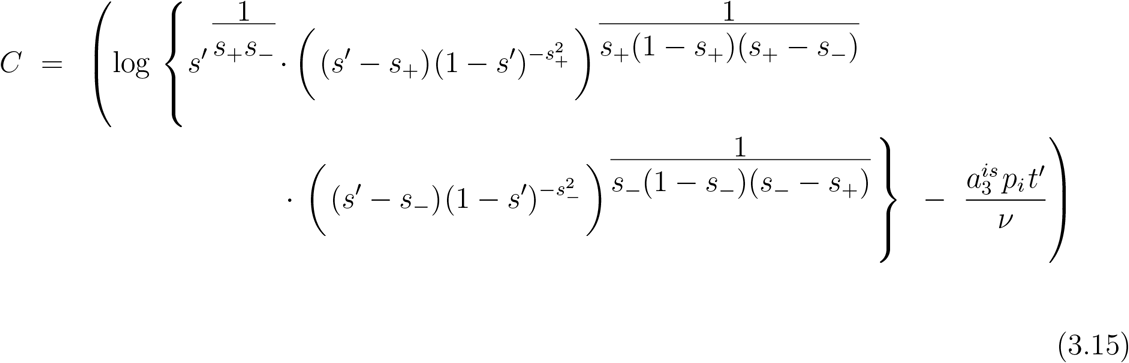

Combining (3.14) and (3.15) we then obtain:

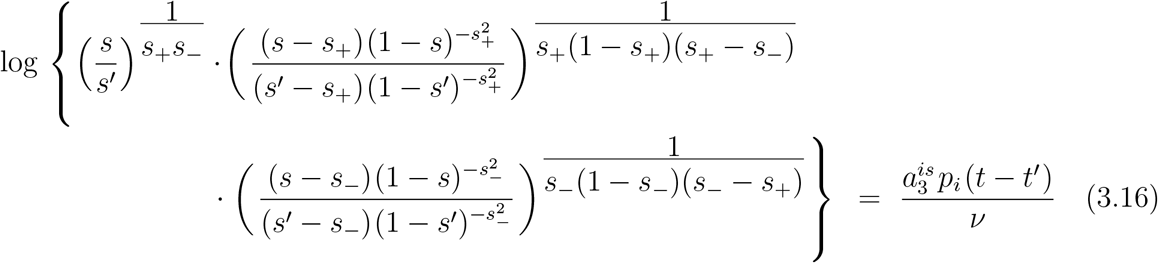

Note that the argument of the logarithm is always real, for when one of the roots *s*_±_ is complex, the other one is its complex conjugate. Solving *s*(*t*) for given *t* directly from this equation cannot be done via algebraic methods and requires a numerical procedure. However, the entire *s*-*t* curve can be obtained straightforwardly by using (3.16) to calculate *t* as a function of *s* and subsequently swap the axes. In principle, the parameters *t*′ and *s*′ can thereby be chosen in an arbitrary way.

The remaining ODE for *s*_*i*_, obtained by truncating terms of order > 3 in the series expansion in (1.43a), reads:

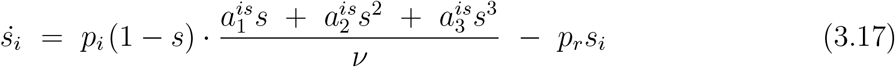

This equation must be solved via numerical integration under substitution of the appropriate values for *s*(*t*) obtained on the basis of (3.16). This then concludes the solution of (1.43a) and (1.43b) for this case.

### c) 2nd-order polynomial approximation of ⟨*s*_*si*_⟩

Truncation of the series expansion of ⟨*s*_*si*_⟩ down to the terms of order ≤ 2 provides a relatively simple approximative expression for *s*_*i*_(*s*) that nevertheless contains a lot of the essential features of exact *s*_*i*_ vs *s* relations, and thus provides an excellent “toy-model” for studying some of the fundamentals of the spread of infectious diseases through a population. This case is basically the limiting case where 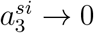 in the 3rd-order expansion case discussed under **a)**.

With 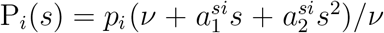 for this case, eq. (3.1) becomes:

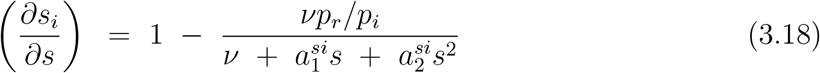

which we rewrite as:

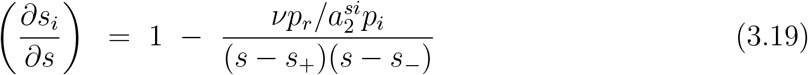

with *s*_+_ and *s*_−_ representing the roots (real and complex) of the equation 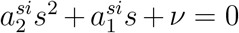, as given by:

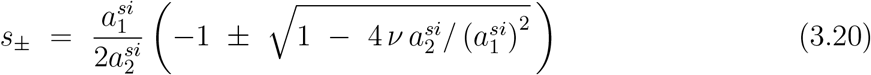

which have the rather convenient property that *s*_+_*s*_−_ is given by the very simple expression 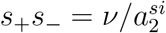, which will be of use later.

eq. (3.19) can be rewritten as:

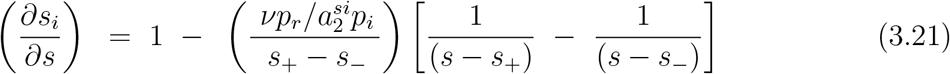

and its integration is straightforward:

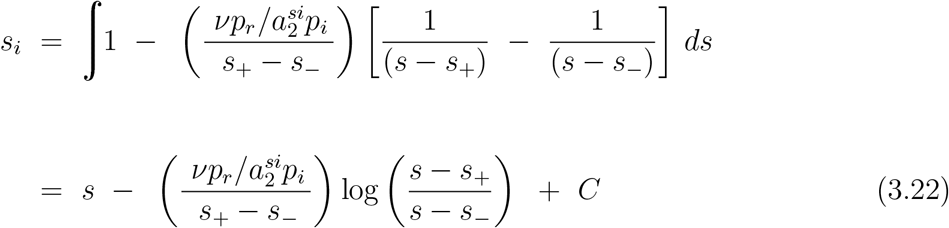

with, again, the complex logarithm function used, and *C* the constant of integration. The latter follows from the (initial) condition that *s*_*i*_ = *s* = *s*_0_ when *t* = 0.

We have:

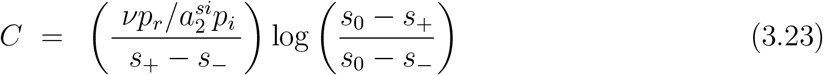

so that (3.22) can be rewritten as:

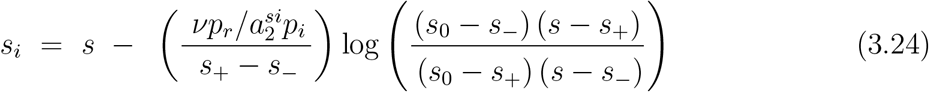

Using this result, the ODE for *s* in this case can then be integrated (which also requires a numerical procedure as in the case for the 3rd-order approximation of ⟨*s*_*si*_⟩ discussed under *a*), thus concluding the solution of the set of coupled ODEs (1.42a,b) in the present approximation.

As a general remark, it should be mentioned that solutions for *s*(*t*) have “physical” meaning only for 0 ≤ *s* ≤ 1, and when they describe a situation where 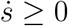, since (by definition) the cumulative number of infections cannot decrease with time. As such, physical solutions are confined to the specific interval of *s*-values where:

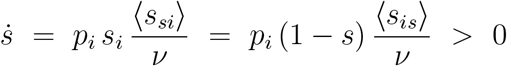

that is, where:

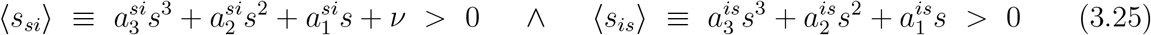

## 4. Simulations

The merits of the extension of the standard SIR-model presented in the previous chapters (essentially consisting of the introduction of the series expansions (1.11),(1.12) and (1.13)) can be demonstrated very well on the basis of results from simulations of the spread (in an SIR context) of infections through entire populations.

For the purpose of the aforementioned simulations, populations were considered in the form of a 2-dimensional (2D) square lattice, each node of a lattice representing a population member, while the edges of the lattice connect the nodes to their 4 nearest-neighbours contacts. Contacts were not restricted to nearest-neighbour contacts only however. To simulate the effects of a wider variety of restrictive social measures, nodes could also be considered as being at the centre of a (2*N* + 1) × (2*N* + 1) square of nodes representing potential contacts (note that there is always an even number of contacts to a single node in such cases). The values of *N* could be chosen at will. The limit of *N* → ∞ was approximated through a simulation where the contacts of a node were selected among *all* other members of the population. Nodes could be labelled as either susceptible (S), infected (I) or removed (R), in accordance with the SIR context chosen as the epidemiological model or setting. Periodic boundary conditions were applied to guarantee that all nodes have a similar “social bubble”, i.e. an equal number of contact-links connected to it.

The spread of an infection through a population can basically be seen as a stochastic process of a Markov type [1]. Such processes are particularly suited for simulations on the basis of a Monte-Carlo scheme (see [2] p. 17ff). With code written in Fortran, the algorithm used here was basically as follows. First of all, the population lattice is brought in its initial state by labelling all population members (nodes) as susceptible (S), except for a fixed number of randomly selected population members (nodes) that will be labelled as infected (I) and serve as initial infections. Random selection of nodes is done by calculating their 2D coordinates on the square lattice on the basis of 2 (pseudo-) random numbers provided by the build-in random-number generator of the compiler. Then the simulation of the actual spread of the infections through the population begins and proceeds in the following way. A member of the population (node) is selected at random. Then a 2nd node is selected in the same way from the nodes in the contact environment of the 1st node (that is, from the nodes linked to the 1st node as its possible contacts). If the 1st node is labelled infected (I) and the 2nd node is susceptible (S), or vice versa, a pseudo-random random number *r* is generated and compared to the transmission probability *w*_*i*_ = *p*_*i*_/2 (chosen by the user as a constant). If *r* ≤ *w*_*i*_ the infection is passed on to the susceptible node by labelling it as infected as well (instead of susceptible). For the purpose of simulating infection removal, another node (population member) is then selected, again at random. If this node turns out to be labelled as infected, a new random number *r* is generated and compared to the (user-defined) constant of infection removal *p*_*r*_. If *r* ≤ *p*_*r*_, the node is relabelled as a removed infection (R). This entire procedure of infection and subsequent removal is repeated a vast number of times.

When such a procedure is applied to each population in a large ensemble of *N*_*e*_ populations that are all in same state, an average number of *N*_*t*_ ≈ *N*_*e*_ *p*_*i*_ *s*_*i*_ ⟨*s*_*si*_⟩ of infections will be transmitted, whereas an average total of *N*_*r*_ ≈ *N*_*p*_ *p*_*r*_ *s*_*i*_ active infections will be removed. It is easy to see therewith that successive application of this procedure to a single population simulates the process of infection and removal described by the master equations (1.38) and (1.39).

The spread of the infection can be followed at arbitrary time scales by regularly monitoring the status (S,I or R) of all nodes in the population. The unit of time is itself the subject of a certain arbitrariness as well in this respect. In can be defined as corresponding to a fixed but arbitrarily chosen number of successive contacts made. In the simulations presented throughout this paper, the unit of time was taken such that it spans a number of contacts equal to the number of nodes/members in the population. So, in a single unit of time each member of the population makes exactly 1 contact on average.

Fig. 4.1 illustrates the results of a simulation for a case where *p*_*r*_ = 0 and contacts of the nodes were selected throughout the *entire* population. Every node is a potential contact to every other node therewith. Such cases represent the equivalent of the so-called mean- or molecular-field cases in the theory of (magnetic) phase transitions [3]. They are limiting cases, for which the standard SIR-model represented by eqs. (1.6) and (1.7) is actually exact. As such, they make an excellent test case to verify whether the simulation scheme described in the above may be a useful validation tool for models that go beyond the standard SIR-model.

**Fig. 4.1:**
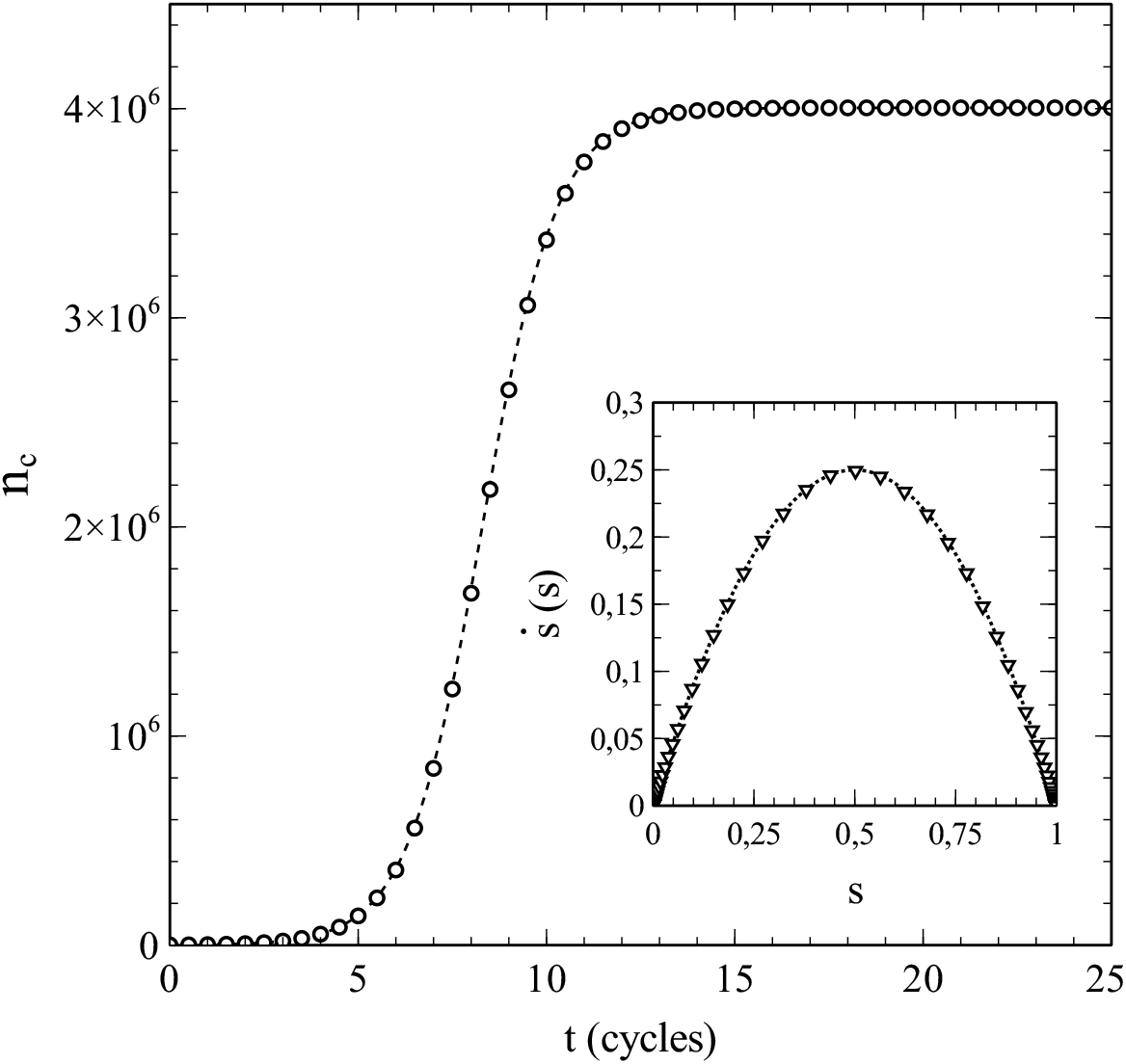
Number of cumulative infections *n*_*c*_ as a function of time (main figure) and 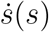 (inset) obtained from a simulation with node-contacts selected throughout the entire population network (2D square lattice). Dashed/dotted curves: standard SIR-model. Parameters: population size *n* = 2001^2^, transmission probability *w*_*i*_ = *p*_*i*_/2 = 0.5, decay/removal constant *p*_*r*_ = 0, number of initial infections *n*_0_ = 999.

Simulations presented throughout this paper were generally carried out on population lattices with a number of nodes typically in the order magnitude of 10^6^. The data presented in fig. 4.1 for instance where obtained from a simulation where the population was represented by a 2001 × 2001 square lattice (i.e. *n* = 4004001 nodes). These are quite large population population sizes indeed, which comes with the advantage that simulations become less prone to the typical finite-size effects that often complicate the interpretation of Monte-Carlo simulations for systems of relatively small size (see [2] p. 35ff). It should also be noted that such population sizes are actually quite realistic. A 2001 × 2001 square lattice consists of a number of nodes in the order of the size of the population of a country like Norway for example [4].

The simulation data presented in fig. 4.1 are in perfect agreement with the standard SIR-model. The dashed curve in the main figure shows the cumulative number of infections vs. time as obtained by solving eq. (1.7) for the same initial conditions and parameters used in the simulation (i.e. *n*_0_ = 999 so that *s*_0_ = *n*_0_*/n* = 2.4950 · 10^−4^, *p*_*i*_ = 2*w*_*i*_ = 1, *p*_*r*_ = 0). The solutions of the standard SIR-model are given by a so-called logistic function [5] in this case:

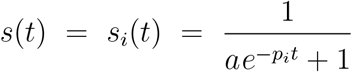

where *a* = (1 − *s*_0_)*/s*_0_. The simulated data follow the dashed curve remarkably well, and create confidence therewith in the adequacy of the implemented simulation scheme. An even more significant match with the standard SIR-model can be observed in the variation of 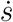 vs. *s* shown in the inset of fig. 4.1. The standard SIR-model yields 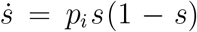 for *p*_*r*_ = 0 (see eqs. (1.6) and (1.7)), which is represented by the dotted curve in the inset. The datapoints (▽) show 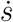 as obtained from a numerical evaluation from the simulated data. The (near) perfect agreement between the simulated data and the standard SIR-model is again obvious. As such, we may conclude that the simulations provide very reliable data for this case. It should also be noted in this respect that the large size of the populations used in the simulations already seems to pay off in the absence of any visual stochastic noise in the simulated data (which smoothly follow the dashed/dotted curves).

Simulations of cases where *p*_*r*_ ≠ 0 confirm the adequacy of the simulation scheme even more. When contacts to a single node are again selected from the entire population, the standard SIR-model applies to these cases as well. Figs. 4.2a/b show the results of simulations for the same (initial) conditions as the results shown in fig. 4.1, except that *p*_*r*_ = 0.5 instead of *p*_*r*_ = 0. Figs. 4.2a shows the simulated number of cumulative infections *n*_*c*_, and fig. 4.2b the number of active infections *n*_*i*_ = *ns*_*i*_, in both cases as a function of time. The dotted curves represent the corresponding numerical solutions of the system of differential equations (1.6) and (1.7) for respectively *n*_*c*_ and *n*_*i*_ (n.b. remind that *n*_*c*_ = *ns n*_*i*_ = *ns*_*i*_).

**Fig. 4.2:**
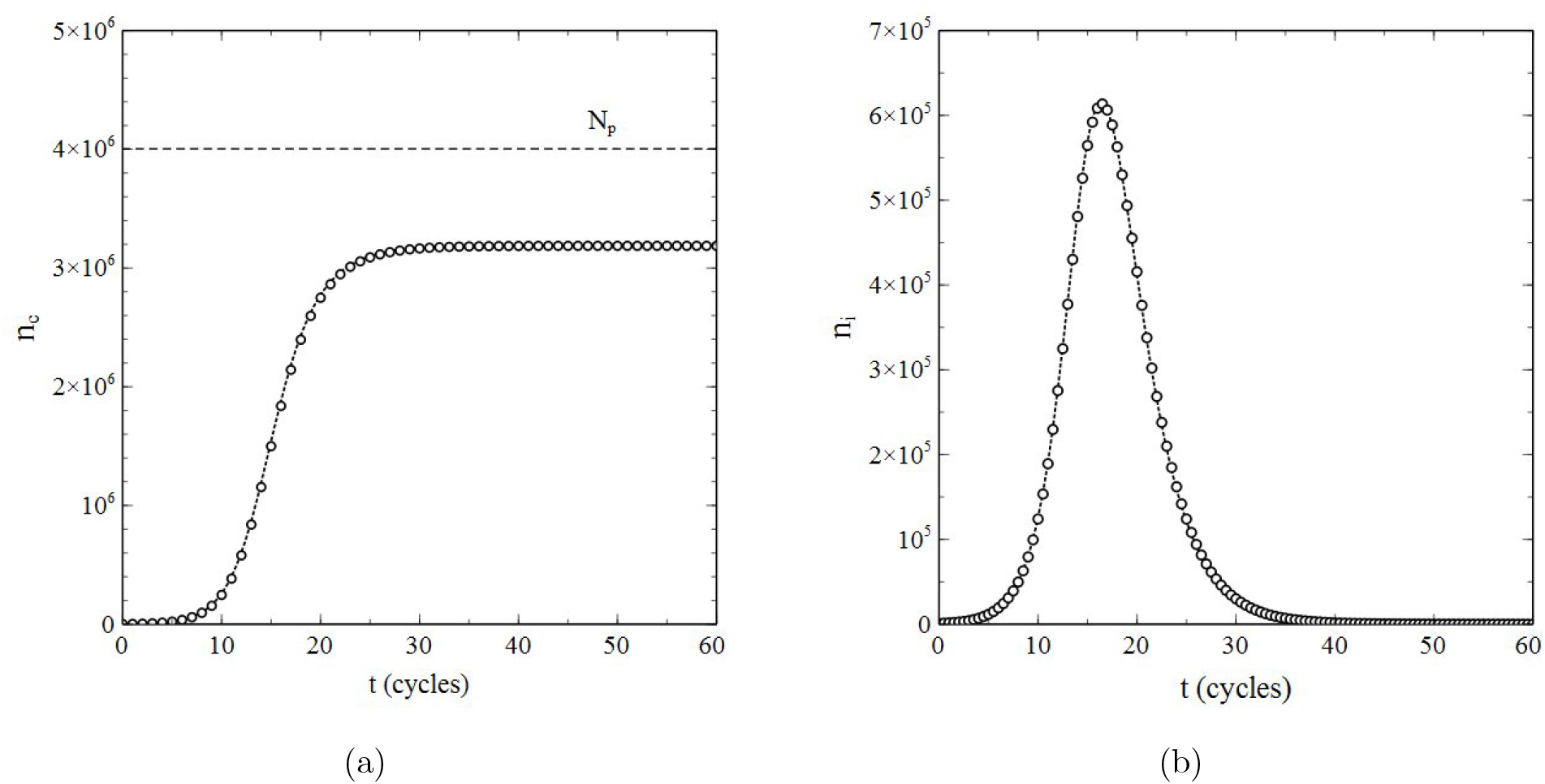
Data obtained from a simulation with *p*_*r*_ ≠ 0 and node-contacts selected throughout the entire population network (2D square lattice) for **a**): number of cumulative infections *n*_*c*_ as a function of time, and **b**): number of active infections as a function of time. Dotted curves: standard SIR-model. Parameters: population size *N*_*p*_ = 2001^2^, transmission probability *w*_*i*_ = *p*_*i*_/2 = 0.5, decay/removal constant *p*_*r*_ = 0.5, number of initial infections *n*_0_ = 999.

Due to the removal of active infections, not the entire population gets infected during the epidemic in this case ^3^, and the cumulative number of infections will reach a final value *n*_*e*_ < *n* (see the dashed horizontal line in fig. 4.2a indicating *n* = 2001^2^).

The simulated data in figs. 4.2a/b match the curves given by the standard SIR-model to a high degree of accuracy. We may therefore conclude that not only the stochastic nature of infection transmission, but also the stochastics of infection removal (decay) have been implemented correctly and realistically in the simulation scheme. This is further corroborated by fig. 4.3, which shows both 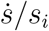 and 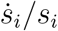 as a function of *s*, as derived on the basis of the data presented in fig. 4.2a/b via a simple numerical evaluation of 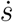 and 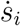 (*s*_*e*_ = *n*_*e*_*/n* indicates the maximum rate of cumulative infections reached). The data thus obtained agree very well with the standard SIR-model (where 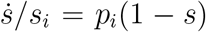 and 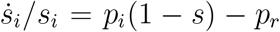: each set of datapoints obviously follows the straight line that the standard SIR-model predicts for it, especially for mid-range values of *s*. Only at the very edges of the *s*-interval that applies, some stochastic noise becomes noticeable. This is due to the fact that both at the beginning as well as at the end of any (real) epidemic, the number of active infections is relatively low and therefore subject to (temporal) fluctuations. The fact that this phenomenon apparently presents itself also in the simulation process deserves attention, since it does not reveal any shortcomings in either the algorithms used in the simulation or their implementation. On the contrary, it is rather to be considered as a realistic artefact of an appropriate simulation of the stochastic processes involved in an actual epidemic.

**Fig. 4.3:**
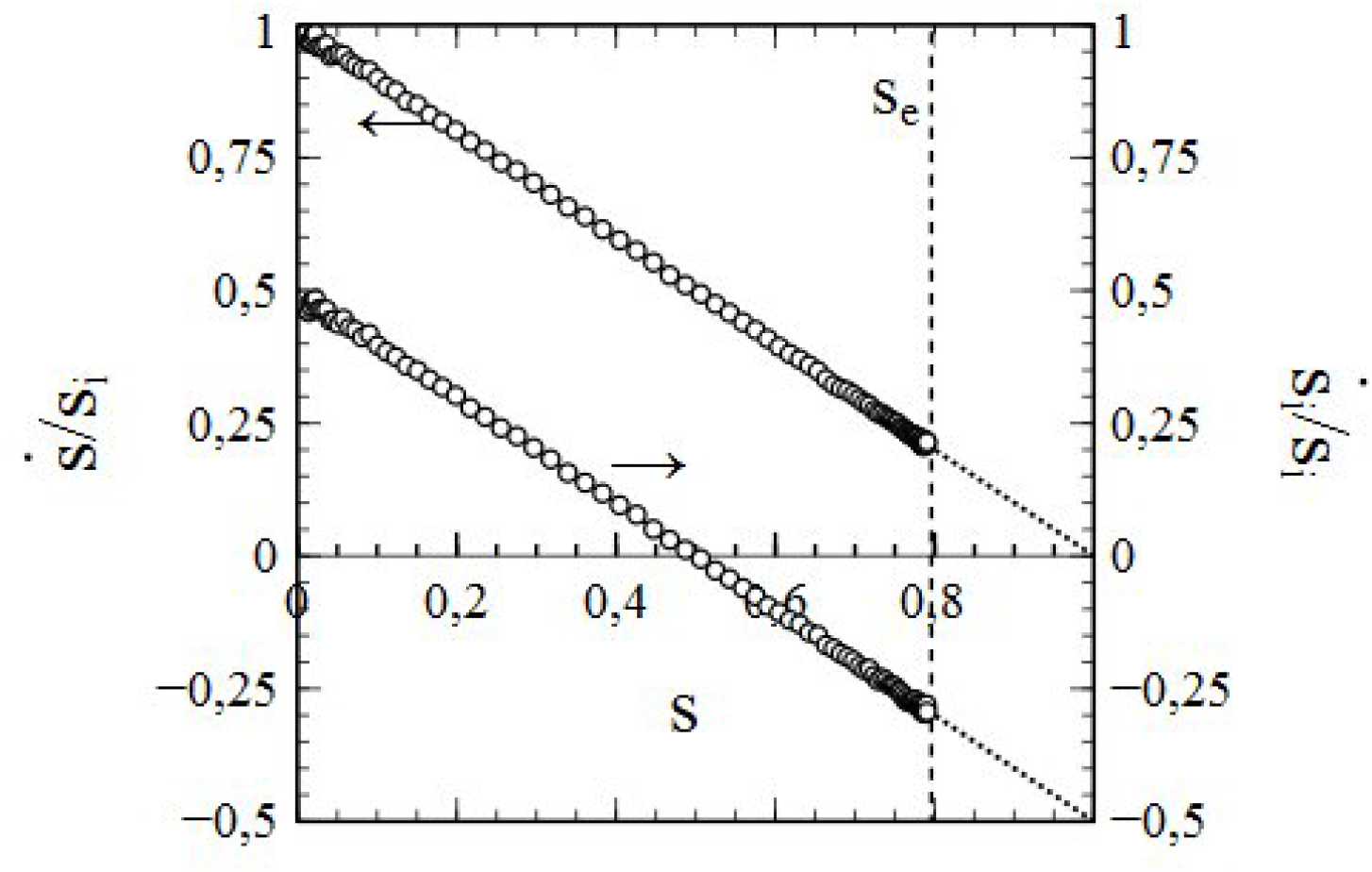
Simulated data for 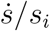 (left vertical axis) and 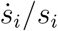 (right vertical axis), obtained from the same simulations as the data in fig. 4.2. Dotted lines: standard SIR-model (i.e. 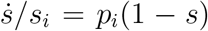 and 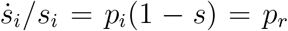 (extrapolated to *s* = 1)). Dashed vertical line *s* = *s*_*e*_: maximum *s* reached during the epidemic. Parameters: the same as for fig. 4.2.

However, as mentioned earlier, the standard SIR model is only a (mean-field like) approximation. Its breakdown comes when the social bubble of the nodes is increasingly reduced from an environment that spans the entire population (in which case the standard SIR model is exact) to smaller environments that contain only a limited number of nodes serving as contacts. This is clearly illustrated in figs. 4.4a/b, which respectively show ⟨*s*_*si*_⟩ and ⟨*s*_*is*_⟩ as a function of *s* for a series of simulations with *p*_*r*_ = 0, so that the entire population becomes infected in the end and *s* varies between 0 and 1 as a consequence. The contacts of each node were selected from a (2*N* + 1) × (2*N* + 1) square of nodes surrounding it. For each simulation, a different value of *N* was taken, so that the size of the social bubble of the nodes (given by *ν* = (2*N* + 1)^2^ − 1) differed per simulation. The values of *N* varied *N* = 2 to *N* = 16 (that is, the size of social bubbles varied from 24 to 1088). The dotted lines ⟨*s*_*si*_⟩ = 1 − *s* in fig. 4.4a and ⟨*s*_*is*_⟩ = *s* in fig. 4.4b represent the standard SIR model. The departure (with increasing *N* (and *ν*)) in the behaviour of ⟨*s*_*si*_⟩ and ⟨*s*_*is*_⟩ with *s* from the mean-field characteristics of the standard SIR model cannot be missed. This observation strongly indicates that the incorporation of the influence of the structure of the social networks and the size of the social-bubbles into the analysis is not merely an exercise, but rather a matter of plain necessity, and that the standard SIR model has serious shortcomings in this respect.

**Fig. 4.4:**
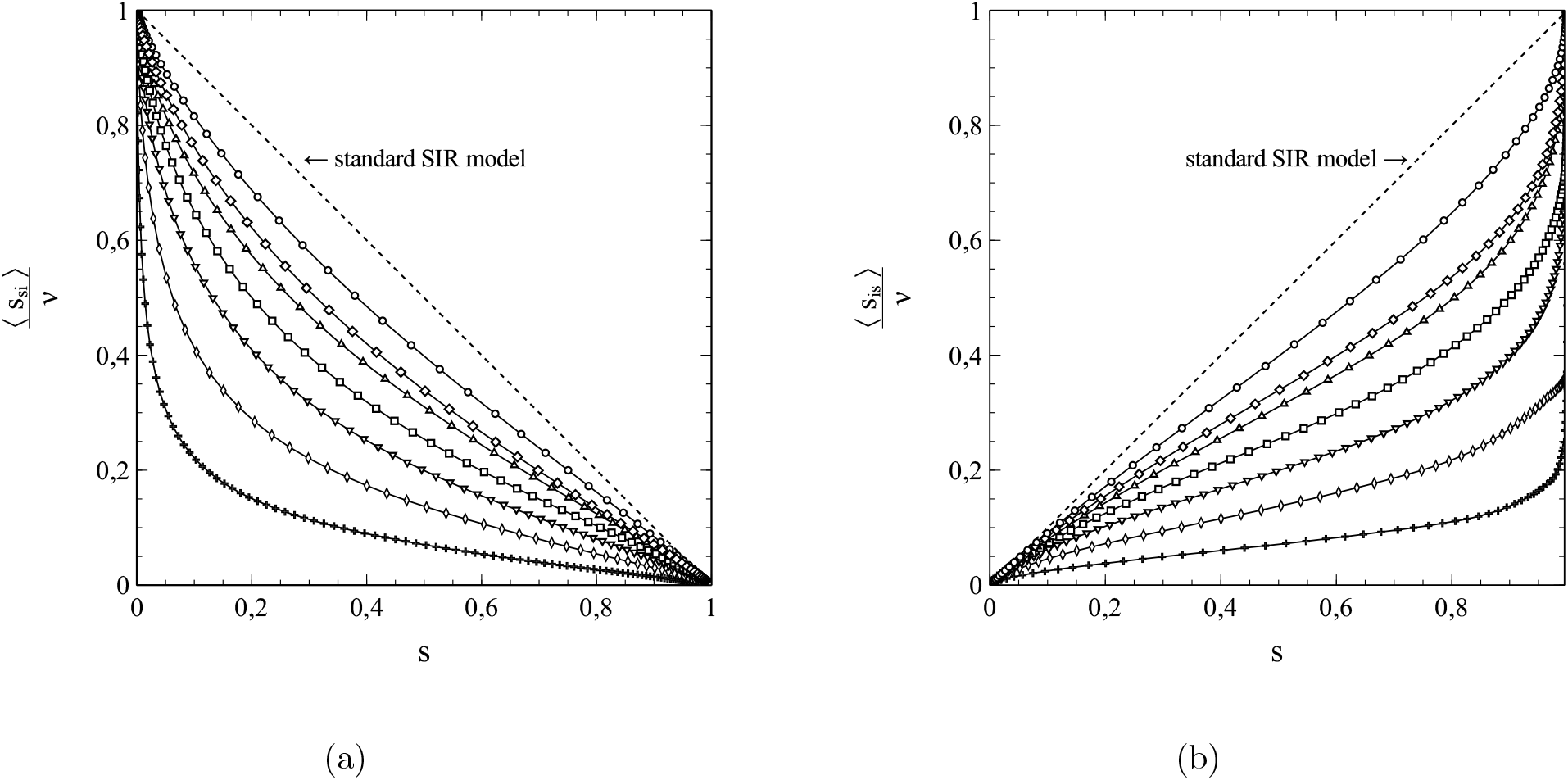
⟨*s*_*si*_⟩ (**a**) and ⟨*s*_*is*_⟩ (**b**) as a function of *s*, for a series of simulations with social bubbles consisting of (2*N* + 1) × (2*N* + 1) squares with *N* = 16 (∘), *N* = 12 (o), *N* = 10 (△), *N* = 8 (□), *N* = 6 (▽), *N* = 4 (◊), *N* = 2 (+). Parameters: population size *n* = 2001^2^, transmission probability *w*_*i*_ = *p*_*i*_/2 = 0.5, decay/removal constant *p*_*r*_ = 0, number of initial infections *n*_0_ = 999.

The usefulness of the method, presented in chapter 1, of dealing with the network structure via (truncated) series expansions in *s* for ⟨*s*_*si*_⟩ and ⟨*s*_*is*_⟩ can be illustrated well by deriving values of the expansion coefficients from the simulated data for either ⟨*s*_*si*_⟩ or ⟨*s*_*is*_⟩, taking these values as input for calculations of *s* as a function of time (*t*) (by solving either (1.42a,b) or (1.43a,b)), and comparison of the results with the *s* − *t*-data obtained from the simulations. It turns out that the variations of ⟨*s*_*is*_⟩ with *s* shown in fig. 4.4b can be described very well by a 3rd-order polynomial of the type 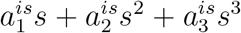 for all cases simulated (note that the standard SIR model is in fact a limiting case here with 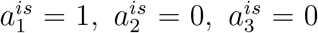). This is clearly illustrated by fig. 4.5a and fig. 4.5b, in which the results of the best fits of the expansion coefficients 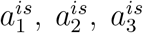 to the data for ⟨*s*_*is*_⟩ vs *s* obtained from the simulations for *N* = 6 and *N* = 10 are shown as representative examples. It is easy to see that, with the best-fitting values taken for 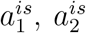 and 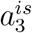^*s*^, the 3rd-order polynomials (dashed curves) describe the simulations (solid curves) to quite an acceptable level indeed. This is also true for the other cases investigated in this respect (i.e. *N* = 1, 2…16). That the best-fitting values for 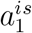 thus obtained provide by themselves an excellent reflection of the breakdown of the standard SIR model deserves special attention here. Fig. 4.6 shows these values as a function of *ν* (i.e the number of contacts per node *ν* or, equivalently, the social-bubble size). For very large values of *ν*, the value of 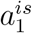 reaches towards its asymptotic value 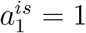 given by the standard SIR model (which represents the limiting case for *ν* → ∞). In the lower *ν*-regime however, the value of differs significantly from its mean-field value 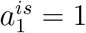, and upon decreasing *ν* well below *ν* ≈ 300 there is actually a collapse that disqualifies the standard SIR model even as an approximation in this regime of *ν*-values.

**Fig. 4.5:**
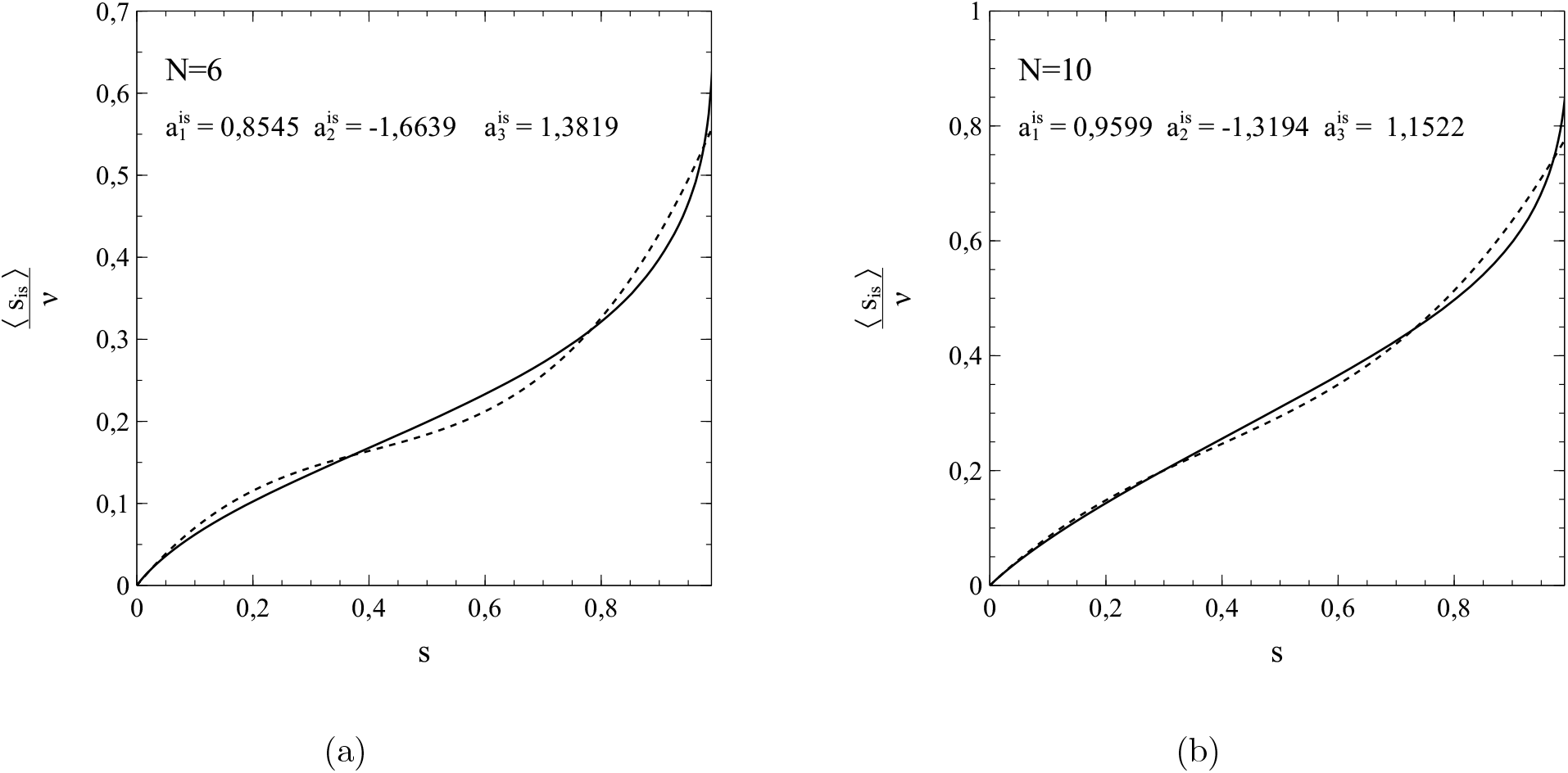
3rd-order polynomial fits (dashed curves) of data (solid curves) for ⟨*s*_*is*_⟩ vs *s* from simulations with *p*_*r*_ = 0 and *N* = 6, *ν* = 168 (**a**) and *N* = 10, *ν* = 440 (**b**). Other parameters: the same as for fig. 4.2. Best-fitting values for 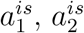 and 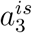 indicated in each figure.

**Fig. 4.6:**
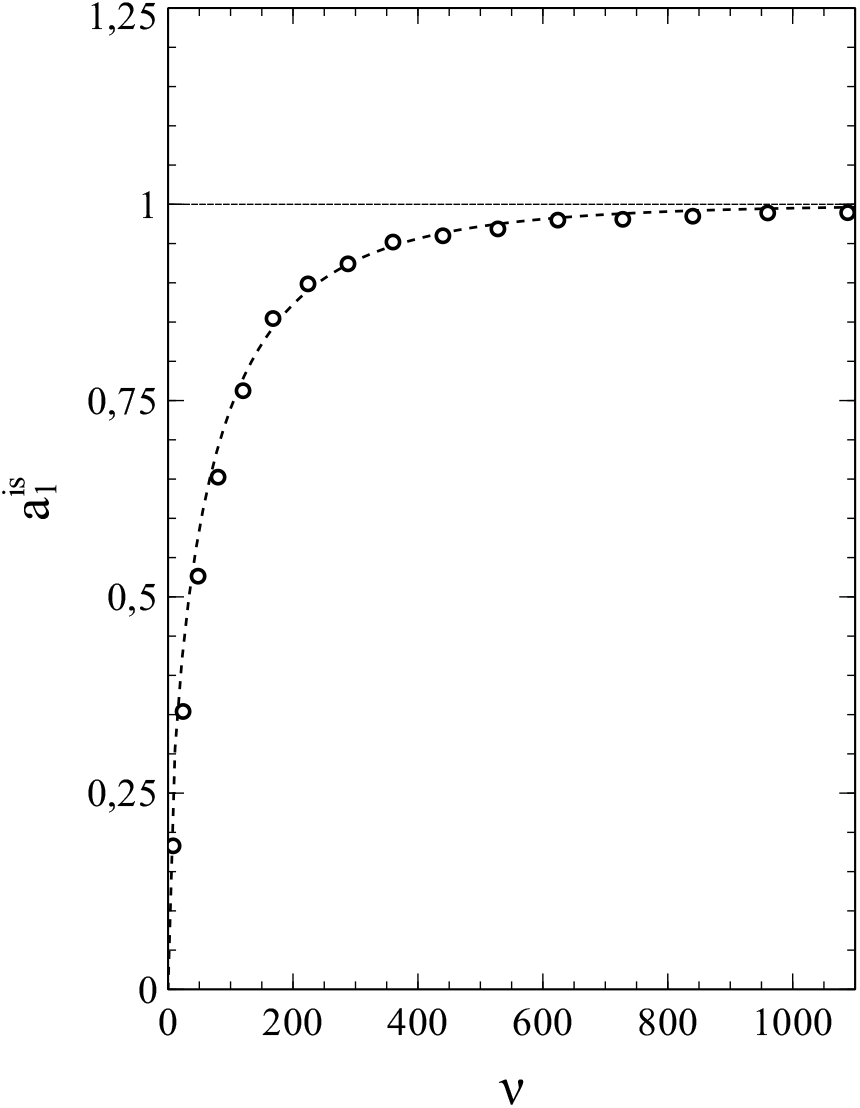
Variation of 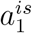 with *ν*

When *p*_*r*_ = 0 (and therefore *s*_*i*_ = *s*), the differential equations for *s*_*i*_ become identical to those for *s*, so that, depending on whether we use an expansion for respectively ⟨*s*_*si*_⟩ or ⟨*s*_*is*_⟩, we only have to solve either (1.42b) or (1.43b) to obtain *s* as a function of *t*. For *N* = 12, 10, 8, 6, 4, 2 (*ν* = 624, 440, 288, 168, 80, 24), the variation of *s* with *t* was calculated by numerically solving (1.43b) (see section 3b for details) for the best fitting values of 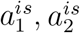 and 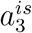 for each *N* (as obtained from the previously mentioned fits of ⟨*s*_*is*_⟩ vs *s*). The results are shown in figs. 6a-f. In each case, the marked datapoints represent the simulations and the dashed curves the respective solutions of (1.43b). The dotted curves relate to the results given by the standard SIR model for the particular set of parameters used here (i.e. *w*_*i*_ = *p*_*i*_/2 = 0.5, *s*_0_ = 2.4950 · 10^−4^). The agreement between the solutions of (1.43b) and the simulated data is equally noticeable as the discrepancy that grows (with increasing *N*) between them and the results from the standard SIR model. The significance of this observation is twofold. One one hand it shows that the method of expressing ⟨*s*_*is*_⟩ as a series expansion in *s* has its merit. On the other hand, it further corroborates our previous observations about the inadequacies of the standard SIR model and the mean-field approach that underlies it.

When *p*_*r*_ ≠ 0 (so that *s*_*i*_ ≠ *s*), we have to solve either both equations (1.42a) and (1.42b) or both equations (1.43a) and (1.43b) simultaneously. Using 3rd-order polynomial approximations for ⟨*s*_*is*_⟩ vs *s* is not a viable option however. The reason is that ⟨*s*_*is*_⟩ drops sharply towards zero upon approaching *s* = *s*_*e*_ (as a consequence of the removal of infections). At low to intermediate values of *s*, ⟨*s*_*is*_⟩ may still be approximated well by a 3rd-order polynomial as a function of *s* (as in the *p*_*r*_ = 0 case), but the approach of *s* = *s*_*e*_ is accompanied by a rather steep drop in ⟨*s*_*is*_⟩ towards zero (for an example see fig 4.8). The resulting functional dependence of ⟨*s*_*is*_⟩ on *s* over the entire interval *s*_0_ ≤ *s* ≤ *s*_*e*_ can no longer be appropriately described by a 3rd order polynomial in *s*, and using (1.43a,b) is therefore not an option. Fortunately, the dependence of ⟨*s*_*si*_⟩ on *s* does show the desired characteristics and can be approximated fairly well in terms of a 3rd-order polynomial, at least for *N* - and *ν*-values not too low (see fig. 8). We can therefore use (1.42a,b) to investigate the cases where *p*_*r*_ ≠ 0. Such cases are of particular additional interest, since they offer an extra possibility to demonstrate the merits of expressing ⟨*s*_*si*_⟩ or ⟨*s*_*is*_⟩ as series expansion in *s*, by showing that they not only allow for an accurate reproduction of the simulated *s* − *t* curves (cumulative infections) but of the *s*_*i*_ − *t* curves (active infections) as well. The procedure for this is conceptually similar to the one followed in the above for the *p*_*r*_ = 0 cases. We fit the simulated ⟨*s*_*si*_⟩ vs *s* data with a polynomial of the type 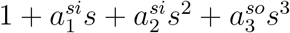 and take the best fitting values of the coefficients 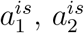 and 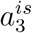 as input for solving the differential equations (1.42a,b) via the algebraic/numerical method outlined in section 3a.

Fig. 4.9 shows the results obtained in this context for *w*_*i*_ = *p*_*i*_/2 = 1, *p*_*r*_ = 0.25 and *N* = 12, 10, 8 (*ν* = 624, 440, 288). The graphs in the left column show the simulated data (marked as grey circles) of *s*_*i*_ vs *t*, as well as the corresponding results obtained on the basis of the standard SIR model for the parameters involved (solid curves). The graphs in the right column show the same simulated data (also marked in grey) as in the graph to their left, but then with the solution of (1.42a,b) (solid curve) for the values of 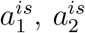 and 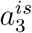 best fitting to the respective ⟨*s*_*si*_⟩ vs *s* data obtained from the simulations. The left column shows again a dramatic failure of the standard SIR model. In contrast, the column to the right shows an excellent (*N* = 12) to still quite reasonable (*N* = 8) match between the simulated data and the solutions of (1.42a,b). This includes the position of the maximum so dramatically and consistently missed by the standard SIR model in the left column.

In case of the cumulative infection rate *s* vs *t*, the agreement between the simulated data and the corresponding solutions of (1.42a,b) is even slightly better than in the case of the active-infection rate *s*_*i*_. This is clearly shown in fig. 4.10. The solutions of (1.42a,b) (solid lines) follow the simulated data (markers) extremely well. We also see that with decreasing *N*, the curves show a tendency to shift to the right along the *t*-axis. A similar tendency can be observed in the curves for *p*_*r*_ = 0 shown in fig. 4.7. This tendency can be understood as a direct manifestation of network and correlation effects. For instance, when the social bubbles become *smaller*, the relative *decrease* of the number of susceptibles that an active infection has left in its bubble after transmitting its infection to one of its contacts becomes *larger*. For smaller social bubbles, there is also an increased tendency towards the formation of small clusters of active infections sharing parts of their social bubble with other active infections. This typically leads to the kind of slow-down of the spread of the infection that we see in fig. 4.10. The solutions of (1.42a,b) follow this process perfectly well, in contrast to the standard SIR model which, from its very concept, does not account for network and correlation effects at all.

**Fig. 4.7:**
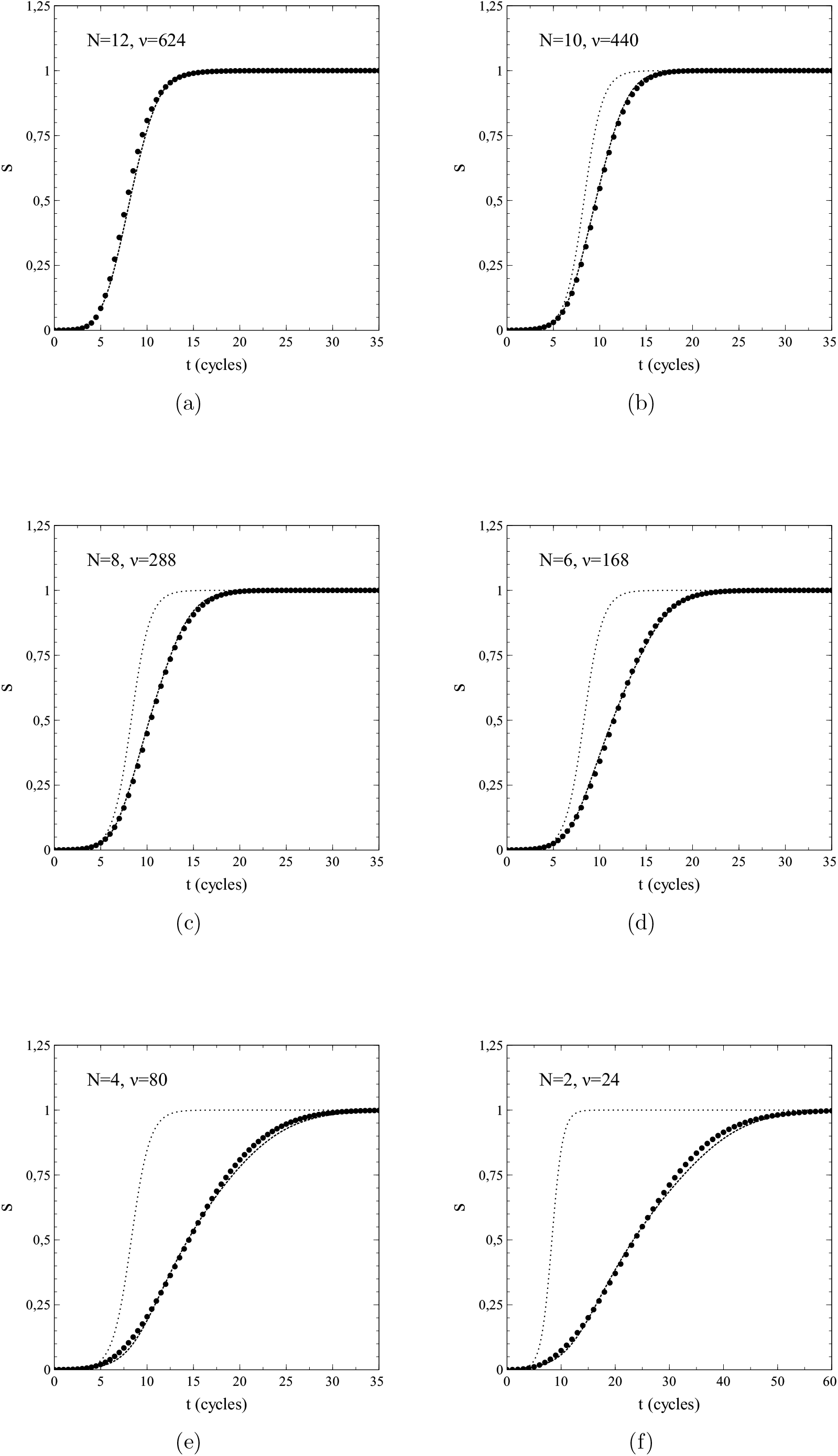
*s* vs *t* for *p*_*r*_ = 0 and *N* = 2, 4, 6, 8, 10, 12. Dashed curves: fit. Dotted curves: standard SIR model (not indicated for *N* = 12)

**Fig. 4.8:**
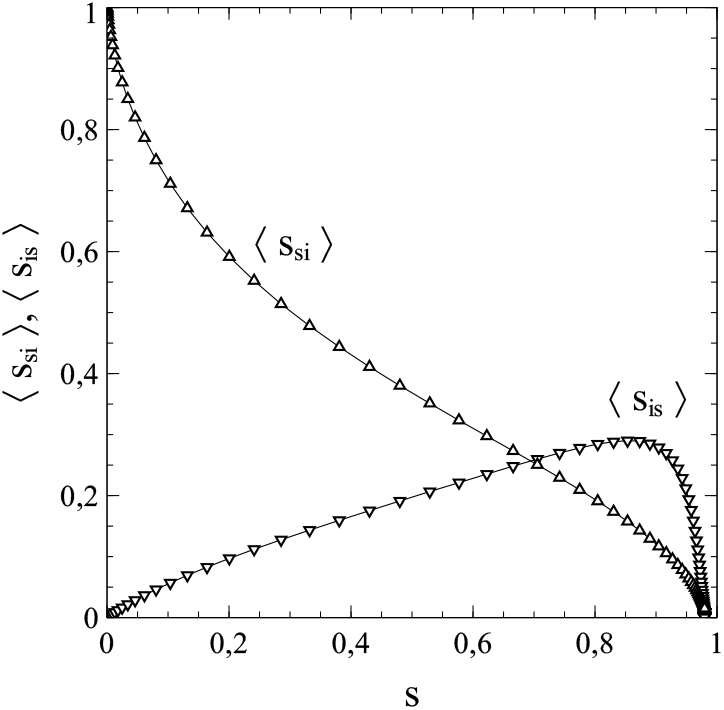
Variation of ⟨*s*_*si*_⟩ and ⟨*s*_*is*_⟩ with *s* for *w*_*i*_ = *p*_*i*_/2 = 0.5, *p*_*r*_ = 0.25 and *N* = 10.

In conclusion we can say that the approximation of ⟨*s*_*si*_⟩ and ⟨*s*_*is*_⟩ as series expansions in *s* works quite satisfactory, especially in the *p*_*r*_ = 0 cases but also when *p*_*r*_ ≠ 0, provided *N* (or, more general *ν*) is not too small in the latter cases. Simulations show that the effects of a finite size of the social bubbles becomes noticeable at fairly large sizes already. Even for *N* = 12, a situation where the total number of possible contacts of a single node is as large as 624, both the variations of *s* and *s*_*i*_ with time (*t*) show significant quantitative variations from the mean-field behaviour that applies in the limiting case *N, ν* → ∞ and for which the standard SIR model is exact. Now, in real life, a number of 624 is a very large size for the social bubble of an average single member of a population when considered in an epidemiological context. From an epidemiological viewpoint, the social bubble of an individual in a population contains only those members of the population contacted intensively enough by the individual on a regular basis to make a transmission of an infection carried by one of the contacting members to the other possible (albeit not necessarily certain). As such, the social-bubble size depends on the critical exposure/uptake for the pathogen involved, defined as the exposure/uptake necessary for a full blown infection to develop in a population member: a lower critical exposure increases the social-bubble size. Also the route of transmission affects the social bubble size (airborne pathogens have their own notoriety in this respect). However, a number well in the hundreds for the (average) epidemiological social-bubble size in a population seems quite on the high side for even the more infectious of pathogens. Nevertheless, even in the cases of such large social bubbles, there is a substantial discrepancy between the actual time evolution of the infection numbers and the one provided by the standard SIR model for the applying parameter values. This has serious consequences in relation to the extraction of values for *p*_*i*_ and *p*_*r*_ from field-data about the spread of an actual infection. We see from figs. 4.7, 4.9 and 4.10 that the *qualitative* behaviour of the actual infection data does not differ significantly from that found for the mean-field case on the basis of the standard SIR model. It may seem tempting therefore to fit the standard SIR model to the actual data via adjustment of *p*_*i*_ (and optionally *p*_*r*_ where relevant). Especially in cases where only data on *s* vs *t* are used, this may result in fits that reproduce the field data fairly well. However, it is easy to see that the values for *p*_*i*_ or *p*_*i*_*/p*_*r*_ thus obtained substantially underestimate the actual values (a reduction of the social bubble size also reduces the growth-rate 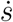 of the number of cumulative infections for given *p*_*i*_ or *p*_*i*_*/p*_*r*_). This is a serious problem indeed, since *R*_0_ = *p*_*i*_*/p*_*r*_ is often taken for the basic reproduction number (see section chapter 5 section d)), which plays an important role in practice for the assessment of the severeness of an outbreak/epidemic or of the risks associated with a particular pathogen in itself. More reliable estimates for *p*_*i*_ and *p*_*r*_ can be obtained, at least in principle, via the method of series expansions outlined in this paper or via direct simulation. The problems do not end there however. It looks like the coefficients of the series expansions for ⟨*s*_*si*_⟩ and ⟨*s*_*is*_⟩ cannot be calculated easily via simple algebraic methods or easily implemented numerical methods, at least not for *any* network of arbitrary structure (for the purpose of this chapter the coefficients were extracted from data generated by rather CPU intensive simulations for instance). This is a serious issue, since the structure and topology of the population network are expected to have a profound impact on at least the quantitative aspects of the spread of an infection, but perhaps also on the qualitative aspects. In connection to this we may refer to the Ising problem, where the dimensionality of the lattice (which affects, for instance, the number of nearest neighbours to a site/node) has major implications even for the qualitative behaviour of the system under consideration. The 1-dimensional Ising model does not show any order at finite temperatures (no matter how low) [6], whereas the 2- and 3-dimensional versions of the model do exhibit ordering phenomena at temperatures above zero, but with different values of the corresponding critical temperature and critical exponents [7], the latter putting the 2 versions in different universality classes. Such observations are typical for systems with network or lattice features, and there is no reason to assume that population networks make an exception in this respect, especially since we will see in the coming sections that the spread of infectious diseases may be associated with its own kind of critical phenomena. Therefore, the extraction of *p*_*i*_ - and *p*_*r*_-values from field data for the purpose of obtaining highly accurate estimates is a quite a problematic affair troubled by fundamental difficulties. In order to obtain reliable values for instance, such an extraction cannot go without obtaining estimates for the coefficients in the series expansions of ⟨*s*_*is*_⟩ or ⟨*s*_*si*_⟩ as well, either from field- or simulated data.

**Fig. 4.9:**
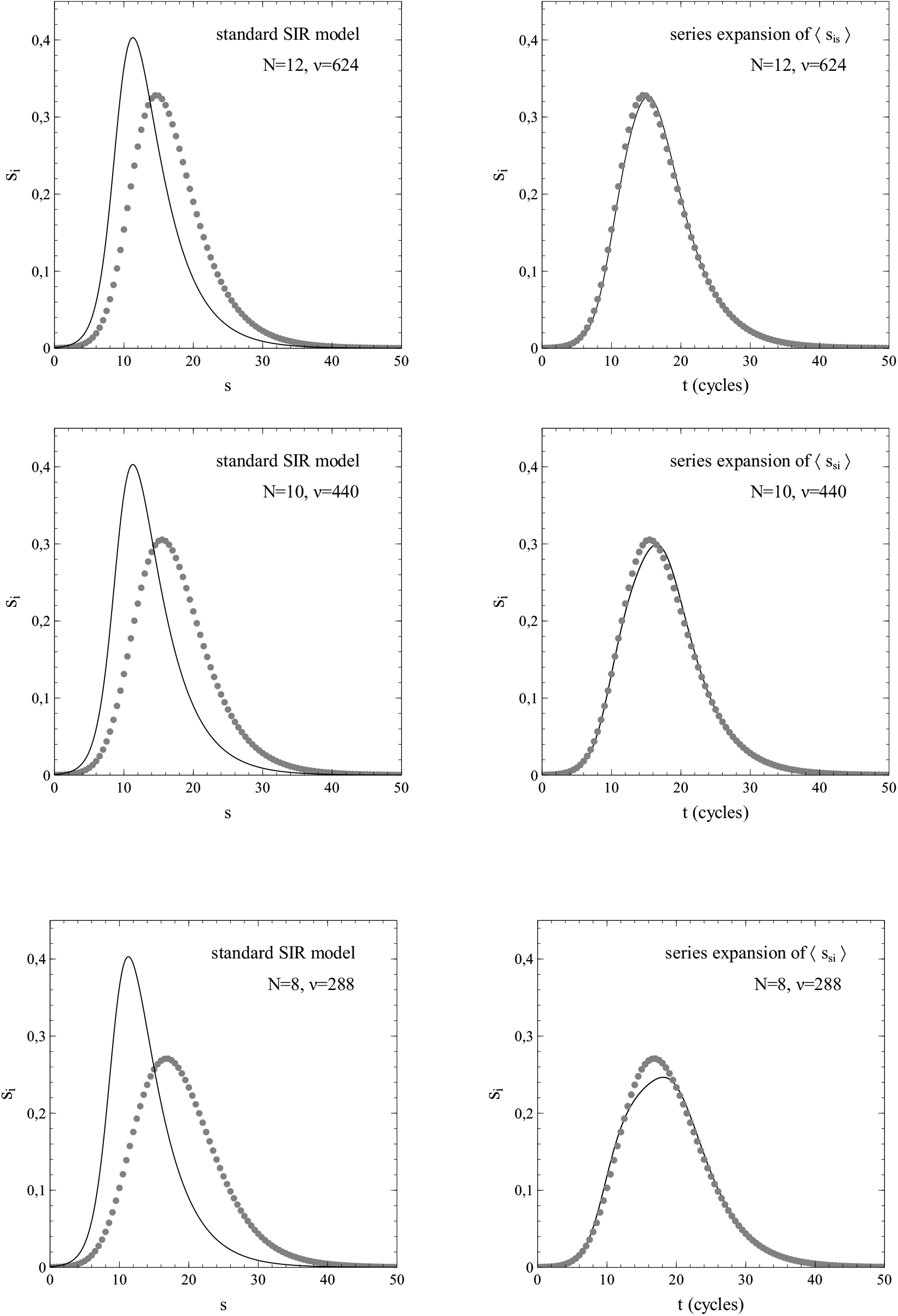
*s*_*i*_ vs *t* for *p*_*r*_ = 0.25 and *N* = 8, 10, 12. Left column: simulated data (markers) and standard SIR model (solid curve). Right column: simulated data (markers) and model based on series expansion of ⟨*s*_*si*_⟩. Other parameters: same as in fig. 4.2.

**Fig. 4.10:**
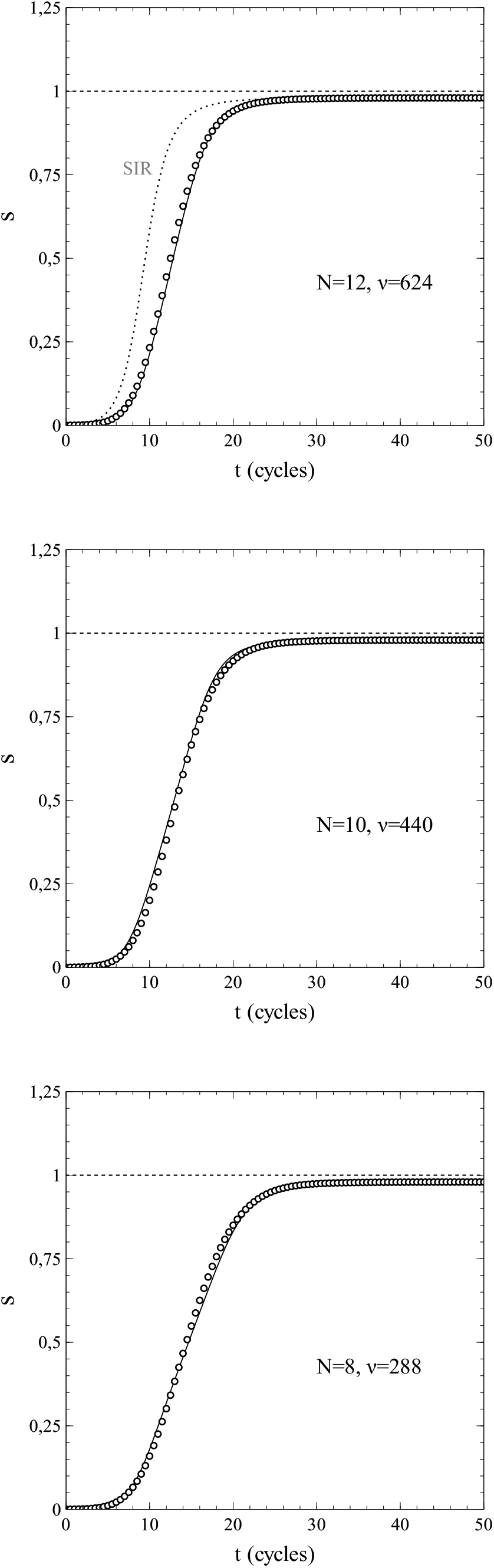
*s* vs *t* for *p*_*r*_ = 0.25 and *N* = 12, 10, 8 (other parameters: same as in fig. 4.2.). Simulated data (markers) and solutions of (1.42a,b) (solid curves). Dotted curve in upper figure: standard SIR model (as indicated in grey).

## 5. Properties of the model and its solutions: conditions for an epidemic, effects of infection removal

Even without solving the sets of differential equations represented by (1.42a,b) and (1.43a,b) completely, certain results key in the evolution of an epidemic can be derived from them.

### a) Criterion for an epidemic to develop from a limited number of infections

From (1.42a) we immediately see that for 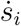 to be positive for *s* → 0 (and thus for sufficiently low values of *s*):

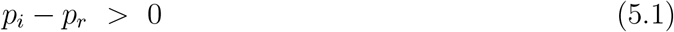

This is an interesting result, which actually implies a basic criterion for the possibility that a number of initial infections, so limited that *s*_*i*_ = *s*_0_ ≈ 0, will grow into a rampant epidemic or not. Only when the rate of transmission per active infection (*p*_*i*_) is higher than the rate of removal (*p*_*r*_), the number of active infections *ns*_*i*_ (and therewith also the cumulative number of infections *ns*) will grow even on the basis of just a very few initial active infections.

All this may sound plausible, but the inequality (5.1) not only provides a mathematical foundation to common sense in this respect, but also points directly towards general strategies that can be deployed during the onset of an epidemic. To reduce the spread of infection one may first of all refer to protective measures or reducing the frequency of social contact (i.e. reducing *f*_*cn*_). The effect of these is a reduction of *p*_*i*_. The more effective they are, the more they will reduce *p*_*i*_ and the slower the infection will propagate at given *p*_*r*_. On the other hand one may look at cures and medication (when available). The sooner active infections can be eliminated the larger *p*_*r*_ will become, thus hampering the spread of the infection. In *theory*, the possibility even exists of smothering a major outbreak well before it even started: if by taking appropriate measures the value of *p*_*i*_ − *p*_*r*_ can be made *negative* (*p*_*i*_*/p*_*r*_ < 1), a large scale epidemic might be averted. For that to achieve by protective measures alone, it is important to mention that it is not necessary to reduce *p*_*i*_ to zero. Only a reduction of *p*_*i*_ sufficient enough to make *p*_*i*_ − *p*_*r*_ negative (i.e. *p*_*i*_*/p*_*r*_ < 1) will do. It is stressed however that when such measures fail and (5.1) is actually met, sooner or later the active infection rate will grow vigorously (in fact exponentially) with time, as may be inferred from (1.42a). For *s* ≪ 1, eq. (1.42a) reduces to 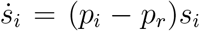, the solution of which is an exponential function of *t*, and when *p*_*i*_ − *p*_*r*_ > 0 the result will be an exponential *increase* in time of the active infection rate that may easily grow to epidemic proportions.

An important observation to be made here is that there is no reference to the structure of the individual social networks in the inequality (5.1): the criterion it represents follows independently of the social fabric of the population. As such, at least within the context of the model, (5.1) is universal and applies to *all* populations, irrespectively of their (social) network structure.

### b) The maximum number of active infections reached during an epidemic

Furthermore, eq. (1.42a) allows us to obtain an expression for the value of *s* at which the overall (global) maximum is reached in the number of active infections. A necessary condition for such a maximum is 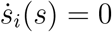, which relates, in case of a global maximum, to an extremum both as a function of *t* and as a function of *s*. From (1.41a) and (1.42a) we see that that 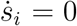 either when *s*_*i*_ = 0 or when P(*s*) = *p*_*i*_ ⟨*s*_*si*_⟩ */ν* − *p*_*r*_ = 0. The first case (*s*_*i*_ = 0) can be discarded, since it relates to the end of the epidemic which, in strict mathematical terms, is (always) approached asymptotically when *t* → ∞.

We will now evaluate the criterion for a maximum in *s*_*i*_ for the case where ⟨*s*_*si*_⟩ is approximated by a 2nd order polynomial in *s*. This case represents the simplest deviation from the standard SIR model possible. However, as a toy model it can be quite instructive. The relevant ODE’s and their solutions are given in section 3c.

When a global maximum in *s*_*i*_ exists in the context of a 2nd-order polynomial approximation of ⟨*s*_*si*_⟩, it must relate to one of the solutions *σ* = *σ*_±_ of the quadratic equation 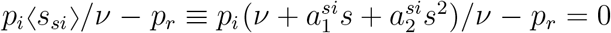. That is:

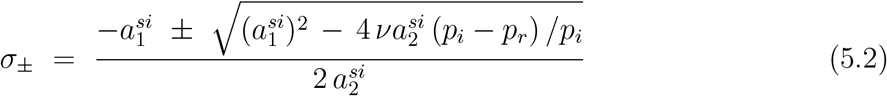

A particular solution *σ*_±_ actually corresponds to (global) maximum when:

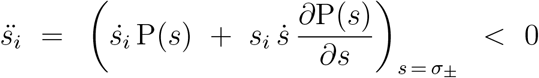

With 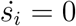 for *s*(*t*) = *σ*_±_ that is:

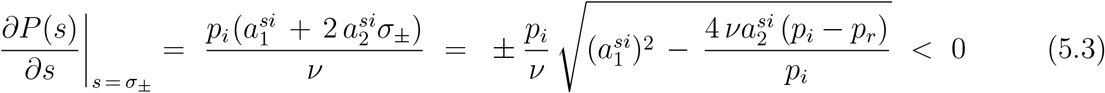

which can be met only in case of the minus sign, i.e. by the solution *σ*_−_, and (of course) only when the term under the square-root sign is non-negative.

Since it seems fairly evident that 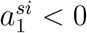 we have to distinguish between only 2 regimes of the expansion parameters, namely 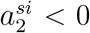 and 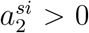. An epidemic requires *p*_*i*_ − *p*_*r*_ > 0 to start and propagate (see section 3a). In such a situation, it can be inferred from (5.2) that when 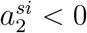 then *σ*_+_ < 0 and *σ*_−_ > 0, and that when 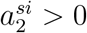 then *σ*_+_ > 0 and *σ*_−_ > 0. Since *σ*_−_ is positive for both 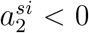 and 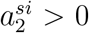, there can be a local maximum in both cases, its value being obtained by substitution of *s* = *σ*_−_ into (3.24):

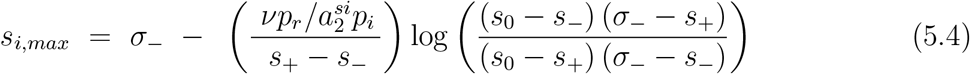

The maximum in *s*_*i*_ results from a competition between the production of new infections 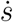 and the removal of already existing infections −*p*_*r*_*s*_*i*_. For *s*-values below *σ*_−_, new infections arise at a higher rate than that existing infections are removed. At *s* = *σ*_−_ the generation of new infections is precisely compensated by the infection removal and a maximum in *s*_*i*_ is reached. For *s* > *σ*_−_, the removal of infections over-compensates the generation of new infections, and a reduction of *s*_*i*_ sets in so that the epidemic or outbreak gradually fades out.

It is emphasized that, in general, the maximum in *s*_*i*_ does *not* mark a simultaneous onset of a decrease in the growth rate of new infections 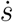.

With 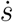 given by (1.32) the maximum in the rate of new infections is given by:

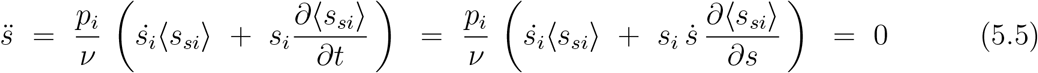

This equation cannot be solved by algebraical methods in general. However, it is straight-forward that when the maximum in *s*_*i*_ (which corresponds to 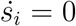) and the maximum in 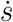 were to occur at to *the same* value of *s*, this would require the 2nd time-derivative of *s*:

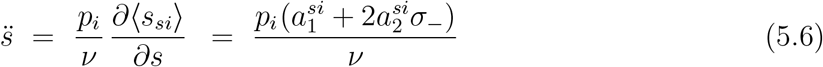

to vanish. Via substitution of *σ*_−_ that requirement is easily restated as:

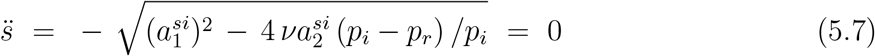

which holds only for very specific parameter combinations that make the term under the square-root sign zero. Hence, the maxima in *s*_*i*_ and 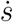 do, in general, *not* occur at the same *s* and *t*. In fact, unless 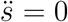, the rate of new infections is always in decline already when *s*_*i*_ reaches its maximum at *s* = *σ*_−_, since 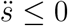 because of the minus sign in front of the square root in (5.7). The maximum in 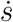 therefore precedes the maximum in *s*_*i*_, at least in the present model. This result may be of importance for policy and decision making during an ongoing epidemic. The observation that the rate of new infections has apparently reached its peak still means that the peak in the number of active infections is not there yet. Since the burden on the healthcare system due to an epidemic is largely determined by the number of active infections, this may have its implications, for instance with respect to matters of healthcare capacity.

### c) The effects of infection removal and the herd-immunity pitfall

The final stage of the epidemic/outbreak is characterised by a stabilisation (asymptotic in time) of *s* at some finite value *s*_*e*_ (n.b. 0 ≤ *s*_*e*_ ≤ 1), whereas the *active* infections fade out (*s*_*i*_ asymptotically approaches zero when *t* → ∞, *s* → *s*_*e*_). More important, the entire spread of the infection gradually comes to a halt, as the rate of infection 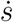 also becomes zero. The latter is in fact *the* quintessential feature of a fade-out of an epidemic, and the influence of the network structure of the population is crucial in it.

The role of the network structure in the evolution of an epidemic can be described in more detail as follows. When the number of active infections approaches zero (*s*_*i*_ → 0), the social network of a susceptible individual will consist more and more of removed infections and (non-infected) susceptibles only. Only a decreasingly small and negligible minority of susceptible individuals is still vulnerable to infection by active infections from within their social network. The root cause of this phenomenon on a “microscopic” scale (that is, on the level of individual nodes) is the removal of infections (*p*_*r*_ ≠ 0), under the assumption that removed infections either relate to individuals that have overcome the infection and acquired immunity, or to individuals that have succumbed to the infection. In both cases, such an individual then corresponds to an “inert” node in the network, unable to become infected again and to pass on the infection to other nodes/individuals. The infection can no longer propagate through the population via such nodes. In fact, active infections may even become surrounded by removed infections (immunized nodes) only, thus providing a shield between that particular infection and the rest of the population (rendering the infection unable to infect other nodes). With time (and therefore with *s*), the number of inert nodes increases to such an extent (relative to the number of active infections *s*_*i*_) that at some level the removal rate exceeds the rate of new infections: after having reached a maximum for *s* = *σ*_−_ (see previous section), the number of active infections begins a steady decrease and the epidemic gradually fades out, as its propagation is more and more hampered by the mechanism described here. As an important corollary of such a mechanism, the epidemic generally comes to an end even before all the members of the population have been infected: the cumulative number of infections then stabilizes at a value *s*_*e*_ ≠ 1. The way in which the spread of the infection is hampered by inert nodes in the population lattice via a blockade of infection routes, makes that the spread of an infection has all the characteristics of a percolation phenomenon. We will discuss this extensively in chapter 8.

In mathematical terms, the end of an epidemic can be defined as the situation where *s*_*i*_ = 0 (which implies also that 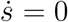 (see for instance (1.42b)). This allows for obtaining an equation for *s*_*e*_. By setting *s*_*i*_ = 0 in (3.24) (2nd order polynomial approximation of ⟨*s*_*si*_⟩), *s*_*e*_ can be identified as the solution *s* of the equation:

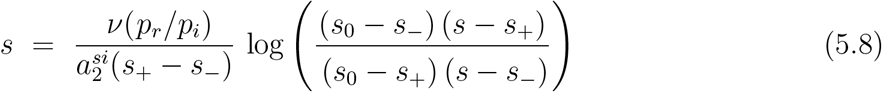

which we reexpress, with 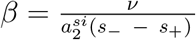, as:

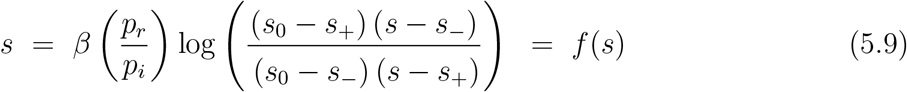

As a condition to be met by *s* = *s*_*e*_, (5.9) can be reexpressed even further, via some algebra, in terms of the inverse of the function *f* (*s*) as:

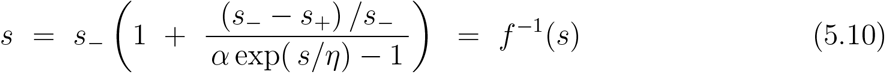

where *η* = −*β*(*p*_*r*_*/p*_*i*_), and the factor *α* is given by:

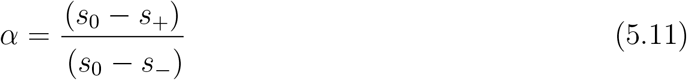

Furthermore, it should be noted that when *p*_*r*_ ≠ 0, the function *f* (*s*) actually represents the number of removed infections at given *s* < 1, which at the end of the epidemic (*s* = *s*_*e*_) becomes equal to the cumulative number of infections. It should be kept in mind that this is not the case for *f* ^−1^(*s*) however. The function *f* (*s*) differs from its inverse, and *f* ^−1^(*s*) has been introduced solely to *re*express the equilibrium condition for *s* implied by (5.9) and *not* to explicitly reexpress the right-hand part of (5.9) in general (i.e. for all *s*). As such the function *f* ^−1^ represents the number of removed infections only for *s* = *s*_*e*_.

eqs. (5.8), (5.9) and (5.10) cannot be solved for *s* via algebraic methods but require numerical or graphical techniques, the latter being quite instructive however. The function *f* ^−1^(*s*) is easier to handle than *f* (*s*) in that respect, mainly because of the divergent behaviour of *f* (*s*) at *s* = *s*_−_ and *s*_+_. Furthermore, *f* ^−1^(*s*) is more appropriate for demonstrating analogies between certain aspects of the dynamics of an epidemic and concepts in thermodynamics and statistical mechanics. It is for these reasons that *f* ^−1^(*s*) was introduced in the first place to serve as a substitute for *f* (*s*) in (5.9).

As a consequence if 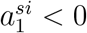 we have *η* ≤ 0, which can be verified easily by substitution of (3.20) for *s*_±_:

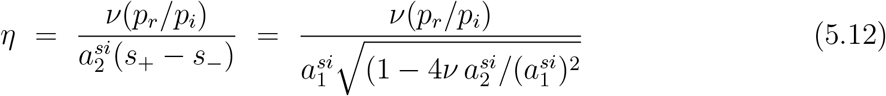

Also, the following inequalities apply to *s*_±_, as can be verified easily on the basis of (3.20) as well:

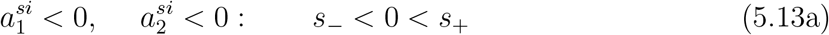

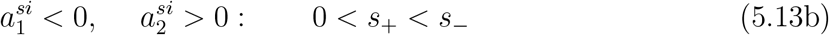

Focussing on the implications of the requirement expressed by (3.25) in case 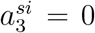, it is straightforward that when 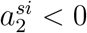 (i.e. when (5.13.a) applies) then 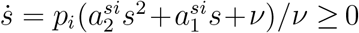 *only for* 0 ≤ *s* ≤ *s*_+_. The same is true when 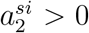 (that is, when (5.13.b) applies). We can therefore conclude that the root *s*_+_ constitutes an upper bound to *s*_*e*_ when *s*_+_ ≤ 1.

We will now investigate the characteristics of the function *f* ^−1^(*s*) in some more detail. For that purpose, we introduce the continuation of the function *f* ^−1^(*s*) on the interval 0 ≤ *s* ≤ *s*_+_ to the function *g*(*s*) on (−∞, ∞):

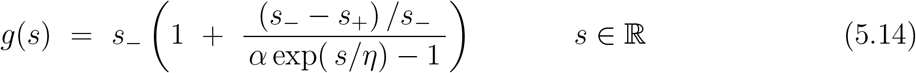

That is, we take the right-hand part of (5.10) as the rule of a function *g*(*s*) with domain ℝ instead of the interval 0 ≤ *s* ≤ *s*_+_.

The 1st derivative of *g*(*s*) is readily obtained as:

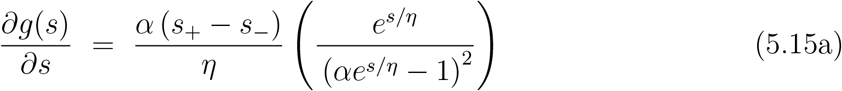

and reexpressed via substitution of 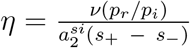 and 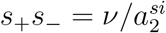 as:

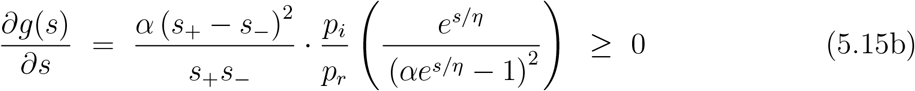

Since *η* ≤ 0 when 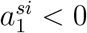 (see (5.12):

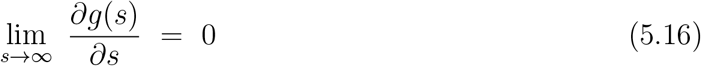

so that we can conclude that *g*(*s*) approaches a horizontal asymptote as *s* → ∞. Furthermore, for those cases relevant, a quick examination^4^ of the 2nd derivative ∂^2^*g*(*s*)/∂*s*^2^ shows that the function *g*(*s*) may have an inflection point *only for negative s*. So, as illustrated in fig. 5.1, for *s* ≥ 0 the function *g*(*s*) is therefore a monotonously increasing, concave function of *s* with a horizontal asymptote:

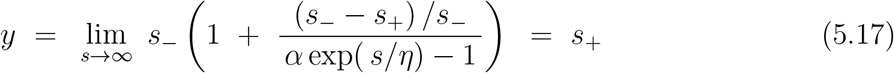

**Fig. 5.1:**
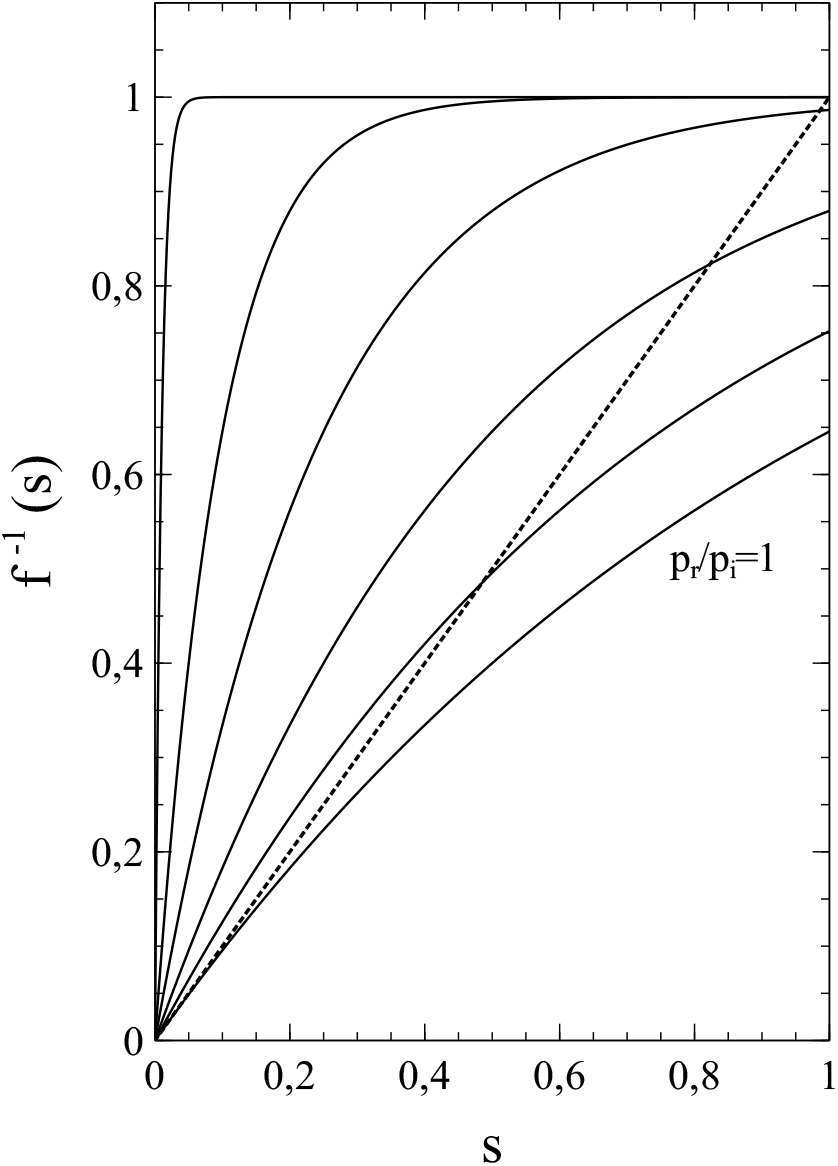
*f* ^−1^(*s*) vs *s* for *p*_*r*_*/p*_*i*_ = 0.01, 0.10, 0.25, 0.50, 0.75, 1 (solid curves) and the straight line *y* = *s* (dotted). Values of parameters: *ν* = 8, *a*_1_ = −7.2, *s*_2_ = −0.8. Solutions of (5.10) correspond to the intersections of the relevant graph of *f* ^−1^(*s*) vs *s* and the line *y* = *s. s* = 0 is always a solution. For *p*_*r*_*/p*_*i*_ < 1 a 2nd solution *s* > 0 exists. With increasing *p*_*r*_*/p*_*i*_, the 2nd solution gradually moves towards *s* = 0. For *p*_*r*_*/p*_*i*_ = 1, both solutions converge into a single solution *s* = 0, the line *y* = *s* being the tangent of the corresponding graph of *f* ^−1^(*s*). For *p*_*r*_*/p*_*i*_ > 1, only *s* = 0 remains as a solution

Furthermore, via straightforward substitution of *s* = *s*_0_ = 0 into (5.14) we get *g*(0) = 0. Hence, for *s* > 0:

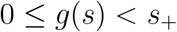

and consequently, for 0 ≤ *s* ≤ *s*_+_:

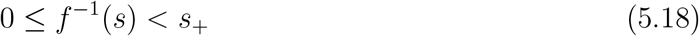

Now, if they exist, the solutions *s* = *s*_*e*_ of (5.10) are given by those intersections of the graph of *f* ^−1^(*s*) vs *s* and the straight line *y* = *s* that take place at an *s*-value in the interval 0 ≤ *s* < *s*_+_ when *s*_+_ < 1, or in the interval 0 ≤ *s* ≤ 1 when *s*_+_ ≥ 1 (see fig. 5.1). Besides *s* = 0 (which is always a solution) there is only a *single* intersection possible at most for *s* > 0 (due to the monotonously increasing concave nature of *g*(*s*), the absence of inflection points for *s* > 0 and the horizontal asymptote of *g*(*s*)). So, if a solution *s* = *s*_*e*_ > 0 of (5.10) exists (and therefore of (5.9), then that is *the only* solution.

Furthermore, when *s*_+_ < 1 then *s*_*e*_ < 1 as a direct consequence of (5.18). The latter mathematically demonstrates the possibility that significant parts of the population may remain uninfected during an epidemic mentioned on page 46. When *p*_*r*_ = 0 however, the entire population actually *will* become infected in the end (that is *s*_*e*_ = 1 when *p*_*r*_ = 0). For the model to be consistent with this, the coefficients 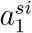 and 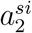 are subject to a constraint in that particular case. From (5.12) it follows that *η* = 0 when *p*_*r*_ = 0. For *s* = 1 to be a solution of (5.10) then requires:

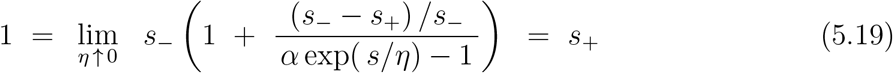

Substitution of (3.20) for *s*_+_ here yields, after some rearrangements, the following relation between 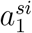 and 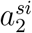:

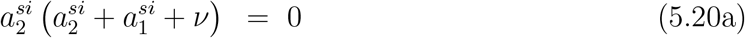

which implies:

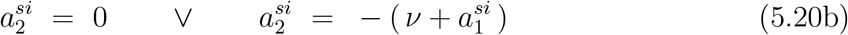

The case 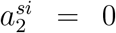 thereby corresponds to the standard SIR-model that we seek to replace by a more general approach in this paper. In contrast, the case 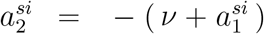 does relate to the generalisation of the SIR-model that accounts for the network structure of the population (albeit in the simplest approximation possible). In fact, the case 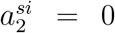, and therewith the standard SIR-model, can be seen as a special case of 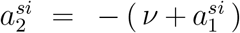 where 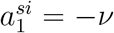 (also see footnote^5^). It is also worth mentioning that fulfilment of the requirement 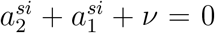 comes down to ⟨*s*_*si*_⟩ = 0 for *s* = *s*_*i*_ = 1, and is therefore in full agreement with the requirement that *s*_*e*_ = 1 for *p*_*r*_ = 0: only when *s* = 1 and the number of susceptible nodes in the network becomes zero, and therefore the entire population has been infected will the spread of the infection come to a halt. In fact, demanding that ⟨*s*_*si*_⟩ = 0 for *s* = 1, would as well have lead us, in a totally valid way, to (5.20b).

It is emphasized that the constraint 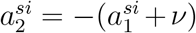 only applies in this particular form in case of an approximative approach where terms of order higher than 2 in the series expansion of ⟨*s*_*si*_⟩ have been truncated.

In the exact case, the demand that ⟨*s*_*si*_⟩ vanishes for *s* = 1 requires cancellation of all the expansion coefficients of the series, so that:

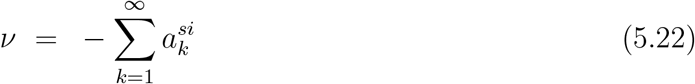

Based on the full series, and therefore exact, this relation holds in the most general sense. On every population network, no matter its structure, an epidemic will develop in accordance with this rule when *p*_*r*_ = 0, thus ensuring that ⟨*s*_*si*_⟩ = 0 for *s* = 1 and therefore *s*_*e*_ = 1.

In general, the coefficients 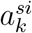 not only depend on the structure of the population network, but also on *p*_*r*_*/p*_*i*_. However, in some cases the dependence on *p*_*r*_*/p*_*i*_ is weak, and in cases where the social network of an individual/node consists of the entire population there is even no dependence on *p*_*r*_*/p*_*i*_ or the network structure at all. In the latter case, the active infections and the removed infections will be distributed randomly over the population, so that (irrespective of *p*_*r*_*/p*_*i*_) the standard SIR-model applies, in which 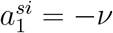 and 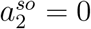.

The effect of *p*_*r*_*/p*_*i*_ ≠ 0 in a case where the coefficients 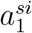 and 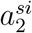 are constants independent of *p*_*r*_*/p*_*i*_ is illustrated in fig. 5.1. From (5.15b) it is clear that ∂*g*(*s*)/∂*s* (and therewith ∂*f* ^−1^(*s*)/∂*s* in the s-interval of relevance) decreases with increasing *p*_*r*_*/p*_*i*_ for given *s*_+_ and *s*_−_ (that is, for given 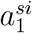 and 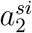). As a result, the intersection of *f* ^−1^(*s*) and the straight line *y* = *s* shifts towards lower s-values when *p*_*r*_*/p*_*i*_ increases, as can be seen in fig. 5.1. So *s*_*e*_ decreases with *p*_*i*_*/p*_*r*_ in those cases. Furthermore, as we will see next, it can even be shown now that also when 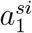 and 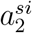 *do* depend on *p*_*r*_ and *p*_*i*_, the value of *s*_*e*_ actually becomes zero at a certain critical value of *p*_*r*_*/p*_*i*_.

Since *f* ^−1^(*s*) is a monotonously increasing, convex function on the *s*-interval of relevance, it is in fact obvious that for all values of *p*_*r*_*/p*_*i*_ for which:

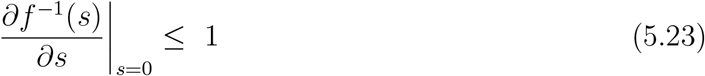

only *s* = 0 remains as a solution of (5.10), so that in all these cases *s*_*e*_ = 0.

Now, the derivative ∂*f* ^−1^(*s*)/∂*s* for *s* = *s*_0_ is readily obtained from (5.15b) as:

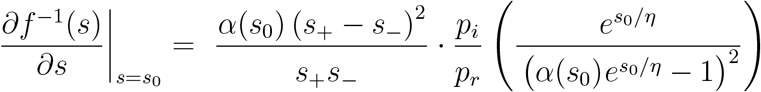

with *α* = *α*(*s*_0_) given by (5.11). Substitution of *s*_0_ = 0 and some rearrangements yields:

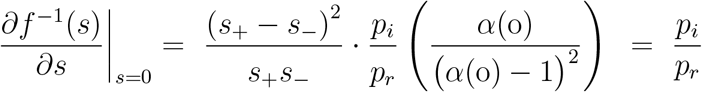

That is:

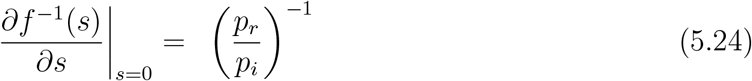

Combining (5.23) and (5.24) we see that a threshold *p*_*r*_*/p*_*i*_ = 1 exists which marks the boundary between a regime where *s*_*e*_ > 0 (when *p*_*r*_*/p*_*i*_ < 1) and a regime where *s*_*e*_ = 0 (when *p*_*r*_*/p*_*i*_ ≥ 1). It should be noticed that this threshold does not depend on 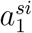 and 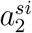 or on *ν*, and is therefore independent of the structure of the population network and thus quite general in nature (at least in the context of our truncated-series model). Related to this, there is a range of values of *p*_*r*_*/p*_*i*_ where *s*_*e*_ is always zero, given by *p*_*r*_*/p*_*i*_ ≥ 1, irrespective of whether 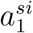 and 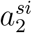 depend on *p*_*i*_ and *p*_*r*_ or not (although such a dependence may have an influence on the value of *s*_*e*_ itself when *s*_*e*_ > 0). As a corollary, when *p*_*r*_*/p*_*i*_ ≥ 1 a small limited number of infections cannot trigger (and develop into) an epidemic that involves large parts of the population. In essence, (5.23) and (5.24) represent the same result as that represented by (5.1). However, it is obtained here in a completely different way that strongly resembles the analysis of magnetic ordering in (ferro-) magnetic systems in terms of the Weiss molecular field theory [1]. As such, this approach anticipates the revelation of a striking analogy between a transition towards herd-immunity and thermodynamic phase transitions to be presented later on.

According to our analysis, an important general effect of infection removal seems to be a decrease of *s*_*e*_ to values less than 1: infection removal suppresses the propagation of infections through the population, to the extend that even part of the population will escape infection. A similar result has been obtained in the past within the context of the standard SIR-model as well. However, the present analysis not only shows that it also applies in a model where the structure of the population network is explicitly taken into account, thus making it a more general result, but also puts it on more solid mathematical foundations.

A legitimate question now, is whether saturation of the cumulative number of infections at a value *s* = *s*_*e*_ corresponds to a state of herd-immunity. The answer to that question largely depends on how we define herd-immunity. For instance, one might think of a state of herd-immunity in the broadest sense possible, namely as a situation in which an infection is unable to further propagate within a population. The answer to the aforementioned question would be affirmative in that case. Such a definition, however, is too general for practical use, as it leaves too much room for ambiguity and some quintessential issues unadressed. The whole point in this respect is that the value of *s*_*e*_ depends on the circumstances under which the infection spreads (that is, on both *p*_*r*_ and *p*_*i*_, as well as on the structure of the population network, which translates into the 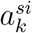). As a result of this, there are serious pitfalls when it comes to rolling back measures aimed at preventing the spread of an infection, especially when the arguments to do so are based on positive expectations concerning the achievement of herd-immunity, however without an adequate notion/definition of its concept. To demonstrate this, consider a situation in which a clear tendency towards saturation of *s* at a particular value *s*_*e*_ is observed after an infection has been spreading for a while under a regime of restrictive social measures. The effect of such measures is twofold thereby: they reduce *p*_*i*_ and they restrict the size of the social networks of individual members of the population, thus affecting (lowering) the value of ⟨*s*_*si*_⟩ at given *s*. When the epidemic has already reached a stage where *s* makes an approach to its asymptotic value *s*_*e*_, the number of active infections *s*_*i*_ may still not be zero but is already over its peak, and therefore :

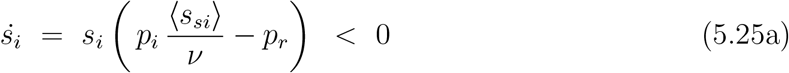

Rolling back social measures at this stage (for instance on the basis of an (inadequate) judgement regarding the achievement of herd-immunity) means that a new regime is entered however, which replaces *p*_*i*_ by 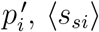, ⟨*s*_*si*_⟩ by 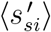 and *ν* by *ν*′, so that the rate of change of *s*_*i*_ now becomes:

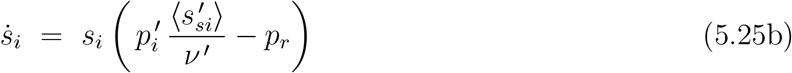

where it should be noted that 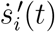 is equal to *s*_*i*_(*t*) at the moment *t* = *t*_0_ when measures are rolled back (that is: 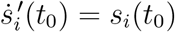. The problem here is that it is not at all certain that 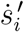 is negative as well (like 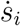). Depending on 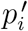 and 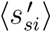 it cannot even be excluded a priori that the term in brackets in (5.25b) is actually positive and consequently 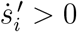. In that case, the spread of the infection wile intensify again into a new wave of (active) infections, which will only attenuate after a new maximum in the number of active infections 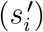 has been reached. Such a scenario is illustrated in fig 5.2, where the results are shown for a simulation where a less tight regime of “social measures” follows upon a significantly more restrictive regime. The spread of the infections starts under a tight regime of social restrictions, in which the contacts of a particular (central) node are selected from a relatively small 2*N* + 2 × 2*N* + 1 square of nodes closest to (i.e. surrounding) the central node with *N* = 1 (the number of contacts to a single node thus being equal to *ν* = 8). The spread of the infection was simulated under this regime to a point where the epidemic had *nearly* come to a halt (i.e. *s* only weakly increasing with time, and *s*_*i*_ almost zero). Then (at *t* = 500) a new, less strict, regime with *N* = 5 was introduced (so that the number of contacts per node increased to *ν* = 24), under which the few active infections left from the 1st regime/wave were given the opportunity to pass on their infection to the remaining susceptibles, thus restarting the epidemic. Fig. 5.2 clearly reflects this. After a near fade-out of the active infections after the 1st regime it takes only a little while after the implementation of the 2nd regime for the epidemic to regain strength, and very soon both the active and cumulative infection rates are clearly on the rise again. The result is that we are confronted with 2 subsequent waves of infections: one rather modest under a regime of tight social measures (*N* = 1) that seems to fade out after a while, and a 2nd one of a much fiercer intensity after a partial roll-back of the measures (*N* = 5), by which almost all members of the population that remained uninfected after the 1st wave become infected in the end. It is for the possibility of scenarios of this kind alone that some doubts are justified about strategies for coping with an epidemic based on the (assumed) achievement of herd-immunity under a regime of (limited) social measures (which, however, is just what a country like Sweden openly advocated and put into practice during the initial months of the Covid-19 pandemic). It is also clear that a better understanding (coupled to a tighter definition) of the concept of herd-immunity is necessary to make it of (safer) practical use.

**Fig. 5.2:**
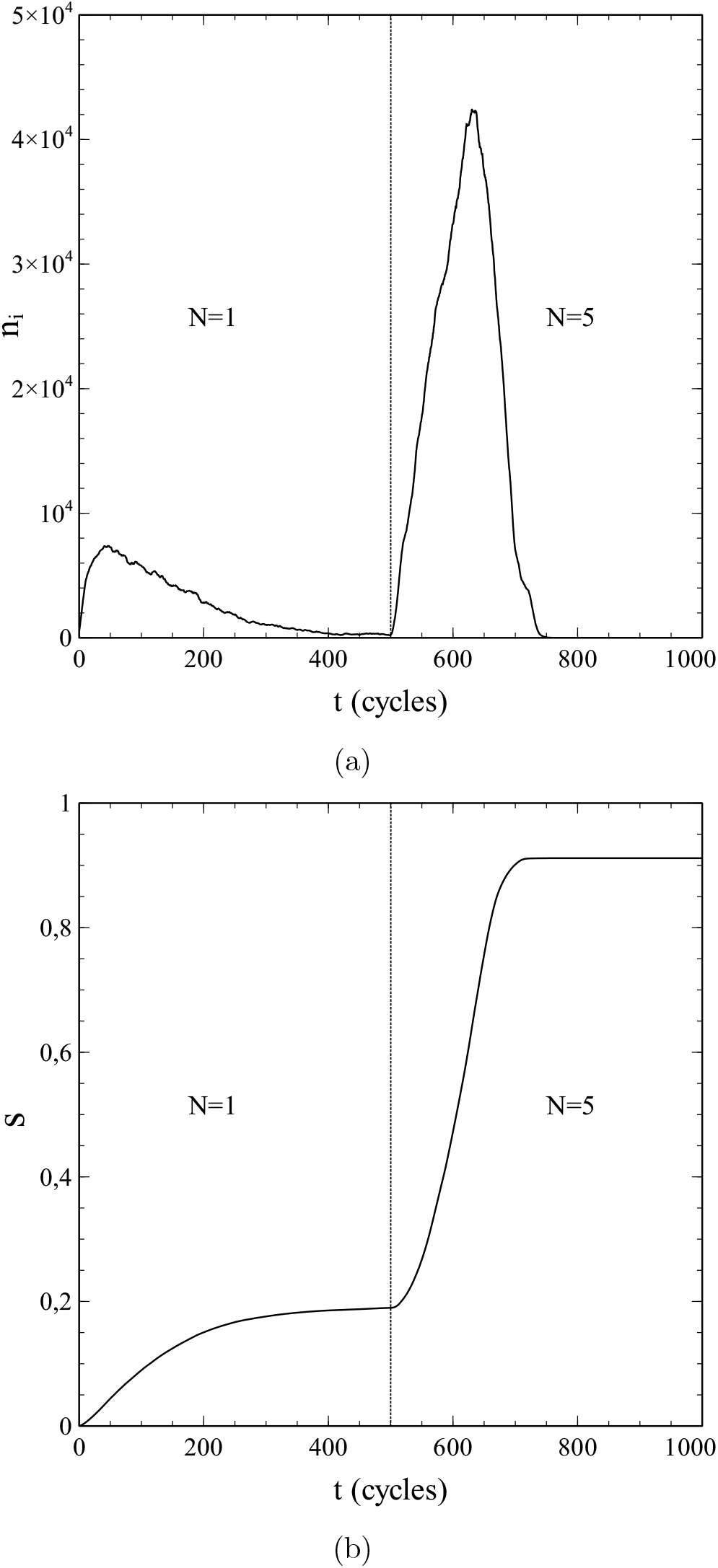
Simulated sequence of infection waves under 2 different regimes of social measures (see main text). **a**) Number of active infections *n*_*i*_ as a function of time (time measured in simulation cycles, i.e. the time in which (on average) each member of the population (node) makes exactly 1 contact). **b**) cumulative infection rate *s* as a function of time. Dotted lines represent the cross-over of social regimes. Parameters: *p*_*i*_ = 0.5, *p*_*r*_ = 0.325, *N*_0_ = 500, population size *N* = 1501^2^.

### d) Reconsidering the meaning of reproduction numbers

It may be clear that, in view of the results presented in this chapter, the often quoted condition *R*_0_ > 1 for an epidemic to get started requires some reconsideration. Via a similar line of though as the one followed in section 5a, both the standard SIR-model and the extended SIR-model presented in this paper lead to the criterion *p*_*i*_*/p*_*r*_ > 1 for an epidemic to evolve from a small number of active infections in an otherwise fully susceptible population. With *R*_0_ given by (2.5), this criterion is equivalent to *R*_0_*/Q*_0_ > 1 rather than *R*_0_ > 1. The criterion *R*_0_ > 1 can only be preserved (as is often done by the way) via the introduction of a rather crude approximation that puts *R*_0_ ≡ *p*_*i*_*/p*_*r*_ (and thus implicitly takes *Q*_0_ = 1). The rationale here is that *τ* = 1*/p*_*r*_ is the average lifetime of an active infection in case of an exponential decay as given by (2.1)^6^. Taking ⟨*s*_*si*_⟩ = constant = *ν*, the number of new infections per unit time due to a single active infection can be taken as a constant as well, which is equal to *p* = *p*_*i*_ so that the total number of new infections due to a single active infection becomes 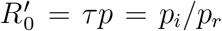 (variations on this simplified scheme exist [2,3] for the purpose of generalisation, but the basic ideas underlying them are the same). It is clear that such an approach basically comes down to substitution of ⟨*s*_*si*_⟩ = *ν* into (2.3) and taking *t*_0_ = 0 :

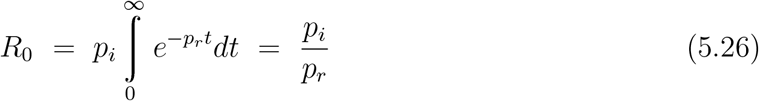

in which case the factor *Q*_0_, accounting for the *s*-dependence of ⟨*s*_*si*_⟩, becomes indeed unity and 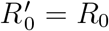. However, in general 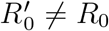 and 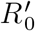 is therefore *not* a reproduction number, but rather an *effective* reproduction number at best (whatever the deeper physical meaning of such a qualification may be). Similar considerations apply to *R* and the often mentioned criterion *R* > 1 for the number of active infections to be on the rise, as we will see at the end of the next chapter on herd-immunity. As such, the interpretation of the dynamics of epidemic outbreaks in terms of reproduction numbers is not entirely unproblematic.

## 6. Defining and understanding herd immunity

### a) Fundamentals

The aim is now to present a strict mathematical definition of herd immunity, in such a way that the result is not only rooted in the basic physical and mathematical principles of epidemic growth, but also makes sense from a practicle point of view.

For this purpose, we consider an epidemic that has been going on for a while under a regime of social and protective measures, such that at some moment *t*_0_ in time, the cumulative number of infections is *s* = *s*_0_ > 0 and the number of active infections *s*_*i*_ = *s*_*i*0_ > 0. Furthermore:

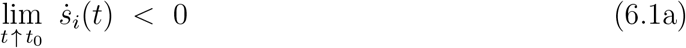

so that the number of active infections is over its peak and declining immediately before *t* = *t*_0_. The average number of s-nodes linked to an i-node immediately before *t*_0_ is given by:

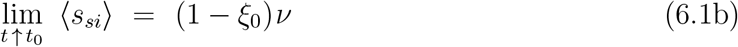

At *t*_0_ a new regime is entered, whether by rolling back of the measures taken, or by some changes in the properties (for instance the transmissibility) of the pathogen that causes the infection. The structure of the population network thus may change, so that *ν* has to be replaced by *ν*′. In addition, possible changes in the protective measures require the replacement of *p*_*i*_ by 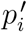. Changes in the the population network also imply the replacement of ⟨*s*_*si*_⟩ by 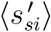 and of 1 − *ξ*_0_ by *λ*′(1 − *ξ*_0_). The parameter *λ*′ thereby accounts for changes in the structure and the topology of the population network and matches the value of 1 − *ξ*_0_ to the new network: 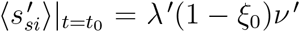. We finally consider 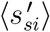 as an expansion around *s* = *s*_0_:

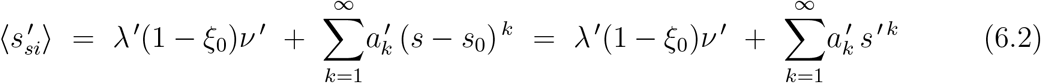

where *s*′ = *s* − *s*_0_.

Substitution of (6.2) into (5.25b) yields for the new regime (*t* ≥ *t*_0_):

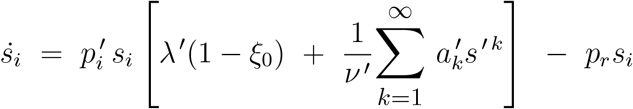

That is:

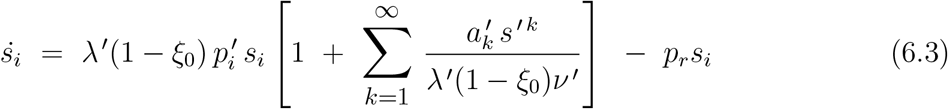

which is, in effect, identical to the expression for 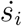 in the case of an infection with transmission probability 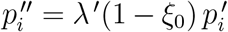 spreading over a network on which the number of nodes linked to a central node is *ν* ^′′^ = *λ*′(1 − *ξ*_0_)*ν*′ (instead of *ν*′). Upon truncating terms of order *k* > 2 in the series expansion in (6.3) we have:

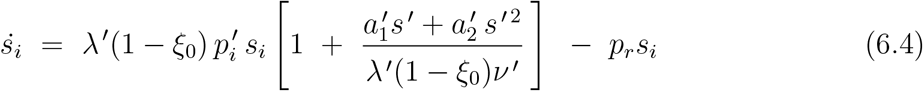

Now, like *s*, the variable *s*′ is a state variable, so that 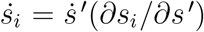. Therefore, upon making the identifications 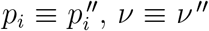 and *s* ≡ *s*′ in (1.42a,b), the differential equation (6.4) can be dealt with in the same manner in which the differential equation of of identical form given by (1.42a) was dealt with in section 3c for the case of a 2nd-order polynomial approximation of ⟨*s*_*si*_⟩. We thus obtain, straightforwardly from (3.18):

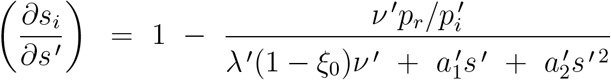

By analogy with (3.19), this can be reexpressed as:

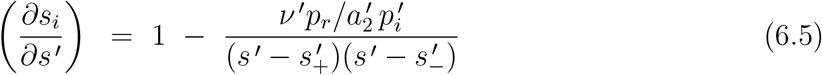

where (see (3.20)):

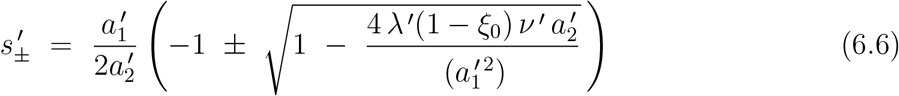

Via some minor rearrangements of (6.5) we obtain the following ODE (compare to (3.21)):

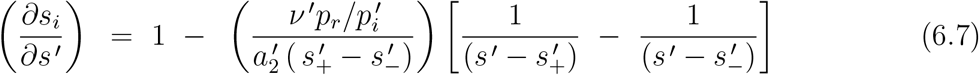

the solutions of which are given by:

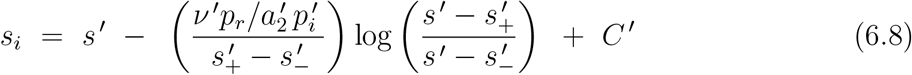

The constant of integration *C*′ follows from the condition 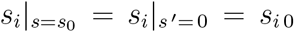 (remember *s*′ = *s* − *s*_0_):

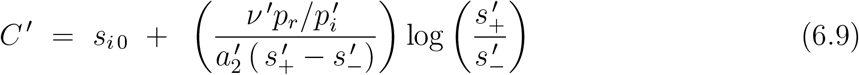

Combining (6.8) and (6.9) we finally obtain:

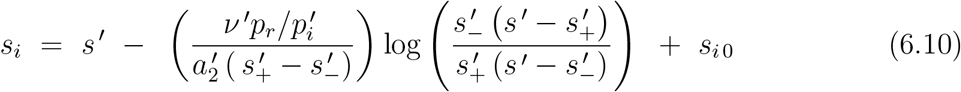

The logarithmic term here on the right-hand side has a clear physical meaning that will prove key to the definition and understanding of herd-immunity. Calling this term *s*_*L*_ we can write (6.10) as:

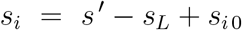

With *s*′ = *s* − *s*_0_ and *s*_*i* 0_ = *s*_0_ − *s*_*r* 0_ that is:

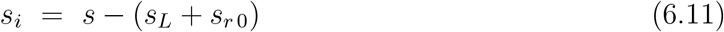

where *s*_*r* 0_ = *n*_*r* 0_*/n* relates to the total number of removed infections at *t* = *t*_0_ (i.e. when *s* = *s*_0_, *s*′ = 0). In general *s*_*i*_(*t*) = *s*(*t*) − *s*_*r*_(*t*), so that (6.11) implies:

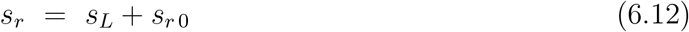

With the given definition of *s*_*r* 0_ it is thus shown that the term:

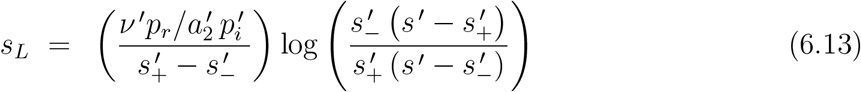

represents the infections removed under the *new* regime (that is, *after t* = *t*_0_). In view of this, an additional quantity of physical relevance becomes evident as well, namely:

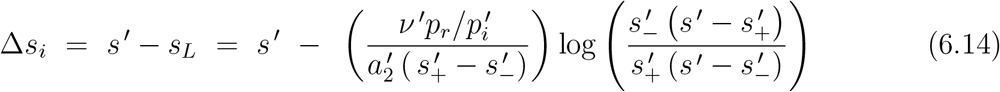

representing, for given *s*′(*t*), the *nett* change in the active-infection number since the new regime took effect at *t* = *t*_0_.

The condition:

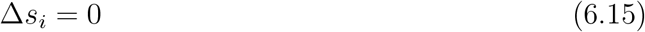

makes an important physical criterion. As stated earlier, we assume a situation where the number of active infections was in (sharp) decline under the old regime prior to *t* = *t*_0_. For that to remain the case under the new regime it is required that Δ*s*_*i*_ < 0 for *all s*′ > 0 (all *t* > *t*_0_). In contrast, when there is an interval 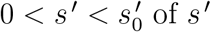 of *s*′-values for which Δ*s*_*i*_ > 0, the number of active infections will initially rise again (maybe even strongly) after the new regime has come into effect. It may be obvious that such a situation is at variance with what one would intuitively think of as a state of herd-immunity. However, a completely different situation occurs when for *all* conceivable regimes to come into effect at *t* = *t*_0_ (most importantly the regime of social normality) Δ*s*_*i*_ is *negative definite* for *s*′ > 0 (that is, Δ*s*_*i*_ < 0 for all *s*′ > 0). A necessary and sufficient condition for such a situation to occur is that solving Δ*s*_*i*_ = 0 for *s*′ yields *s*′ = 0 as the only solution for *any* conceivable regime. Although there will even be new infections after *t* = *t*_0_ in such a case, the total number of *active* infections meanwhile can do nothing then but decrease with *t* and *s*′, and the epidemic is inevitably in state of decline and fading out.

To substantiate these viewpoints mathematically we define, by analogy with (5.9):

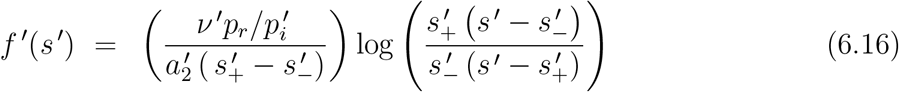

The condition Δ*s*_*i*_ = 0 can then be recast into the form (see (6.14)):

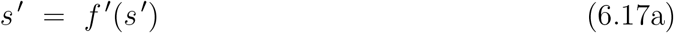

or, equivalently, into:

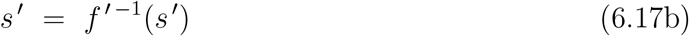

where *f*′ ^−1^(*s*′) represents the inverse of *f*′(*s*′) which, being the analogue of *f* ^−1^(*s*) in (5.10), is readily obtained, with 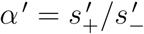 taken as the analogue of *α* (see (5.11)), as:

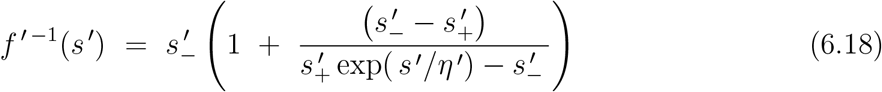

where:

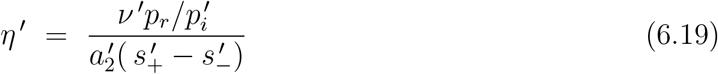

Being identical in their mathematical form, the functions *f*′ ^−1^(*s*′) and *f* ^−1^(*s*) behave in a qualitatively similar way in relation to their respective arguments *s*′ and *s*. Hence, the line of thought followed in section 3c in connection with *f* ^−1^(*s*) applies to *f*′ ^−1^(*s*′) as well. As such, we find equations (6.17a) and (6.17b) to have 2 solutions when ∂*f*′ ^−1^(*s*′)/∂*s*′ |_*s*′=0_ > 1, one of them being *s*′ = 0 and the other one given by the intersection of the line *y* = *s*′. When ∂*f*′ ^−1^(*s*′)/∂*s*′ |_*s*′=0_ ≤ 1 however, only the solution *s*′ = 0 remains and Δ*s*_*i*_ < 0 for *s*′ > 0. When the latter is the case for any regime of social measures, the epidemic is in a stage of inevitable fade-out.

By analogy with (5.15a), the derivative ∂*f*′ ^−1^(*s*′)/∂*s*′ is obtained as:

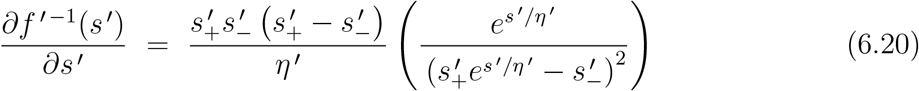

Substitution of (6.19) for *η*′ here, while also using 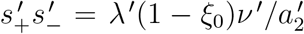 (to be obtained straightforwardly on the basis of (6.6)), then yields:

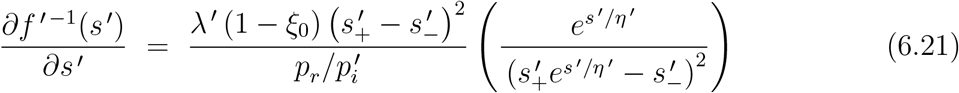

For *s*′ = 0 that is:

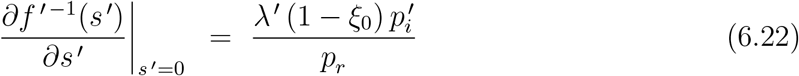

The condition for the epidemic to remain fading out after *t* = *t*_0_ (*s*′ > 0), also under the new regime, becomes therewith:

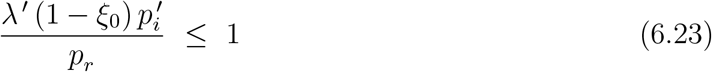

That is:

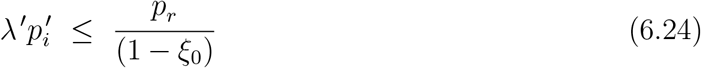

In every new regime for which the product 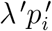 meets this (in)equality, the number of active infections will be subject to a monotonous decrease after *t* = *t*_0_. It is noteworthy that requiring 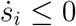 for *s*′ = 0 on the basis of (6.3):

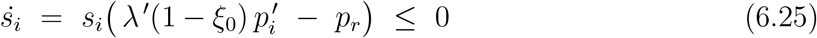

directly leads us to (6.23) and (6.24) as well. However, this procedure leaves us with no clue as to whether Δ*s*_*i*_ is negative definite or not for *s*′ > 0, and therefore does not exclude the possibility, as observed in fig. 5.2, that for some *s*′ the number of active infections will start to rise again (even under the same regime of measures).

If (6.24) also holds for the regime of social normality then lifting restrictive and protective measures is safe, in the sense that it will not lead to a new wave of infections: the rate at which 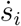 decreases may be less than in a regime with measures in place, but the epidemic will continue to fade-out until the last active infections disappear and *s*_*i*_ becomes zero. One could therefore say that as soon as (6.17a,b) apply, a form of herd-immunity has been achieved, despite the fact that new infections will continue to emerge until *s*_*i*_ = 0, albeit at an increasingly lower rate as *s*′ increases. New infections will cease to emerge as soon as *s*_*i*_ = 0. That is, when (see (6.10)):

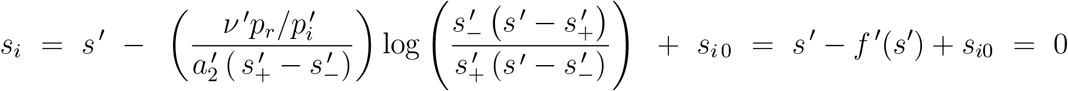

and thus when:

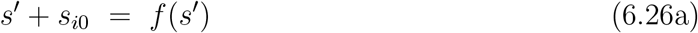

which is equivalent to:

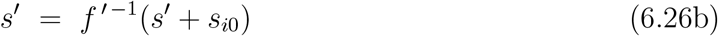

The solution 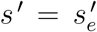 of (6.26a) and (6.26b) relates to *s*_*e*_ in this via 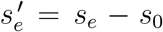, and has to be obtained numerically. The value of *s*_*e*_ = *s*_0_ + *s*′ represents the final cumulative infection rate reached when the epidemic comes to an end. Since at the end of an epidemic all active infections have been removed, it also represents the final rate of removed infections to be reached, i.e. when *s* = *s*_*e*_ = *s*_0_ + *s*′ then *s*_*r*_ = *s*_*e*_.

Now that we have captured the criterion for the end of an epidemic in the mathematical form of (6.26a,b), the relevant question is how “robust” the resulting epidemic state after reaching *s* = *s*_*e*_ is against an influx of new (active) infections from outside of the population, for instance via infected travellers (from outside), or infective population members (re)entering from abroad. In other words: will a new wave of infections start or not, once a (very) small but not insignificant number of new active infections from outside has been introduced into the population?

To answer this question, let *ν*′(1 − *ξ*_*e*_) be the average number of s-nodes linked to an r-node when the epidemic has come to a halt:

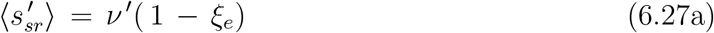

Using the symmetry relation 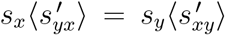 we then get for this case (for which 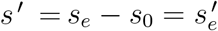):

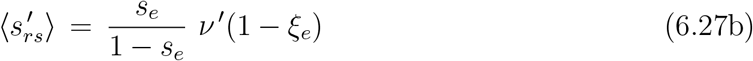

and via (1.17) and (1.18), which reduce to:

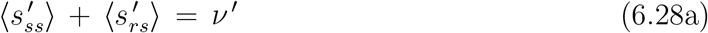

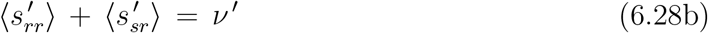

we thus obtain:

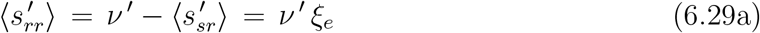

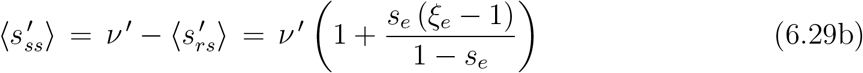

for this case. To distinguish the average coordinations ⟨*s*_*xy*_⟩ and 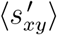 in the earlier wave(s) from those in the new wave, we write the latter as ⟨*σ*_*xy*_⟩ and the coefficients of their corresponding series expansions as *α*_*k*_ (instead of *a*_*k*_). Similarly, we write the cumulative infection rate and the rate of active infections in the new wave respectively as *σ* and *σ*_*i*_. We assume the initial number of new active infections that form the precursor to a possible new wave of infections to be very low (*σ*_0_ = *σ*_*i*0_ << 1, or even *σ*_0_ = *σ*_*i*0_ ≈ 0). Since we also assume the removed infections to have full immunity, a possible new wave of infections will spread exclusively among those members of the population (nodes) that remained uninfected (i.e. susceptible) during the earlier wave(s). Hence, since each newly introduced active infection is considered to replace a susceptible node at random (so that 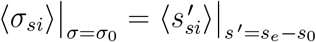) and because *σ*_0_ is considered small enough that to take *σ*_0_ ≈ 0:

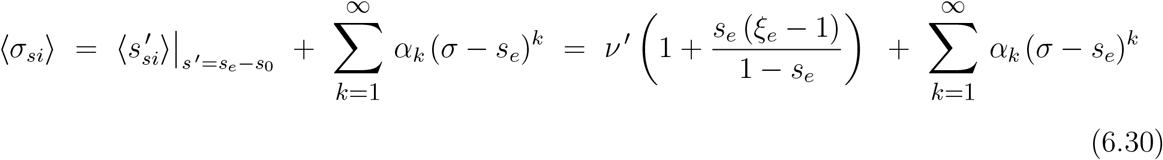

We consider no changes in social measures taken after the first appearance of the new active infections, so that the typical rate of transmission remains 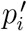, the rate of change of *σ*_*i*_ in the new wave therewith becomes:

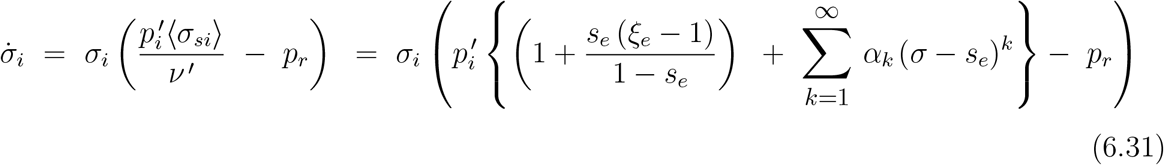

However, a new wave of infections due to a small (almost negligible) number of initial infections *σ*_0_ << 1 will start *only* if 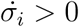 for *s*′ = *s*_*e*_ (i.e. for *σ* = *σ*_0_ ≈ 0).

From (6.31) it is easily inferred that a new wave will therefore *not* emerge when:

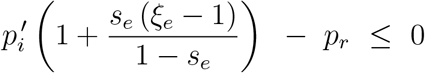

That is, when:

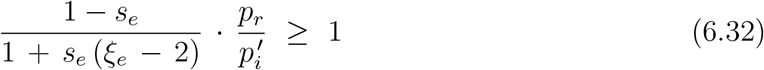

which thus provides us with a proper criterion for “true” herd-immunity that is not only rooted in the basic mechanisms and mathematics of epidemic growth but also connects with our intuitive conception of the phenomenon.

An important insight that immediately follows from (6.32) is that apparently the structure and topology of the population network *do* have an influence in the process of achieving herd-immunity, in contrast to what we have seen earlier in connection with the criterion (expressed by (5.1) and (5.23),(5.24)) for an epidemic to develop from a few initial infections in case of a fully susceptible population (*s*_*s*_ = 1). Key for this observation is the dependence of *s*_*e*_ and *ξ*_*e*_ on the structure of the social networks. Network correlations *explicitly* make their way into the process via *ξ*_*e*_ (whereas they affect the value of *s*_*e*_ in a more implicite manner). These correlations are a typical artefact of network structure of the population and also the percolative nature of the spread of an infection on a population network. Their influence can be understood as follows. We write, for every *s* ≤ *s*_*e*_:

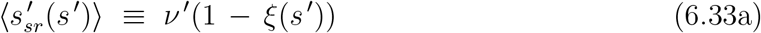

In case of a fully random distribution of the susceptible individuals/nodes over the network 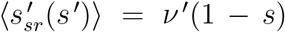. We may therefore write:

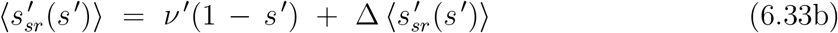

where 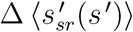 accounts for the correlations (i.e. the deviations from the random distribution). Combining (6.33a) and (6.33b) we have:

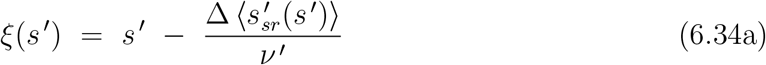

With 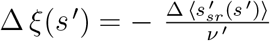 that is:

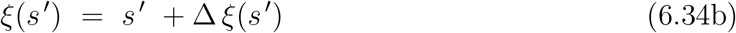

so that *ξ*_*e*_ = *ξ*(*s*_*e*_) = *s*_*e*_ + Δ *ξ*_*e*_.

We thus obtain:

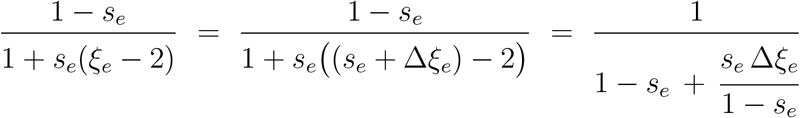

Substitution of which into (6.32) yields:

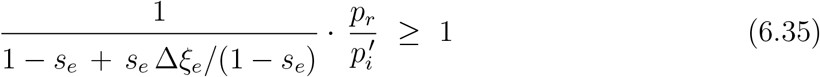

We see that the network correlations explicitly enter this (in)equality on the left-hand side via Δ*ξ*_*e*_. The sign of Δ*ξ*_*e*_ is indicative of whether such correlations support (Δ*ξ*_*e*_ < 0) or counteract (Δ*ξ*_*e*_ > 0) the achievement of herd-immunity. A qualitative argument for the sign of Δ*ξ*_*e*_ in general can be given by considering the influence of correlations on the number of s-r pairs and 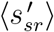.

In the absence of correlations (fully random distribution of r-nodes) the number of s-r pairs for *s*′ = *s*_*e*_ − *s*_0_ is (see (1.14)): 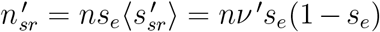, so that 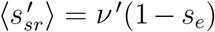. However, due to the percolative nature of the spread of the infection, the r-nodes along its paths are not randomly distributed but form dendritic structures (“trees”). With 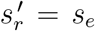 the following inequality then applies to the number of r-r pairs for *s*′ = *s*_*e*_ − *s*_0_:

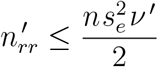

where the equal sign relates to a random distribution of removed infections (each removed infection having *s*_*e*_ *ν* ^′*l*^ other removed infections in its social network), and the inequality applies in case of the (correlated) dendritic structures (the division by 2 corrects for double counting removed infections). With 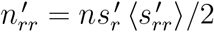 (see (1.15)) we thus obtain (remember 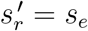):

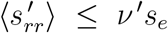

and with 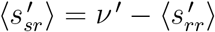 (see (6.28b):

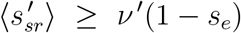

Via 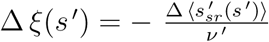and (6.33b) this (in)equality can be transformed into:

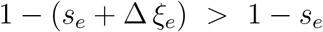

in the presence of correlations (i.e. in cases where Δ *ξ*_*e*_ ≠0). That is:

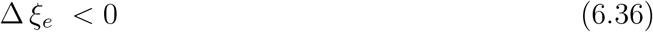

We thus find out that network correlations by themselves *always* contribute to the achievement of herd-immunity in a positive way: a negative Δ *ξ*_*e*_ increases the value of left-hand side of (6.35) with respect to its value for Δ *ξ*_*e*_ = 0 (i.e. for a random distribution without network correlations). It is emphasized however, that network correlations are a contributing factor, rather than a necessary requirement for herd-immunity. After all, is clear that even in the absence of correlations (Δ *ξ*_*e*_ = 0), the sheer increase in the cumulative-infection number *s*_*e*_ alone already leads to an increase in the left-hand side of (6.35). However, indicative of the role of correlations, (6.35) and (6.36) indirectly emphasize the role of percolation effects as well in the establishment of herd-immunity, given the close relation between network correlations and percolation.

### b) Different types of herd-immunity: a classification

It is worth noticing that a distinction can be made on the basis of (6.24) and (6.32) between different types of herd-immunity, all of which having clear, but different, practical implications. First of all, it is important whether (6.24) and (6.32) only hold in regimes of strict social measures, or in every conceivable regime of social measures (including the regime of social normality). In the first case we can speak of “weak” herd-immunity, as opposed to “strong” herd-immunity in the second case. Weak herd-immunity is a contextual phenomenon, and it occurs only by virtue of the restrictions imposed upon the population by a regime of sufficiently strict social measures. There is no guarantee that the population is safe from a restart of the epidemic as soon as social restrictions are (partially) lifted. Only strong herd-immunity can offer such a guarantee. In fact, weak herd-immunity is not what we intuitively associate with herd-immunity, only strong herd-immunity does. However, for the sake of clarity, making a distinction between the 2 forms has its benefits.

Another relevant distinction can be made on the basis of something that we might call the “degree” of herd-immunity. As soon as (6.24) is met, the epidemic is in a phase of inevitable fade-out under the imposed regime of social measures. However, the generation of new infections has still *not* come to a halt (which may still put a burden on the health system for instance). Nevertheless, the risk of the epidemic growing out of control has disappeared. We might call this situation a state of “1st degree herd-immunity”. Such a situation may precede a state of “2nd degree herd-immunity”, which is entered when the number of active infections actually becomes zero *and* the resulting state is one in accordance with (6.32), i.e. a state where, although (very) small pockets of new infections may (re)appear, the population *at large* is “immune” against the build-up of a new wave of infections, at least under the imposed regime of restrictive social measures.

It should be clear that states of weak herd-immunity are deceptive, irrespective of their degree. Although the number of infections may be declining (1st degree) or has come to a fade-out (2nd degree), the risk of a renewed increase in the active infection rate after lifting the social measures is real. A state of weak herd-immunity may therefore be the aim of temporary measures to lift the burden on the health system, but its achievement should by no means taken as a motivation to return to normality. Only when the number of active infections is in decline in a case of strong herd-immunity (of 1st degree) such a return is safe. By definition, the end of the battle against an epidemic outbreak is then marked by a state of strong herd-immunity of 2nd degree: not only has the active infection rate faded-out in such a case, but the population is immune, even under a regime of social normality, against new waves of infections arising out of small contingents of initial infections. Telling the difference between states of weak and strong herd-immunity may not be easy in practice however, especially when the pathogen is (relatively) new and its properties (*p*_*i*_ for instance) insufficiently known.

### c) Herd-immunity and reproduction numbers

In the literature, conditions for herd-immunity and the herd-immunity threshold are often expressed in terms of reproduction numbers. Considering the critical remarks made in chapter 2 and section 5d with respect to the practical use of reproduction numbers, combined with the confusion about the definition of herd-immunity in the literature, a short regression into this subject seems more than appropriate.

Very often, a state of herd-immunity is defined as a state where the number of active infections has reached its peak and is in a state of decline. It may be clear that this is actually what is called a state of 1st-degree herd-immunity according to the classification in the previous section, as opposed to a state of 2nd-degree herd-immunity to be reached when the active infection rate has finally faded-out to zero. The condition for such a state to occur is (when *s*_*i*_ ≠ 0):

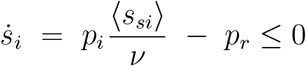

That is:

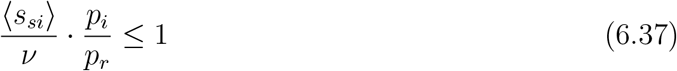

Note that ⟨*s*_*si*_⟩ = ⟨*s*_*si*_(*t*)⟩. We define:

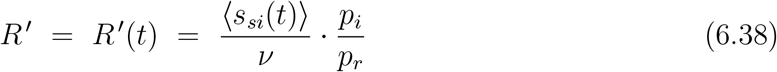

which we reexpress as:

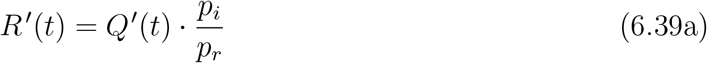

where:

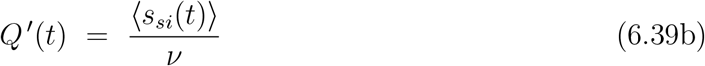

so that the requirement for 1st-degree herd-immunity (6.37) can be rewritten as:

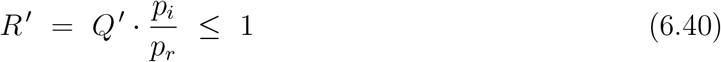

The similarity between (6.39a) and (2.4) is obvious. However, *Q*′ ≠ *Q* in general (see (2.3)), so that *R*′ ≠ *R*. Therefore *R*′ is *not* a reproduction number.

Combining (2.3) and (6.39b) we get:

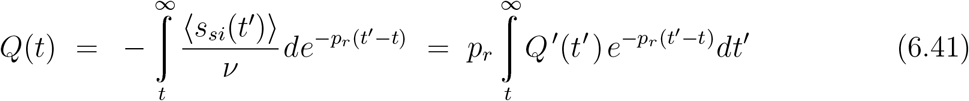

We introduce *Q*_*m*_(*t*):

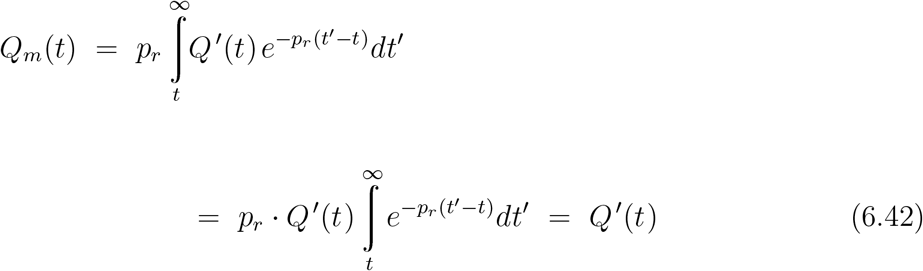

Since *Q*′ (*t*) ≥ *Q*′ (*t*′) for *t*′ ≥ *t* (because ⟨*s*_*si*_(*t*)⟩ ≥ ⟨*s*_*si*_(*t*′)) for *t*′ ≥ *t*), it is easy to see that *Q*_*m*_(*t*) ≥ *Q*(*t*). By combining this result and (6.42) we thus obtain:

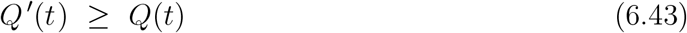

Hence (see (2.4) and (6.39a)), for *all t* > 0 : *R*′ ≥ *R*. So, when the criterion *R*′ ≤ 1 for 1st-degree herd-immunity is met, the criterion *R* ≤ 1 is also met. Vice versa however, *R* ≤ 1 is *not* a sufficient condition for *R*′ ≤ 1. Therefore, *R* ≤ 1 is not a valid criterion for 1st-degree herd-immunity.

## 7. Vaccination

A key lesson from the previous sections is that the pursuit of herd-immunity by allowing the infection to spread through the population may possibly run into serious difficulties. When the infection is allowed to spread under a (strict) regime of social measures there is a risk that the population ends up in a state of apparent herd-immunity that turns out to be false as soon as social restrictions are lifted and the infection rate starts to rise steeply again. On the other hand, letting the infection spread under a normal social regime does lead to herd-immunity, but only at the cost of a very large number of infections, which is unacceptable especially in those cases where the infection is of a kind that causes serious health issues. Therefore, the only way to achieve herd-immunity in a manner that is safe under all circumstances is vaccination.

To describe the effects of large scale vaccination on the susceptibility of a population to epidemic spreading of an infection, we introduce the effectiveness ϵof a vaccine, being the relative decrease of the transmission probability *w*_*i*_ or, equivalently, the relative reduction of the transmission constant *p*_*i*_ = 2*w*_*i*_. The constant of transmission 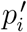 from an active infection to a vaccinated individual is thereby related to the transmission constant *p*_*i*_ from an active infection to an unvaccinated (fully susceptible) individual via:

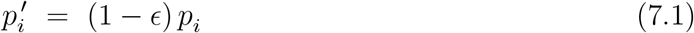

The lesser the protection offered by a vaccine, the lower the value of for that particular vaccine: by definition 0 ≤ ϵ≤ 1, where ϵ= 1 corresponds to a 100% effective vaccine that gives full protection (immunity), whereas ϵ= 0 effectively relates to a case without any vaccination, or to a totally inactive vaccine.

Now, let ⟨*σ*_*si*_⟩ and 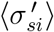 respectively be the average number of unvaccinated nodes and the number of vaccinated nodes linked to an active infection. The rate of change *s*_*i*_ of the active infections in case of a (partially) vaccinated population can then be expressed as:

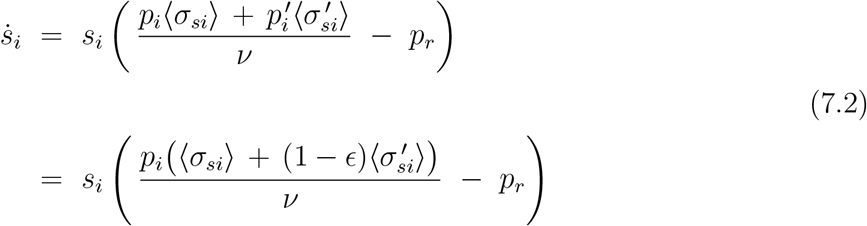

The vaccinated population is assumed to be free from active and removed infections (*s*_*i*_ = *s* = 0) prior to *t* = 0, when a very small number (*s*_*i*_ ≪ 1) of new active infections, randomly distributed over the population, appears.

We write:

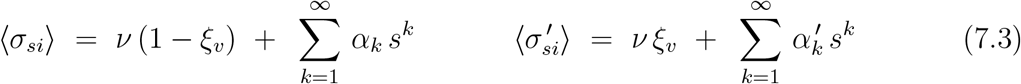

Here *ξ*_*v*_ accounts for the reduction of ⟨*σ*_*si*_⟩ and 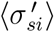at *t* = 0 (*s* = 0) due to vaccination (compare to *ξ*_0_, *ξ*_*e*_ and *ξ*(*s*) in chapter 6). Mathematically, nodes representing a vaccinated individual are equivalent to those representing a removed infection. Substitution of (7.3) into (7.2) yields:

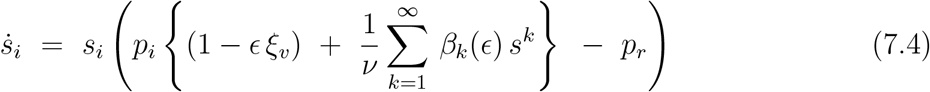

where:

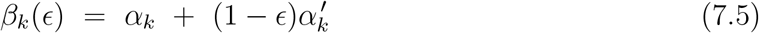

The condition 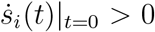 (or equivalently 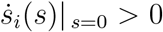 for an epidemic to develop from a few initial infections then implicitely leads to (compare to sections 5a and 5b):

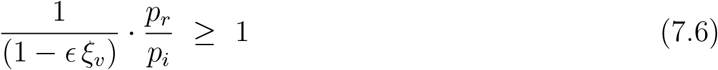

as a criterion for vaccine-acquired herd-immunity. Possible (network) correlations between vaccinated individuals enter the criterion via *ξ*_*v*_. Such correlations may arise (in theory) when the vaccination is carried out according to a non-random scheme. Such a situation seems quite unusual however. We therefore assume that the members of the population are vaccinated at random so that *ξ*_*v*_ = *s*_*v*_, where *s*_*v*_ is the vaccination rate (fraction of the population vaccinated). In that case (7.6) becomes:

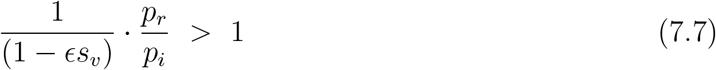

The product ϵ *s*_*v*_ *≡𝔰*_*v*_ can be considered as an effective vaccination rate: vaccinating a fraction *s*_*v*_ of the population with a vaccine having an effectiveness of ϵ< 1 is equivalent to vaccinating a population-fraction 𝔰_*v*_ = ϵ*s*_*v*_ with a vaccine having an effectiveness ϵ= 1. The critical vaccination rate marking the herd-immunity threshold is now straightforwardly obtained from (7.7) as:

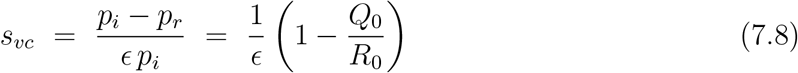

where *R*_0_ = *Q*_0_ · *p*_*i*_*/p*_*r*_ is the basic reproduction number, with *Q*_0_ accounting for the *s*-dependence of ⟨*s*_*si*_⟩ (see chapter 2). In the literature, the herd-immunity threshold is often given as *s*_*vc*_ = 1 − 1*/R*_0_ [see [1] for instance]. This is, strictly speaking, incorrect. Expressions of that form relate to an incorrect/incomplete expression for *R*_0_ or, at best, to an approximation for *R*_0_ (see section 5d), where the *s*-dependence of ⟨*s*_*si*_⟩ is ignored or neglected (leading to *R*_0_ = *p*_*i*_*/p*_*r*_).

The result (7.8) can be reexpressed in terms of the critical *effective* vaccination rate 𝔰_*vc*_ as:

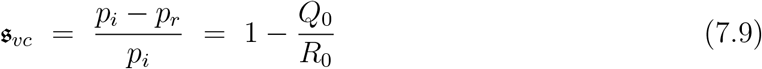

Vaccination-acquired herd-immunity is obtained when the vaccination rate is equal to or larger than the critical vaccination rate, which is equivalent to the *effective* vaccination rate 𝔰_*v*_ = ϵ*s*_*v*_ being equal to or larger than the critical *effective* vaccination rate 𝔰_*vc*_:

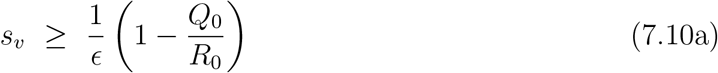

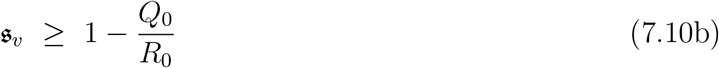

Since 0 ≤ ϵ≤ 1 *and* 0 ≤ *s*_*v*_ ≤ 1 it is easy to see that 0 ≤ *𝔰*_*v*_ ≤ 1. The critical effective vaccination rate 𝔰_*vc*_ = 1 − *Q*_0_*/R*_0_ is the lowest value of 𝔰_*v*_ for which herd-immunity *is* obtained for given *p*_*i*_*/p*_*r*_ = *R*_0_*/Q*_0_. However, from (7.10a) it is evident that its value also equals the lowest value of for which (by vaccinating the entire population so that *s*_*v*_ = 1) herd-immunity *can be* obtained at given *p*_*i*_*/p*_*r*_ (lower values of do not allow for herd-immunity to be obtained for the value of *p*_*i*_*/p*_*r*_ involved, since they would require *s*_*v*_ > 1). We can therefore combine 2 “phase diagrams” into a single figure. Fig. 7.1a. shows the relevant combinations of *p*_*i*_*/p*_*r*_ and 𝔰_*v*_, *as well as* the combinations of *p*_*i*_*/p*_*r*_ and, for which vaccine-acquired herd-immunity *is* or (respectively) *can be* obtained (and for which not). The combinations of *p*_*i*_*/p*_*r*_ and 𝔰_*v*_ are represented as points (*p*_*i*_*/p*_*r*_, *𝔰*_*v*_) in the *p*_*i*_*/p*_*r*_ −*𝔰*_*v*_ plane (horizontal axis and *right* vertical axis), whereas the combinations of *p*_*i*_*/p*_*r*_ and are represented as points (*p*_*i*_*/p*_*r*_, ϵ) in the *p*_*i*_*/p*_*r*_ −ϵ plane (horizontal axis and *left* vertical axis). Vaccine-acquired herd-immunity is possible only for points (*p*_*i*_*/p*_*r*_, *𝔰*_*v*_) and (*p*_*i*_*/p*_*r*_, ϵ) in the grey-shaded area of the combined *p*_*i*_*/p*_*r*_ − *𝔰*_*v*_ and *p*_*i*_*/p*_*r*_ − plane. This area is enclosed by the curves, ϵ*𝔰*_*v*_ = 1, *p*_*i*_*/p*_*r*_ = 1 and the graph of the function *f* : *p*_*i*_*/p*_*r*_ → 1 − *p*_*i*_*/p*_*r*_. Points (*p*_*i*_*/p*_*r*_, *𝔰* _*v*_) in this grey-shaded area correspond to a state of (vaccine-induced) herd-immunity by definition. In contrast, for points (*p*_*i*_*/p*_*r*_, ϵ) in the grey-shaded area, herd-immunity is obtained only when an appropriate vaccination rate 0 ≤ *s*_*v*_ ≤ 1 consistent with (7.10a) is chosen. It should be noted that the region for *p*_*i*_*/p*_*r*_ < 1 is in fact irrelevant in the context of vaccination, since herd-immunity is, so to speak, trivial and inherent to the situation here (an epidemic cannot develop at all when *p*_*i*_ < *p*_*r*_, as outlined previously).

**Fig. 7.1:**
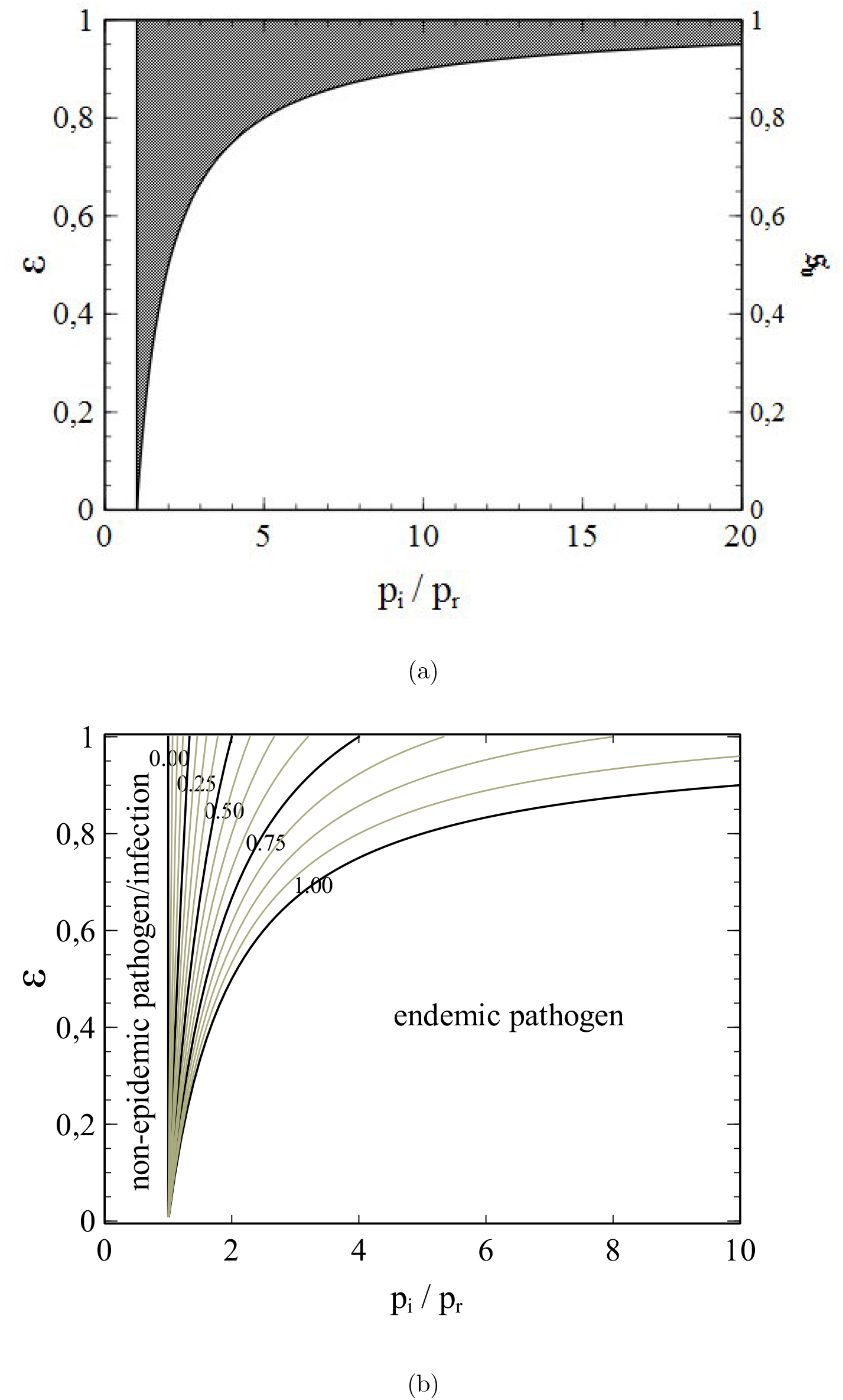
(**a**) Combined *p*_*i*_*/p*_*r*_ − and *p*_*i*_*/p*_*r*_ −*𝔰* “phase-diagrams”. Points (*p*_*i*_*/p*_*r*_, *𝔰*) (right vertical axis) in the grey-shaded area correspond to vaccine-acquired herd-immunity, points (*p*_*i*_*/p*_*r*_, *ϵ*) (left vertical axis) to the *possibility* of vaccine-acquired herd-immunity (via a sufficiently high vaccination rate, the minimum value of which can be read from the diagram in fig 7.1b below). **b**) Contour lines of the vaccination rates necessary for vaccine-acquired herd immunity as a function of *p*_*i*_*/p*_*r*_ and *ϵ*.

In addition, fig. 7.1b shows a contour map of the minimum vaccination rates necessary to obtain herd-immunity (calculated on the basis of (7.10a)) as a function of *p*_*i*_*/p*_*r*_ and ϵ. Adjacent contours correspond to a difference Δ*s*_*v*_ = 0.0625. The contours for *s*_*v*_ = 0, 0.25, 0.5, 0.75, 1 have been specially highlighted in black to serve as visual anchors. The progress of the contour lines clearly illustrates how ever higher vaccination rates become necessary to obtain herd-immunity when *p*_*i*_*/p*_*r*_ is increased while *ϵ* remains constant. Where the line *y* = ϵ= *constant* intersects the curve *y* = 1 − *p*_*i*_*/p*_*r*_ (which relates to *s*_*v*_ = 1) a critical value *r*_*c*_ = *r*_*c*_(ϵ) of *p*_*i*_*/p*_*r*_ is reached: for values of *p*_*i*_*/p*_*r*_ > *r*_*c*_(ϵ) a vaccine with efficiency *ϵ* is unable to provide herd-immunity. Conversely, fig. 7.1b also shows how ever lower values of *ϵ* necessitate ever higher values of *s*_*v*_ to obtain herd-immunity when *R*_0_ is kept fixed, until a critical value ϵ_*c*_ = ϵ_*c*_(*p*_*i*_*/p*_*r*_) = 1 − *p*_*i*_*/p*_*r*_ is reached below which no herd-immunity is possible even for *s*_*v*_ = 1. Points (*p*_*i*_*/p*_*r*_,) in the segment of the *p*_*i*_*/p*_*r*_ − ϵplane enclosed by the curve ϵ= 1− *p*_*i*_*/p*_*r*_ and the horizontal axis (ϵ= 0) therefore relate to a situation where the pathogen involved becomes “endemic”. What is meant by this is that for such combinations of *p*_*i*_*/p*_*r*_ and *ϵ* the spread of the infection cannot be stopped, despite vaccination. When additionally the individuals that have recovered from an infection only obtain a low (partial) immunity and/or loose most of their immunity after longer periods of time, the pathogen will remain circulating among the members of the population. The only way out of this situation is to develop a vaccine with an effectiveness high enough to ensure herd-immunity for a vaccination rate *s*_*v*_ ≤ 1. As long as such a vaccine is not available, the pathogen has to be considered as “endemic”.

It may also happen that a vaccination campaign is undertaken using different vaccines of different effectiveness (as in the case of, for instance, many 2021 vaccination campaigns against Covid-19). In such a situation, (7.2) should be replaced by the more general expression:

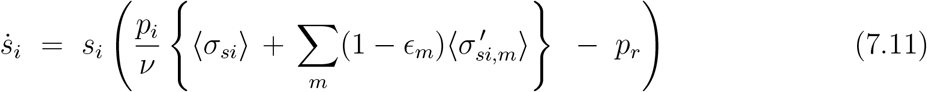

where the summation runs over the different vaccines, which are labelled by the integer m. We replace the equation on the right in (7.3) by:

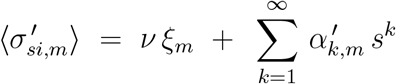

Correspondingly, the equation on the left in (7.3) is replaced by:

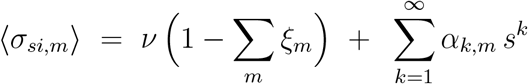

We can now write the generalisation of (7.4) as:

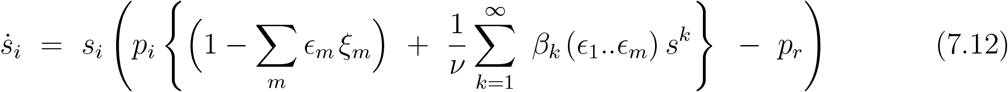

In case of at-random vaccinations that is:

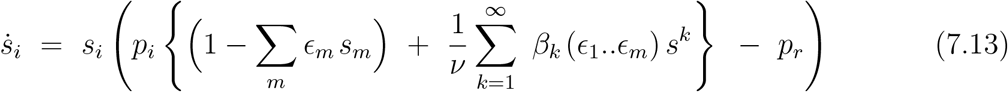

where *s*_*m*_ is the (partial) rate of vaccination with vaccine *m* (that is, the fraction of the population vaccinated with vaccine *m*). In analogy with (7.7), the criterion for herd-immunity now becomes:

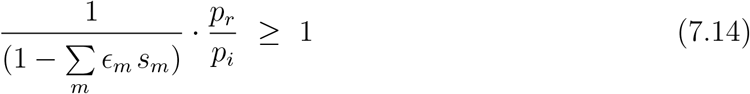

Let the vaccine-averaged efficiency be defined as:

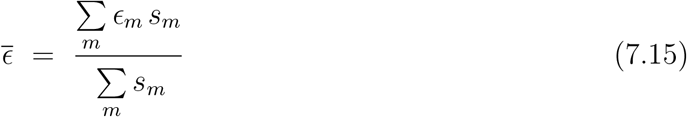

where 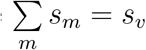 can be identified as the total (cumulative) vaccination rate (i.e. the sum of all the partial vaccination rates).

The criterion (7.14) can then be reexpressed as:

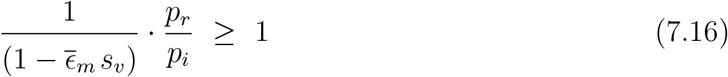

from which the critical vaccination rate follows as:

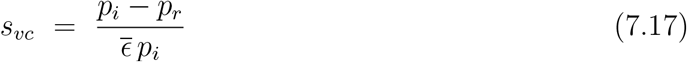

This result is of the same form as (7.8), except that the (single-vaccine) effectiveness has been replaced by the vaccine-averaged effectiveness 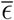. An effective vaccination rate 𝔰_*v*_ can be defined in the same way as previously, giving 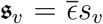, the critical effective vaccination rate 𝔰_*vc*_ being given by (7.9).

It should be noted that the obtained critical vaccination rates do *not* depend on the structure of the social network: as long as the vaccinations are carried out at random, critical vaccination rates are the *same* for all populations irrespective of their social (network) structure.

However, when a vaccination campaign is undertaken during an ongoing epidemic the situation is different, and the structure of the population network *does* have an influence on the threshold for vaccine-acquired herd-immunity, even in case of at-random vaccination.

Suppose that the infection has been spreading from *t* = 0 onwards until at *t* = *t*_0_ a vaccination campaign is started, and that only one type of vaccine is used in this campaign. As a result of the vaccinations, the susceptible part of the population is divided into a vaccinated and an unvaccinated part from *t* = *t*_0_ onwards, where the “bare” transmission constant *p*_*i*_ applies to the unvaccinated part and the reduced transmission constant 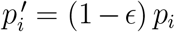 to the vaccinated part. With time, the epidemic comes to a halt under the *combined* influence of infection-removal and the vaccine-related reduction of *p*_*i*_. This situation differs from the case without a vaccination campaign during the epidemic, since in that case the epidemic comes to a halt due to infection removal *only* (and it’s effects on infection percolation).

In general, especially when vaccinations are randomly distributed across the population, the vaccination rates among the susceptibles and the removed infections differ after the end of the epidemic. These rates are represented respectively by 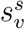 and 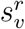. Due to the lower transmission probability that vaccinated susceptibles are subject to compared with non-vaccinated ones 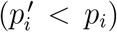, eventually (with time) the inequality 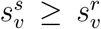 will apply. Vaccinated susceptibles will get infected (if at all) in lower numbers than the non-vaccinated ones so that they will become overrepresented among the non-infected individuals and underrepresented among the infected (the opposite being the case for the non-vaccinated susceptibles). The partial vaccination rates 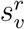 and 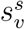 are related to the total vaccination rate *s*_*v*_ for the entire population via:

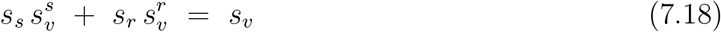

Hence, the inequality 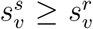 implies that 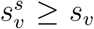: the total vaccination rate *s*_*v*_ is in fact a lower bound for the partial vaccination rate 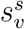 of the non-infected (s) part of the population after a (1st) wave of infections has passed.

since infection removal is considered to leave an individual with full immunity, a potential next wave (due to new pockets of active infections after the fade-out of the preceding wave) will spread exclusively among the members of the non-infected (and therefore still susceptible) part of the population. To describe the dynamics of this next (2nd) wave we introduce, in addition to 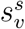, the fraction 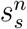 of non-vaccinated (*n*) individuals among the susceptible part of the population after the 1st wave (nb: 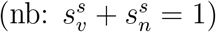). We also introduce 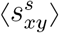, representing the average number of nodes of type *x* linked to a node of type *y* on the network formed by the nodes still uninfected (susceptible) after the 1st wave (i.e. before the 2nd wave starts due to newly introduced active infections). *After* the emergence of new active infections, the node-types can be: vaccinated (*v*), non-vaccinated (*n*), active infection (*i*) and removed infection (*r*).

Now, let *s*_*e*_ be the final value of *s* and ⟨*s*_*sr*_⟩ = *ν*(1 − *ξ*_*e*_) when the 1st wave has come to a halt and just before the start of the 2nd wave (compare to (6.27a)). As shown in the previous chapter, the value of ⟨*s*_*ss*_⟩ is then given by (compare to (6.29b)):

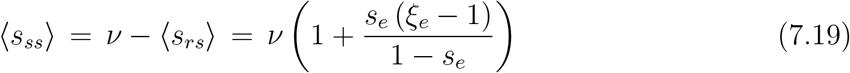

We also have (see Appendix 1):

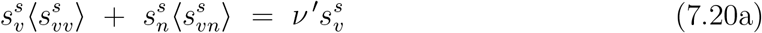

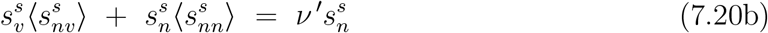

where *ν*′ = ⟨*s*_*ss*_⟩, and as such directly follows from (7.19) as:

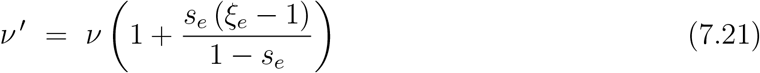

The left-hand parts of (7.20a,b) can be interpreted, respectively, as the average number of *v*-nodes linked to an *s*-node (7.20a) and the average number of *n*-nodes linked to an *s*-node (7.20b). Since we assume that the small number of new active infections is distributed randomly among the members of the susceptible part of the population, these new active infections will therefore be linked, on average, to 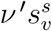 vaccinated susceptibles and to 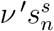 non-vaccinated susceptibles. Hence:

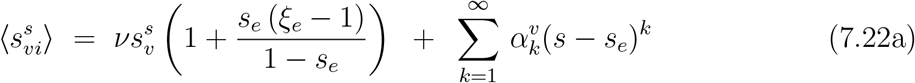

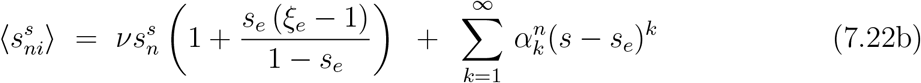

with *s* representing the cumulative infection rate over both the 1st *and* 2nd wave combined. The rate of change of the active infections is given by:

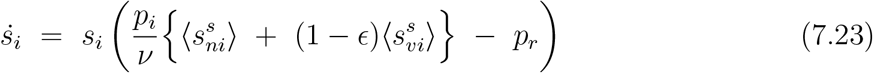

Substitution of (7.22a), (7.22b) and 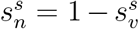 into (7.23) yields, after some rearrangements:

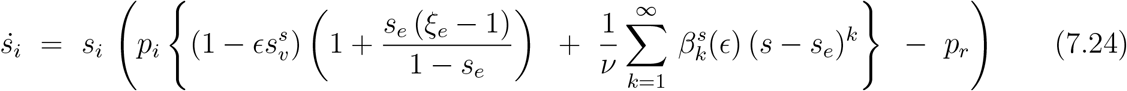

where 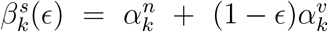.

Demanding 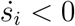 for *s* = *s*_*e*_, the criterion for herd-immunity is straightforwardly obtained from (7.24) as:

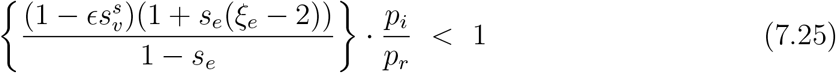

That is, we will have herd-immunity when:

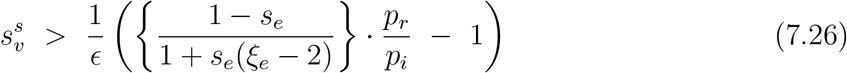

A generalisation of this result to the case of multiple vaccines with different ϵis rather straight-forward. Since 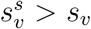, the right-hand part of this inequality can be considered as a critical value of the global vaccination rate *s*_*v*_ beyond which herd-immunity is assured.

The network structure manifests itself in this case through the values of *s*_*e*_ and *ξ*_*e*_, and is therewith a decisive factor in the achievement of herd-immunity, with a direct influence on the herd-immunity threshold. As a consequence, problems may thus arise similar to those outlined in the previous chapter in connection with the achievement of spontaneous herd-immunity. A fade-out of the number of active infections under a regime of restrictive measures, even when combined with a vaccination campaign, is *not* a guarantee that herd-immunity is being achieved. Lifting the restrictive measures to regain a regime of social normality may be accompanied by a new rise in the active-infection numbers to such an extent that even a new wave of infections cannot be ruled out in advance. Everything will depend on the values of 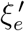 and 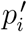 that replace *ξ*_*e*_ and *p*_*i*_ in the new regime entered after lifting the restrictions. If (7.26) is not met for 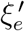 and 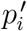 then a new wave of infections is inevitable upon lifting restrictions, despite the vaccinations administered so far (which will simply be too low in number for the establishment of herd-immunity in such a case). This has important consequences for efforts to prevent epidemic outbreaks by means of vaccination. A prophylactic vaccination campaign will provide the herd-immunity it is aiming at *only* when the resulting vaccination rate exceeds the herd-immunity threshold for a situation of social normality. If the latter is not the case, a transition from a regime of social restrictions to social normality (or a milder regime) may be followed by (significant) increases in the infection numbers, despite the vaccination campaign and an apparent fade-out of the infection rates prior to the moment of rolling back the restrictions.

## 8. Percolation

### a) The percolation transition and its relevance in the context of vaccination

So far, herd-immunity has been presented purely as a consequence (or, merely, a side-effect) of infection removal, even when network effects are involved. However, an additional independent mechanism for herd-immunity is brought about in the form of percolation by the network structure and topology typical of populations. Percolation on a lattice or network can be understood as the formation of paths along nodes of a particular type, or as the formation of (isolated) *clusters* of nodes of a certain type (either enclosed by nodes of a different type, or cut-off from the rest of the network). The formation of paths or clusters can be the result either of (random) removal/replacement of nodes or the (random) removal of links/bonds. The first case is referred to as *site-percolation* whereas the second case is called *bond-percolation* (see [1] for a basic but detailed outline of concepts and theory). Percolation phenomena play a role in many branches of the natural sciences and technology, ranging from solid-state physics (magnetic dilution) and chemistry (polymerisation) to electrical engineering (random electrical networks). It is by the very nature of the problem that the relevance of percolation appears almost self-evident in the context of epidemic infection-growth as well. Surprising it is therefore that the subject has been given fairly little attention in the epidemiological literature, despite the fact that it has been demonstrated that the percolation paradigm has its (potential) merit for the field (for example through the analysis by Davis et. al [2] of the spread of yersinia pestis (plague) among populations of great gerbils). However, a conceptually simple phenomenon at first glance, percolation is a notoriously difficult subject for mathematical analysis. Despite the fact that seminal results have been achieved during the 1950s and onwards (see [3] for an in-depth review), specific problems often defy solution by analytical means and can only be dealt with through the use of computational methods (in particular (statistical) simulations). Therefore, the emphasis of this chapter will, out of necessity, for a significant part be on computational results.

Percolation seems a particularly relevant concept in relation to (random) vaccination, especially when, as from now on, an “ideal” vaccine is considered with 100% efficiency (that is, ϵ= 1 (see previous chapter)). Nodes in the population network are randomly immunized and are no longer susceptible to infection. They can no longer become infected and, equally important, they can no longer pass on the infection to other (susceptible) nodes. They are, so to say, “inert” nodes in the network, in contrast to the “active” nodes which are either already infected or still susceptible to infection. It is easy to see that this situations corresponds, in fact, to nothing less than a genuine case of site-percolation.

An essential phenomenon to be considered now is the so-called *percolation transition*. A general feature of both bond- and site-percolation on a lattice or network, the percolation transition marks a sharp change, upon increasing the number of inert nodes, between a regime where a majority of the active nodes forms a “macroscopic” cluster of proportions comparable to those of the entire network, to a regime where the active nodes are split up in clusters of much smaller dimensions (of, for instance, no more than a few nodes). The critical value *x*_*c*_ of the fraction of inert nodes in the network at which the transition takes place is commonly referred to a the percolation threshold. A phase-transition in the true physical and thermodynamic sense, the percolation transition comes with all the typical characteristics of a thermodynamic phase transition, such as universality and scaling invariance (see [1] chapter 7). Its relevance to the problem of epidemic infection growth and herd-immunity is evident. Below the percolation threshold the infection is easily passed on throughout the entire population network. Even when the initial number of active infections is low, a vast number of nodes may eventually be reached by the infection. However, for values of *x*_*c*_ above the percolation threshold, the infection chains sooner or later run into an inert node that blocks any further propagation of the infection along that particular chain. Especially when the number of initial infections is very low, only a (very) minor fraction of the nodes will be reached by the infection and the number of accumulative infections will remain low.

The role of percolation in an epidemiological setting and its relation to herd-immunity can be demonstrated quite well via carefully thought out simulations of the evolution of an epidemic on a simplified network. A convenient choice for such a network is the 2D square lattice already introduced in chapter 4, with the nodes representing the individual members of the network. Links between nodes, representing the possibility of contact and, inherently, a route of infection transmission, can be chosen in any arbitrary way in order to simulate the effects of differences in the size and structure of the social networks of the individual members of the population. A particular benefit of such simulations is that different mechanisms can be “turned on and of” at will by an appropriate choice of their corresponding parameters, thus enabling a targeted investigation of their particular role and influence (or those of other mechanisms).

To separate the influence of infection-removal from that of percolation phenomena, the evolution of 4 epidemics was simulated for different values of the rate of (random) vaccination *x*_*v*_ on a 2D square lattice of 2001×2001 nodes while putting *p*_*r*_ = 0 (i.e. no infection removal). Periodic boundary conditions were again applied. The initial states of the population *t* = 0 at the start of each epidemic was constructed by randomly labelling nodes as vaccinated until the desired vaccination rate was reached, followed by a random selection of non-vaccinated nodes to be labelled as active infections, thus providing the “seeds” for the epidemic.

Figs. 8.1a/d show the end-status of the nodes in the population after each epidemic has come to a halt. In these simulations, nodes were considered to be linked only to their direct nearest neighbours, a situation resembling the conditions under a (very) strict lock-down. Nodes are labelled red when infected, black when vaccinated and white when still uninfected. In cases like these, where there is no infection removal, the infection spreads through the entire cluster of susceptible nodes surrounding each initial (seed) infection until the boundary of the cluster (formed by vaccinated nodes) is reached (that is, when there actually is such a boundary instead of a continuous cluster). The fragmentation of the bulk cluster of infected (red) nodes into clusters of ever smaller size upon increasing *x*_*v*_ is clearly visible. The results shown are consistent with the value of the site-percolation threshold of a 2D square lattice of *x*_*c*_ ≈ 0.41 as reported in the literature for the case of random blocking of sites [4].

**Fig. 8.1:**
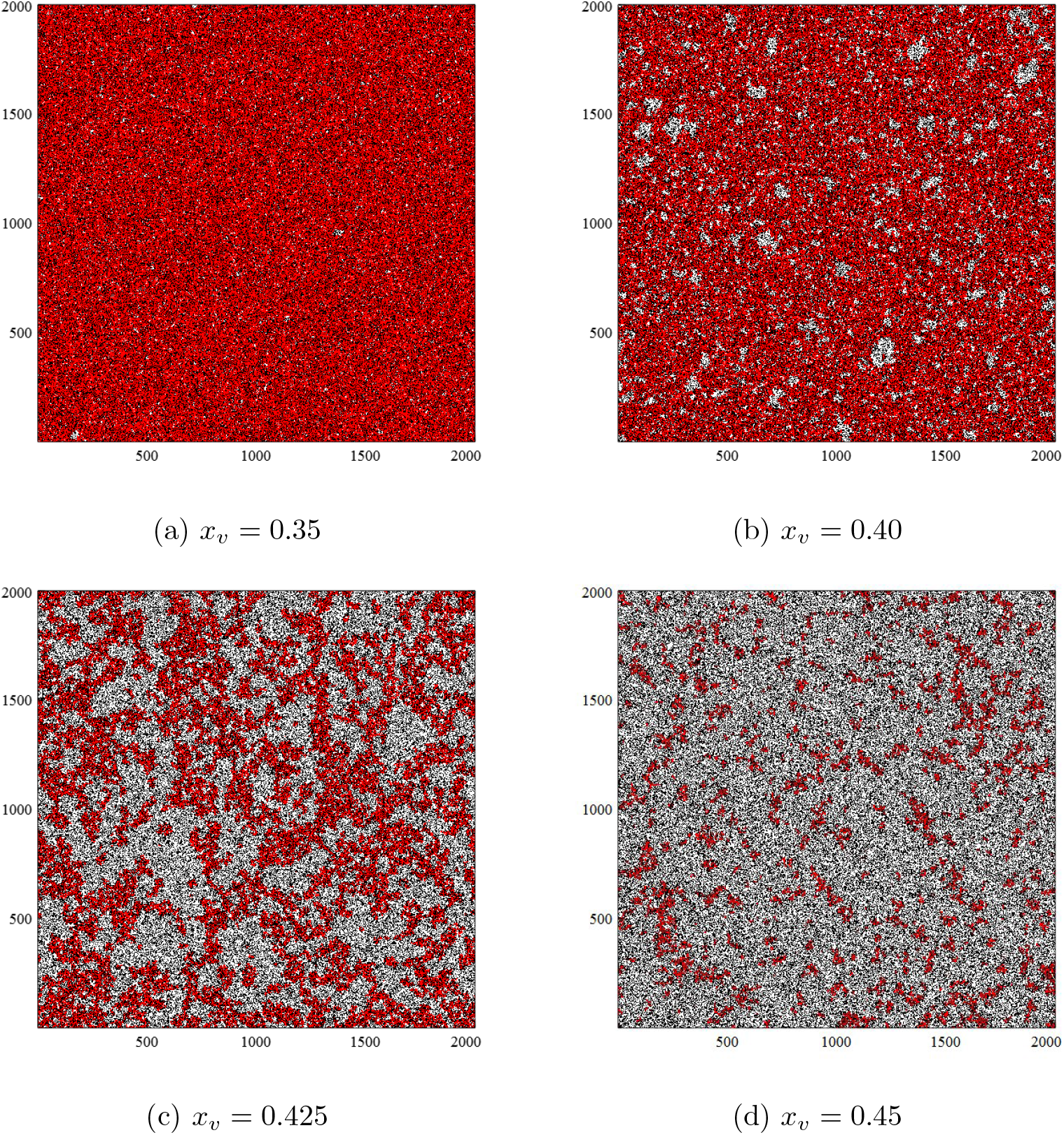
End-status (after fade-out of the epidemic) of the nodes in a model-population consisting of a 2D square lattice with nearest-neighbour contacts for different rates (*x*_*v*_) of random vaccination. The different nodes types are distinguished by the colour of the square unit cell that surrounds them (red: infected nodes, black: vaccinated nodes, white: susceptible nodes)

To allow for a more detailed impression of the effect of vaccination at the level of the individual nodes, enlarged smaller sections of the respective network end-states shown in figs. 8.1a/d are represented in figs. 8.2a/d. It is clearly recognisable how, with increasing *x*_*v*_, more and more paths along susceptible nodes become interrupted by vaccinated nodes, even to the level that susceptible nodes and actually entire clusters of susceptible nodes become fenced-in by a closed “ring” or even a cluster of vaccinated nodes, thus shielding the susceptible nodes involved from active infections outside the cluster. Only infections from *inside* such enclosed clusters of susceptibles may lead to a spread of the infection to other members of the cluster. When *p*_*r*_ = 0, eventually *all* the nodes in a susceptible cluster will become infected in the end. But, whereas below the percolation threshold *x*_*c*_ this implies that a majority if the non-vaccinated part of the population (if not the entire part) will become infected, only a small (possibly negligible) minority of the non-vaccinated individuals will become infected when *x*_*v*_ > *x*_*c*_, provided that the number of initial infections is (sufficiently) low.

**Fig. 8.2:**
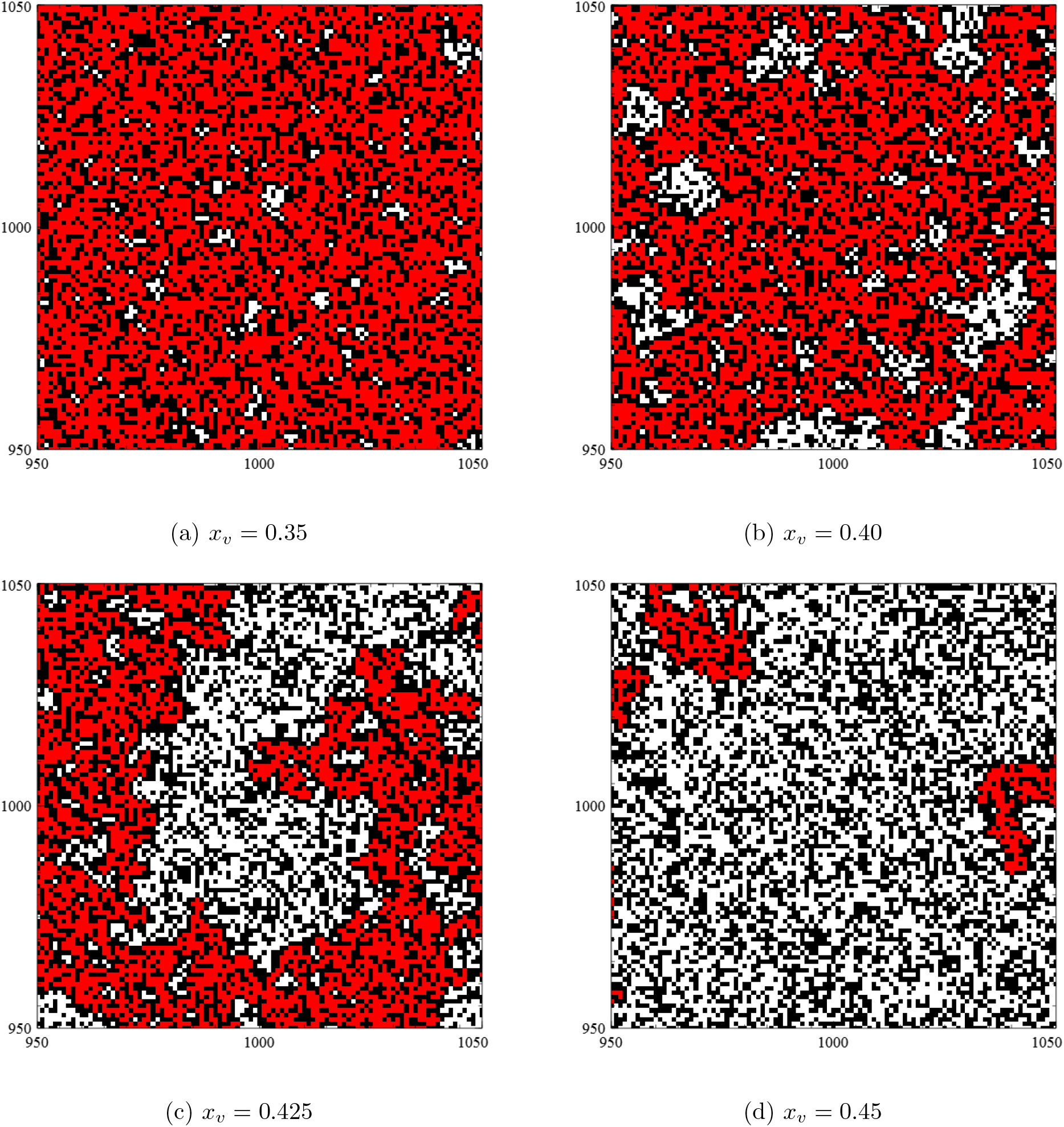
Close-up of the model-populations shown in figs. 8.1a/d (red: infected nodes, black: vaccinated nodes, white: susceptible nodes)

In a partially vaccinated population, each initial infection will “land” in one of the remaining clusters of (non-vaccinated) susceptibles. There it will start transmitting the infection via its social contacts, thus initiating the spread of infection through the cluster. However, once an entire cluster of susceptibles has become infected, further propagation of the infection will stop at the cluster boundaries formed by the closed ring of vaccinated nodes. Below the percolation threshold (*x*_*v*_ < *x*_*c*_) cluster sizes (from here onwards defined as the number of nodes in each cluster) are quite large (of the same order of magnitude as the size of the population) so that the final (cumulative) number of infections will be large too. Beyond the percolation threshold however (*x*_*v*_ > *x*_*c*_), cluster sizes are modest or even very small. Since the infection will be limited only to those clusters embracing one or more initial infections, the final number of infections will remain low when the number of initial infections is low.

In view of these considerations, we expect the cumulative number of infections at the end of an epidemic to scale with a properly weighted average ⟨*S*_*c*_⟩ of the size *S*_*c*_ of the clusters of susceptibles directly after vaccination (before the start of the epidemic). An appropriate and meaningful choice for such an average is obtained by taking the average over the fractions of the susceptible part of the population accounted for by the individual clusters:

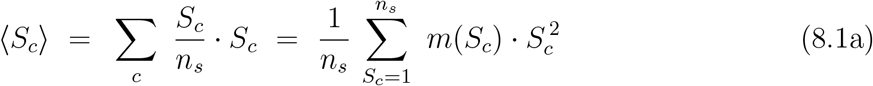

where the first summation runs over all susceptible clusters (*c*), with *n*_*s*_ representing the total number of susceptible nodes in the population, whereas the second summation runs over all cluster sizes, with *m*(*S*_*c*_) *∈* ℕ representing the actual number of clusters of size *S*_*c*_ present in the population network (so that *m*(*S*_*c*_) · *S*_*c*_ is the total number of nodes belonging to clusters of size *S*_*c*_). A quantity of even more significance is obtained when the cluster sizes *S*_*c*_ and their average ⟨*S*_*c*_⟩ are themselves considered relative to the total number *n*_*s*_ = *n*(1 − *x*_*v*_) of susceptible nodes at a given *x*_*v*_. That is, when we introduce the *relative* cluster size *S*_*c*_*/n*_*s*_ and its average ⟨*S*_*c*_⟩*/n*_*s*_. Being equal to 1 in case of only one single macroscopic cluster of a size comparable to the size of the population, but approaching zero in cases where there are only very small clusters of a few nodes embedded in a very large population, the relative clusters size *S*_*c*_*/n*_*s*_ can be seen as an *order parameter* (*r*_*p*_) for percolation on a lattice or network. From (8.1a) we immediately get:

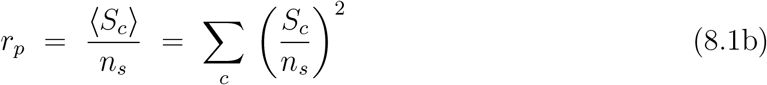

Fig. 8.3a shows ⟨*S*_*c*_⟩*/n*_*s*_ as a function of *x*_*v*_ for a 2D square lattice with nearest-neighbour contacts. Numerical data were obtained by generating a randomly vaccinated population for a series of *x*_*v*_ throughout the entire interval 0 < *x*_*v*_ < 1 and subsequent application of a computational algorithm for cluster-identification to each population, thus providing the necessary input for calculating ⟨*S*_*c*_⟩ and ⟨*S*_*c*_⟩*/n*_*s*_. The result is typical of a system showing a percolation transition. Below *x*_*v*_ = *x*_*c*_ ≈ 0.41 the average relative cluster size is close to unity. When *x*_*v*_ approaches *x*_*v*_ = 0.41 a gradual decrease in ⟨*S*_*c*_⟩*/n*_*s*_ sets in that culminates in a sharp drop at *x*_*v*_ = *x*_*c*_ ≈ 0.41 marking a transition to a regime marked by small clusters (of even negligible relative size) at higher values of *x*_*v*_.

**Fig. 8.3:**
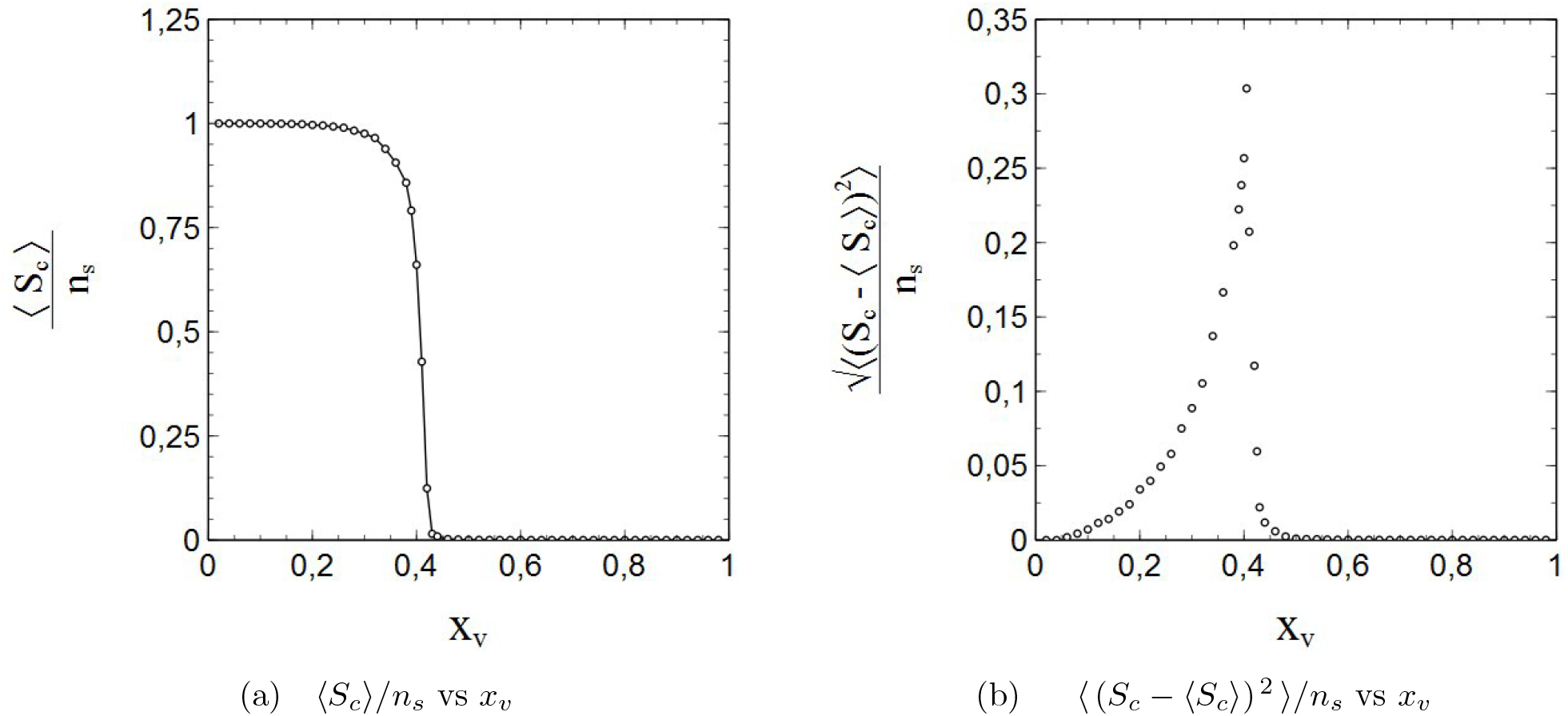
Average relative cluster size (a) and its standard deviation (b) vs *x*_*v*_ for a 2D square lattice with nearest-neighbour contacts

Furthermore, the fact that percolation transitions (including the one shown in fig. 8.3a) are true phase transitions in a statistical physical sense is reflected in fig. 8.3b, showing the standard deviation 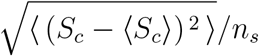 of the relative cluster size as a function of *x*_*v*_. With 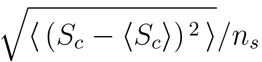 being a measure for the (average) size of the “fluctuations” in *S*_*c*_*/n*_*s*_, its behaviour as a function of *x*_*v*_ is typical of physical systems undergoing a phase transition of so-called second order, as reflected in the distinctive lambda-shape of the curve in fig. 8.3b, with a clear peak at *x*_*v*_ = *x*_*c*_ that also marks a discontinuity in the derivative.

### b) The correlation length

In the (modern) theory of phase transitions, the size of the fluctuations is intimately related to the correlation length [5], as such that the increase and divergence of the fluctuations (and other quantities) upon approaching the percolation transition is directly connected to a divergence of the correlation length.

For percolation phenomena, the correlation length *ξ* is defined (see [1] p. 64) as an appropriate average of a typical measure of length *R*_*c*_ of the isolated *finite* clusters (the large infinite “bulk-cluster” being excluded from the average):

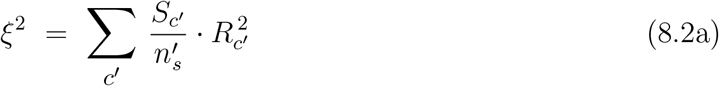

where the index *c*′ runs over the finite clusters and 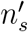 represents the total number of susceptible nodes contained in the finite clusters. Assuming only one single bulk cluster and representing its size by *S*_0_, we can write 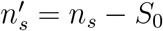. With 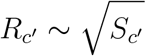 we can thus rewrite (8.2a) as:

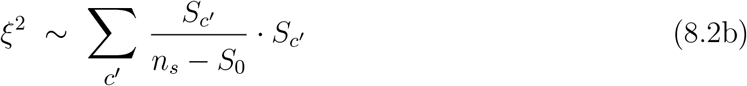

Calculation of the correlation length from computer simulations of lattices (or networks) randomly filled with susceptible and vaccinated nodes comes with a conceptual problem however. The culprit in this is the bulk cluster. In an infinite lattice, the bulk cluster is infinitely large too. In contrast, lattices simulated on a computer can *never* be infinite. The amount of available memory imposes an (absolute) upper bound upon their size, and even for sizes significantly lower than the largest size allowed for by the available memory computation times may become impractically long. Hence, computationally simulated lattices and networks are of finite and considerably limited size, and so are the clusters on them to be identified as simulated bulk clusters. The actual core of the problem here resides in the identification of these bulk clusters. Simply taking the largest cluster size for the size of the bulk cluster will not work, since there is always a largest cluster size below as well as above the percolation threshold. A constraint that the largest cluster must be “very large” in order to qualify as bulk clusters will simply replace one problem for another, since “very large” is a highly arbitrary qualification and therewith a complication in itself. Calculation of the average of the (squared) fluctuation size does not come with such difficulties however, as it involves a summation over *all* clusters, including the bulk cluster (hence the index *c* instead of *c*′):

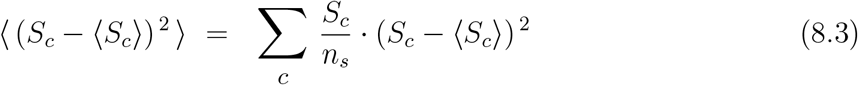

It would therefore be highly significant if we could indeed (and generally) relate every peak in 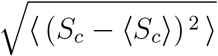 to a sharp increase (or divergence) in the correlation length, no matter the context or case in which such a peak emerges. Using somewhat pragmatic arguments it can be shown that this is actually the case.

First of all, note that 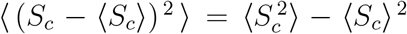. Via its definition, 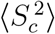 can be expressed as:

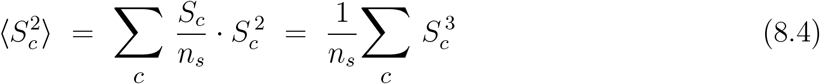

whereas ⟨*S*_*c*_⟩ follows directly from (8.1a). We fairly assume that there is only one single bulk cluster. We can then reexpress (8.1a) and (8.4) as:

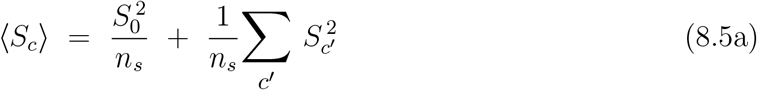

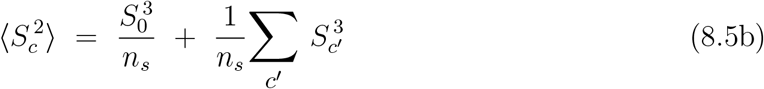

with *c*′ running over all *non-bulk* clusters. Based on considerations similar to those that led us to a scaling relation between ⟨*S*_*c*_′⟩ and *ξ* (as expressed by (8.2b)) we expect that 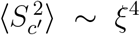. After all *S*_*c*_(*S*_*c*_′) can be considered as the square of a characteristic length of a cluster *c* (*c*′), and *ξ* by definition as the average of such a length. Hence, the following scaling relationship applies to the sum in (8.5b):

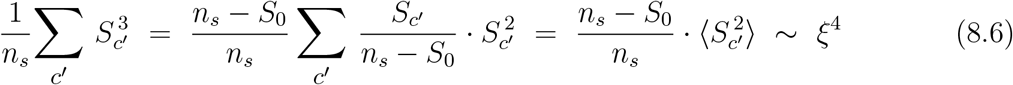

Note that 0 ≤ (*n*_*s*_ − *S*_0_)*/n*_*s*_ ≤ 1. Restating the scaling relationships (8.2b) and (8.6) as:

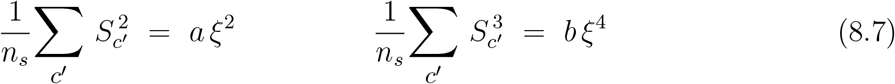

(with *a* and *b* in the order of unity) we can eventually rewrite (8.5a) and (8.5b) as:

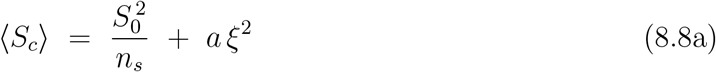

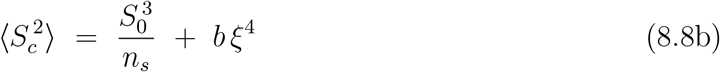

from which we obtain, via direct substitution and some minor rearrangements:

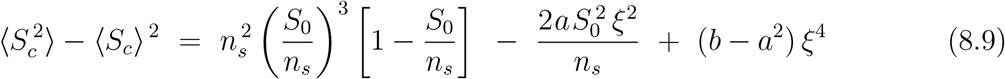

The ratio *r* = *S*_0_*/n*_*s*_ in (8.9) is generally referred to as the percolation order parameter in the literature (see [1] p. 151). It equals 1 for complete percolation (*S*_0_ = *n*_*s*_) and vanishes when the bulk cluster collapses at the percolation transition. It is closely related to the order parameter *r*_*p*_ defined via (8.1b)). The 1st term on the right-hand side of (8.9) can be expressed as 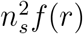, with *f* (*r*) a function of *r* given by *f* (*r*) = *r*^3^(1 − *r*). This function has 2 roots, respectively at *r* = 0 and *r* = 1, and a local maximum at *r* = 3/4. Upon approaching the percolation transition, *S*_0_ decreases more and more towards zero, and with it also *r* and *f* (*r*). As a result, the 1st term on the right-hand side of (8.9) decreases towards zero upon approaching the percolation transition (instead of increasing or even diverging). That the same applies to the 2nd term as well is nearly self-evident (since the correlation length *ξ* diverges at the percolation transition). Sufficiently close to the percolation transition we can therefore write:

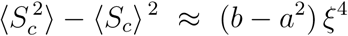

thus obtaining:

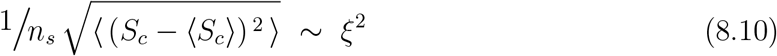

for the average size of the fluctuations represented by the standard deviation of the cluster size. Hence, a sharp increase or divergence in 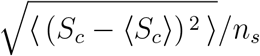 upon approaching the percolation threshold indeed relates to an increase or divergence in the correlation length *ξ* (and vice versa). A divergence in the correlation length is considered to be the quintessential feature of the critical phenomena that go with 2nd-order phase transitions. Therefore the lambda-shaped peak in fig. 8.3b can be considered as a direct manifestation of the very nature of the percolation transition, which is that of a 2nd order phase transition. It is emphasized however that (8.10) is quite a general result and therefore its use is not limited to phenomena entirely driven by percolation (as we will see in the next section).

### c) The vaccination-induced herd-immunity threshold as the critical point of a 2nd-order phase transition

As mentioned earlier, the benefit of numerical simulations is that mechanisms can be switched on and off, so that their role and influence can be investigated separately. By taking *p*_*r*_ = 0 in the simulations, the influence of vaccination and the role of vaccination-related percolation phenomena in the evolution of an epidemic can be isolated and illustrated (as shown by figs 8.1a-d and 8.2a-d in the previous section).

Now, let *n*_*e*_ represent the final number of cumulative infections after an epidemic has come to a halt and *n*_*s*_ = *n*_*p*_(1 − *x*_*v*_) the number of susceptible nodes left after vaccination. Fig. 8.4a shows, for *p*_*r*_ = 0, the *relative* rate *n*_*e*_*/n*_*s*_ = *s*_*e*_/(1 − *x*_*v*_) of cumulative infections at the end of an epidemic as a function of the rate of random vaccination *x*_*v*_ for the 2D square lattice with nearest neighbour interactions under consideration ^7^. Data were obtained by (again) generating a randomly vaccinated population for given *x*_*v*_ followed by a random labelling of a fixed number of sites as active infections to serve as seeds for the epidemic, and then letting the epidemic spread until it has faded out and come to a halt. We thereby assume that vaccinated nodes are fully immune to infection. The propagation of the infection itself is simulated in the same way as in the simulations presented in chapter 4, namely by repetitive random selection of nodes, checking whether a selected node is infected, randomly selecting one of its nearest neighbours, check its status (*s, i* or *v*) and turn it into an active infection as well when it is found to be susceptible *and* a generator of pseudo random numbers outputs a number lower than the given transmission probability *w*_*i*_. The number used in the simulations of the initial infections (seeds) was taken to be *n*_0_ = 10^3^. Fig. 8.4b shows the data on *s*_*e*_/(1 − *x*_*v*_) vs *x*_*v*_ shown in fig. 8.4a together with the relative size ⟨*S*_*c*_⟩*/n*_*s*_ of the susceptible clusters after vaccination and prior to the epidemic. The obvious similarity between the curves in fig. 8.4b in both a qualitative and quantitative sense cannot be overlooked and is in agreement with the conjecture that *n*_*e*_*/n*_*s*_ scales with ⟨*S*_*c*_⟩/*n*_*s*_. This conjecture can also be made plausible through simple though somewhat crude arguments. Consider the population of susceptible nodes to be split-up into *N*_*c*_ clusters of different sizes (*S*_*i*_) according to a cluster-size distribution of some kind (i.e. prior to the introduction of the initial infections). The number of initial infections *n*_0_ is small but large enough for the initial infections to “sample” the cluster-size distribution when they are randomly distributed over the susceptible nodes. The total size *S* of all clusters together is equal number of susceptible nodes *n*_*s*_:

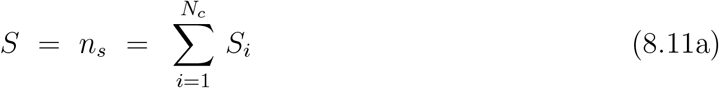

where the summation runs over all clusters (and *N*_*c*_ represents the total number of clusters). The total number of initial infections is equal to the sum of all the occupation numbers^8^ *n*_0,*I*_ of the individual clusters at *t* = 0 (note that, by definition, 0 ≤ *n*_0,*i*_ ≤ *S*_*i*_):

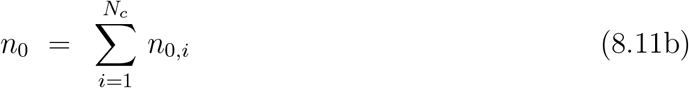

**Fig. 8.4:**
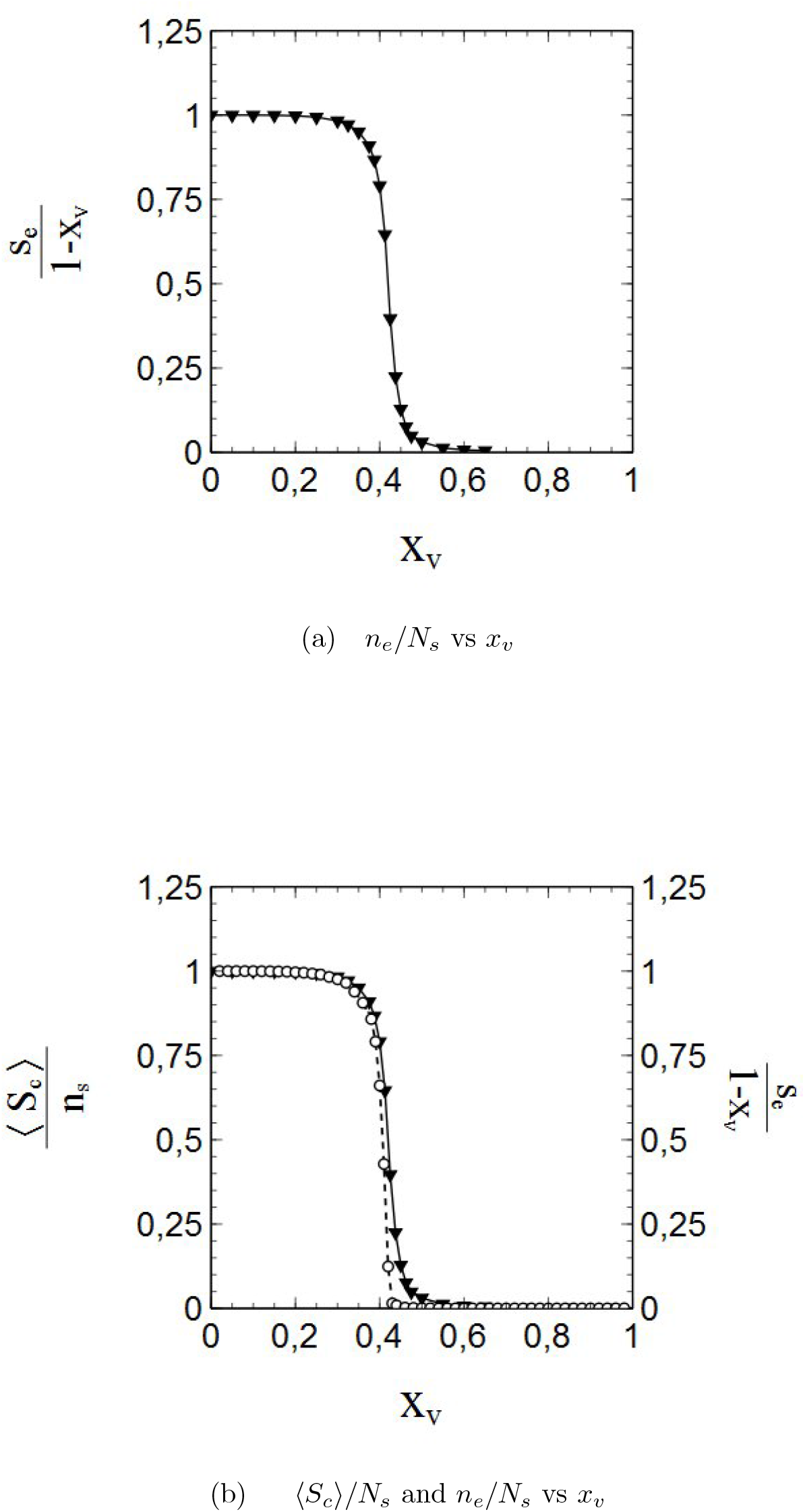
(**a**) Relative rate of cumulative infections *n*_*e*_*/n*_*s*_ = *s*_*e*_/(1 − *x*) vs *x*_*v*_ and (**b**) relative rate of cumulative infections *n*_*e*_*/n*_*s*_ = *s*_*e*_/(1 − *x*) vs *x*_*v*_ (black triangles/solid line) compared to the relative average cluster size ⟨*S*_*c*_*/n*_*s*_⟩ vs *x*_*v*_ (open circles/dashed line)

We introduce {*n*_0,*i*_} = {*n*_0,1_, *n*_0,2_, *n*_0,3_…*n*_0,*N*_} as the set of occupation numbers of the clusters. For a given set of occupation numbers (i.e. for a given distribution of initial infections) the cumulative number of infections at the end of an epidemic 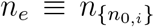 can be written as (remember that all clusters containing one or more initial infections will become infected):

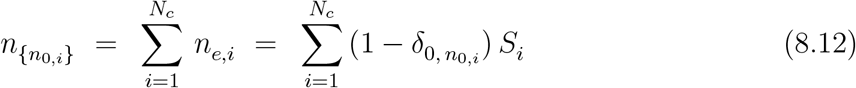

Here the *n*_*e,i*_ represent the number of cumulative infections in the *i*th cluster at the end of the epidemic, and 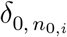 the Kronecker-delta:

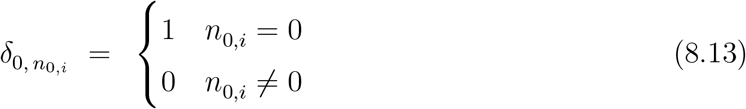

We now introduce 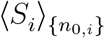 as the average, for given {*n*_0,*i*_}, of the size of the *cumulatively infected* clusters over the number of *cumulatively infected* nodes at the end of the epidemic:

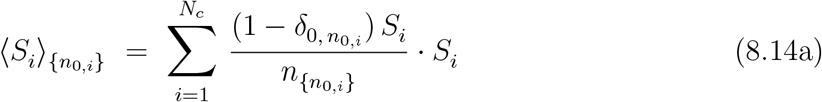

which can be reexpressed, by substitution of (8.12) for 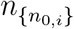, as:

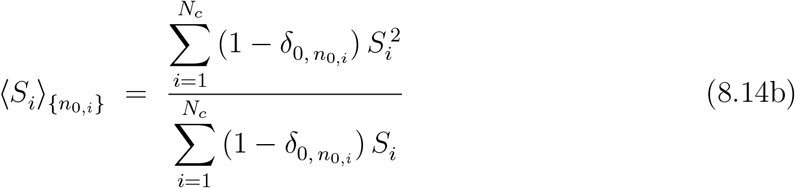

Since the initial infections sample the cluster-size distribution (when sufficiently large in number), the average 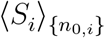 can be considered as a fair approximation of the previously introduced average ⟨*S*_*c*_⟩ of the size of the susceptible clusters after vaccination and prior to the epidemic. Hence, an approximation for the ratio 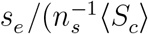 of the normalised infection rate 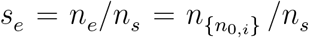 and the normalised average cluster size ⟨*S*_*c*_⟩*/n*_*s*_ easily follows. Combining (8.12) and (8.14b) yields:

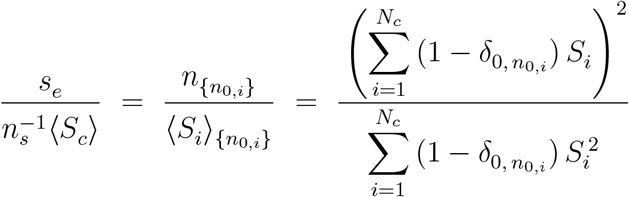

so that, via some algebraic rearrangements, we get:

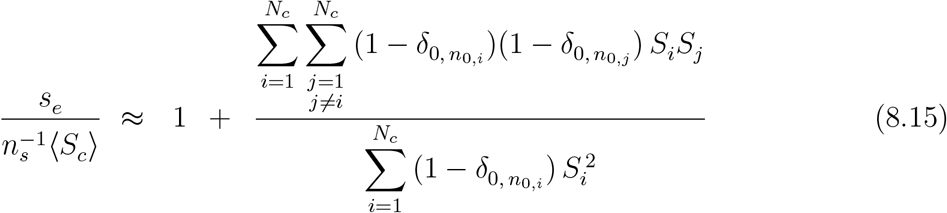

At low vaccination rates, a (vast) majority of the susceptible nodes forms a large (bulk) cluster of a size comparable to the size of the population network. With increasing vaccination rates, more and more clusters separate from the bulk cluster and become isolated. However, the size of these “secondary” clusters is much smaller (up to orders of magnitude even) than that of the bulk cluster. Note that the bulk cluster does not contribute to the numerator of (8.15) but does contribute to the nominator of (8.15). As such, the products *S*_*i*_*S*_*j*_ in the terms these smaller clusters contribute to the numerator in (8.15) will lose against the 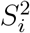 term contributed by the bulk cluster to the nominator in (8.15). In addition, due to their small size only a (small) minority of the the separated clusters will include an initial infection, especially when *x*_*v*_ approaches the critical region near the percolation transition where the number (but not the typical size) of the separated clusters increases significantly. Moreover, the probability that 2 specific secondary clusters (to be identified by their indices *i, j*) *both* include an initial infection is (very) low. Hence, many of the product terms 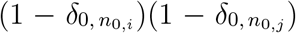 in the numerator of (8.15) will be zero over the entire range of *x*_*v*_-values up to the critical region *and beyond*. Altogether, it strongly looks as though (8.15) can be reduced to *s*_*e*_/⟨*S*_*c*_⟩ ≈ 1 and that *s*_*e*_ scales indeed with the average cluster size ⟨*S*_*c*_⟩.

The divergence, as a function of *x*_*v*_, in the average of the fluctuation in the size ⟨*S*_*c*_⟩ of the susceptible clusters shown in fig. 8.3b is also reflected in the average fluctuation size for the *infected* clusters present in vaccinated populations after an epidemic has faded out. fig. 8.5 shows √⟨(*S*_*c*_ − ⟨*S*⟩)^2^⟩/*n*_*s*_ for these infected clusters vs *x*_*v*_. The divergence clearly stands out, and the behaviour of √⟨(*S*_*c*_ − *(S)*)^2^⟩*/N*_*s*_ as a function of *x*_*v*_ is typical therewith of a phase transition. The dotted lines in fig. 8.5 further substantiate this viewpoint. They mainly serve as guides to the eye, but have been calculated by adjusting the parameters *c*′, *c*′′, *ν* and *x*_*c*_ of the function:

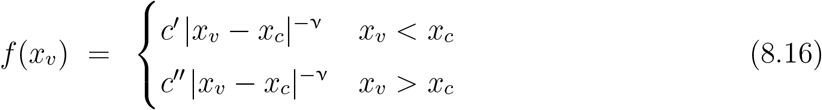

to the datapoints. The function *f* (*x*_*v*_) represents a scaling law of the type typically associated with the critical phenomena complementing a 2nd-order phase transition, with ν representing a (so-called) critical exponent. The dotted lines fit the datapoints quite well, and √⟨(*S*_*c*_ − *(S)*)^2^⟩*/n*_*s*_ shows therewith the appropriate scaling behaviour expected in connection with a 2nd-order phase transition. As such, the onset of vaccination-induced herd immunity can be perceived in itself as a phase transition of 2nd order (especially since we previously saw that the divergence in √⟨(*S*_*c*_ − *(S)*)^2^⟩*/n*_*s*_) relates to a divergence in the correlation length). fig. 8.5 provides insufficient data though to obtain an accurate estimate for *x*_*p*_ and ν.

**Fig. 8.5:**
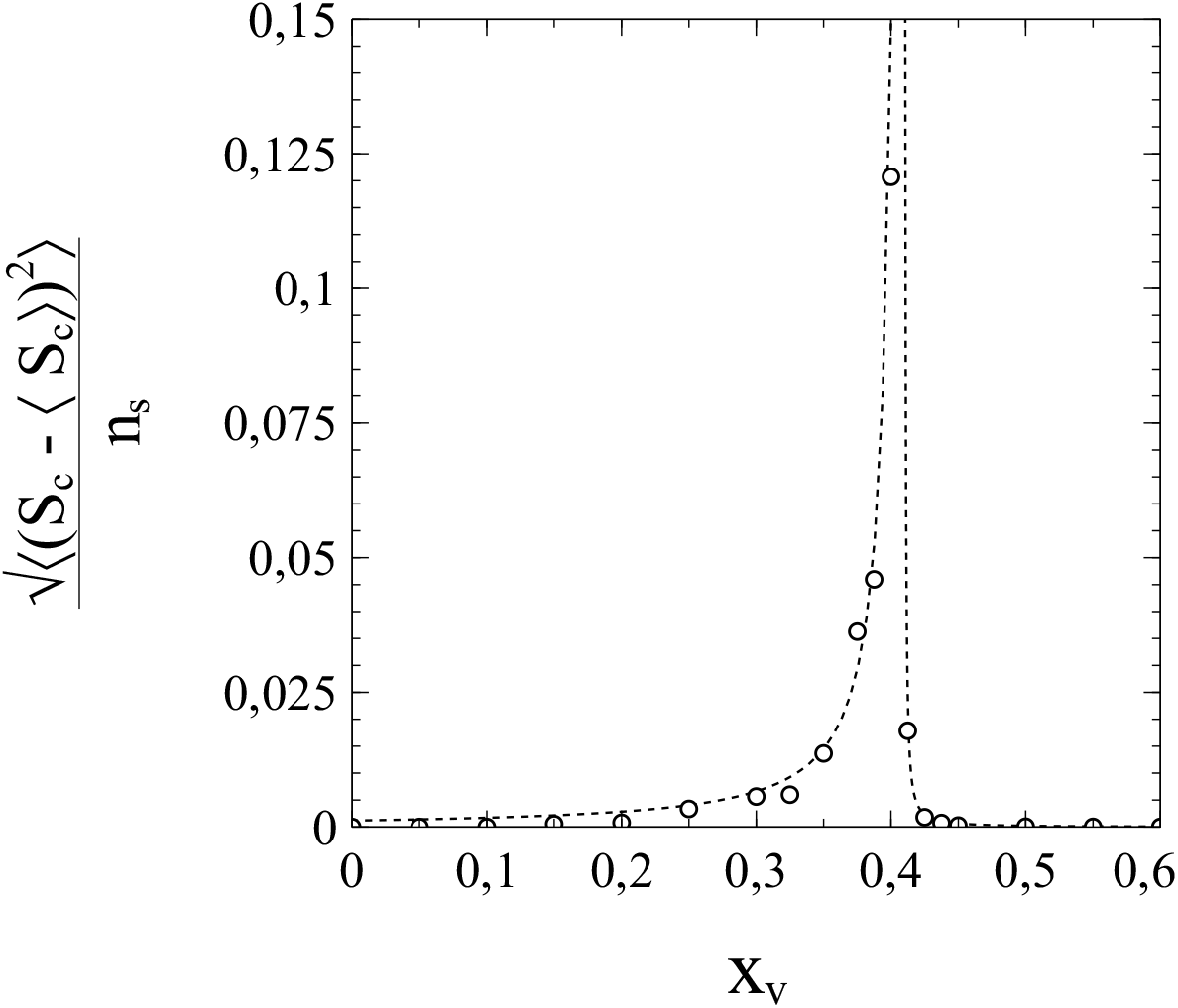
Average of the fluctuations in the size of the clusters of cumulative infections after fade-out of the epidemic vs vaccination rate *x*_*v*_. Dotted curves are guides to the eye.

### (d) Vaccination-induced percolation transitions in the framework of the SIR model

The effects of percolation in vaccinated populations can also be dealt with in the context of the SIR model, provided we modify the standard SIR model a little further beyond the modifications already presented in chapter 1.

Percolation phenomena directly affect the (average) number of susceptible contacts ⟨*s*_*si*_⟩ of an active infection, and they can be counted for as such via the coefficients of the expansion of ⟨*s*_*si*_⟩ in *s*. Close to the percolation threshold (i.e. to the critical vaccination rate), ⟨*s*_*si*_⟩ decreases sharply with any further increase in *s* when *s* → *s*_*e*_, since the active infections will then approach the cluster boundaries formed by the vaccinated nodes. When *s* = *s*_*e*_, the active infections will actually reach the cluster boundaries so that ⟨*s*_*si*_⟩ vanishes. For the SIR model to be consistent with this, constraints must be imposed upon the coefficients of the truncated series expansions of ⟨*s*_*si*_⟩ to be used, such that the following condition applies:

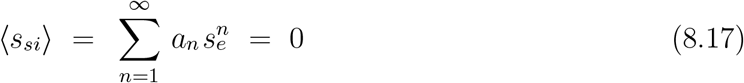

However, to properly deal with percolation phenomena and describe them correctly in terms of a modified SIR model, some further adjustments of the model seem to be inevitable. The reason is that we have to deal with 2 qualitatively different regimes, respectively below the percolation transition (*x*_*v*_ < *x*_*p*_) and above the percolation transition (*x*_*v*_ > *x*_*p*_). Below the percolation transition the bulk cluster dominates, whereas, in the absence of a bulk cluster, the secondary clusters take over in the regime above the transition. It seems appropriate therefore, to split the clusters into a subset consisting of the cluster(s) of (equal) maximum size (which consists of a *single* bulk cluster for sufficiently low *x*_*v*_), and a subset comprised of all other (secondary) clusters. The contributions to *s* (and *s*_*e*_) from both respective subsets are then treated separately. For purpose of the latter we introduce:

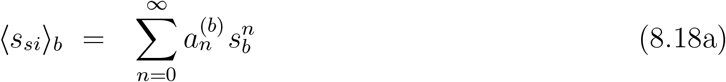

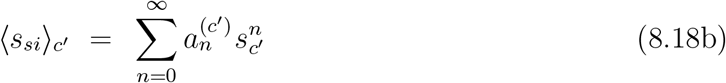

where ⟨*s*_*si*_⟩_*b*_, ⟨*s*_*si*_⟩_*c*_′ represent the average number of *s* nodes in contact with an *i* node on respectively the bulk (maximum-sized) cluster(s) (labelled b), and the secondary clusters (labelled *c*′). The variables *s*_*b*_ and *s*_*c*_′represent, the respective occupation rates, *relative to n*_*s*_, of the susceptible nodes in the bulk (maximum-sized) cluster(s), and those in the secondary clusters. Note that, by definition, a node in one cluster/subset has *no* contact with nodes in the other clusters/subsets, so that we can expand ⟨*s*_*si*_⟩_*b*_, ⟨*s*_*si*_⟩_*c*_′ in a single variable only (*s*_*b*_ and *s*_*c*_′ respectively).

Now, let *S*_*b*_ represent the combined sizes of the clusters of maximum size (that is, the size of the single bulk cluster when existent, or the sum of the nodes contained in the multiple of maximum-sized clusters) and ⟨*S*_*c*_′⟩ the average size of the secondary clusters:

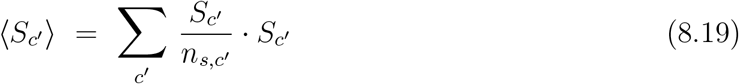

where *c*′ runs over the secondary clusters, and *n*_*s,c*_′ represents the total number of susceptible nodes in the secondary clusters before the infection spreads. Note that ⟨*S*_*c*_′⟩ is therewith an average over the susceptible nodes in the secondary clusters only. Both *S*_*b*_ and ⟨*S*_*c*_′⟩ are functions of *x*_*v*_. The number of nodes in the bulk (maximum-sized) cluster(s) is of course equal to *S*_*b*_. Following section 8a, we also introduce the *relative* cluster sizes:

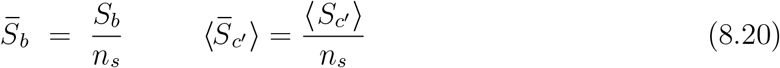

The total number of susceptible nodes in the network is given by:

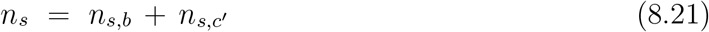

where *n*_*s,b*_ represents the number of susceptibles in the bulk cluster(s) prior to the start of the epidemic (note that *n*_*s,b*_ = *S*_*b*_ actually). We can rewrite this relation in terms of the relative *total* sizes 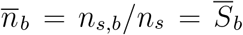 and 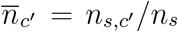 of respectively the bulk (maximum-sized) cluster(s) and the secondary clusters as:

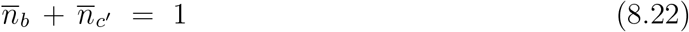

It is easy to see that in each separate regime (below or above the percolation threshold) the values 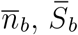 and 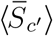 will be unique for each value of *x*_*v*_. Stated differently: 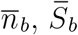 and 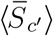are bijective functions of *x*_*v*_ on the intervals 0 ≤ *x*_*v*_ ≤ *x*_*p*_ and *x*_*p*_ < *x*_*v*_ ≤ 1. They can therefore be considered as state variables for the subsets of clusters to which they relate. As a corollary, the coefficients 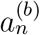 in the series for ⟨*s*_*si*_⟩_*b*_ in (8.18a) can be expanded themselves in either 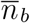 or 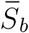, just like the coefficients 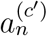 in the series for ⟨*s*_*si*_⟩_*c*_′ in (8.18b) can be expanded in 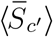. The variable 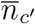 does it qualify as a proper state variable however, since the secondary clusters prevail above the percolation threshold (*x*_*v*_ > *x*_*p*_) and contain all susceptible nodes, making *n*_*c*_′ */n*_*s*_ = 1 for all *x*_*v*_ in the interval *x*_*p*_ < *x*_*v*_ ≤ *x*_*p*_. Furthermore, it should be kept in mind that, since we deal with 2 qualitatively different *x*_*v*_-regimes, each regime may require its own series expansion of the 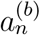 and 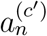. Expansions of 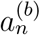 and 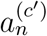 in terms of, respectively, 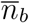and 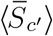 seem to be the most elegant and convenient path to follow.

It is at this point where constraints of a kind similar to (8.17) come into play. First of all, we note that in case of vaccination at random:

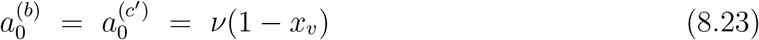

The series expansion for ⟨*s*_*si*_⟩_*b*_ can then be written as:

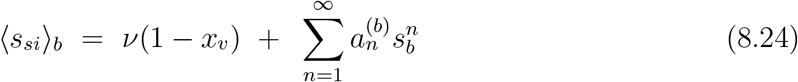

When the bulk cluster is entirely infected 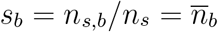 (compare with (8.17)):

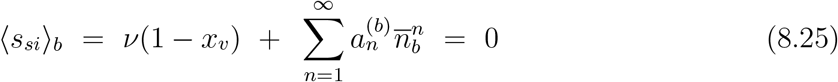

which, by introducing a function 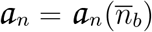, such that:

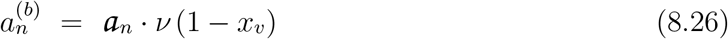

can be reexpressed as:

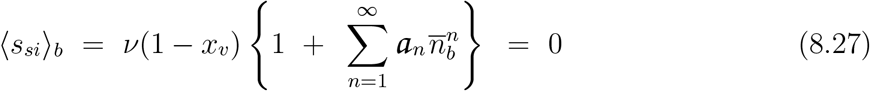

For this condition to be met, a power-series expansion of the function 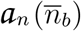 in 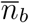 cannot have the form of a Taylor series, since ⟨*s*_*si*_⟩_*b*_ should vanish when 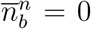 (which will be the case upon reaching the percolation threshold *x*_*v*_ = *x*_*c*_). A Laurent series is to be used instead:

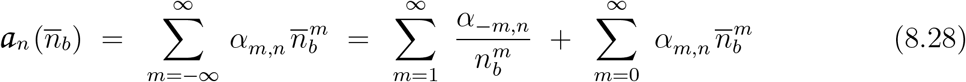

which also includes negative powers of *n*_*b*_. Substitution of (8.28) into (8.27) then yields for *x*_*v*_ ≠ 1, and with *i* = *n* and *j* = *n* + *m*:

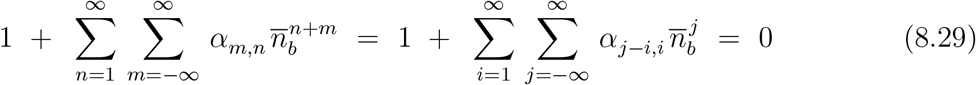

Collecting terms with equal powers of *n*_*b*_ we get, for *j* = *n* + *m* = 0:

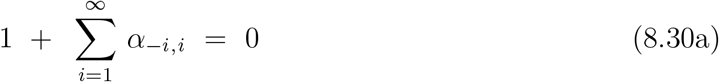

and for *j* = *n* + *m* ≠ 0:

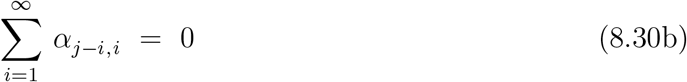

Following the approach in section 3c, by assuming that we can neglect terms of 3rd and higher order in the series expansion of ⟨*s*_*si*_⟩_*b*_, only 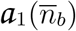 and 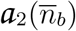 will be relevant. For *n* = 1 and *n* = 2 (8.28) then yields, respectively:

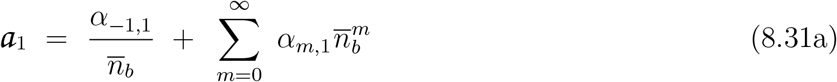

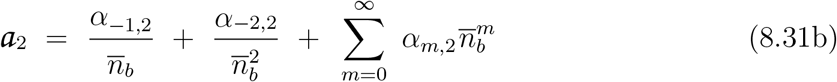

Note that *α*_*m,n*_ = 0 for *m* < −1 when *n* = 1, and *α*_*m,n*_ = 0 for *m* < −2 when *n* = 2 since otherwise ⟨*s*_*si*_⟩_*b*_ would diverge for *n*_*b*_ → 0. For the 2nd-order polynomial approximations of ⟨*s*_*si*_⟩_*b*_ considered here, the following constraint must apply when 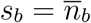 (see (8.25)):

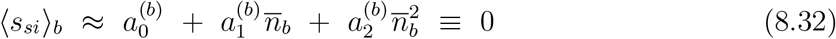

so that in combination with (8.26) we get:

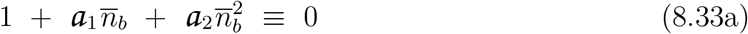

That is:

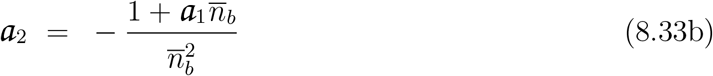

Substitution of (8.31a) then yields:

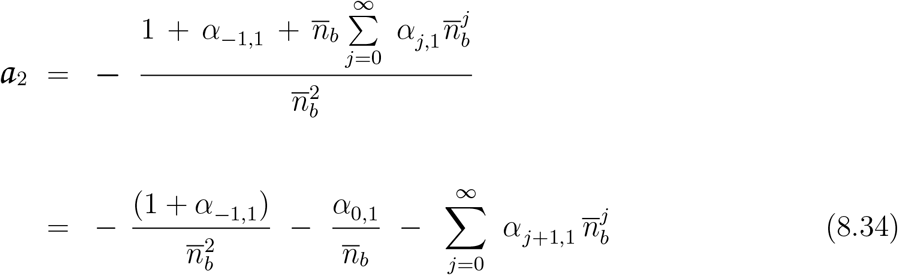

from which, by comparison with (8.31b), *α*_−1,2_ and *α*_−2,2_ can be identified as:

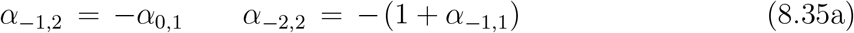

a result in agreement with (8.30a,b). Also by comparison of 8.34 and 8.31*b*, the *α*_*j*,2_ for *j* ≥ 0 follow as:

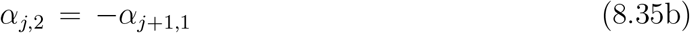

By combining (8.26), (8.31a,b) and (8.34), the Laurent series in 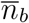 for 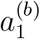 and 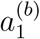 can thus be written as:

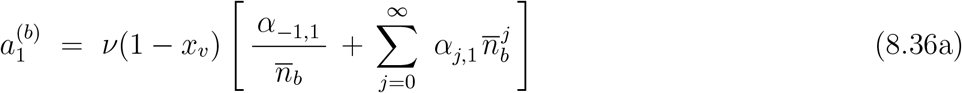

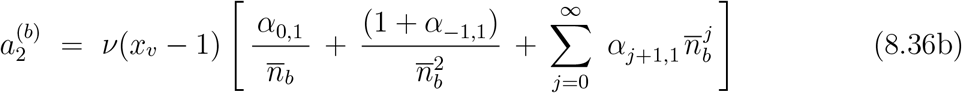

Series expansions for the 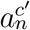 in (8.18b) follow along similar lines. There is an issue to be considered however. To obtain the counterpart of (8.25) for the secondary clusters, we must be able to specify the total number of nodes in the subset of secondary clusters that will become infected in the absence of infection removal. We will therefore deal with this issue first.

Consider a fairly large number *n*_0_ >> 1 of initial infections randomly distributed among the unvaccinated nodes. The most likely distribution of these infections over the unvaccinated *clusters* that follows is one in which a (sufficiently large) cluster of size *S*_*c*_ will, on average, contain a number of *n*_0,*c*_ = *n*_0_ · (*S*_*c*_*/n*_*s*_) initial infections. When there is a bulk cluster of unvaccinated nodes present (*x*_*v*_ < *x*_*p*_) it will therefore almost certainly contain more than a single initial infection, since *S*_*b*_ is of the same order of magnitude as *n*_*s*_ (or even (almost) equal *n*_*s*_), and *n*_0_ >> 1. Without infection removal, a contribution of a total of *n*_*s,b*_ = *S*_*b*_ cumulative infections due to infection of the entire bulk cluster is certain therewith.

Estimating the contribution to the cumulative infection rate from secondary clusters is a different matter however. Since secondary clusters can be very small, the equation *n*_0,*c*_ = *n*_0_ · (*S*_*c*_*/n*_*s*_) for the most likely number of initial infections in a cluster does not necessarily apply to them. For instance, although *n*_0_ · (*S*_*c*_*/n*_*s*_) ≠ 0, it is highly conceivable that a significant part of the smaller secondary clusters will in fact remain without any (initial) infections (especially when *n*_0_ << *n*_*s*_). And although, provided the distribution of cluster sizes is known, the combinatorics of the problem *is* tractable,^9^ it does not provide us with concise algebraic results.

A useful alternative can be based though on the fact that 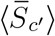 is a state variable: the cluster distribution is unique for each *x*_*v*_, and so is the average (relative) cluster size. The importance of this is that the average cluster size directly affects the expectation value 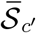 of the relative *total* size of the secondary clusters that contain at least 1 single initial infection. The value of 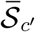 is crucial, since it represents the number, relative against *n*_*s*_, of all the nodes that will get infected upon the spread of the infection (when *p*_*r*_ = 0). That is, it represents the end-value *s*_*c*_′_,*e*_ of the cumulative relative infection rate *s*_*c*_′ of the secondary clusters after the epidemic has come to a halt (0 ≤ *s*_*c*_′ ≤ *s*_*c*_′_,*e*_ with 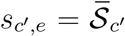). In addition, 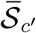 is also determined by *n*_0,*c*_′, the number of initial infections distributed among the nodes of the secondary clusters. We can therefore consider 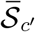 to be a function of both 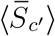 and *s*_0,*c*_′ = *n*_0,*c*_′ */n*_*s*_. That is: 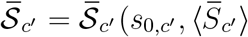. As such, we will express the coefficients 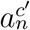 of the series expansion of ⟨*s*_*si*_⟩_*c*_′ in (8.18b) as series in terms of 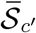, and therewith in terms of both 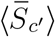 *and s*_0,*c*_′. For purpose of the latter we will first express 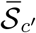 in terms of 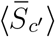 and *s*_0,*c*_′.

We reasonably assume 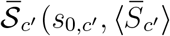 to be a continuous function on the relevant part 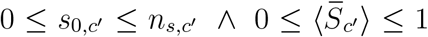 of its domain, and that the Taylor series of 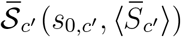 in 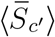 and *s*_0,*c*_′ exists on this domain part, so that:

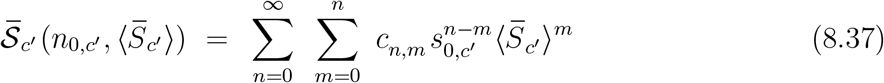

Since 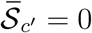 for *s*_0,*c*_′:

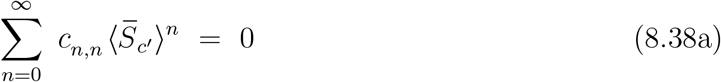

Furthermore, since 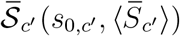 is a continuous function on the relevant part of its domain, and since 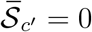 for 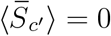, it is not difficult to see that for *all s*_0,*c*_′ :

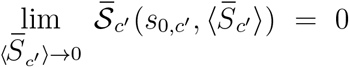

Hence:

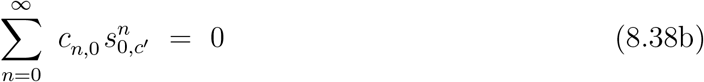

Based on (8.38a) and (8.38b), the series expansion in (8.37) can be reduced to:

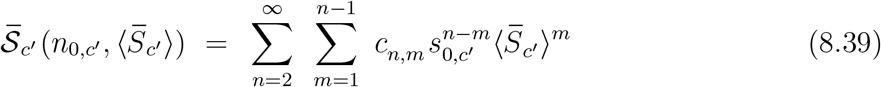

leaving (with *i* = *n* − *m* and *j* = *m*) a summation over terms 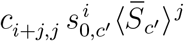 with *i, j* > 0 (as required by (8.38a) and (8.38b)). We assume that a sufficient part of the secondary clusters, and also the number of initial infections *n*_0,*c*_′ = *n*_*s*_*s*_0,*c*_′ are small enough to neglect all terms with *i* > 1 and *j* > 2. We can therefore put:

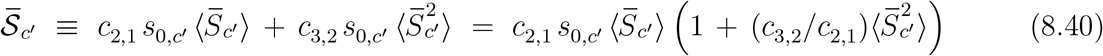

The derivation of the coefficients 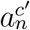 as a function of 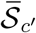 is somewhat tedious but similar in line to the derivation of the coefficients 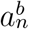 as a function of *n*_*b*_. It basically comes down to replacing 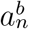 by 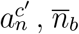 by 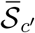 and the coefficients *α*_*n*_ by the coefficients 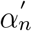 related to the series expansion of the 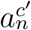 in 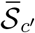. A detailed outline is presented in the Appendix 2. The result for 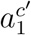 and 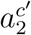 reads:

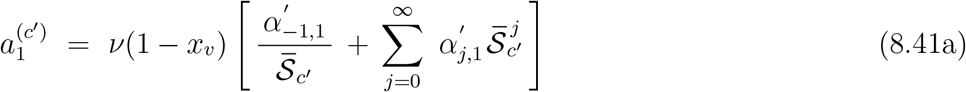

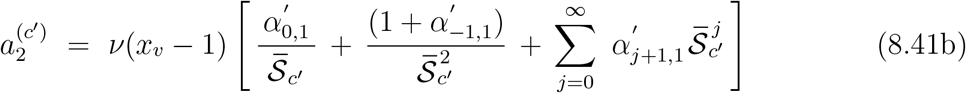

With 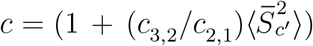, substitution of (8.40) here for 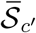 finally yields 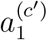 and 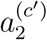 in terms of *s*_0,*c*_′ and 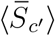:

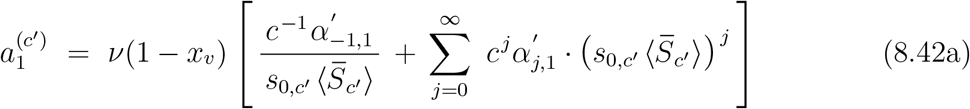

and:

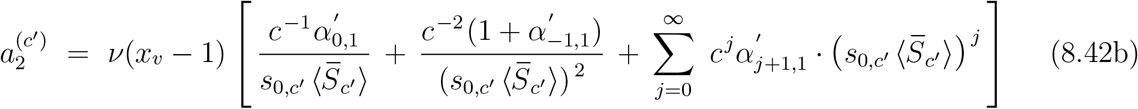

When we neglect terms in the summations over j with *j* > 1 (which implies that we assume 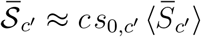, eqs. (8.42a) and (8.42b) reduce to:

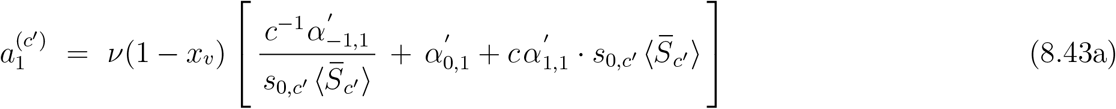

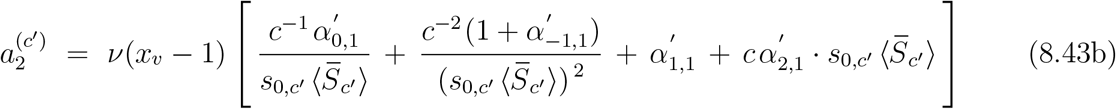

Furthermore we can also neglect/drop the terms linear in 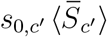 here, since *s*_*c*_′ is positive, and 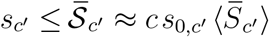. As a result of that (8.43a) and (8.43b) then reduce to:

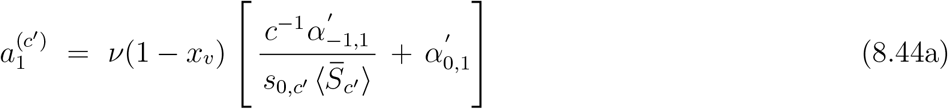

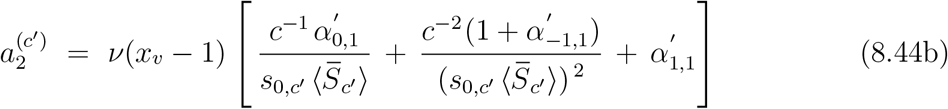

The approximative/truncated series expansions for ⟨*s*_*si*_⟩_*b*_ and ⟨*s*_*si*_⟩_*c*_′ can now readily be obtained. Combining (8.25) and (8.36a,b) we get:

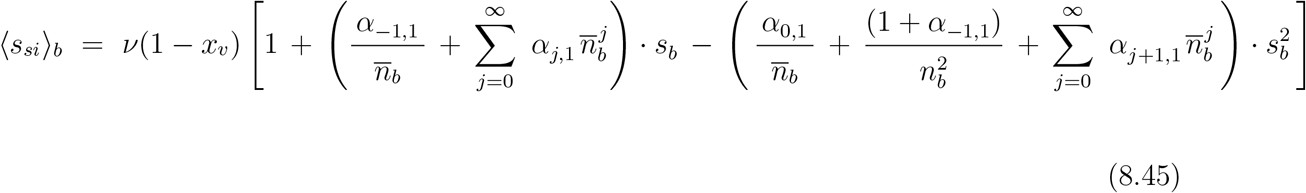

And, consistent with the condition (see Appendix 2) that for 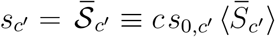:

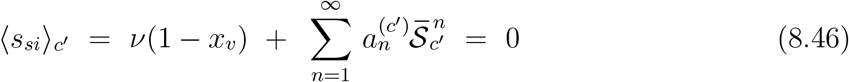

combining (8.18b) and (8.44a,b) yields for ⟨*s*_*si*_⟩_*c*_′, up to 2nd-order in *s*_*c*_′ :

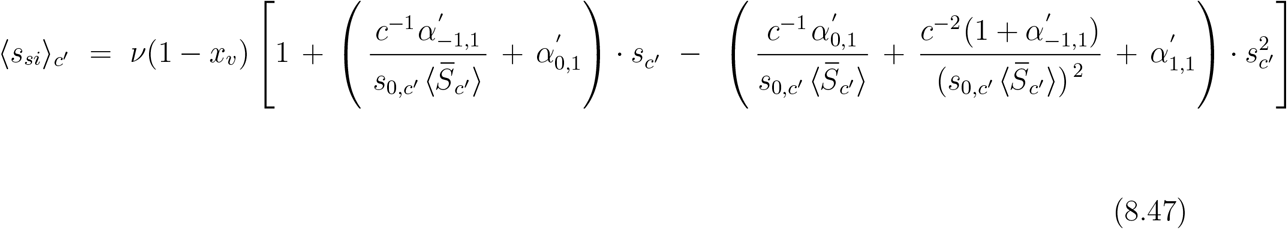

With ⟨*s*_*si*_⟩_*b*_ and ⟨*s*_*si*_⟩_*c*_′ given by (8.45) and (8.47) respectively, the sets of differential equations describing the temporal evolution of the respective infection rates of the bulk cluster (*s*_*b*_) and the secondary clusters (*s*_*c*_′) can be written as:

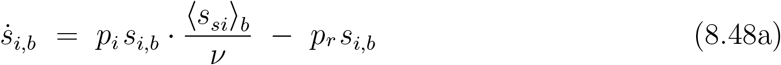

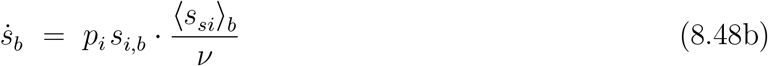

where *s*_*i,b*_ represents the contribution to the active infection rate composed of active infections in the bulk cluster, and:

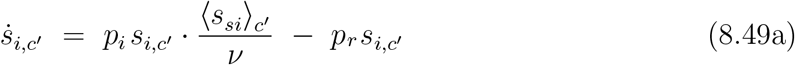

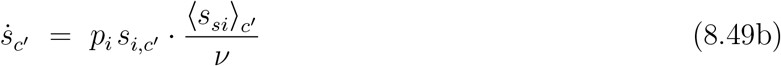

with *s*_*i,c*_′ representing the contribution to the active infection rate composed of active infections in the secondary clusters. The solutions if these differential equations are controlled by the total number of initial infections *s*_0_, and by the ratio according to which these infections are (randomly) distributed among the sites of the bulk and the secondary clusters. Consider *s*_0_ being split-up into a part *s*_0,*b*_, accounting for the initial infections that “land” in the bulk cluster, and a part *s*_0,*c*_′ that accounts for those initial infections in the secondary clusters: *s*_0_ = *s*_0,*b*_ + *s*_0,*c*_′. Since the bulk cluster is (very) large relative to *s*_0_, we can write:

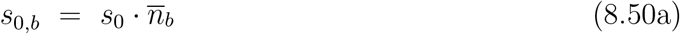

which immediately leads us also to:

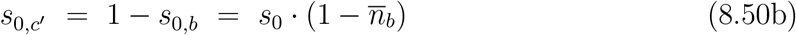

For any choice of parameters, and with (8.50a,b) serving as initial conditions, the pairs of differential equations (8.48a,b) and (8.49a,b) can be solved independently for respectively *s*_*b*_, *s*_*i,b*_ and *s*_*c*_′, *s*_*i,c*_′ via a numerical procedure, provided that *n*_*b*_ and 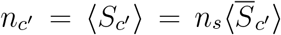 are known as a function of *x*_*v*_. The latter requirement can be fulfilled by determining the bulk-cluster size for a very large number of *x*_*v*_-values between 0 and 1, thus resulting in a dataset to be used as input from which, for any value of *x*_*v*_, proper estimates for *n*_*b*_ and ⟨*s*_*si*_⟩_*c*_′ can be obtained via interpolation (note that *n*_*c*_′ = *n*_*s*_ − *n*_*b*_).

Figs 8.6a,b respectively show, as a function of *x*_*v*_, the absolute size of the bulk cluster *n*_*b*_ and the absolute size of the combined secondary clusters *n*_*c*_′ as obtained from simulations for a population network consisting of 2D square lattice of 751 × 751 nodes with nearest-neighbour contacts (in which case the percolation threshold is approximately *x*_*c*_ = 0.41). The variation of *n*_*b*_ and *n*_*c*_′ with *x*_*v*_ clearly reflects the existence of a percolation threshold. The size of the bulk cluster decreases linearly with *x*_*v*_ at low vaccination rates. Upon approaching the percolation threshold *x*_*v*_ = *x*_*c*_ ≈ 0.41, the decrease of *n*_*b*_ with *x*_*v*_ becomes increasingly nonlinear until, as a typical feature of a percolation transition, the size of the bulk cluster vanishes completely at *x*_*v*_ = *x*_*c*_. The variation of *n*_*c*_′ with *x*_*v*_ is complemetary to this. At low values of *x*_*v*_ the combined size of the secondary clusters is almost zero. Upon approaching the percolation threshold, *n*_*c*_′ starts to increase, and at *x*_*v*_ = *x*_*c*_ there is a steep jump as a corrolary of the percolation threshold, which is then followed by a transition towards a regime of linear decrease at higher values that persists up to *x*_*v*_ = 1. Figs 8.6c,d respectively show the *relative* variations 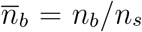 and 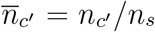 of the bulk and the combined secondary cluster sizes. The figures are quite illustrative, since they show that below the percolation threshold the vast majority of the susceptibles is in the bulk cluster, whereas for *x*_*v*_ > *x*_*c*_ the susceptibles are predominantly (if not entirely) in the secondary clusters. The simulations not only provide us with the required relation between *n*_*b*_ and *x*_*v*_ necessary as input for solving the differential equations (8.48a,b) and (8.49a,b), but also with the relation between 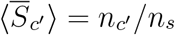 and *x*_*v*_ required in that respect (since *n*_*c*_′ = *n*_*s*_ − *n*_*b*_). The dashed curve in fig. 8.7a shows the end-value *s*_*e*_ of the cumulative infection rate *s* = *s*_*b*_ + *s*_*c*_′ as a function of *x*_*v*_ for the case *p*_*r*_ = 0, as obtained from (numerical) solutions of (8.48b) and (8.49b) (remember that *s*_*i*_ = *s* when *p*_*r*_ = 0, so that solving (8.48a) and (8.49a) for *s*_*i,b*_ and *s*_*i,c*_′ is not necessary in this case). The relevant parameters 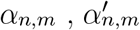 and *c* were adjusted such that the solutions of the ODE’s fit well to the *s*_*e*_ values directly obtained from the simulations (indicated by markers): it is obvious that the agreement is very acceptable indeed. To illustrate the influence of the bulk cluster size and the total size of the secondary clusters on *s*_*e*_, figs 8.7b shows the dashed curve in fig. 8.7a (obtained from the ODEs) compared to the 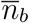 vs *x*_*v*_ curve in fig 8.6c. Similarly, fig. 8.7c shows the comparison of the dashed curve in fig. 8.7a to the average size of the secondary clusters obtained from the simulations. It is obvious that for lower values of *x*_*v*_ up to the percolation threshold, *s*_*e*_ follows the size of the bulk cluster (fig. 8.7b). At the percolation threshold the secondary clusters take over and *s*_*e*_ very accurately follows the average size of the secondary clusters (fig. 8.7c). This is perfectly in accordance with our expectations. As such, the results presented in figs. 8.6a-d and figs. 8.7a-c clearly illustrate that with the extension presented in the present section, the theoretical framework outlined in chapter 1 is also able to account for percolation phenomena in general, and even allows for decribing the effects of percolation transitions.

**Fig. 8.6:**
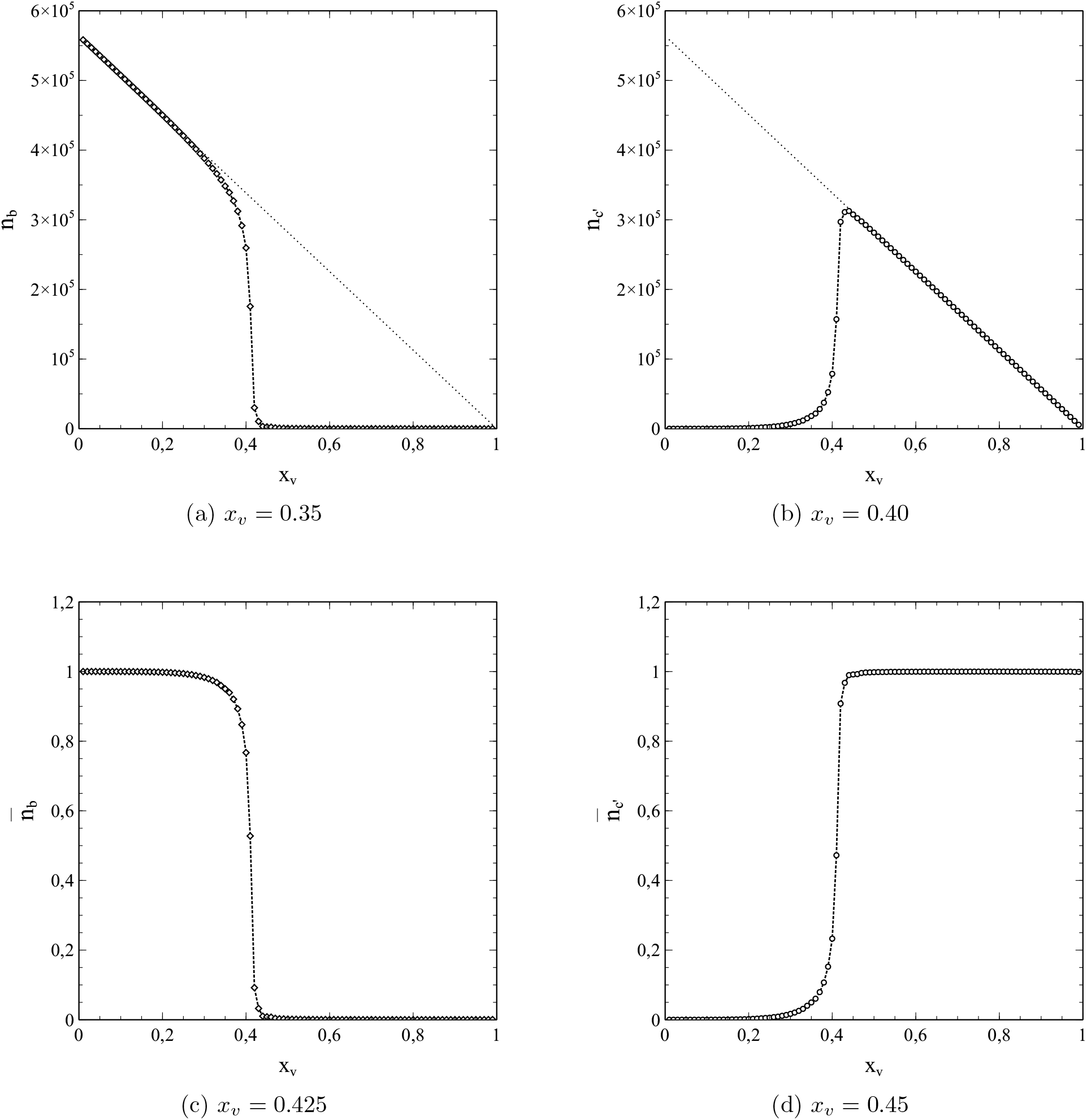
Variation of *n*_*b*_ (a) and *n*_*c*_′ (b) with *x*_*v*_ as obtained from simulations for a 751 × 751 square lattice with nearest neighbour interactions. Dotted lines represent the total number of susceptibles. Figs c) and d): resulting variations of 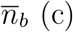 and 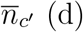 with *x*_*v*_ obtained from the data represented in a) and b) respectively.

**Fig. 8.7:**
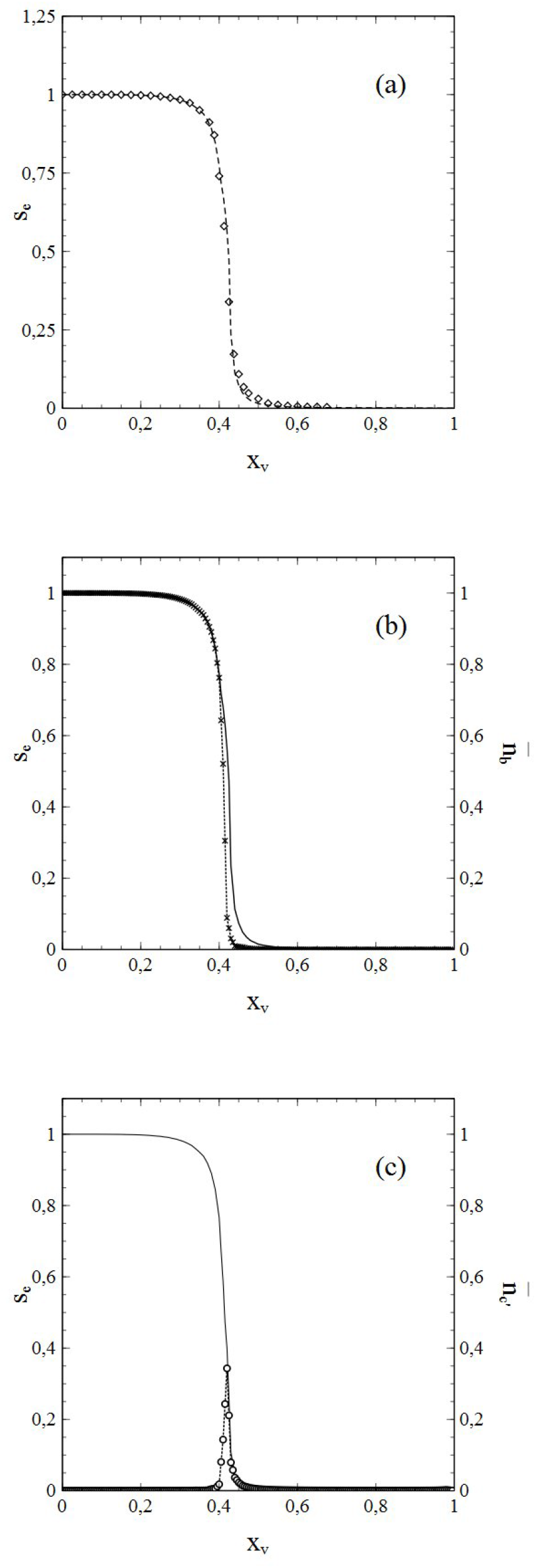
**(a)** End-value *s*_*e*_ of cumulative infection rate obtained from ODE vs *x*_*v*_ (solid curve) compared to data from simulations (markers) for *p*_*r*_ = 0 and *n*_0_ = 125. **(b)** End-value *s*_*e*_ represented by solid curve in (a) (left vertical axis) compared to 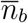 from simulations (dotted curve with markers, right vertical axis). **(c)** End-value *s*_*e*_ represented by solid curve in (a) (left vertical axis) compared to 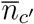 from simulations (dotted curve with markers, right vertical axis)

### e) Percolation phenomena in unvaccinated populations

The occurrence of percolation phenomena is not restricted to (partially) vaccinated populations. Depending on the circumstances, manifestations of percolation are possible in unvaccinated populations as well. Especially in cases were the the social bubble of the individuals is (strongly) reduced (reduced number of links per node if the network), percolation phenomena may play a crucial role for the patterns in which the infection spreads throughout the population, as well as for the extend to which the infection spreads. The role of percolation is expected to *in*crease when the number of links per node *de*creases. As such we have on one end of the spectrum the Ising-like networks with nearest-neighbour contacts only, where the effects of percolation are the strongest, whereas on the other end we have the extreme limit where the social bubble of an individual (node) consists of the *entire* population. The latter case is in fact “percolation free” (showing no typical percolation effects that is), since an infected node can reach any susceptible node and transmit its infection to it, no matter where this susceptible node is located within the population. One might even say that there is actually no population network in that case, and that therewith the necessary physical pre-requisite for percolation phenomena (a network with only a limited number of links/contacts per node) is in fact missing. That this represents a special case indeed is reflected in the fact that, for instance, when *p*_*r*_ = 0 the entire population will get infected in the end in such a case.

Due to the tendency to become more important when the number of the contact-links of the nodes decreases, it may be obvious that percolation is particularly an issue in connection with lock-downs. The stronger the lock-down conditions become, and therewith the limitation of the social bubbles of individuals, the stronger the influence of percolation phenomena on the spread of the infection will be. For policy makers it is important to be aware of this, since we will see that percolation effects may cause the infection rates to behave in ways that can be quite confusing, thereby posing a risk of misinterpretation in connection with critical issues like, for instance, herd-immunity.

An important manifestation of percolation effects consists of an additional reduction, with increasing cumulative infection rate *s*, of the (average) number ⟨*s*_*si*_⟩ of susceptible nodes that can be contacted by an active infection in the earliest stages of the epidemic (i.e. for (very) low *s* values). Typical examples of this phenomenon obtained from simulations are shown in figs. 8.8a-d, where ⟨*s*_*si*_⟩ is shown as a function of *s* for 4 social bubbles different in size. The social bubbles of a node thereby consist of all the other nodes in a (2*N* + 1) × (2*N* + 1) square surrounding the node (the node itself being at the center of the square). The dashed lines represent ⟨*s*_*si*_⟩ in the percolation free case (*N* = ∞) for which the standard SIR-model is exact. Figs 8.8a-d respectively correspond to *N* = 12, *N* = 8, *N* = 4 and *N* = 2. The parameters *p*_*i*_ and *p*_*r*_ gave been chosen such that *p*_*r*_*/p*_*i*_ = 0.75.

**Fig. 8.8:**
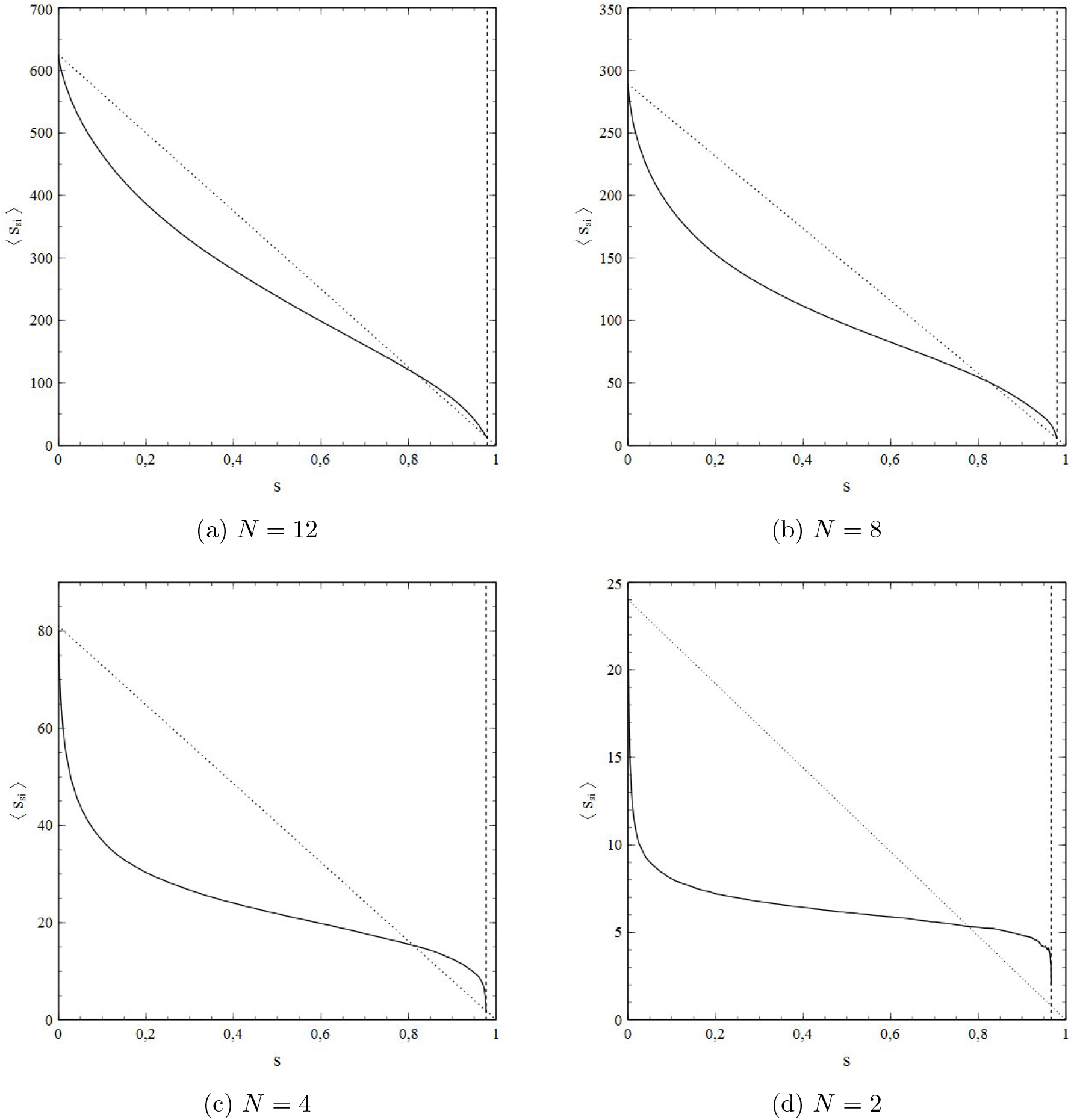
⟨*s*_*si*_⟩ for different social-bubble sizes ((2*N* + 1) × (2*N* + 1) square), dashed lines represent the percolation free case *N* = ℞ which is covered by the standard SIR model.

The effects of *N* on ⟨*s*_*si*_⟩ in fig. 8.8a-d are obvious, especially in the low-*s* regime: with decreasing *N* the reduction of ⟨*s*_*si*_⟩ in the lower *s*-regime becomes increasingly stronger. This behaviour can be understood as follows.

Let *s*_0_ be the initial infection rate at *t* = 0 so that for *t* > 0 we can write *s* = *s*_0_ + *s*′ (*s*_0_, *s*′ *«* 1). We consider both *s*_0_, *t* (and consequently *s*′) to be so small that we can neglect the probability that more than 1 infection can be found within the social bubble of each node. We implicitly assume as well therewith that *N* is small enough to rule out any overlap between the social bubbles of the initial infections. At *t* = 0 each initial infection has *ν* susceptible nodes in its social bubble. Upon infecting one of its contacts that number reduces by 1 to *ν* − 1. It is easy to see that the newly infected node has a similar number of susceptibles in its own social network (the node via which the infection was transmitted being the only non-susceptible one in that network). Writing the number of initial infections that has *not* passed on its infection at given *s*′ as 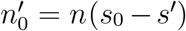 (with *n*_0_ = *ns*_0_ being the number of intial (active) infections), so that the number of initial infections that has actually infected another node can then be written as 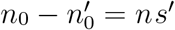, the change in ⟨*s*_*si*_⟩ upon a the change *s*′ in *s* can be written as:

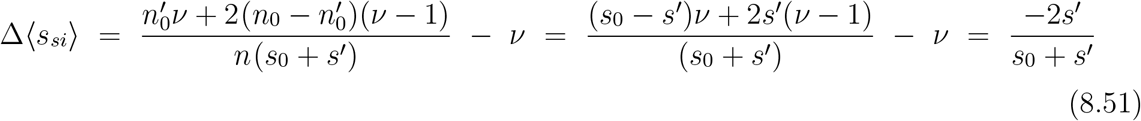

Hence:

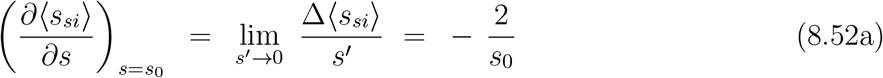

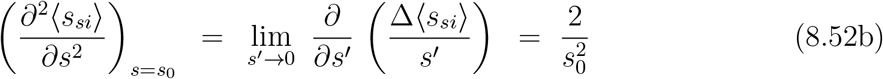

so that:

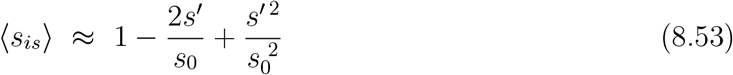

This is quite a remarkable result, which shows that the *s*-dependence (*s*′-dependence) of ⟨*s*_*is*_⟩ for sufficiently small *s*_0_ and *s*′ directly depends on the number of initial infections *s*_0_, the derivative of ⟨*s*_*is*_⟩ with respect to *s*′ for *s*′ = 0 being given by ∂ ⟨*s*_*is*_⟩ /∂*s*′ = −2*/s*_0_. Even more striking is the fact that according to (8.53) ⟨*s*_*is*_⟩ should not depend on *N* (that is, not on the size of the social bubble). To verify the correctness of (8.53) in general, fig. 8.9 shows the variation of ⟨*s*_*is*_⟩ with *s*′ in the low-*s*′ regime for *N* = 2 at 8 different values of *s*_0_, varying between *s*_0_ = 10^−4^ and 8·10^−4^. The curves show a qualitative behaviour consistent with (8.53). Moreover, the inset shows the estimated values of −∂ ⟨*s*_*is*_⟩ /∂*s*′ obtained from the slope (at very low *s*′) of the curves in the main figure (triangles), and the variation of 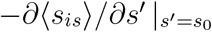 with *s*_0_ to be expected on the basis of (8.53) 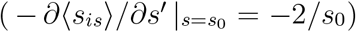, as represented by the solid curve. It is obvious that the agreement is excellent, so that we conclude that (8.53) is indeed a correct representation of ⟨*s*_*is*_⟩ at low *s*′. Hence, 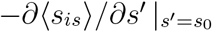 is the same for all N. However, *ν* depends on *N* (*ν* = 4*N* (*N* + 1)), so that the *relative* differential variation of ⟨*s*_*is*_⟩ with *s*′ near *s*′ = 0 varies significantly with *N* :

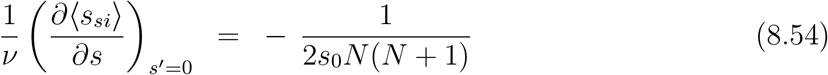

**Fig. 8.9:**
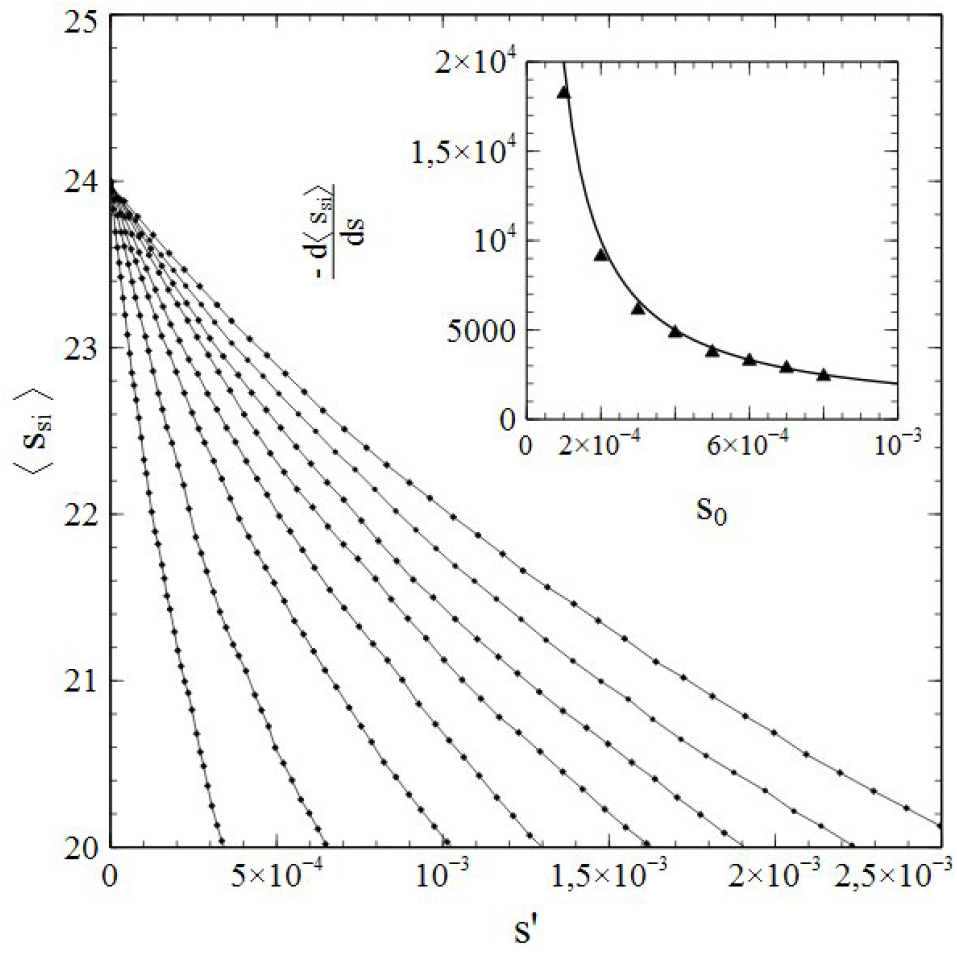
⟨*s*_*si*_⟩ as a function of *s*′ for different *s*_0_ (main figure) and the corresponding values of −∂ ⟨*s*_*is*_⟩ /∂*s*′ vs *s*_0_ (inset)

This variation becomes increasingly stronger with decreasing *N*. Hence the observed trend in the reduction of ⟨*s*_*si*_⟩ with *N* in the lower *s*-regimes in figs. 8.8a-d.

The steep drops for sufficiently low *N* in ⟨*s*_*si*_⟩ in the low-*s*′ regime of the kind shown in figs 8.8a-d (and explained on the basis of (8.53)) may have important consequences for the evolution of an epidemic. In cases of small social bubbles, and in the very early stage of an epidemic, the rate of change 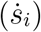 of the active infections can be expressed as (see chapter 1):

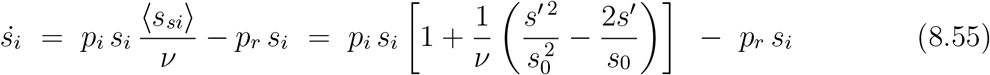

An extremum in *s*_*i*_ will occur when 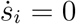. That is, when:

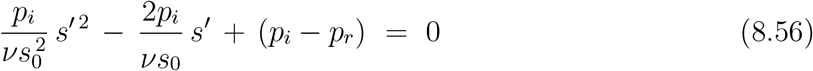

Solving for *s*′ is nearly trivial and yields:

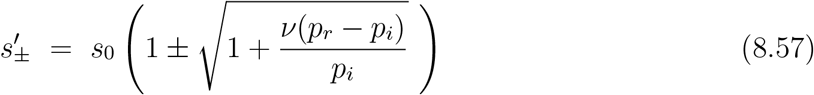

which relates to real solutions for *s*′ when:

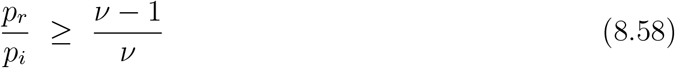

However, 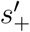 is an improper solution since 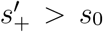, and any *s*′ > *s*_0_ is in conflict with the requirement that there is not more that 1 active infection per social bubble for (8.53) to apply (bear in mind here that *s*′ = *s*_0_ corresponds to a situation where *each* initial infection has infected exactly 1 contact in its bubble). We are thus left with 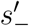 as the only solution that may apply, which requires not only (8.58) to be met, but also that 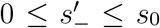. It is easy to show that the latter is the case when *p*_*r*_*/p*_*i*_ ≤ 1. So 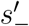 corresponds to an extremum in *s*_*i*_ when^10^:

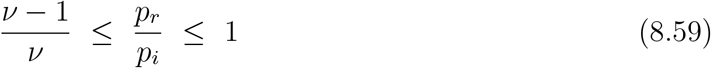

It is rather straightforward to show that this extremum is actually a maximum. We have:

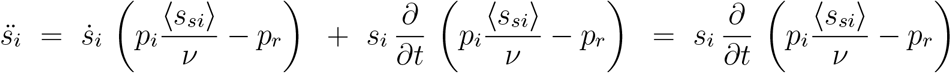

Note that both 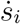 and the term in brackets here must be zero for an extremum in *s*_*i*_ (see (8.55)). We thus obtain:

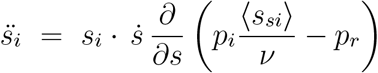

Substitution of (8.53) for ⟨*s*_*si*_⟩ then yields:

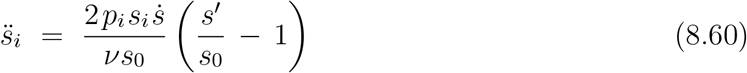

which is negative since 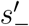 (be aware that 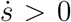). As such the extremum in *s*_*i*_ can be identified indeed as a maximum. This is a crucial observation indeed, since it implies that the number of active infections will reach its maximum shortly after the onset of the spread of the infection, when 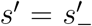 the increase *s*′ in the cumulative number of infections is even less than the number of the initial infections therewith (since 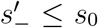). After reaching that maximum, *s*_*i*_ will then drop (sharply) again, so that the total number of cumulative infections will eventually come to a halt at a value of *s* = *s*_0_ + *s*′ which will be of the order of a few times *s*_0_ but not more. One could therefore say that under those circumstances the epidemic comes to an early stop by percolation effects alone, which smother the epidemic well before it gained any noticeable strength. However, a scenario like this is possible only for values of *p*_*r*_*/p*_*i*_ within the range given by (8.59), and is therefore limited to cases of very small social bubbles (i.e low values of *ν*). As such, the scenario may only apply to (very) tight lockdown conditions. For instance, on a square lattice with 4 nearest-neighbour contacts only (8.59) becomes 3/4 ≤ *p*_*r*_*/p*_*i*_ ≤ 1, representing a fairly narrow window of *p*_*r*_*/p*_*i*_-values. For *ν* > 4, that window is even smaller and its lower bound even closer to 1. Hence, an active implementation of the scenario, in order to prevent an outbreak from developing into a full-blown epidemic, may not only require a strong reduction of the social-bubble size (a necessary requirement) but, depending on the infecting pathogen and its typical *p*_*r*_-value involved, also necessitate bringing *p*_*r*_*/p*_*i*_ within the appropriate range (closer to 1) by additional protective measures specifically aimed at reducing *p*_*i*_ (note that *p*_*r*_, as an intrinsic property of the involved pathogen, cannot be “engineered”).

Even in those cases where percolation phenomena do *not* prevent an epidemic from passing the initial start-up hurdles, their presence may have a (significant) moderating influence on the evolution of the epidemic. The reason for this is (again) the steep decrease, as shown in fig. 8.8a-d, in the variation of ⟨*s*_*si*_⟩ with *s* that occurs for lower values of *N*. The effect of such a decrease is a significant reduction, at low *s*-values already, of the rate at which the infection is transmitted for given *p*_*i*_. As a result, the maximum for given *p*_*i*_ and *p*_*r*_ in the active-infection rate *s*_*i*_ is pushed towards lower values of *s* so that the epidemic will fade-out earlier and at a lower number of accumulative infections. This can be demonstrated as follows. With:

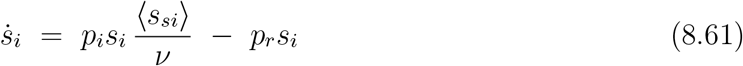

the condition 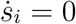 for a local extremum in *s*_*i*_ can be expressed straightforwardly as:

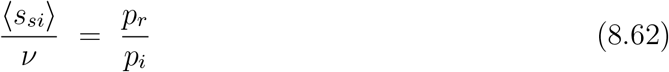

(note that 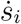 also vanishes when *s*_*i*_ = 0 but that case relates to *t* → ∞, i.e. to the fade-out of the epidemic (see section 5b)). It is easily shown that the local extrema are in fact maxima by using:

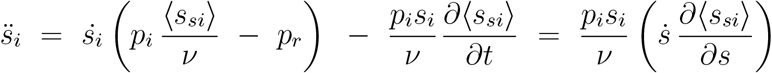

Since 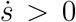 for all *t*, and ∂ ⟨*s*_*si*_⟩ /∂*s* < 0 for all *s*, we get 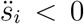 for all *t*. The condition (8.62) therefore relates to a maximum, and the active infection rate *s*_*i*_ will grow as long as ⟨*s*_*si*_⟩ */ν* > *p*_*r*_*/p*_*i*_. However, as soon as (8.62) is met and *s*_*i*_ reaches its maximum therewith, the epidemic will go into remission and gradually fades out. The steeper the decrease of ⟨*s*_*si*_⟩ with *s*, the lower the value of *s* at which (8.62) is met for given *p*_*i*_ and *p*_*r*_ will be. Consequently, it is easy to see that reducing the social-bubble size (i.e. reducing *N* or, generally, *ν*) will lead to a suppression of the epidemic itself. The root cause of this phenomenon is, at a deeper level, a percolation-related suppression of the average number of susceptible contacts ⟨*s*_*si*_⟩.

It may be obvious from (8.61) and (8.62) that also in the cases under consideration here, where the evolution of an epidemic is dominated by percolation phenomena, the ratio *r*_*p*_ = *p*_*r*_*/p*_*i*_ plays a crucial role in that evolution. All the cases that relate to the same value of *r*_*p*_ but to different magnitudes of *p*_*r*_ and *p*_*i*_ = *p*_*r*_*/r*_*p*_ are in fact equivalent. For given *r*_*p*_, the only effect of variations in the magnitudes of *p*_*r*_ and *p*_*i*_ consists of a rescaling of the time-axis. For different *r*_*p*_-values however, both qualitative and quantitative differences in the evolution of the epidemic are to be expected. To illustrate this, fig. 8.10 shows the variation of the end-value *s*_*e*_ of the accumulative rate of infections as a function of *r*_*p*_ = *p*_*r*_*/p*_*i*_, as well as the average cluster size ⟨*S*_*c*_⟩ normalised to the number of nodes in the population *n*. The data were obtained from simulations based on a population represented by a rectangular 2D square lattice of 1501 × 1501 nodes with nearest-neighbour contacts only (Ising-like case).

**Fig. 8.10:**
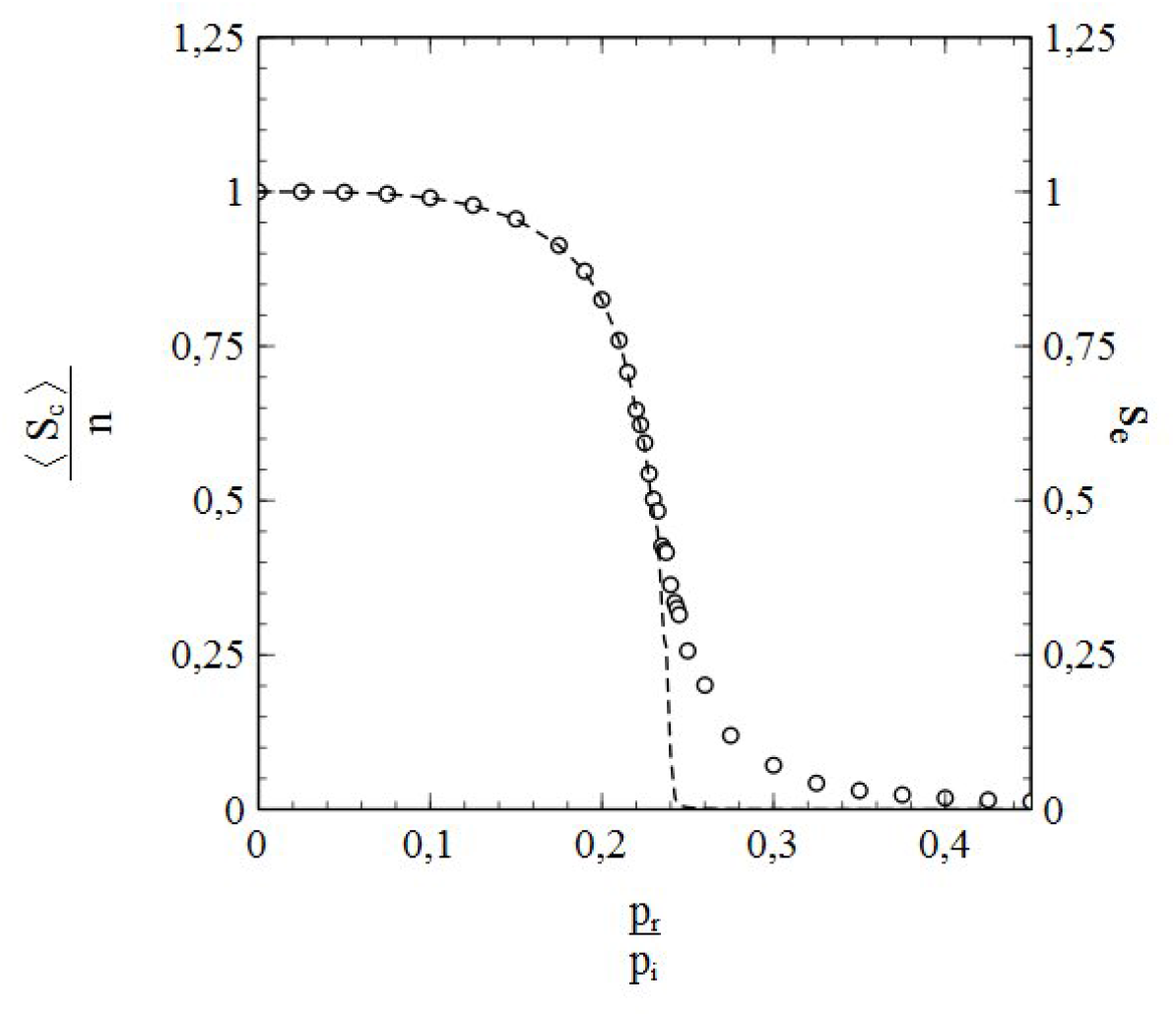
End-value *s*_*e*_ of the accumulative infection rate (open circles/right axis) and normalised average cluster size ⟨*S*_*c*_⟩ */n* (dashed curve/left axis) versus *p*_*r*_*/p*_*i*_

Not entirely unexpected, both the value of *s*_*e*_ and the average cluster size show a continuous decrease with increasing *p*_*r*_*/p*_*i*_. An interesting feature thereby is that the average cluster size steeply declines at relatively low values of *p*_*r*_*/p*_*i*_ already, and then collapses at the (relatively low) value *r*_*p*_ = *p*_*r*_*/p*_*i*_ ≈ 0.235. The value of *s*_*e*_ follows this tendency and bends down sharply with ⟨*S*_*c*_⟩, until it reaches a point of inflection at basically the same value of *r*_*p*_ that marks the collapse of ⟨*S*_*c*_⟩. It then begins a gradual fade-out towards zero, which is almost complete at *r*_*p*_ = 0.5. The low value of *r*_*p*_ at which *s*_*e*_ reaches the inflection point and ⟨*S*_*c*_⟩ collapses is particularly noteworthy. In the percolation-free case without vaccination (*ν, N* = ℞ and *x*_*v*_ = 0), the critical point at which *s*_*e*_ becomes zero (so that an epidemic spread of the infection is no longer possible) is given by *r*_*p*_ = 1 (see sections 5a, 5c). Furthermore, in case of vaccination, the percolation-free case requires a vaccination rate of *x*_*v*_ ≈ 0.75 for the critical value of *r*_*p*_ to drop to *r*_*p,c*_ ≈ 0.235 (the required vaccination rate follows from *p*_*r*_*/p*_*i*_ = 1 − *x*_*v*_ (see chapter 7)). In the Ising-like case however, such a reduction of the critical value of *r*_*p*_ is observed without any vaccinations whatsoever. Hence, in cases of small social bubbles, percolation and infection removal apparently team-up and combine their moderating effects on the spread of an infection.

It may be obvious that the inflection point in the *s*_*e*_ vs *r*_*p*_ curve and the collapse of ⟨*S*_*c*_⟩ at the same value of *r*_*p*_ are related and, in fact, symptoms of the same underlying phenomenon. This is further corroborated by the *r*_*p*_ dependence of the standard deviation of the cluster size. Fig. 8.11 shows the *r*_*p*_-dependence of √ ⟨(*S*_*c*_ − ⟨*S*_*c*_⟩)^2^⟩ corresponding to the data in fig. 8.10. A divergence emerges precisely at the critical value of *r*_*p*_ marking the collapse of ⟨*S*_*c*_⟩ and the point of inflection in the *s*_*e*_ vs *r*_*p*_ curve. The drawn curves in fig. 8.11 thereby represent a scaling law of the type (8.16) and serve as a guide to the eye. Such observations unambiguously point towards a (2nd-order) phase transition taking place at *r*_*p*_ ≈ 0.235, since we have seen in section 8b that a divergence in the standard deviation of the cluster size (i.e. a divergence in the fluctuation size) directly relates to a divergence in the correlation length (the quintessential feature of a 2nd-order phase transition). Both the collapse of ⟨*S*_*c*_⟩ and the inflection point in the *s*_*e*_ vs *r*_*p*_ curve are essential features of this phase transition as well.

**Fig. 8.11:**
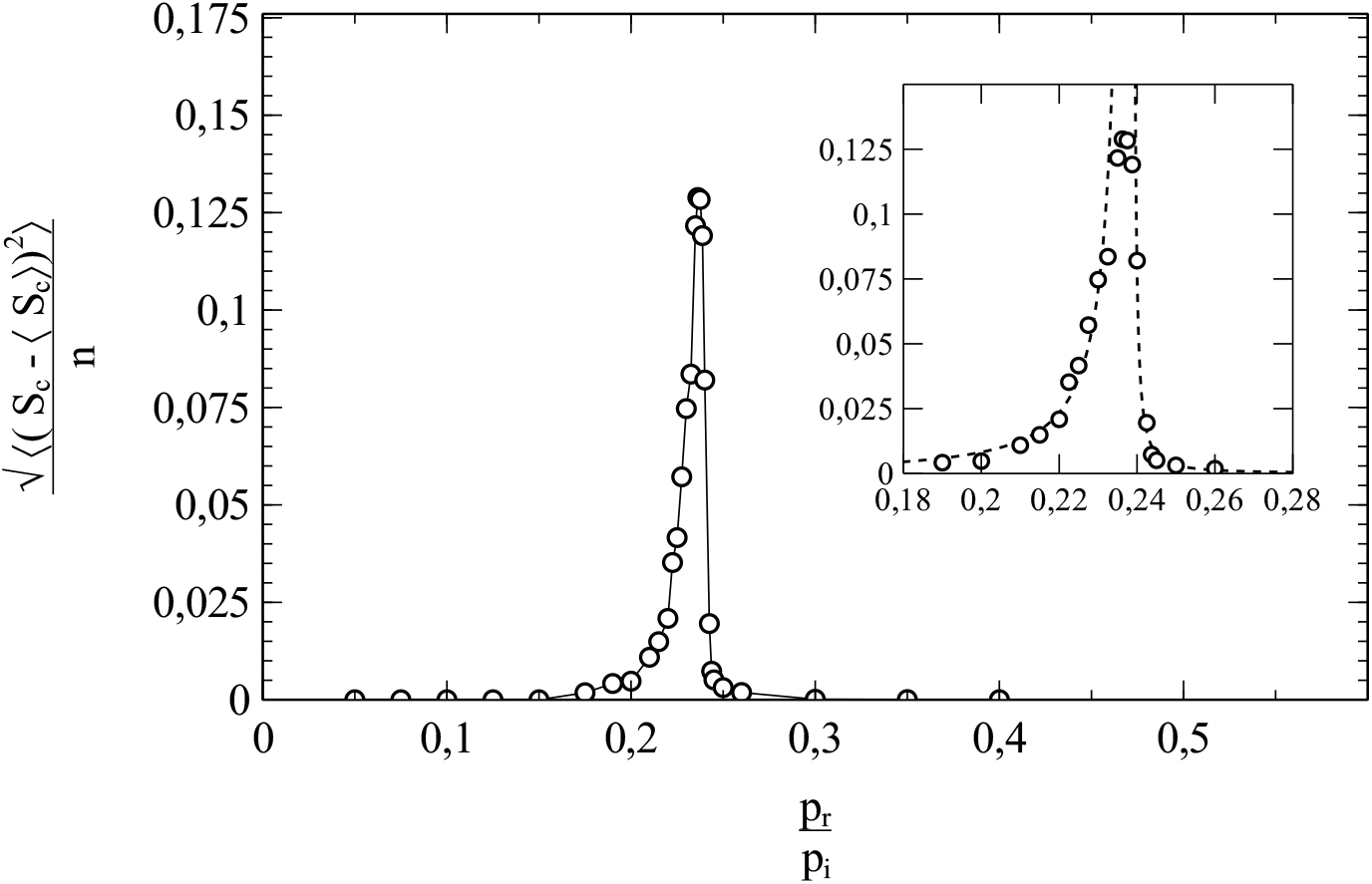
Main figure: normalised standard deviation 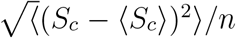 of the cluster size as a function of *p*_*r*_*/p*_*i*_. Inset: close-up of the critical region.

As to the very nature of the phase transition at *r*_*p*_ ≈ 0.235, and the mechanism driving it, it turns out that we are dealing in fact with a percolation transition in its own right. An indication for this is obtained by plotting 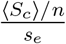 against *s*_*e*_ for a range of different *r*_*p*_-values. The advantage of the division of ⟨*S*_*c*_⟩ */n* by *s*_*e*_ is that the cluster size is related to the internal spatial arrangement of the contingent of cumulative infections (for instance, when 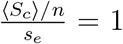 the cumulative infections form a single bulk cluster) and that more subtle details of the *r*_*p*_-dependence of ⟨*S*_*c*_⟩ are enhanced. Fig. 8.12 shows the result of such a plot for the data represented in fig. 8.10. We see that 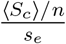 remains almost zero for low values of *s*_*e*_ and then increases quite steeply over a narrow range of *s*_*e*_-values towards a value 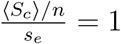, at which it then remains when *s*_*e*_ increases further. The curve seems to have an inflection point at approximately *s*_*e*_ = 0.4 (dotted line), which corresponds more or less to the middle of the narrow window of *s*_*e*_-values in which 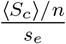 increases from almost zero to 1. The value *s*_*e*_ ≈ 0.4 that marks the inflection point is not without meaning. First of all it agrees with the value of *s*_*e*_ at the inflection point of the *s*_*e*_ vs *r*_*p*_ = *p*_*r*_*/p*_*i*_ curve in fig. 8.10. It also agrees (within a very narrow margin) with the site-percolation threshold *x*_*c*_ for a 2D square lattice with only nearest-neighbour contacts, in the case where we begin with a completely *filled* lattice and then randomly remove/block a fraction of sites *x* (for *x* > *x*_*c*_ the *remaining* or *un*blocked sites no longer percolate in that case). As a logical consequence of the latter, if what we are dealing with here is truly a percolation transition, it *has to be* a transition in the subpopulation complementary to the subpopulation of cumulative infections, i.e. a percolation transition in the subpopulation of *susceptibles*. This may seem awkward at first, since at lower values of *s*_*e*_ the cumulative infections will come in dense isolated clusters of neighbouring nodes surrounding the (single) initial infection (the one that provided the seed of the cluster), whereas the majority of susceptible nodes still forms a large bulk cluster. However, it can be shown that the phase transition indeed consists of a collapse of this very same bulk cluster of susceptibles.

**Fig. 8.12:**
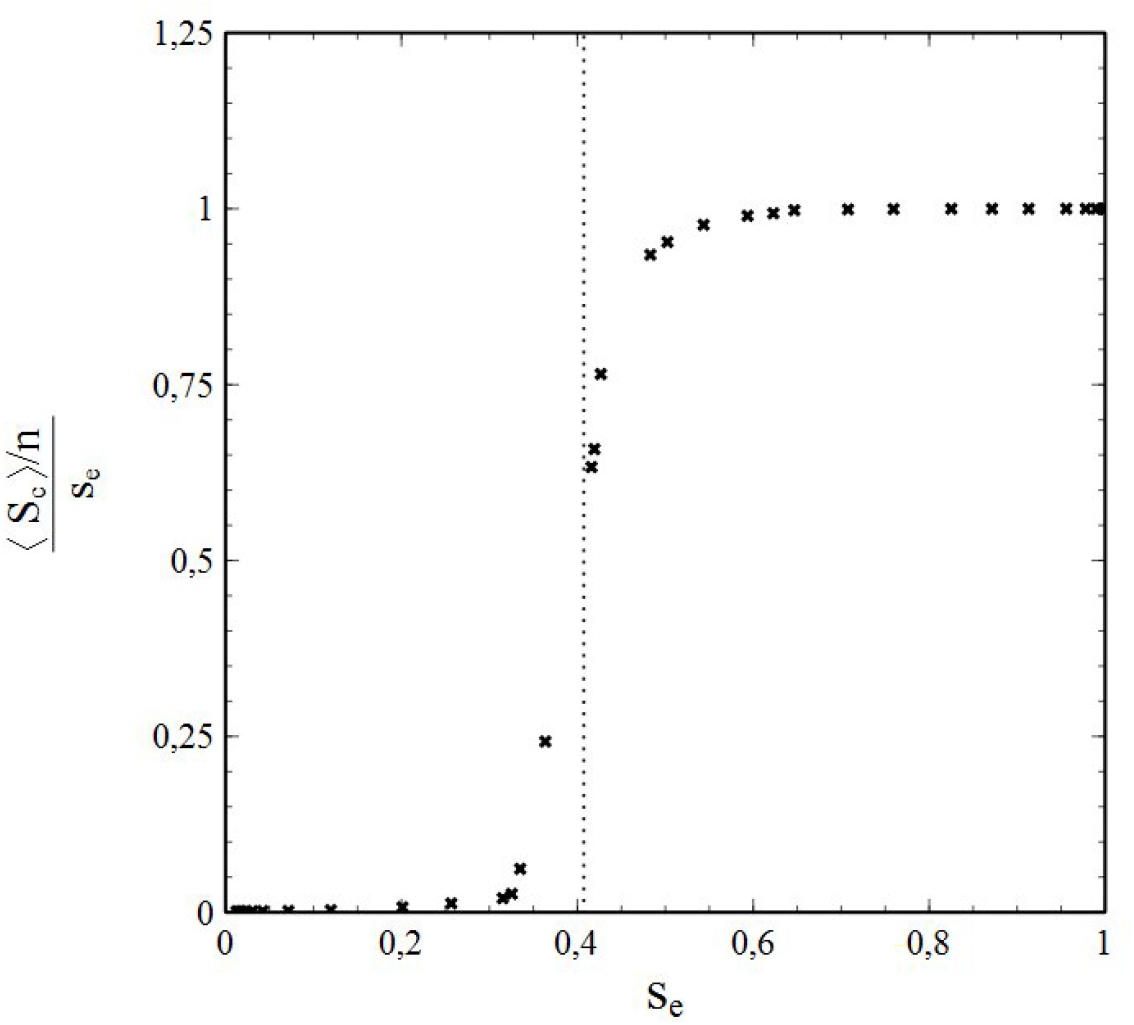
Plot of 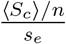 vs *s*_*e*_ (dotted line marks the inflection point)

For the ease of visualisation, we represent the 2D square lattice of nodes by a 2D lattice of square tiles (the size of a tile being that of a lattice/unit cell of the node lattice) in such a way that each node corresponds to the center of a single tile (see fig. 8.13a).

**Fig. 8.13:**
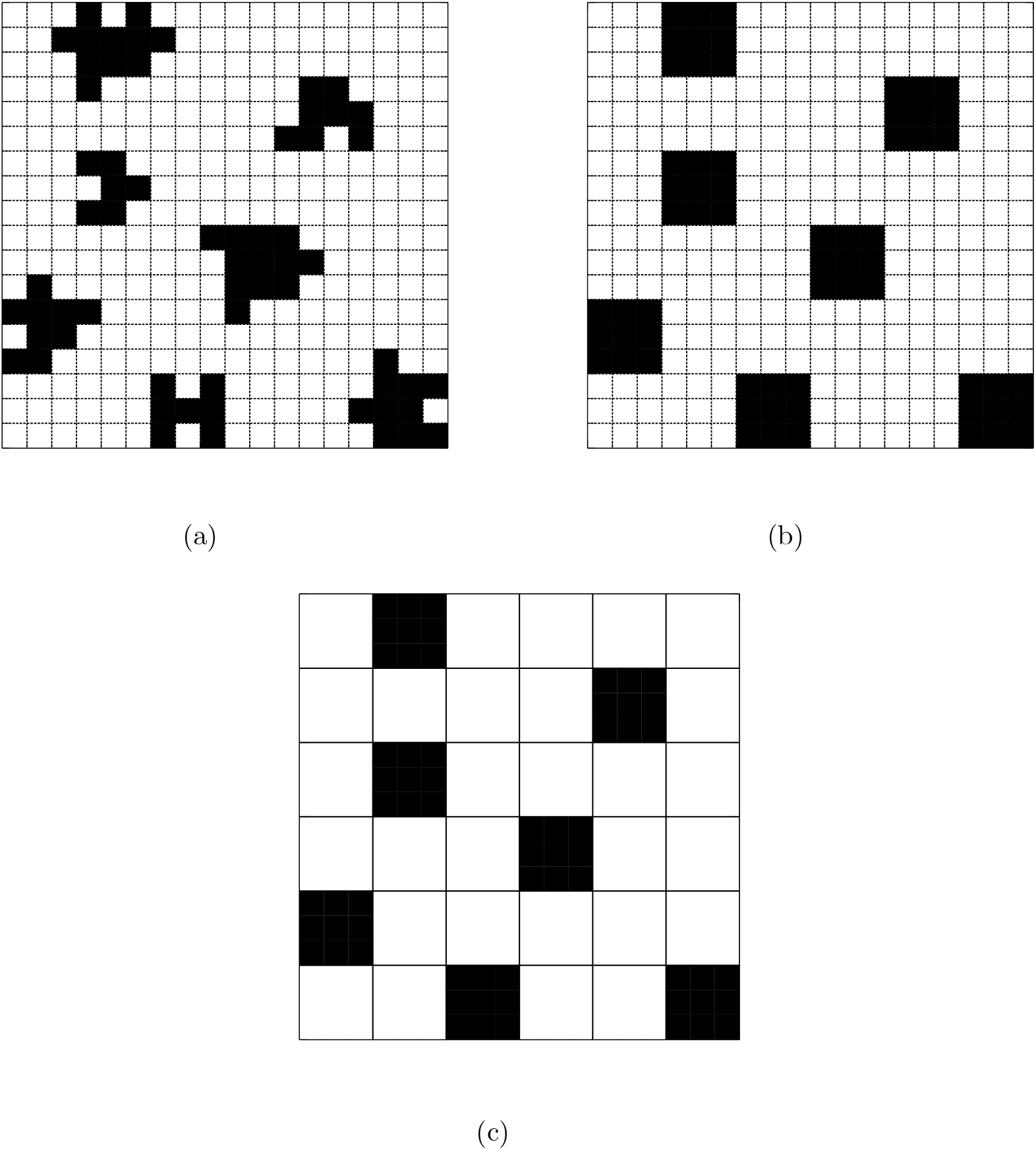
Construction of block-tiles and renormalisation of the 2D square population lattice: **(a)** actual clusters (cumulative infections) on the original (2D square) lattice, **(b)** block-tiles effectively replacing the clusters on the original lattice, **(c)** block-tiles on the renormalised (2D square) lattice

Now, as soon as the spread of the infection starts at *t* = 0, clusters of isolated (cumulative) infections begin to grow around each of the initial infections (see fig. 8.13a for a pictorial impression). The number of clusters thereby equals the number of initial infections *ns*_0_. It is also important to be aware of the fact that the distribution of initial infections is assumed to be random, and that consequently also *the distribution of the clusters over the (population) lattice will be random*.

A suitable definition of the lattice position of an *isolated* cluster is thereby given by the cluster’s “center of mass” 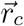:

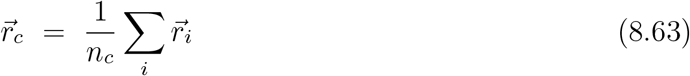

since it not only matches our intuitive notion of the concept of cluster position closely, but also makes sense from a more mathematical standpoint. The 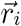 thereby represent the positions of the individual nodes (or tile centres) in a cluster, and *n*_*c*_ the number of nodes/tiles in a cluster (i.e. the size of a cluster). The index *i* runs over the individual nodes in the cluster. Note that the center of mass of a cluster is independent of the representation of the population lattice: a lattice of nodes will give the same 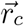 as the corresponding lattice of tiles, since the position of the nodes is coincident with the centres of the corresponding tiles. In practice, 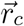 for a particular cluster of larger size is expected not to differ too much from the position 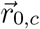 of the initial infection from which the cluster has grown (since the differences 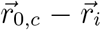 will tend to cancel out for larger cluster sizes). Once the clusters of cumulative infections have grown somewhat larger, we expect their individual sizes to differ only marginally (since all clusters grow under the same stochastic conditions), and the statistical spread in the cluster sizes low enough to consider all clusters to be of the same size as the average cluster size.

Now let us imagine that each isolated cluster of cumulative infections is being replaced by a square block of tiles (see fig. 8.13b), the number of tiles in a block being (within as close a margin as possible) equal to the average number of tiles in a cluster. The center of each square block is taken in accordance with the center of mass 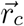 of the cluster it replaces. Next, we consider a new 2D square lattice, made-up of tiles the size of the aforementioned (cluster-replacing) square blocks. We will refer to these tiles as “block tiles” (by analogy with a conceptually similar construct called “block spins”, introduced by L. Kadanoff in his approach to order-disorder transitions in Ising spin systems [5]). A block tile is considered to be corresponding to a *single* node type only (susceptible or (cumulative) infection).

The purpose of the lattice of block tiles is to obtain a simplified representation of the original (i.e. the real) population lattice of nodes/tiles while maintaining the essential features of the actual lattice, but with less tiles (i.e. with less degrees of freedom). The positions of the block tiles related to the cumulative infections are therefore taken to be as closely as possible to the position of the square blocks of tiles introduced to replace the clusters of cumulative infections in the original lattice. The result is a 2D square lattice of block tiles with a *random* distribution of tiles representing clusters of cumulative infections in the original lattice (see fig. 8.13c).

By definition, the size of a block tile is given by the average cluster size ⟨*S*_*c*_⟩. The number *n*′ of block tiles in the new lattice and the number *n* of nodes/tiles in the original lattice are related via *n*′ ⟨*S*_*c*_⟩ = *n*, so that the ration between *n*′ and *n* is given by:

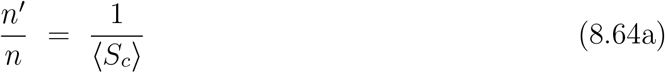

The number 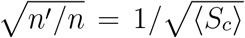 can be considered thereby as a scaling factor relating the typical length scales of both lattices (length of a tile edge, resp. block tile edge). Therefore, the block-tile concept de facto corresponds to a renormalisation transformation of the original 2D square lattice.

Furthermore, it is obvious that the fraction *s*′ = *s*_0_ *n/n*′ of block tiles representing a cluster of cumulative infections in the renormalised lattice is (and must be) *equal to the cumulative infection rate s of the population: s*′ = *s*. An explicit expression for *s*^*l*^ obtained by using (8.64a) reflects this:

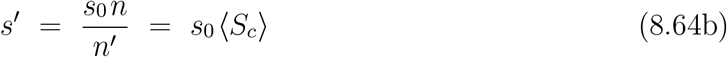

since *s* = *s*_0_ ⟨*S*_*c*_⟩ in the case of isolated infected clusters.

Together, (8.64a) and (8.64b) are the mathematical expression of a pivotal insight. With increasing cluster size (i.e increasing *s*) the total number *n*′ of (block) tiles in the renormalised lattice decreases (as expressed by 8.64a)). The number of infected tiles remains the same however, since the number of the clusters they represent is independent of *s* and equal to the number of initial infections *s*_0_ *n*. The decrease of the total number of tiles is therewith entirely due to a decrease of the number of susceptible tiles. Hence, the fraction *s*′ of infected (block) tiles in the renormalised lattice *increases* with increasing cluster size and *s*, whereas the fraction of susceptible tiles *decreases*. This is expressed by (8.64b), showing that the fraction of infected (block) tiles in the renormalised lattice is in fact equal to the fraction *s* of infected nodes (tiles) in the real population lattice, the fraction of susceptible tiles being 1 − *s*.

A legitimate question now is: will block tiles of a certain type percolate at some critical value of *s*? Since the block tiles are randomly distributed over the renormalised lattice, the answer is yes. Starting as a single bulk cluster, consisting of the entire population (except for the initial infections) when *s* = *s*_0_, the *susceptible* tiles (randomly distributed like their infected counterparts) reach their percolation threshold *x*_*c,s*_ ≈ 0.5927 on the original 2D square lattice with nearest neighbour contacts when the fraction of *infected* tiles reaches the critical value *x*_*c,i*_ = 1 − *x*_*c,s*_ ≈ 0.4073. The result is a percolation transition of the *susceptible* tiles: for *s* < 1 − *x*_*c,s*_ = *x*_*c,i*_, a large bulk cluster of susceptible tiles exists (which collapses when *s* = *x*_*c,i*_), whereas for *s* > 1− *x*_*c,s*_ = *x*_*c,i*_ the contingent of susceptible tiles is split-up into clusters of smaller size (i.e. significantly smaller than the size of the lattice). However, the crossing of the aforementioned percolation threshold by the susceptible tiles also has important implications for the *infected* tiles, since the renormalised lattice is a bit odd. Its size is *not constant* when expressed in the lattice’s very own “natural” units of area and length (given by a single block tile and the length of the edge of a block tile respectively) but varies with *s*. This is (of course) closely intertwined with the fact that the total number *n/* ⟨*S*_*c*_⟩ of block tiles in the renormalised lattice decreases a function of *s*. As a result, the lattice shrinks with increasing *s* when measured in its natural units of size. Since the number of infected tiles is constant, their average separation (measured in natural length units) must therefore decrease with increasing *s*. Therefore, when a *s* reaches a critical value when *s* = *x*_*c,i*_, a majority of the infected block tiles comes in touch with other infected block tiles, thus forming (percolating) paths of infected block tiles. This is in fact how the percolation of the *susceptible* tiles is broken. We see therewith that the critical value *s* = *x*_*c,i*_ = 1 − *x*_*c,s*_ ≈ 0.4073 marks in fact 2 percolation transitions, one in the sublattice of (cumulative) infections and one (in the opposite direction) in the sublattice of susceptibles. This can be clearly seen when we consider what happens at cluster level in the original lattice when *s* reaches *x*_*c,i*_ = 1 − *x*_*c,s*_. It is not difficult to recognise that percolation of block tiles corresponds to percolation of *clusters* in the original lattice. We thus can say that block-tile renormalisation transforms cluster percolation in the original lattice into tile percolation in the renormalised lattice. At *s* = *x*_*c,i*_ ≈ 0.4073 the majority of infected clusters merges into a single cluster, splitting-up the remaining susceptibles into smaller clusters enclosed by cumulative infections.

This can be illustrated when we compare the average size of the cluster(s) of cumulative (removed) infections ⟨*S*_*cr*_⟩ to that of the cluster(s) of susceptibles ⟨*S*_*cs*_⟩ as a function of *s*_*e*_ (the end-value of *s*) and 1 − *s*_*e*_. Fig. 8.14 shows ⟨*S*_*cr*_⟩ and ⟨*S*_*cs*_⟩ as a function of *s*_*e*_ and 1 − *s*_*e*_, normalised respectively against 1 − *s*_*e*_ and *s*_*e*_ (compare to fig. 8.12). The data were obtained from similar simulations (same parameters) as the data represented in figs. 8.10, 8.11 and 8.12. Fig. 8.14 clearly shows the symmetry involved: when *s*_*e*_ approaches *x*_*c,i*_ ≈ 0.4073 (lower horizontal scale) then ⟨*S*_*cr*_⟩ */s*_*e*_ (right scale) starts to undergo a rather steep transition, from ⟨*S*_*cr*_⟩ */s*_*e*_ ≈ 0 in the low *s*_*e*_ regime, to an almost “saturated” value ⟨*S*_*cr*_⟩ */s*_*e*_ = 1 for higher values of *s*_*e*_ (see also fig. 8.12). Parallel to this, ⟨*S*_*cs*_⟩ /(1 − *s*_*e*_) undergoes a similar steep transition in the opposite direction: when *s*_*e*_ crosses the threshold *x*_*c,i*_ ≈ 0.4073, its complement 1 − *s*_*e*_ (upper horizontal scale) crosses the percolation threshold *x*_*c,s*_ = 1 − *x*_*c,i*_ ≈ 0.5927 of the susceptible nodes and ⟨*S*_*cs*_⟩ /(1 − *s*_*e*_) (left scale) changes from ⟨*S*_*cs*_⟩ /(1 − *s*_*e*_) = 1 to ⟨*S*_*cs*_⟩ /(1 − *s*_*e*_) ≈ 0.

**Fig. 8.14:**
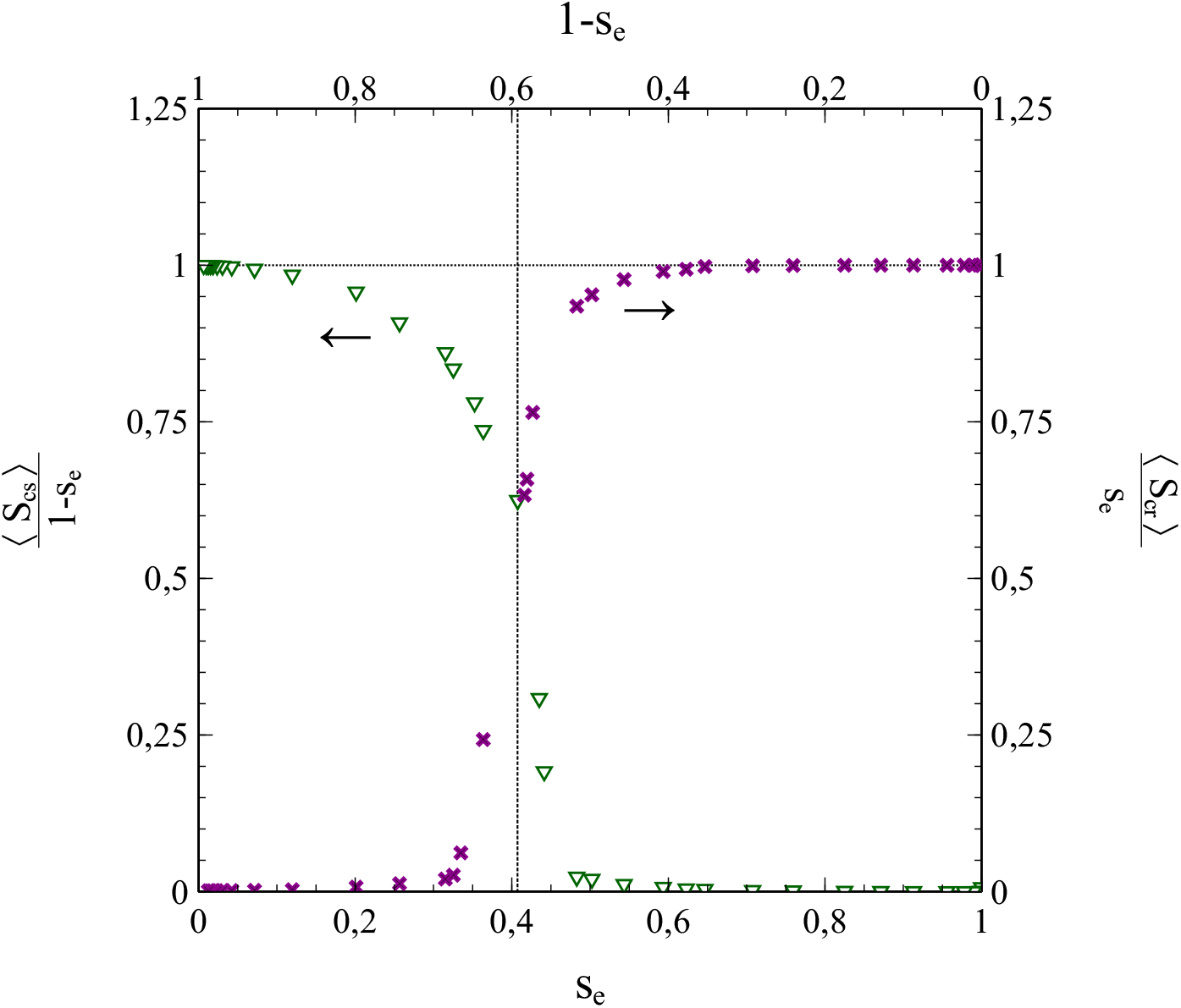
Plots of 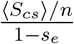 (left axis (see arrow)) and 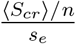 (right axis (see arrow)) vs *s*_*e*_ (bottom axis) and 1 − *s*_*e*_ (top axis)

Furthermore, in a series of pictures, fig. 8.15 shows the actual process of coalescence of the cumulative infections into a large bulk cluster (accompanied by a split-up of the bulk cluster of susceptibles into isolated smaller clusters) as it happens in reality (i.e. on the real, non-renormalised, population lattice). We see that for *p*_*r*_*/p*_*i*_ = 0.4, (fig. 8.15a) the epidemic is largely suppressed by the decay of active infections, and *s*_*e*_ does not exceed any further than a value of *s*_*e*_ = 0.0184. As a result, the clusters of cumulative infections remain very small and isolated. With decreasing *r*_*p*_ = *p*_*r*_*/p*_*i*_, the influence of infection removal diminishes, the value of *s*_*e*_ increases and the average size of the clusters of cumulative infections grows. Nevertheless, when *p*_*r*_*/p*_*i*_ decreases from *p*_*r*_*/p*_*i*_ = 0.4 to *p*_*r*_*/p*_*i*_ = 0.3 the corresponding increases in the infected-cluster size and *s*_*e*_ are still modest (a difference of Δ*r*_*p*_ = −0.1 in *r*_*p*_ results in an increase of only Δ*s*_*e*_ = 0.053 in *s*_*e*_). There is also still a bulk cluster of susceptibles in this case. However, when *p*_*r*_*/p*_*i*_ is further reduced, the effects of increased percolation take over. The size of the infected clusters and *s*_*e*_ increase more and more over ever smaller intervals of *p*_*r*_*/p*_*i*_. Ever more infected clusters merge into larger clusters, while the bulk cluster of susceptibles is gradually split-up into smaller clusters, until at *p*_*r*_*/p*_*i*_ ≈ 0.235 (fig. 8.15f) the percolation threshold has been reached and the cumulative infections form a large bulk cluster, whereas the susceptible nodes are confined to smaller (secondary) clusters. The differences between fig. 8.15e and fig. 8.15f are particularly noteworthy in this respect, since they illustrate the sharpness of the actual transition: a very minor difference in *r*_*p*_ = *p*_*r*_*/p*_*i*_ of only Δ*r*_*p*_ = −0.025 makes the difference between predominantly isolated clusters of cumulative infections, accompanied by a bulk cluster of susceptibles, and the opposite situation.

**Fig. 8.15:**
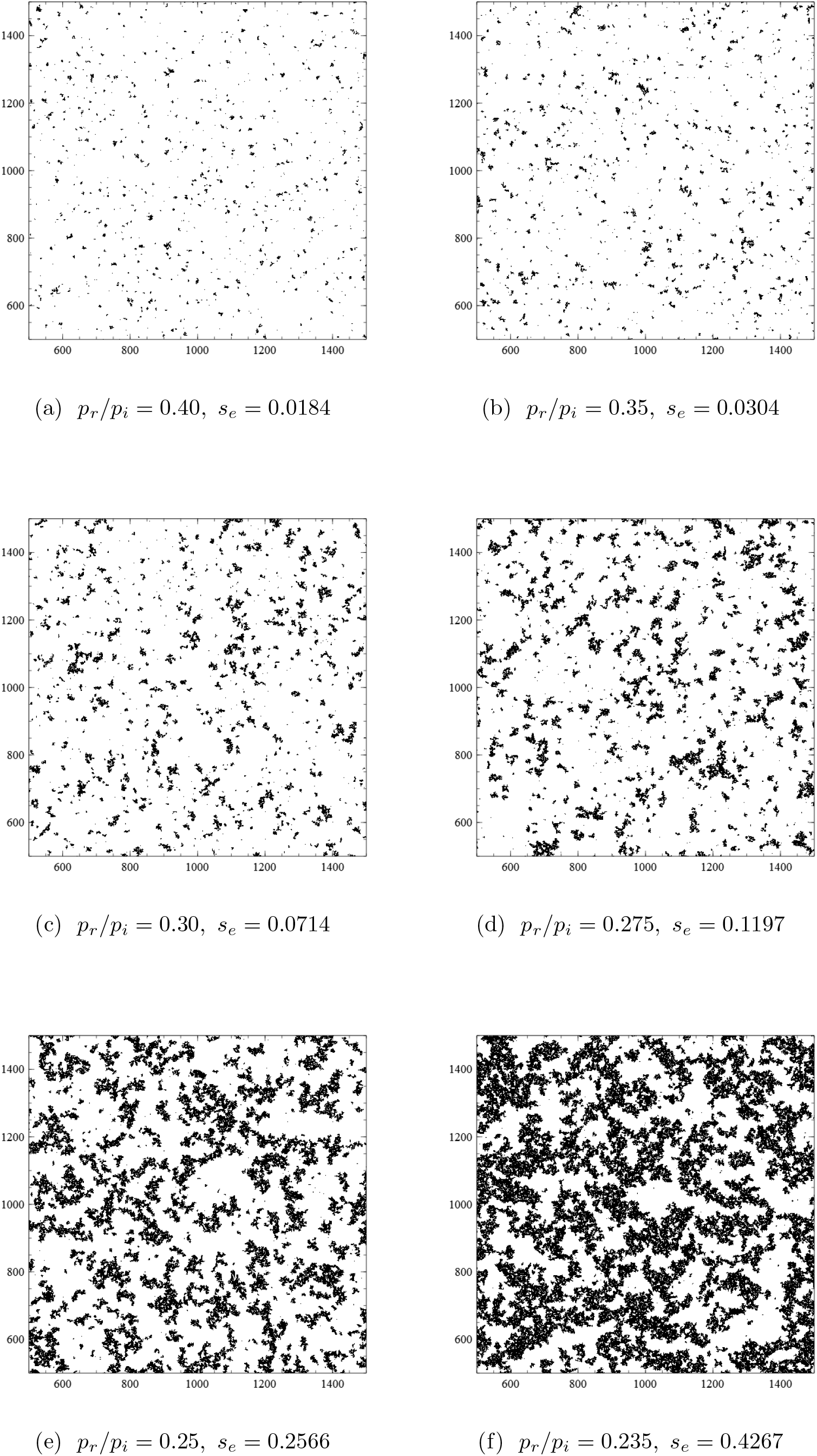
Status of the population after fade-out of the epidemic for the *r*_*p*_ = *p*_*r*_*/p*_*i*_ values and the corresponding *s*_*e*_ values indicated. Black: (cumulative) infections, white: susceptibles. The gradual confluence of the clusters of infections with decreasing *r*_*p*_ (increasing *s*_*e*_) is obvious. Case (f) represents the crossing of the percolation threshold(s).

The remaining question now is how, also at intermediate values of *s*, percolation phenomena can have such a significant effect on the production of new infections that they can compete with (and even compensate) infection removal. The answer here is in the effect of percolation on ⟨*s*_*si*_⟩ or, equivalently, on⟩ *s*_*is*_ ⟨. This is illustrated best by considering ⟨*s*_*is*_⟩ as a function of *s*. Fig. 8.16 shows ⟨*s*_*is*_⟩ vs *s* for *p*_*r*_ = 0 (so that *r*_*p*_ = 0) as its main figure. We clearly see that ⟨*s*_*is*_⟩ has a point of inflection of such a kind that ∂ ⟨*s*_*is*_⟩ /∂*s* increases with *s* for higher values of *s*. Below the point of inflection ∂ ⟨*s*_*is*_⟩ /∂*s* decreases with *s*. Numerical evaluation of ∂ ⟨*s*_*is*_⟩ /∂*s* (inset in fig. 8.16) shows that the inflection point occurs at *s* ≈ 0.4, and corresponds therewith to the percolation transition described at the foregoing pages. Apparently, the formation of secondary clusters of susceptible nodes, and their enclosure by closed “fronts” of active infections from the bulk cluster of cumulative infections, has a positive effect on ⟨*s*_*is*_⟩. This seems quite logical, since the infection now closes in on the susceptible nodes from all directions so to say. The resulting boost in ⟨*s*_*is*_⟩ directly relates to a boost in the growth rate of the rate of infections given by 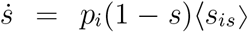 (see chapter 1). When *p*_*r*_ is large enough to keep *s* below *x*_*p,i*_, the epidemic will be unable to take advantage of this mechanism. However, as soon as *s* breaks through the percolation threshold (*s* ≥ *x*_*c,i*_) the number of (cumulative) infections will get the aforementioned boost, and so will the value of *s*_*e*_ in the end.

**Fig. 8.16:**
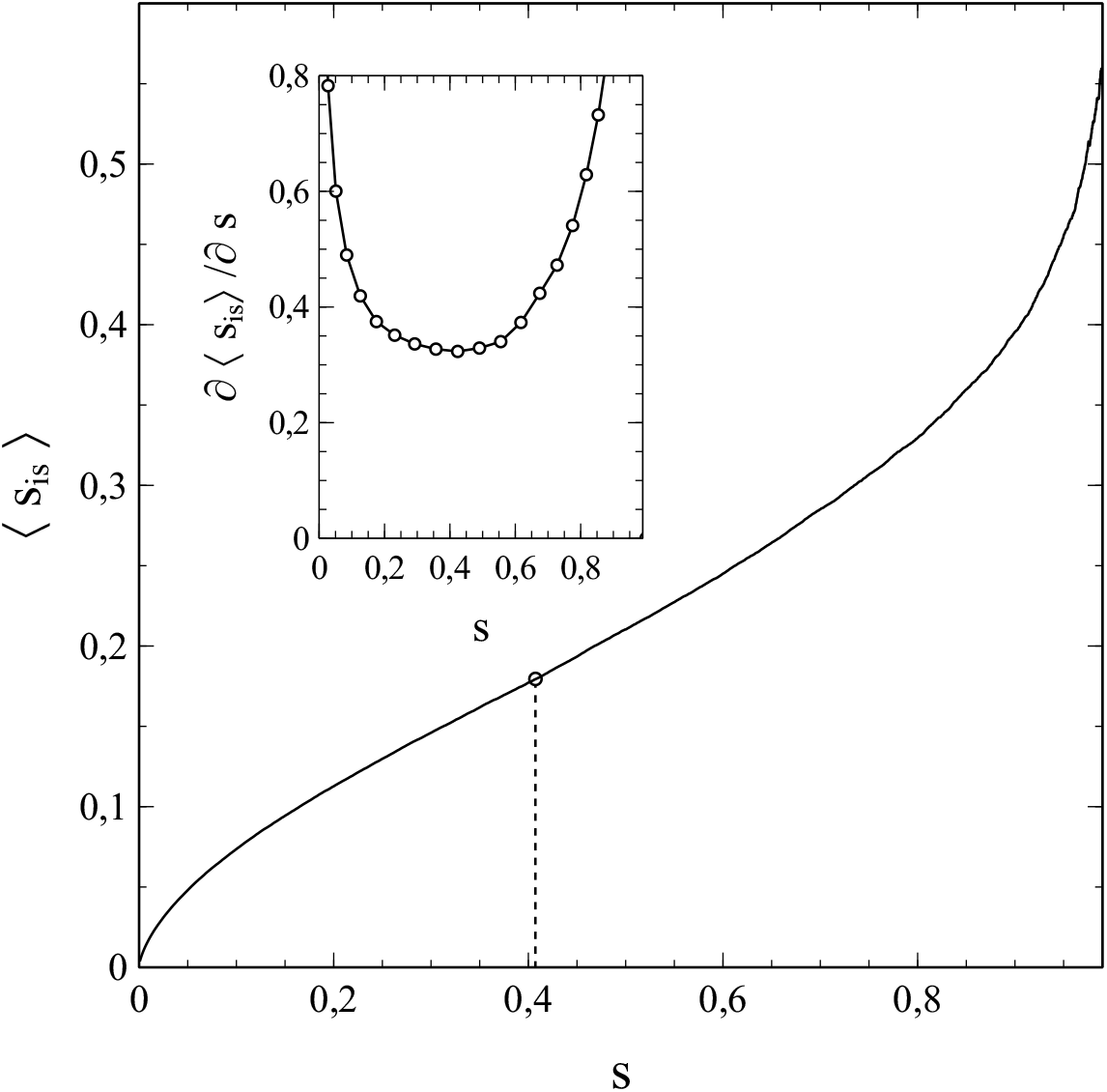
⟨*s*_*is*_⟩ vs. *s* for *p*_*r*_ = 0 (main figure) and ∂ ⟨*s*_*is*_⟩ /∂*s* vs. *s* (inset). Dot and dashed line mark the point of inflection.

The Ising case with only nearest-neighbour contacts represents a rather extreme example of contact limitation, as applied in only the strictest of lock-downs. Percolation shapes the evolution of the epidemic on par with infection removal in this case. However, the effects of percolation seem to be important under less strict lock-down conditions too, although their magnitude decreases significantly when the number of close contacts (*ν*) of the population members increases (especially in the lower range of *ν*-values). Some essential characteristics of the results for the Ising case seem to remain however, irrespective of the value of *ν*.

To give an mathematical description for what (approximately) happens in non-Ising cases we introduce the area *A*_*c*_ of a cluster, i.e. the area within the 2D population lattice that can be attributed in a meaningful way to a particular cluster. Although a concept that is easily to grasp on an intuitive basis, a precise definition of *A*_*c*_ is subject to quite some arbitrariness. A possible definition that also resonates with intuition could be that *A*_*c*_ stands for the largest area that can be fenced-in by a closed circuit of straight lines connecting nodes of a certain type in the cluster, thereby enclosing all other nodes of that type in the cluster (see fig. 8.17). It is obvious that a cluster area defined in this way generally contains nodes of both types (cumulative infections and susceptibles). We therefore introduce the filling factor *ϕ*_*c*_ of a cluster, which is the fraction of the nodes/tiles contained by *A*_*c*_ that relate to the node-type of interest. As such, the cluster size *S*_*c*_ is related to the cluster area via *S*_*c*_ = *ϕ*_*c*_ *A*_*c*_.

**Fig. 8.17:**
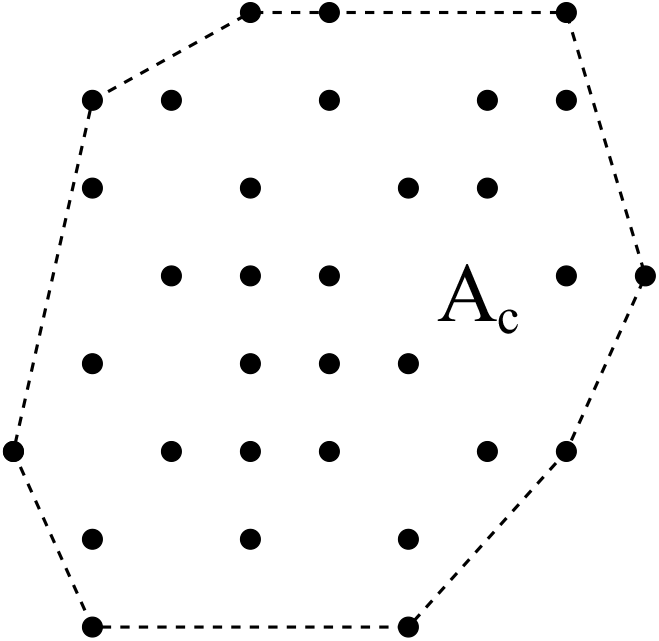
Definition of cluster area *A*_*c*_ as the largest area that can be fenced-in by straight lines (dotted) connecting elements of the cluster

We express the area *A*_*c*_ in the number of tiles in the original population lattice covered by it. Let ⟨*A*_*c*_⟩ be the average of *A*_*c*_ defined as (compare with (8.1a)):

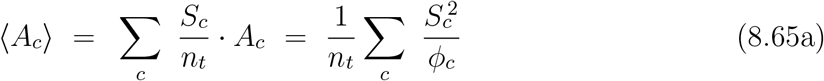

which represents an average over the size *S*_*c*_ of the clusters normalised against the total number *n*_*t*_ of nodes of the type of interest in the network, with *c* running over all clusters of the node-type of interest.

The average cluster size is given by:

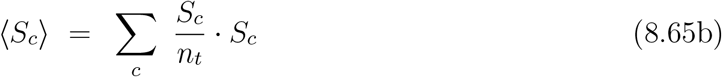

Assuming that *ϕ*_*c*_ is a constant (written simply as *ϕ*) for all clusters, combining (8.65a) and (8.65b) yields a relation between the average cluster area and the average cluster size:

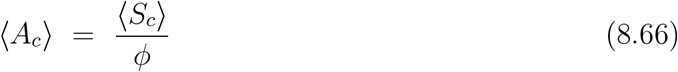

The nodes of our interest here are those related to the cumulative infections. An issue to be dealt with in this context is the distribution of the cumulative infections over the clusters that they concentrate in at low values of *s*. Those clusters start at an initial infection and then grow outwards. With time (moderate *s*) there will be a fairly high density of (cumulative) infections close to the center of mass 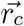 of the cluster, forming a dense nucleus of infections around 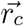. Moving further away, the density decreases however, and near the cluster boundaries only a few (cumulative) infections are present per unit of area. This is clearly shown in fig. 8.18, which pictures the simulated node-status in a population section of 700 × 700 nodes at *s* ≈ 0.24 in the absence of infection removal (i.e. *p*_*r*_ = 0) for a case where the contact bubble of a particular node consists of the nodes in a 11 × 11 square centred around that node. Black tiles/dots represent the infections, the white areas the susceptibles. The individual clusters are easily recognisable, as well as their tendency to merge.

**Fig. 8.18:**
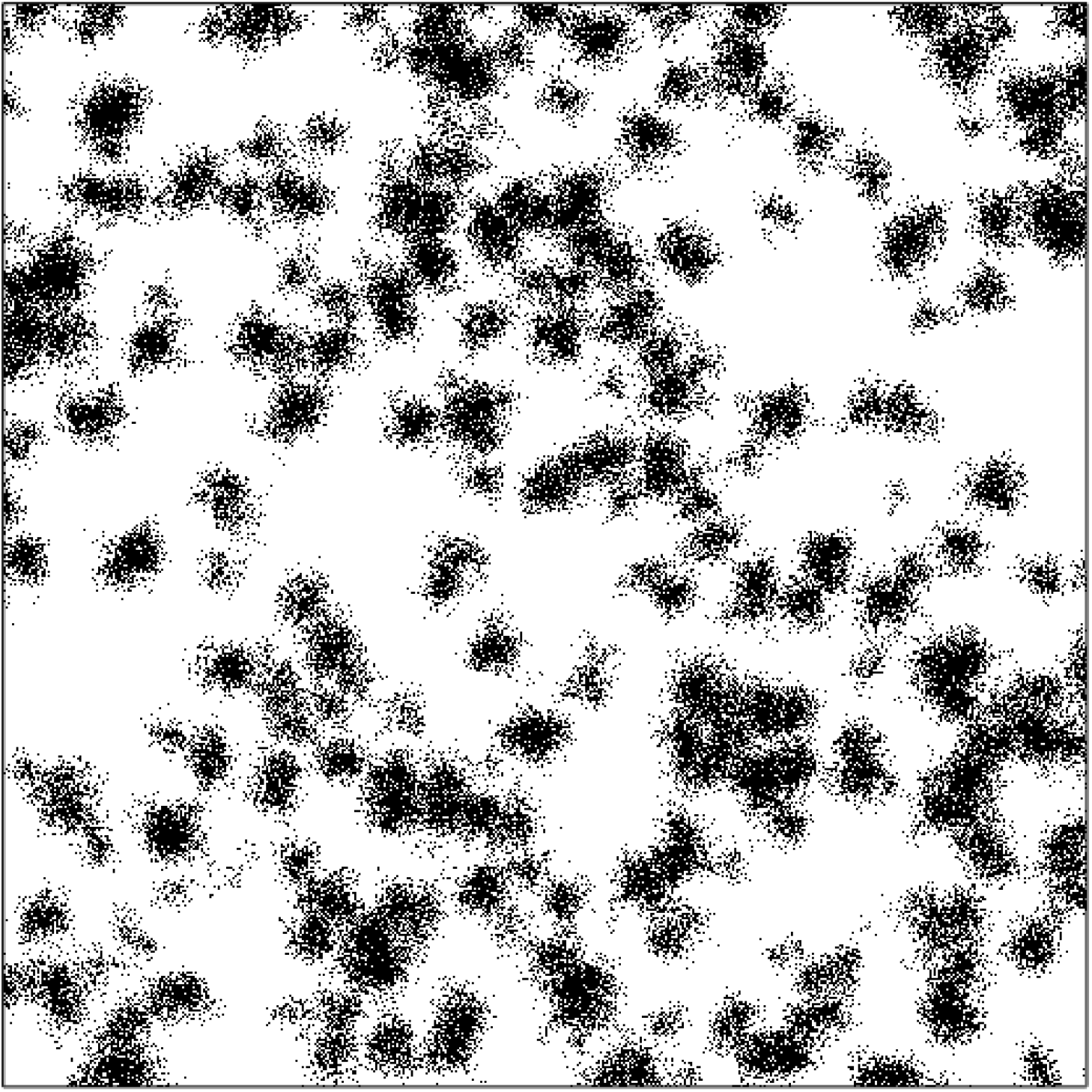
Status of nodes in a 700 × 700 section of a population (black: cumulative infections, white: susceptibles)

However, the problem is how to define and identify the confluence of 2 clusters. That is, when can we say that 2 clusters have merged? As with the definition of the cluster area, there is a degree of arbitrariness also in this matter. We cope with the issue in a somewhat pragmatic way by introducing an effective area *A*_*e*_:

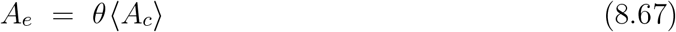

where *θ* represents a parameter between 0 and 1 (0 ≤ *θ* ≤ 1). The purpose/effect of *θ* is to lower the area that actually represents a cluster. Its value is such that 2 clusters become *indistinguishable* roughly when the distance between their respective centres of mass 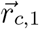 and 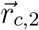 relates to *A*_*e*_ = *θ* ⟨*A*_*c*_⟩ as:

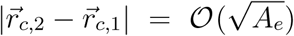

where 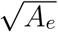 should be considered as a typical measure of the (effective) length/diameter of a cluster. Combining (8.66) and (8.67) we thus obtain for *A*_*e*_:

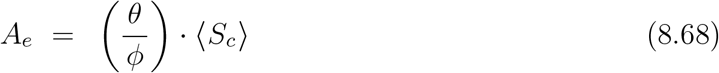

It is this effective cluster area that defines the block tiles in a newly constructed 2D square lattice: a block tile consists of a square arrangement of a number of original lattice tiles as close as possible to *A*_*e*_. The number *n* of tiles in the original lattice and the number *n*′ of block tiles in the lattice after renormalisation are related via *n*′ *A*_*e*_ = (*θ/ϕ*)*n*′ ⟨*S*_*c*_⟩ = *n*, so that we obtain:

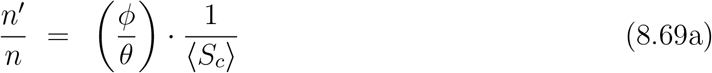

as a generalisation of (8.64a). A generalisation of (8.64b) is also obtained straightforwardly:

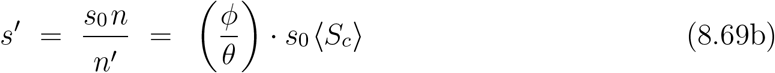

so that (with *s* = *s*_0_ ⟨*S*_*c*_⟩):

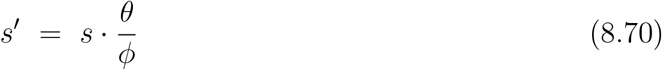

The block tiles related to the clusters of cumulative infections break the percolation of the susceptible block tiles when 1 − *s*′ = *x*_*c,s*_ ≈ 0.5927 (*s*′ = *x*_*c,i*_ ≈ 0.4073), which (as seen previously) corresponds to a collapse of the bulk cluster of susceptibles and the formation of a bulk cluster of cumulative infections. Note that by the very definition of a cluster, the existence of a bulk cluster also implies that its members are in a state of percolation.

To illustrate the influence of the size of the social bubble (value of *ν*), fig. 8.19 shows both the normalised average cluster size ⟨*S*_*c*_⟩ */n* and *s*_*e*_ as a function of *p*_*r*_*/p*_*i*_ for the case where the contact environment (social bubble) of a node consists of its nearest and next-nearest neighbours (i.e. the nodes in the (2*N* + 1) × (2*N* + 1) square with *N* = 1 surrounding the node, which itself is at the center of the square). We see that ⟨*S*_*c*_⟩ */n* and *s*_*e*_ behave as a function of the critical value of *p*_*r*_*/p*_*i*_ = *r*_*p*_ in a similar way as in the Ising case (see fig. 8.10).

**Fig. 8.19:**
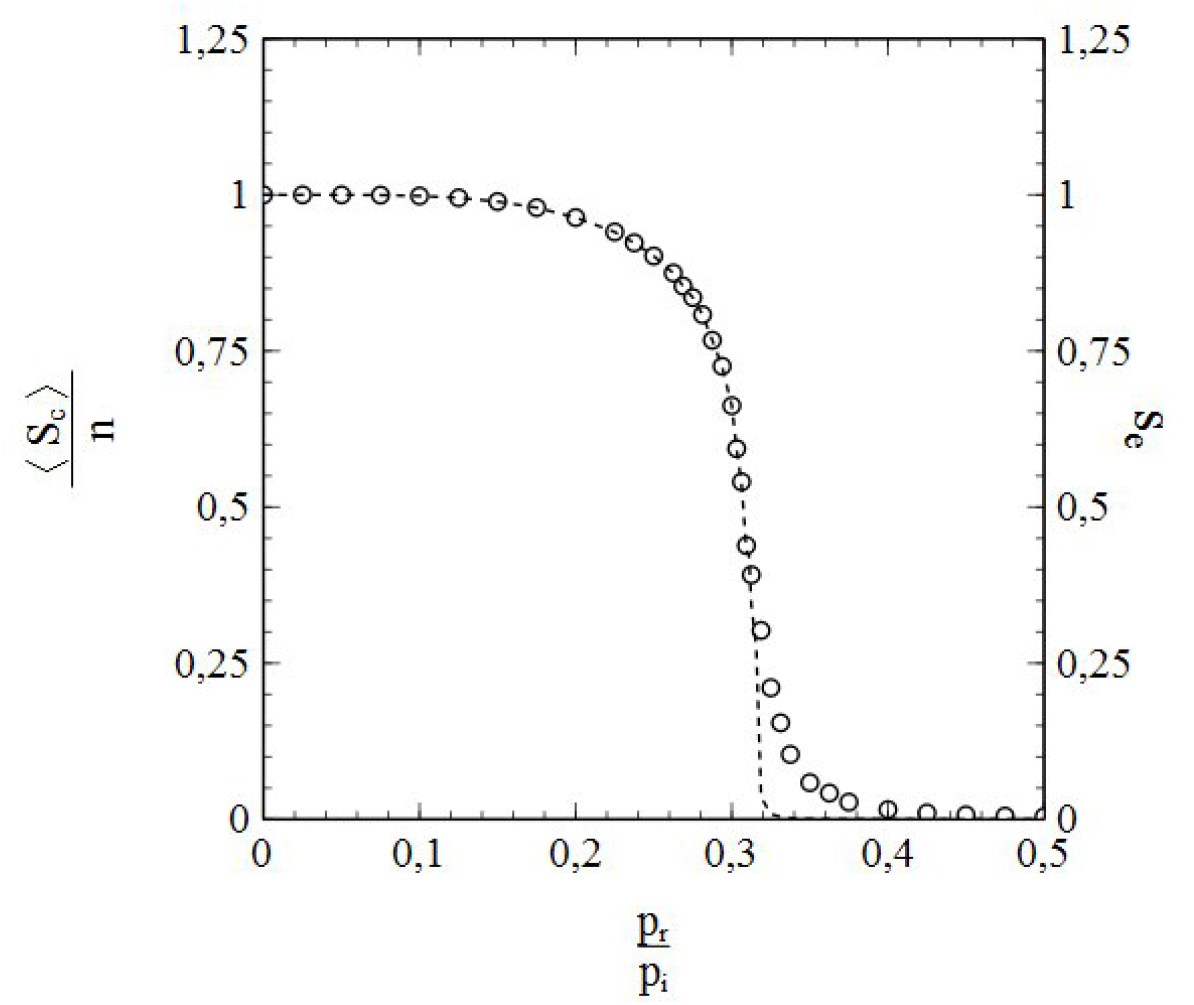
End-value *s*_*e*_ of the accumulative infection rate (open circles/right axis) and normalised average cluster size ⟨*S*_*c*_⟩ */n* (dashed curve/left axis) versus *p*_*r*_*/p*_*i*_ for a case with nearest- and next-nearest neighbour contacts.

However, compared to the Ising case, the critical value *p*_*r*_*/p*_*i*_ ≈ 0.32 that marks both the collapse of the bulk cluster of cumulative infections and the inflection point in the *s*_*e*_ vs. *p*_*r*_*/p*_*i*_ curve is higher than in the Ising case. Apparently, the effects of percolation phenomena have become less strong as a result of the changes in lattice topology and coordination number (the number of contacts per node *ν* is 8 in this case vs. 4 in the Ising case).

The qualitative behaviour of the curves in fig. 8.19 suggests that there may be a second-order phase transition involved in this case as well. This is confirmed by fig. 8.20, which shows ⟨(*S*_*c*_ − *(S*_*c*_))^2^⟩ vs. *p*_*r*_*/p*_*i*_. The critical value of *p*_*r*_*/p*_*i*_ ≈ 0.32 observed in the data in fig. 8.20 appears to be related to a divergence of ⟨(*S*_*c*_ − *(S*_*c*_))^2^⟩ */n*, which indeed provides us with conclusive evidence for a second-order phase transition taking place at *p*_*r*_*/p*_*i*_ ≈ 0.32.

**Fig. 8.20:**
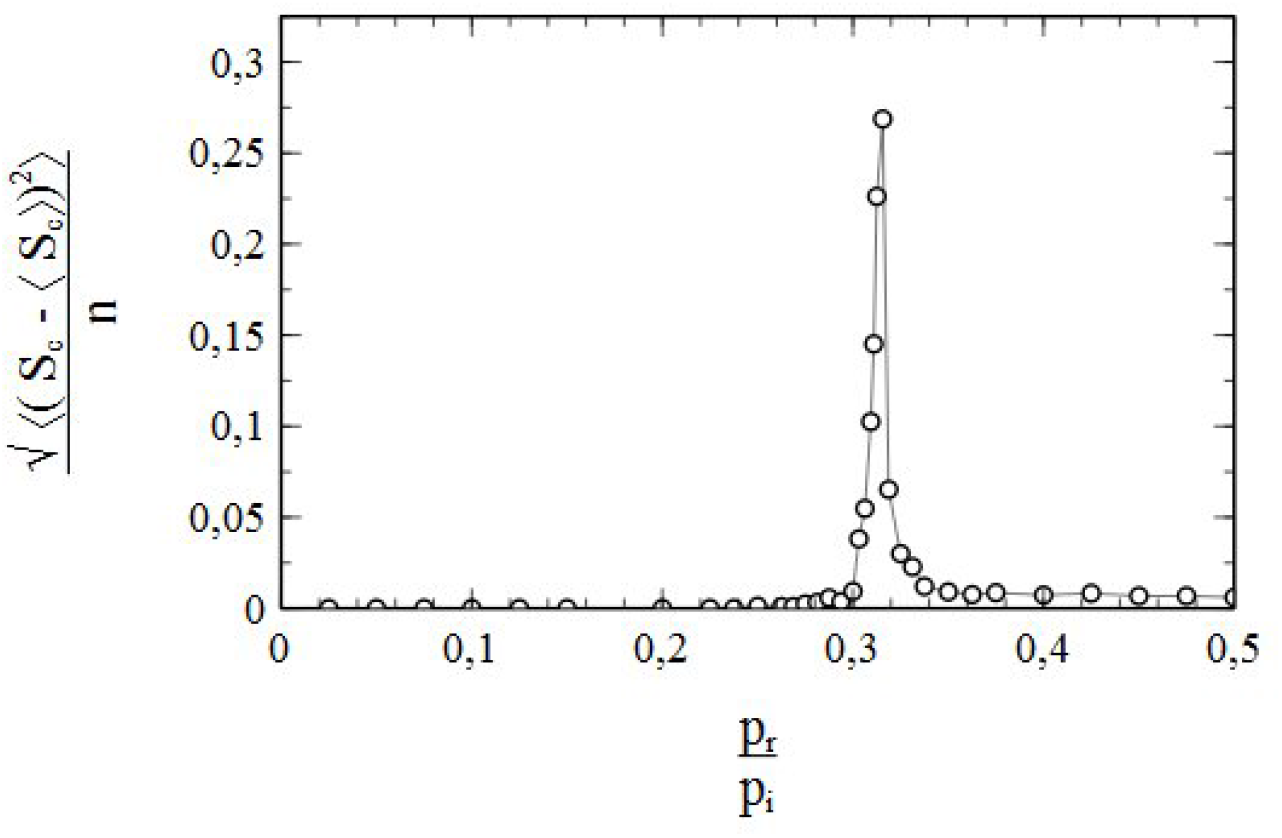
Main figure: normalised standard deviation 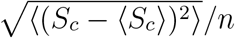 of the cluster size as a function of *p*_*r*_*/p*_*i*_ for a case with nearest- and next-nearest neighbour contacts.

Furthermore, it looks like the inflection point in the *s*_*e*_ vs. *p*_*r*_*/p*_*i*_ curve in fig. 8.19 is due to the same mechanism as the one responsible for the inflection point of a similar kind in fig. 8.10. To demonstrate this, fig. 8.21 shows ⟨*s*_*is*_⟩ vs *s* for this case. The resemblance to fig. 8.16 is obvious: like the curve in fig. 8.16, the curve in the main figure of fig. 8.21 also has a point of inflection (at approximately *s* = 0.37), and behaves similarly to the curve in fig. 8.16, both below and above *s* = 0.37. The inflection point is again to be considered as a result of the type of percolation transition that we described in the above via block-tile renormalisation of the population lattice: at *s* ≈ 0.37 the cumulative infections reach percolation while the percolation of the susceptibles is broken. As a consequence, like in the Ising case, percolation-induced boosting of the infection rate will also occur here for *s* > 0.37. Hence the inflection point in the *s*_*e*_ vs. *p*_*r*_*/p*_*i*_ curve in fig. 8.19. It is worth mentioning that apparently *θ/ϕ* ≈ 1 for this case, since the value *s* ≈ 0.37 differs only marginally from the critical value *s*′ ≈ 0.4073 that marks the percolation threshold of the block tiles representing the clusters of cumulative infections (see (8.70)).

**Fig. 8.21:**
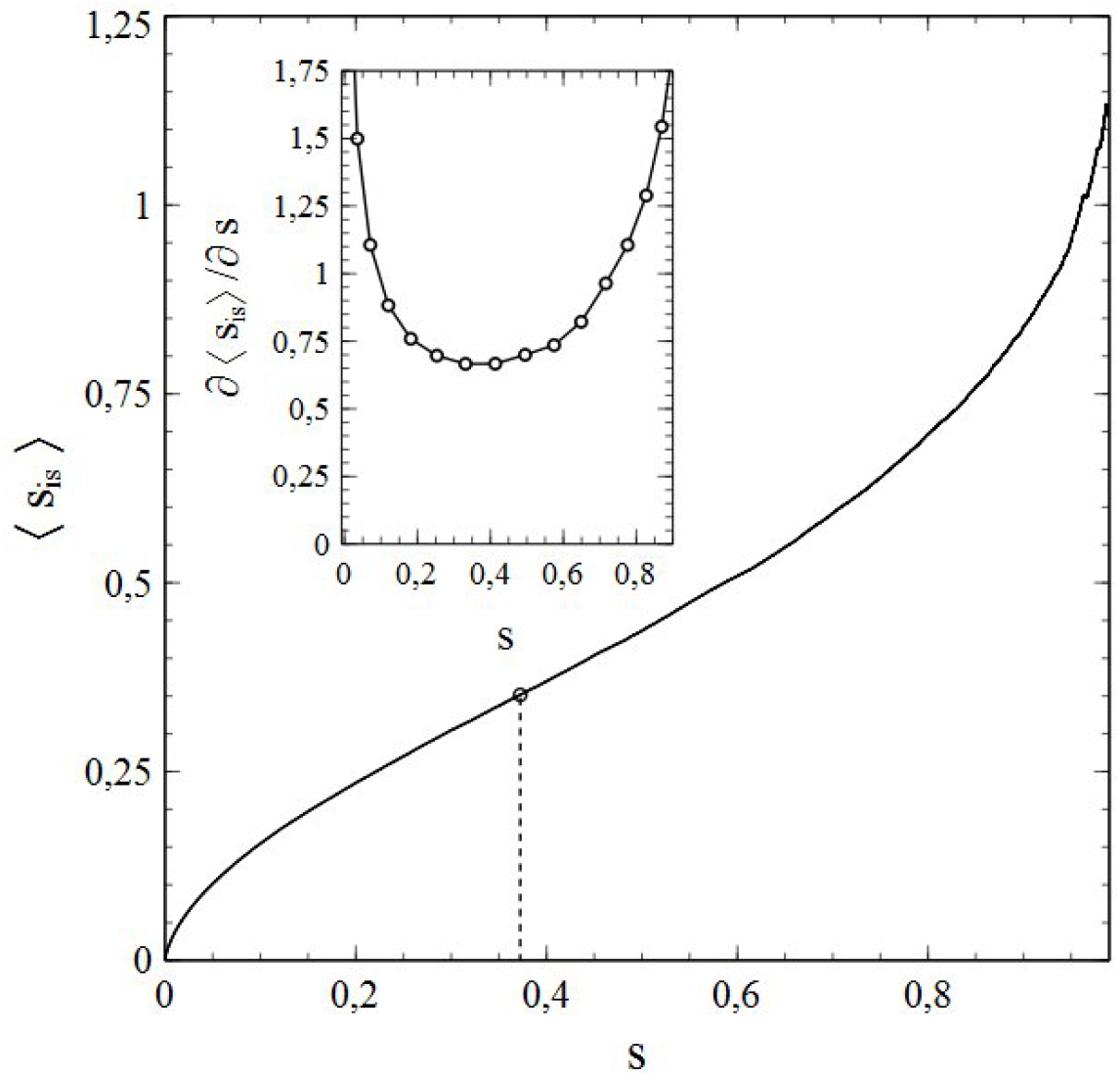
⟨*s*_*is*_⟩ vs. *s* for *p*_*r*_ = 0 (main figure) and ∂ ⟨*s*_*is*_⟩ /∂*s* vs. *s* (inset) for the case with nearest- and next-nearest neighbour contacts. Dot and dashed line mark the point of inflection.

By comparison of the data in fig. 8.10 and fig. 8.19, it is obvious (and not entirely unexpected) that the influence of percolation effects becomes smaller when the social bubbles of the members of the population increase: an increase of *ν* from 4 to 8 leads to a significant increase in the critical value of *r*_*p*_ = *p*_*r*_*/p*_*i*_ from *r*_*p*_ ≈ 0.235 to *r*_*p*_ ≈ 0.32. It is easy to see that when the social bubbles are increased ever further, this trend will remain and take the form of an asymptotic approach of the percolation-free case (*ν, N* = ℞) for which *r*_*p*_ = 1 (see section 5). In this connection it is also obvious that for all social bubbles of *finite* size the critical values of *r*_*p*_ will relate to a phase transition of 2nd order involving (and partially driven by) a collapse/formation (depending on whether *r*_*p*_ is resp. increased or decreased) of a bulk cluster of cumulative infections.

Being the limit for *ν, N* → ℞, the percolation-free case stands out against the latter. The cluster-concept has no relevance in this case, since all cumulative infections are part of one single (bulk) cluster by definition (irrespective of *S*_*c*_). Consequently, there is neither a percolation threshold in this case, nor a divergence of a correlation length (which can be considered as infinite *for all s* in fact). A parallel to thermodynamic systems with cooperative long-range interactions presents itself here. Mean-field methods give an exact description of the thermodynamic behaviour (including phase transitions) of these systems when the interaction range becomes infinite. The individual entities that make-up the system (like spins, molecules etc.) thereby interact with an effective interaction (mean field), which is described in terms of the average state of each entity in the system. As such, there is no meaningful notion of a correlation length (except that it is infinite), and fluctuations on finite length scales are averaged out without any loss of relevant information. Mean-field methods can be exact in situations with long- or infinite-range interactions between the entities in a thermodynamic system are infinite [6].

It is not difficult to see that the percolation-free case shows a strong resemblance to this picture, and that the standard SIR model represents the equivalent of an *exact* mean-field approach in this. The standard SIR model sets ⟨*s*_*si*_⟩ and ⟨*s*_*si*_⟩ equal to, respectively, the probability *p* = 1 − *s* that a node is still in a susceptible state and the probability *p*^*l*^ = *s* that a node has already been infected: ⟨*s*_*si*_⟩ = (1 − *s*), ⟨*s*_*si*_⟩ = *s*. It is easy to recognise that these substitutions are the conceptual analogue of the introduction of a mean-field. Like the mean-field methods, in most cases the standard SIR model is an approximation. However, when the social bubbles of the nodes extend to the entire population (so that the “range of social interactions” becomes infinite) these identities become exact and the SIR model gives an exact description of the evolution of the epidemic, thus accentuating the analogy between the model and mean-field methods for systems with cooperative interactions. It is evident that the similarities in concept and role between the standard SIR model and the mean-field methods for phase transitions in physical systems with cooperative interactions are in accordance with the viewpoint that the onset/disappearance of herd immunity corresponds to a (2nd order) phase transition.

### f) The SIR model and percolation in unvaccinated populations

On the basis of the previous section, the standard SIR model seems (and actually *is*, as we will see in this section) inadequate for describing percolation phenomena of the kind that we encountered in the previous section. However, the more general SIR model outlined in chapter 1 is in fact able to account for the effects of percolation in both vaccinated (see section 8d)) and unvaccinated populations. The latter can be illustrated best by using an approximation of ⟨*s*_*is*_⟩ in the form of the truncated series expansion:

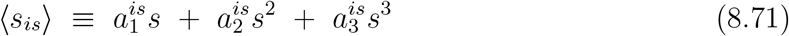

where the coefficients are fitted to the ⟨*s*_*is*_⟩ vs. *s* data represented in fig. 8.21, yielding 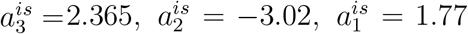. With these values taken for 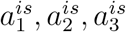, the approximation (8.71) reproduces the data in fig. 8.21 fairly well (see fig. 8.22a). The reason why an approach based on an expansion of ⟨*s*_*is*_⟩ (instead of ⟨*s*_*si*_⟩) is suited best here is twofold. First of all we have seen in the previous section that the behaviour of ⟨*s*_*is*_⟩ as a function of *s* provides a very clear indicator for the influence of percolation in both a qualitative *and* quantitative sense (inflection point). Secondly, the variation of ⟨*s*_*is*_⟩ with *s* can be approximated/described well in terms of a truncated series expansion with far less terms than the variation of ⟨*s*_*si*_⟩ with *s*. The fit of the data presented in fig. 8.21 already demonstrated that a 3rd-order polynomial in *s* provides a good approximation for ⟨*s*_*is*_⟩ in this respect. This allows for the (semi) algebraic solution of the differential equations involved, as outlined in section 3b. In principle, the parameters *t*′ and *s*′ as they occur in eq. (3.16) can thereby be chosen in an arbitrary way. However, making this choice can be somewhat tricky. As we see from fig. 8.22a, the relative difference between the actual value of ⟨*s*_*is*_⟩ obtained from simulation and the best fitting approximation based on (8.71) is quite substantial at very low values of *s* (near *s* = 0). Calculations based on (3.16) have shown that these fairly large (relative) variations in ⟨*s*_*is*_⟩ considerably compromise the calculated *s*-*t* relation and its agreement with the simulated data. We must therefore choose the point (*t*′, *s*′) at not too low a value of *t*′(that is, well within the regime where the fitted 3rd-order polynomial (8.71) does provide a good approximation (with only minor relative differences) for ⟨*s*_*is*_⟩).

**Fig. 8.22:**
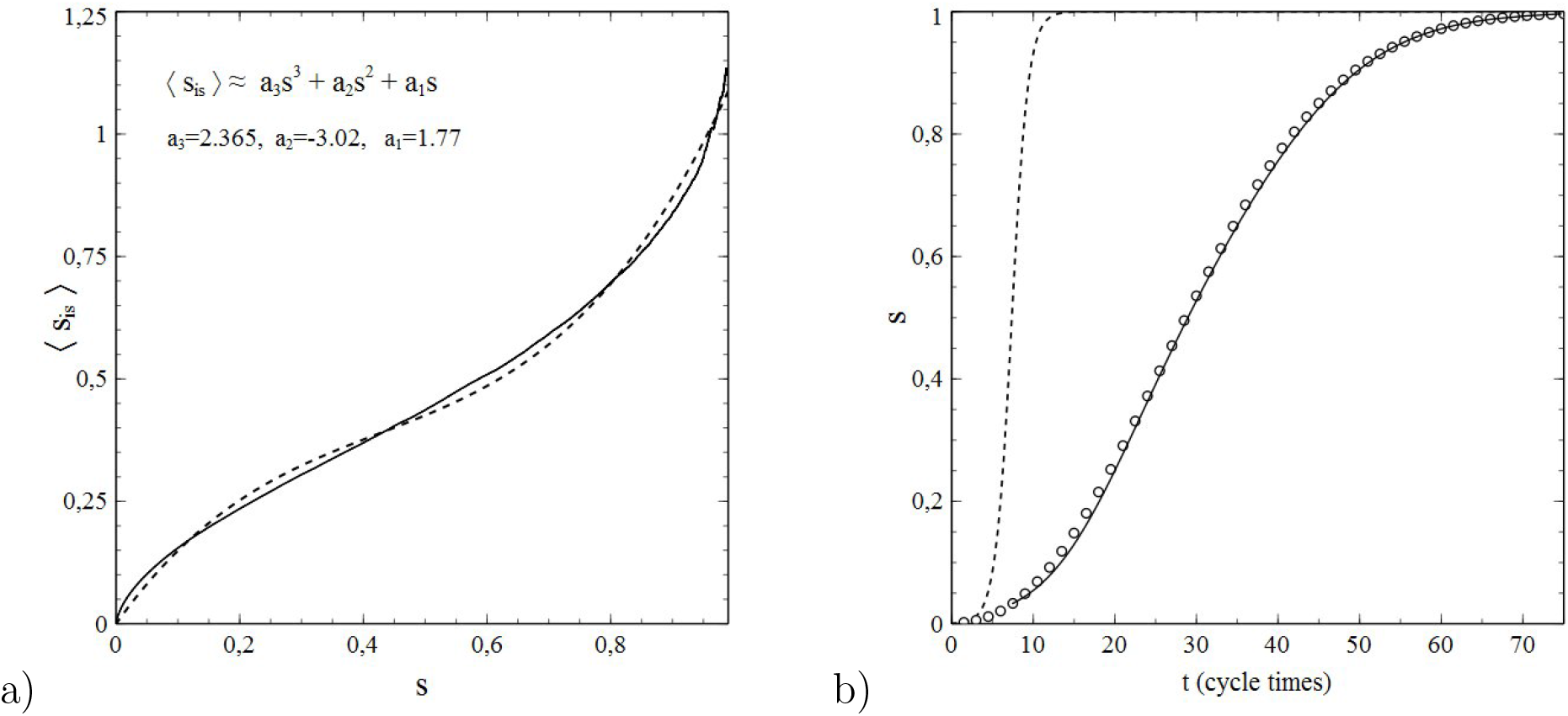
**a)** Result of a fit of the coefficients *a*_1_, *a*_2_, *a*_3_ in (8.71) to the simulated data for ⟨*s*_*is*_⟩ vs. *s* represented in fig. 8.21. Solid curve: simulated data. Dashed curve: approximation on the basis of (8.71) with best fitting *a*_1_, *a*_2_, *a*_3_ (values indicated). **b)** *s* vs *t*. Open circles: simulation, Solid curve: modified SIR model (based on eq. 3.16 under substitution of the results for *a*_1_, *a*_2_, *a*_3_ from the fit presented under **a)**), Dashed curve: standard SIR model

Fig. 8.22b shows simulated data for *s* vs. *t* along with a solution of the differential equations (3.9a,b), given by (3.16) under substitution of the values for *a*_1_, *a*_2_, *a*_3_ obtained from the fit to the ⟨*s*_*is*_⟩ vs. *s* data presented in fig. 8.22a. Time is measured in “cycle units”, each of which corresponds to a full “cycle” of *n* simulated contacts^11^, so that in one single cycle unit each member of the population makes, on the average, one single contact. The value of *t*′ chosen is *t*′ = 7.5 (in cycle units). The agreement between the solution of the modified SIR model (with fitted parameters) and the simulated data is splendid. This example emphasises therewith not only the ability of the modified SIR model to account for percolation effects rooted in lattice correlations, but also its necessity as a replacement for the standard SIR model in those cases where a strong influence of percolation phenomena is to be expected. The inadequacy of the standard SIR model is illustrated thereby by the dashed curve in fig. 8.22b, which shows *s* vs. *t* calculated on the basis of the standard SIR model for the same *p*_*i*_ as used in the simulation. The discrepancy with the simulated data and the modified SIR model cannot be missed and reveals, in fact, a complete failure of the standard SIR model for this case.

### g) Combined effects of vaccination and social-network restrictions

We now focus on situations where vaccination and percolation effects due to limitations of the social-bubble size combine, so that their effects may add-up or even strengthen one another. fig. 8.23 shows the values of *s*_*e*_ for various vaccination rates *x*_*v*_ as a function of *p*_*r*_*/p*_*i*_ for a simulated epidemic on a 2D square lattice with nearest and next-nearest neighbour contacts (*N* = 1) and vaccine efficiency ϵ = 1 (full immunity when vaccinated). All curves show a continuous transition as a function of *p*_*r*_*/p*_*i*_, from a regime of high *s*_*e*_ values to a regime of low and even negligible *s*_*e*_ values. The transitions are quite sharp (especially for low values of *x*_*v*_) and as such they display the phase-transition behaviour that we have seen in the previous sections. The data for *x*_*v*_ = 0 have already been presented in fig. 8.19, together with the variation of the average size of the clusters of cumulative infections indicative of a percolation transition. The transition points *r*_*c*_ (that is, the critical *r*_*p*_ = *p*_*r*_*/p*_*i*_) are identified as the inflection points of the curves. Note that also the maximum values of *s*_*e*_ (at *p*_*r*_*/p*_*i*_ = 0) show an expected decrease with *x*_*v*_ (which appears to be (almost) linear).

**Fig. 8.23:**
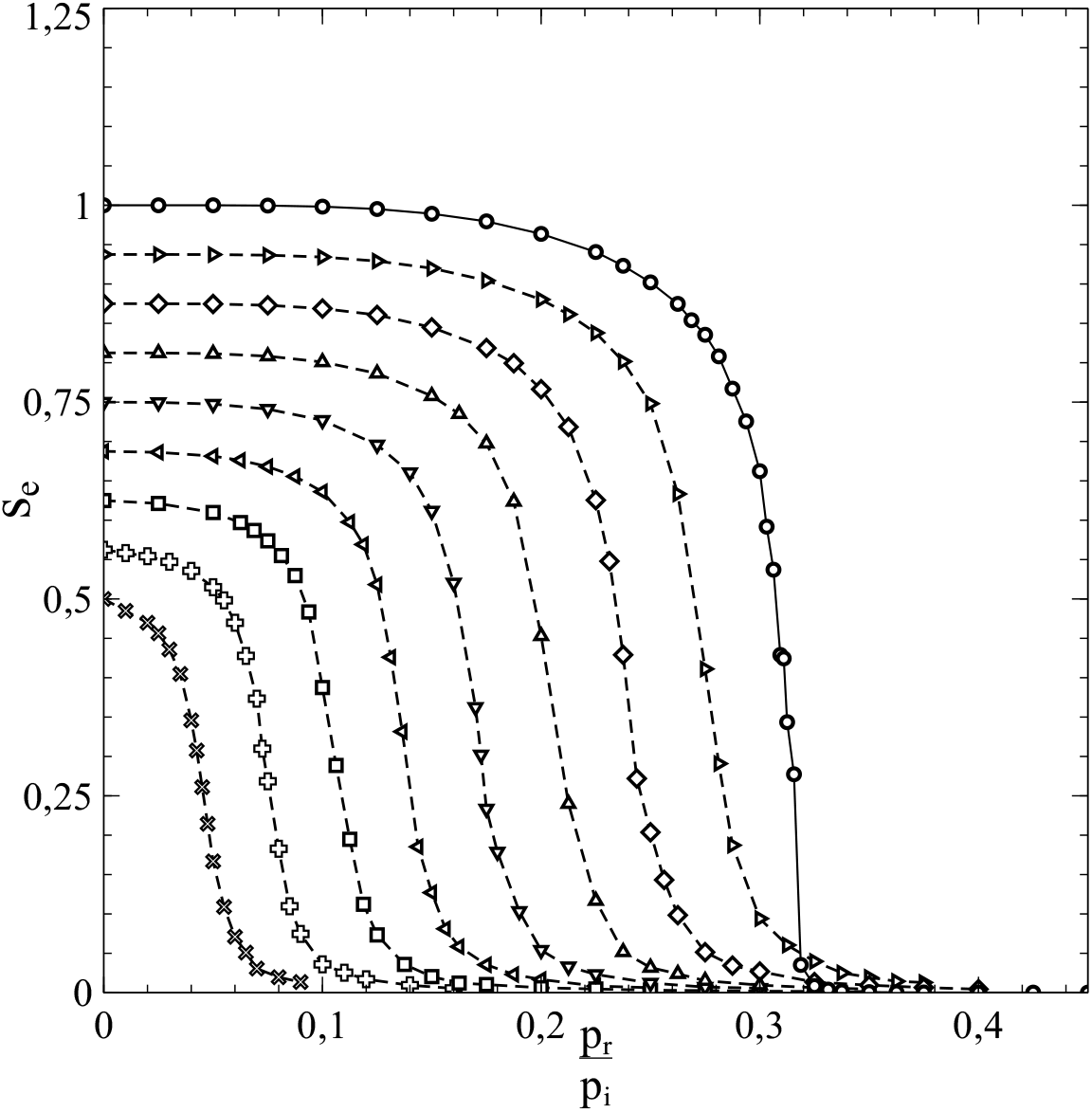
Variation of *s*_*e*_ for *x*_*v*_ = 0 (∘), *x*_*v*_ = 0.0625 (*▹*), *x*_*v*_ = 0.125 (◊), *x*_*v*_ = 0.1875 (△),*x*_*v*_ = 0.25 (▽), *x*_*v*_ = 0.3125 (◃), *x*_*v*_ = 0.375 (◻), *x*_*v*_ = 0.4375 (+) *x*_*v*_ = 0.5 (×)

Although way below the critical value *p*_*r*_*/p*_*i*_ = 1 for removal-related herd-immunity already for *x*_*v*_ = 0 (see chapters 5 and 6), the values of *r*_*c*_ become even significantly lower than that with increasing *x*_*v*_, and even approach zero when *x*_*v*_ approaches *x*_*v*_ = 0.593 *≡ x*_*p*_, which corresponds to the percolation threshold *x*_*c,s*_ = 1 − *x*_*p*_ = 0.407 for the susceptible nodes at the onset of the epidemic in case of nearest and next-nearest neighbour contacts [4]. We clearly see vaccination and the effects of size reduction of the social bubbles team-up in bringing *r*_*c*_ down and suppressing the spread of an infection. This “bundling of forces” can be understood as follows. Vaccination acts in a twofold manner by reducing the number of new infections possible per unit of time, and by blocking/reducing paths along which the infection can propagate through the population. The latter mechanism is a true percolation effect and, as such, enhanced by reductions of the social-network size.

Fig. 8.24 shows the variation of *r*_*c*_ as a function of *x*_*v*_ as obtained from the data in fig. 8.23. The variation appears to be (almost) perfectly linear and with *r*_0_ representing the value of *r*_*c*_ for *x*_*v*_ = 0 it can be described by the empirical relation:

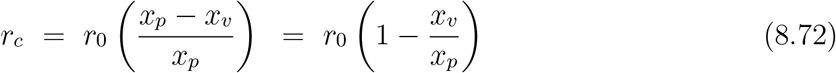

**Fig. 8.24:**
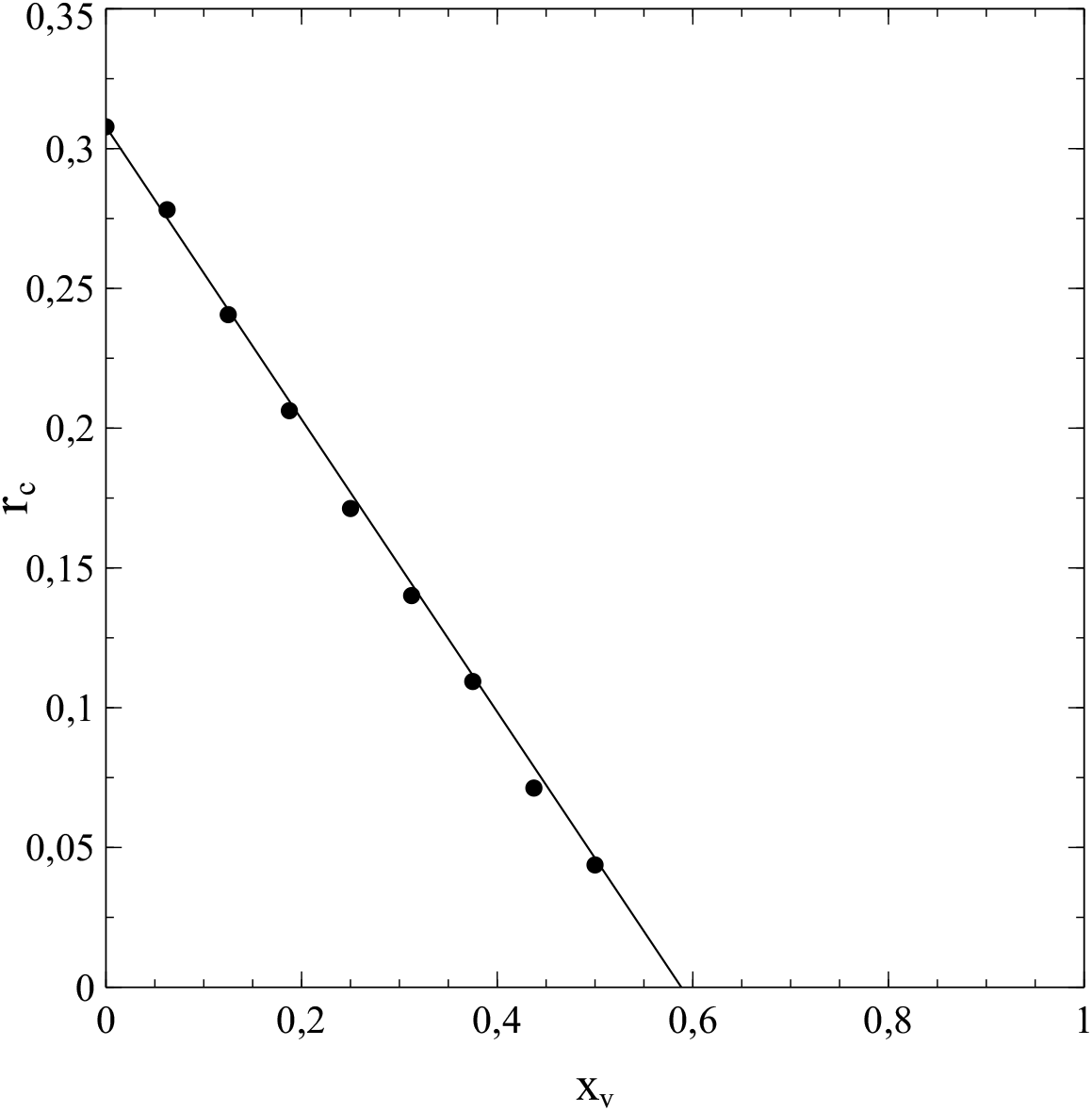
Variation of *r*_*c*_ with *x*_*v*_

This result implies the following criterion for (vaccine-acquired) herd-immunity:

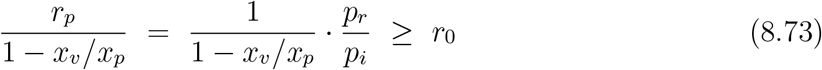

At least from a phenomenological point of view, we may consider this relation as a generalisation (for percolation effects) of the results on herd-immunity presented in chapters 5, 6 and 7 (see formula (5.1), (6.24) and, with = 1, (7.7)). It seems that a herd-immunity threshold generally exists, and takes the mathematical form of a critical value *r*_*c*_ of the ratio between *p*_*r*_ and *p*_*i*_, if we think of herd-immunity as a situation where a bulk cluster of cumulative infections is no longer possible and only a minor part of the population will get infected (both in case of strong herd-immunity under a regime of social normality and in case of weak herd-immunity under a regime of social measures (see chapter 6)). When *p*_*r*_*/p*_*i*_ > *r*_*c*_ there can be no large scale propagation of infections throughout the population. The herd-immunity threshold for *x*_*v*_ = 0 is given by (written as) *r*_0_. In general 0 ≤ *r*_0_ ≤ 1. The exact value of *r*_0_ results from the specific percolation phenomena involved, and is therefore typical of the specific (finite) size and structure of the social bubbles of the population members. As a result of vaccination, the herd-immunity threshold decreases in accordance with (8.72). Comparison of (7.7) and (8.73) suggests that the ratio 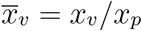 thereby acts as an *effective* vaccination rate (conceptually comparable to 𝔰_*v*_ = ϵ*x*_*v*_ in (7.7)) and may therefore be considered as just that. As such (at least when ϵ= 1) the result (7.7) may be regarded as merely a special case for *r*_0_ = 1 and *x*_*p*_ = 1 (so that 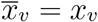) of a more general result given by (8.73).

Instead of a rigorous mathematical treatment of the observed (linear) functional relationship between *r*_*c*_ and *x*_*v*_, a simple qualitative analysis that makes the specific form of (8.72) at least plausible can be given as follows. The curves in fig. 8.8 show that, even when *x*_*v*_ = 0, a reduction of the size of the social bubbles the nodes may lead to a very strong decrease of ⟨*s*_*si*_⟩ with *s* at (very) low *s*-values. Upon increasing *s* for (very) low *N* and starting form *s* ≈ 0, a kind of plateau in the ⟨*s*_*si*_⟩ vs *s* curves is reached already at *s*-values only marginally higher than *s* = 0. Let *s*′ be a typical value for *s* in this respect. For *s* > *s*′ the change in the active-infection rate per unit of time can be written as:

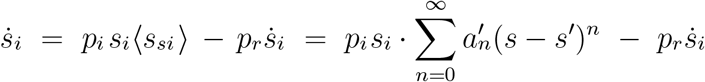

For *x*_*v*_ = 0, the coefficient 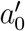 is identified as the parameter *r*_0_, so that for *s* = *s*′ and *x*_*v*_ = 0:

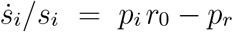

In general, 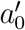 will depend on *x*_*v*_, and we write:

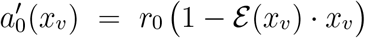

The terms ℰ (*x*_*v*_) ·*x*_*v*_ represents an effective vaccination rate (compare with 𝔰_*v*_ = *s*_*v*_ in chapter 7), where the function ℰ (*x*_*v*_) accounts for an additional decrease (due to percolation effects) of the average number of susceptibles (linked to a single active infection) that may become part of a bulk cluster of cumulative infections. In the absence of percolation effects ℰ (*x*_*v*_) = 1 so that the effective vaccination rate is equal to the actual vaccination rate *x*_*v*_, whereas in cases where percolation effects are present ℰ (*x*_*v*_) > 1, and the effective vaccination rate will be larger than the actual vaccination rate in those cases. Obtaining the exact mathematical form of the function ℰ (*x*_*v*_) is a difficult problem. However, *x*_*v*_ = *x*_*p*_ marks the threshold beyond which there will no longer be a bulk cluster of cumulative infections. Hence 1 − ℰ (*x*_*p*_) · *x*_*p*_ = 0, so that we obtain ℰ (*x*_*p*_) = 1*/x*_*p*_. Consequently, when we consider ℰ (*x*_*v*_) to be a differentiable function of *x*_*v*_ for 0 < *x*_*v*_ < *x*_*p*_, we can express ℰ (*x*_*v*_) as a series expansion in *x*_*v*_ − *x*_*p*_ of the form:

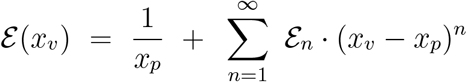

in order to account for *x*_*v*_-dependence of ℰ (*x*_*v*_) explicitly. However, fig. 8.24 suggests that, apparently, the contribution to ℰ (*x*_*v*_) due to terms of the order *n* ≥ 1 is very modest and even negligible for all *x*_*v*_, and that only the 0*th*-order term 1*/x*_*p*_ of the series expansion prevails. This then leads us straight to the empirical relation (8.72).

### h) Herd-immunity and percolation

It may be clear from the previous sections in this chapter that percolation phenomena are a contributing factor to the achievement of herd-immunity. Even in unvaccinated populations the critical value of *r*_*p*_ = *p*_*r*_*/p*_*i*_ (beyond which the cumulative infection rate will stick at a very low value) is significantly lowered when the social bubble size is reduced. This is clearly illustrated by the data presented in fig. 8.10 and fig 8.19 in section 8e. These show, for the both cases *ν* = 4 and *ν* = 8, a critical value of *r*_*p*_ much lower than the value *r*_*p*_ = 1 expected in the percolation free case (*ν* → ℞), even when *x*_*v*_ = 0. It is emphasised that this reduction is caused by percolation effects alone, that is, by the increased reduction of the number of susceptibles in the social bubble of an active infection for given *s* arising from the reduction of the size of the social bubbles *ν* and the increased influence of lattice correlations that goes with such a reduction, as well as by cluster-percolation effects discussed in section 8e.

Furthermore, the reduction of the critical *r*_*p*_-value becomes even (much) stronger in vaccinated populations, as shown by the data presented in fig. 8.23 and 8.24, and percolation effects are responsible for this observation. The increased influence of percolation effects in these cases may be inferred from section 8c, where it is shown that an epidemic becomes actually impossible when the percolation threshold of population lattice is passed, even without infection removal. When *p*_*r*_ ≠0, the effects of infection removal and percolation apparently team-up to reduce the critical value of *r*_*p*_ significantly more than the reduction resulting from each of these mechanisms separately (that is, when only one of these mechanisms would apply while the other does not, like in the case of infection removal in a percolation-free case with *ν* → ℞ or the case of percolation effects in the absence of infection removal (*p*_*r*_ = 0).

Finally, it seems that the onset of herd-immunity is in fact related to a 2nd-order phase transition, in which the average cluster-size drops to (almost) zero, whereas the fluctuation size, reflected by 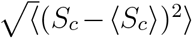, and the correlation length *ξ* diverge. It seems therefore that the herd-immunity transition is accompanied by a genuine percolation transition, and that percolation effects are therewith an integral part of the road towards herd-immunity. They are at least a “side-effect”, but in many cases, especially when the social bubbles are small, also a contributing cause. The entwining of the herd-immunity transition with a percolation transition to the extend that one may even argue that the herd-immunity transition is in fact a percolation transition epitomises the essential role played by percolation phenomena in the achievement of herd-immunity, even in those cases where the primary mechanism driving the transition is infection removal. It may be clear that percolation effects contribute to the achievement of all types of herd-immunity that can be identified on the basis of the classification scheme presented in section 6b.

## 9. Epilogue

### Summary, conclusions and suggestions for further research

The foregoing chapters not only confirm previously known results and phenomena while putting them on more solid mathematical grounds (like the criterion for epidemic spread of an infection), but they also present a number entirely new results and viewpoints.

First of all, the inadequacy of the standard SIR-model, especially under certain conditions (strict lock-downs and the strong limitations of social contacts inherent to them), is revealed. Under very strict lock-down conditions, the failure of the standard SIR model becomes quite dramatic, as demonstrated by the results of simulations presented in chapter 4 and their comparison to predictions by the standard SIR-model for the same sets of parameters as those used in the statistical simulations. As such, the need becomes clear for a new paradigm that allows for either an extension or a correction of the standard SIR-model to address these issues. In Chapter 1 such a change of approach is actually presented by considering the population as a network, and through a particular focus on the parameters ⟨*s*_*si*_⟩ and ⟨*s*_*is*_⟩ and their expression as series expansions in the cumulative infection rate *s* (which itself acts as the analogue of a state parameter in thermodynamic systems). In contrast to the standard SIR-model, this new approach enables an appropriate reproduction of the data obtained from simulations in both a qualitative and quantitative way, even when strict social measures are in place that significantly reduce the number of contacts of each member of the population. However, modelling the spread of infectious diseases through a set of appropriately constructed differential equations is still surrounded by fundamental difficulties. The problem in this respect mainly lies with the influence of the network details on the actual spread of the infection. The complexity of network phenomena is notoriously difficult to describe in terms of an exact algebraic approach. The approach presented in this paper bypasses this issue by the use of series expansions. However, the coefficients in these series expansions cannot be calculated via an ab-initio scheme but their values have to be derived from already existing data (either from simulations or field-data) in such a way that they provide a proper agreement between the solutions of the differential equations and the (actual) data already available from other sources (see chapter 4). Nevertheless, even the insight that the standard SIR-model has serious shortcomings caused by the network structures of populations, and the awareness of the serious fundamental issues that these network structures bring about in relation to the modelling of infection spread in general, has its owns merits. It comes therefore as a bit of a surprise that it can be shown (as in chapter 5) that the criterion for an epidemic to develop from a limited number of initial active infections is actually *independent* of the structure of the population network (which, at first glance, seems to defy intuition).

Understanding the vital role of the network structure on the quantitative aspects of the spread of an infection also leads us to valuable insights regarding the unexpected and undesired effects of changing (scaling down) the regime of social restrictions, even when (or, better stated: especially when) the number of cumulative infections seems to stabilise or saturate (see section 5c). The fade-out of an epidemic is contextual, in the sense that the social-network structure has a direct influence on the total number of cumulative infections reached during a wave of infections that takes place under a specific regime of social restrictions. A change towards a regime of less social restrictions (larger social bubbles) when the cumulative number of infections seems to stabilise while there are still some residual active infections present, may in some cases directly lead to a restart of the epidemic, and to a renewed increase of the number of active and cumulative infections (of potentially dramatic proportions). The relevance of these insights to policy makers is evident. Viewpoints as these are therefore among the key results presented in this paper, as they also provide an illustration of the epistemic difficulties that exist in relation to the concept of herd-immunity and its understanding, and of how an inappropriate conception of herd-immunity may lead to the wrong social policies under the circumstances given.

One of the most urgent scientific issues with regard to herd-immunity is its definition: herd-immunity is still somewhat ill defined. To meet the need for a definition that is not only of practical use but also rooted in the fundamental mechanisms that govern the spread of infections, a classification (based on the results in chapters 4 and 5) is presented in section 6b. A distinction is made between weak and strong herd-immunity of either 1st or 2nd degree. Weak herd-immunity corresponds to a situation as described in the above, where there is a saturation of the cumulative infection rate under a regime of social restrictions, but with the prospect of a new wave of infections once the restrictions are (partially) lifted. In contrast, strong herd-immunity relates to a stable situation where even a return to a situation of social normality will not lead to a new spark in the number of active infections and a new wave of infections. The distinction between 1st and 2nd degree applies to both weak and strong herd-immunity. In a case of 1st degree herd-immunity (weak or strong) the number of active infections is over its peak and in decline: the epidemic is (under the circumstances given) in an inevitable state of fading-out. The complementary case where an epidemic (again under the circumstances given) has actually reached its end (so that the number of active infections is (close to) zero and the cumulative infection rate has reached saturation) is referred to a state of 2nd-degree herd-immunity. It may be clear that the best (i.e. safest) policy, at least from an epidemiological point of view, should aim at achieving strong herd-immunity of (preferably) 2nd degree.

The herd-immunity classification scheme was primarily conceived with a picture in mind of waves of infections that gradually come to a halt (mainly due to infection removal), thus resulting in a state of weak or strong herd-immunity. A less troublesome way to obtain a state of herd-immunity is vaccination. By vaccinating a sufficiently large proportion of the population with a vaccine of sufficiently high effectiveness *ϵ*, states of weak or even strong herd-immunity can be obtained. In a vaccine-induced state of weak herd-immunity, the vaccination campaign has to be complemented with social measures to prevent further spreading of an infection. In vaccine-induced cases of strong herd-immunity, the population is safe against an epidemic even without any social measures in place. It is evident that a vaccination campaign should aim at the latter. Chapter 7 deals extensively with the effects of vaccination. A criterion was obtained for a critical vaccination rate that represents the minimal vaccination rate for which immunity for the population at large (without social measures) against the outbreak of an epidemic is guaranteed (and therewith a state of strong herd-immunity is obtained). The line of thought to obtain such a criterion was similar to the one followed in section 5a and, as a consequence, the critical vaccination rate thus obtained is independent of the structure of the social network. The effectiveness of the vaccine is an important parameter here and it explicitly enters the expression derived for the critical vaccination rate. In relation to this it appeared useful to introduce an effective vaccination rate 𝔰_*vc*_ = ϵ*s*_*v*_. A crucial observation (see fig. 7.1) is that for each *p*_*i*_*/p*_*r*_ a minimum effectiveness and a minimum effective vaccination rate exist below which a state of strong herd-immunity cannot be obtained, so that the pathogen involved has to be considered as endemic: unless vaccines of higher efficiency can be developed, vaccination is unable to provide (strong) herd-immunity and the pathogen will spread through the entire population unless appropriate social measures aimed at reducing *p*_*i*_*/p*_*r*_ are taken. The possibility of such a scenario may also become of relevance when new variants of a pathogen emerge: a vaccine that provides (strong) herd-immunity against one variant may not do so against a newer variant, the result being a new wave of infections.

A link exists between the criterion for an epidemic to evolve from a modest number of initial infections (and correspondingly also the criteria for herd-immunity) and the value of the (basic) reproduction number *R* (*R*_0_). However, the relation between the two is less straightforward than often assumed (see chapter 2 and sections 5d and 6c). Starting from its definition, the reproduction number *R* can be expressed as *R* = *Q · p*_*i*_*/p*_*r*_, where *Q* accounts for the depletion of the reservoir of susceptibles (the basic reproduction number follows as *R*_0_ = *Q*_0_ · *p*_*i*_*/p*_*r*_, where *Q*_0_ is the value of *Q* at the beginning of an outbreak (*t*_0_ = 0)). The criterion *p*_*i*_*/p*_*r*_ > 1 for an epidemic to evolve is therefore not equivalent to *R* > 1 or *R*_0_ > 0 (which as often assumed however). Only when *Q* = 1 (*Q*_0_ = 1) both criteria are equivalent. The issue can only be resolved by changing the definition of the reproduction number (by simply putting *R, R*_0_ *≡ p*_*i*_*/p*_*r*_), or by relating the criterion *p*_*i*_*/p*_*r*_ > 1 to the reproduction number by writing it as *R/Q* > 1 (*R*_0_*/Q*_0_ > 1). The often cited criterion *R, R*_0_ > 1 is therefore to be considered only as a crude approximation obtained under neglect of the depletion of the reservoir of susceptibles during the course of an outbreak (see section 5d).

The role of the network structure is already a central theme in chapters 1 to 7. After all, since the population is considered as a lattice or network, and since it is not difficult to imagine that the structure of such a population network will have at least some influence on the evolution of an epidemic, the effects of the details of the network have to be explicitly considered and accounted for, in order to obtain a realistic analysis. The results in the afore-mentioned chapters clearly demonstrate this. However, there is barely another phenomenon where network effects manifest themselves in such an all-important way as in the percolation transition, the relevance of which to vaccination scenarios is almost self-evident by its very nature. Chapter 8 deals extensively with the phenomenon therefore. Via cluster-identification algorithms applied to data from simulations, a clear link is shown between vaccination-induced percolation transitions and the propagation of an infection through a population. It is shown that the fluctuations in the size of the final clusters of cumulative infections diverges when the percolation threshold is reached. It is also shown that such a divergence in the size-fluctuation can be related *in general* to a divergence in the correlation length (see section 8b). A divergence in the correlation length is a defining signature of a second-order phase transition in physical/thermodynamic systems. As such, the vaccine-induced herd-immunity threshold can be identified as the critical point of such a 2nd-order phase transition. This represents an entirely new fundamental viewpoint on herd-immunity as a phenomenon. With some extensions, the framework outlined in chapter 1 is able to cope with the observed percolation phenomena, including the percolation transition itself. That is, the variation as a function of the vaccination rate of the end-value of the number of cumulative infections, as well as its collapse at the percolation threshold can be adequately described. However, the variation in (average) cluster size and the divergence of the correlation length are beyond the scope of the presented framework. It should be realised however that these are difficult topics that cannot be caught easily (if at all) in a tractable set of formulas and require a numerical approach almost by nature. In section 8e it is shown that the occurrence of percolation phenomena is not restricted to vaccinated populations alone. Including even percolation transitions, they also occur in unvaccinated populations, where they manifest themselves through a collapse of the end-value of the cumulative infection rate at a critical value of *r*_*c*_ = *p*_*r*_*/p*_*i*_ accompanied by a divergence in the size fluctuation of the clusters of cumulative infections, which is indicative of a 2nd-order phase transition also in these cases. Cluster percolation plays a decisive role here (see figs. 8.12 and 8.14 and the related discussion in section 8e). As such we arrive at a new fundamental viewpoint on herd-immunity in general: the herd-immunity threshold marks a 2nd-order phase transition related to an underlying percolation transition, no matter whether vaccinations are involved or not. Also in case of the unvaccinated populations, the modified SIR-model of chapter 1 gives an adequate description of the evolution of an epidemic, even when there is a percolation/herd-immunity transition.

The results in this paper were predominantly obtained under the assumption that both infection removal and vaccination provide full immunity. Only in chapter 7 on vaccination, the possibility of vaccines with an effectiveness *ϵ* less than 1 (i.e. less then 100%) was considered and also part of the analysis. In practice, both infection removal and vaccination may not result in full immunity of each population member involved however, especially when new variants of a pathogen with (slightly) different properties emerge to which the immune system does not have a fully effective response (like in the case of Covid-19 for instance). It is therefore a legitimate question whether the results and conclusions presented in this paper would be different when infection removal and/or vaccination would only result in “partial” immunity, that is when the population members that were vaccinated or the ones that have overcome an infection are not fully immune but may instead be susceptible to re-infection, albeit with a lesser transmission probability than the one that applies to the unvaccinated and also still uninfected (thus fully susceptible) population members. The analyses in the previous chapters offer enough insights to allow for some expectations to be mentioned regarding this issue. For this purpose it is important to notice that although the transmission probability that applies in relation to vaccinated population members and removed infections may be nonzero, a certain “stopping power” is provided by a reduction of the transmission probability: a route of infection along members of the the population may still pass a particular vaccinated node or removed infection, but with much greater difficulty (i.e. much less a probability) than when the node in question was still fully susceptible. Percolation effects are therefore still a very real possibility, and the growth-rate of the number of active infections is (substantially) reduced when the vaccination rate and/or the fraction of removed infections are/is high. The effects of partial immunity are therefore expected to be quantitative rather than qualitative. To corroborate these expectations, an extension of the framework presented in this paper to account also for cases of partial immunity would make an excellent suggestion for further research therewith. The lines along which such an extension may be conceived seem clear and could take the form of the introduction of a second transmission parameter 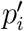 next to *p*_*i*_ (such that 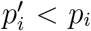) which specifically applies to contacts between an active infection and a vaccinated population member or a removed infection. The spirit of such an approach was actually advanced already in chapter 7 when we dealt with cases where *ϵ* < 1 (eq. (7.2) ff). It is rather straightforward to take this approach one step further and to apply it to cases of partial immunity in general. It should be mentioned however, that those results presented in this paper that were obtained on the basis of the assumption of full vaccine- or removal-acquired immunity represent fair and meaningful approximations for those cases where a substantial (near full) immunity can be obtained.

All together we may conclude that the results in this paper provide quite some new insights in the spread of infectious diseases and the evolution of an epidemic. As stated earlier, it is at these insights that this paper aims particularly, thus leading to new methods to forecast the evolution of ongoing epidemics. In fact, the paper more than once points out that the goal of making such forecasts is quite ambitious and is met with serious fundamental issues. However, the author believes that the incorporation of the methods and viewpoints outlined in the previous chapters could lead to valuable improvements of the models and methods presently in use for modelling the spread of infectious diseases. The subject of the paper is also quite timeless indeed. At the very moment that this text is written, the Covid-19 pandemic seems to be in remission, but it is too early to tell what the near future will bring. In addition, it would be dangerously naive to assume that Covid-19 will be the last of the great pandemics, and that the SARS-CoV-2 family of viruses will be the last pathogens with the potential of causing pandemic outbreaks. Finally, it should also be realised that the viewpoints in this paper equally apply to outbreaks of plant diseases in, for instance, densely packed mono-cultures and may therefore be of relevance in an agricultural context as well.

## Appendix 1

### An eigenvalue equation relating the *s*_*x*_ and the <*s*_*yx*_>

We suppose 3 types of nodes *a, b, c*. Using (1.14) and (1.15):

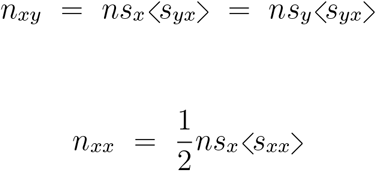

we can rewrite the relation between the numbers of *aa, ab* and *ac* pairs, respectively represented by *n*_*aa*_, *n*_*ab*_ and *n*_*ac*_:

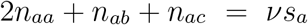

as follows:

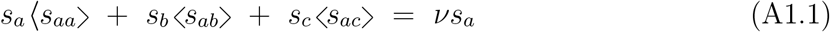

Upon systematic permutation of *a, b, c* in (A1.1) we also obtain:

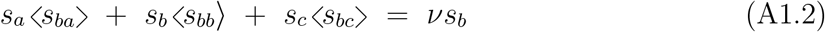

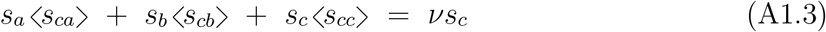

We can write (A1.1), (A1.2) and (A1.3) in matrix form as:

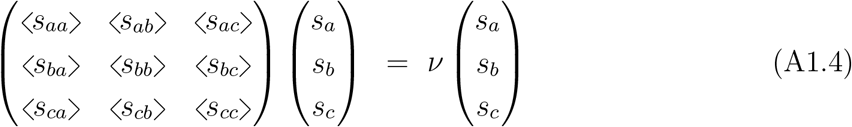

which is essentially an eigenvalue equation with the number of links per node *ν* as the eigen-value. When the number of nodes of a certain type, say *c*, is zero (i.e. *s*_*c*_ = 0 so that ⟨*s*_*xc*_⟩ = ⟨*s*_*cx*_⟩ = 0 for *x* = *a, b, c*) we get the simplified eigenvalue equation:

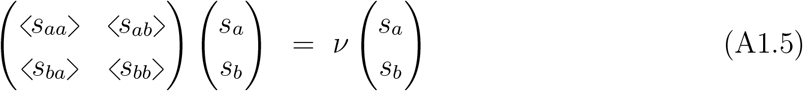

## Appendix 2

### Derivation of the coefficients 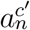

Analogous to (8.24) we write ⟨*s*_*si*_⟩_*c*_′ as:

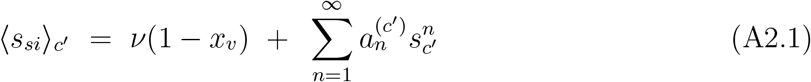

When 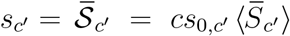 the following condition must then be met:

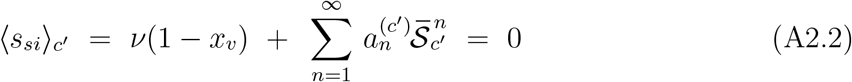

Introducing the function 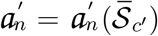 as the analogue of the previously introduced function *a*_*n*_ = *a*_*n*_(*n*_*b*_):

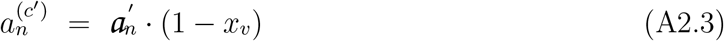

we reexpress (A2.2) as:

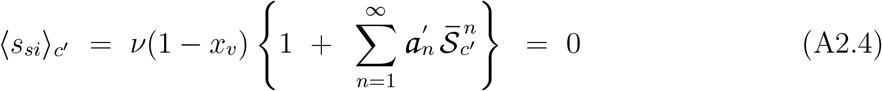

The Laurent series of 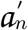 in 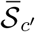 is written as:

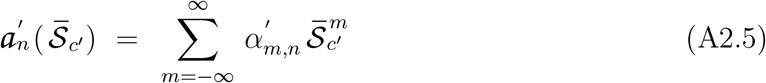

Substitution of (A2.5) into (A2.4) then yields, with *i* = *n* and *j* = *n* + *m*:

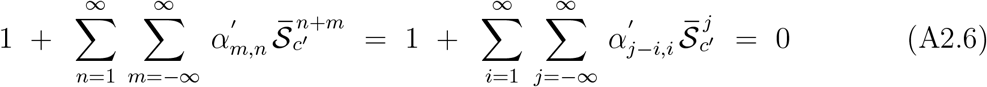

Collecting terms with equal powers of 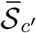 we get, for *j* = *n* + *m* = 0:

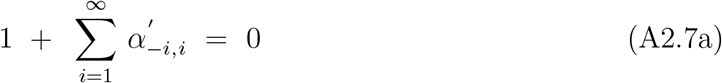

and for *j* = *n* + *m /*= 0:

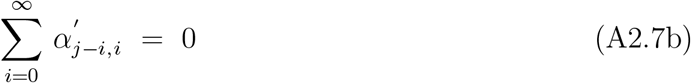

Neglecting terms of 3rd and higher order in 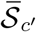 in the series expansion of ⟨*s*_*si*_⟩_*c*_′, only 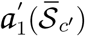 and 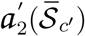 remain relevant. For *n* = 1 and *n* = 2 we get from (A2.5):

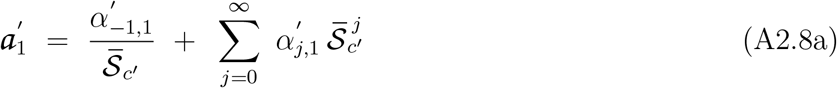

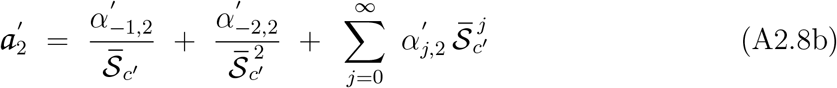

Analogous to (8.32), for the 2nd-order polynomial approximations of ⟨*s*_*si*_⟩_*c*_′, the following constraint must apply with 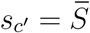 (see (A2.2)):

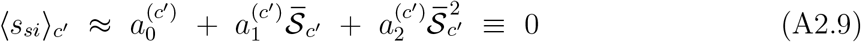

so that in combination with (A2.3) we get, for *x*_*v*_ ≠ 1:

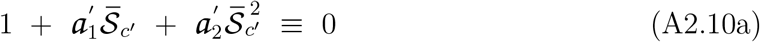

That is:

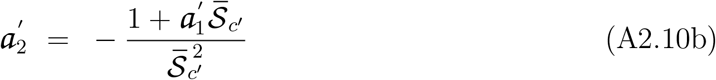

Substitution of (A2.8a) then yields:

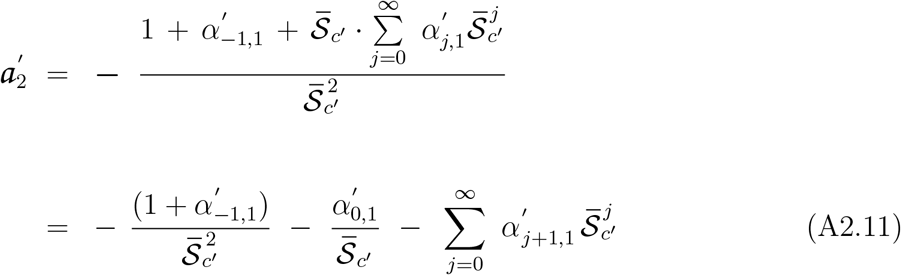

from which 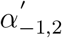 and 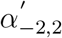 can be identified as:

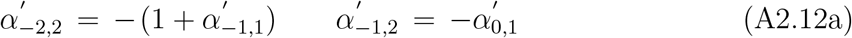

respectively in agreement with (A2.7a) and (A2.7b). For the 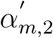 when *m* ≥ 0 we get from comparison of (A2.11) and (A2.8b):

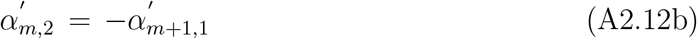

By combining (A2.4), (A2.8a) and (A2.11), the Laurent series in 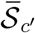 for 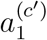 and 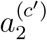 can thus be written as:

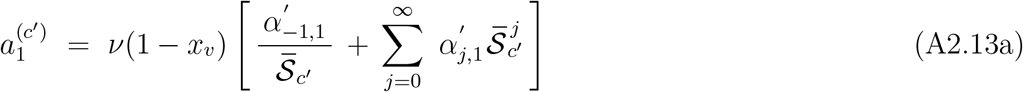

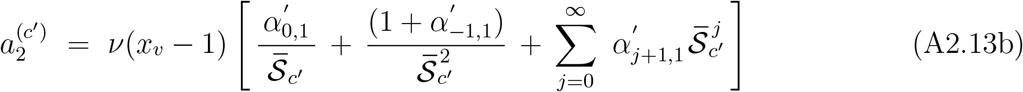

## Footnotes

1 A variation on the famous sentence by Richard Hamming: “The purpose of scientific computing is insight, not numbers”.

2 Taking an arbitrary moment in time instead of *t* = 0 as a reference for the calculation of the constant of integration will be of use in chapter 7.

3 This phenomenon will be addressed extensively in Chapter 6

4 The 2nd derivative ∂^2^*g*(*s*)/∂*s*^2^ is given by: 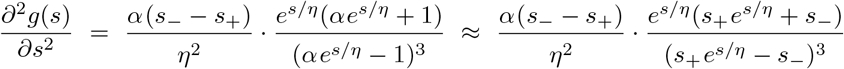 where the approximation on the right applies when *s*_0_ << 1 or, more in particular, when *s*_0_ << |*s*_+_|, |*s*_−_| (which is basically the regime we focus on). Provided that *s*_+_ ≠ *s*_−_, the numerator of the 2nd term of the approximation on the right is positive or negative definite when *s*_+_, *s*_−_ < 0 or *s*_+_, *s*_−_ > 0. When *s*_+_ and *s*_−_ have different sign (i.e. when (5.13a) applies), the numerator may change sign only for some *s* < 0 (since *η* ≤ 0), whereas the denominator does not change sign for any *s* ∈ ℝ. Therefore, *if* ∂^2^*g*(*s*)/∂*s*^2^ changes sign in the cases of our interest, it can only be when *s*_+_ and *s*_−_ have different sign, and only for some *s* < 0.

5 Be aware that when 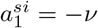 and 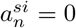 for *n* > 1, eqs. (1.42a) and (1.42b) respectively reduce to eqs. (1.6) and (1.7) of the standard SIR-model

6 The equation for *τ* in this case is: 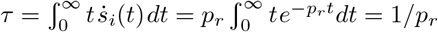

7 Note that *n*_*e*_*/n*_*s*_ relates to the final cumulative infection-rate *s*_*e*_ = *n*_*e*_*/n* and the vaccination rate *x*_*v*_ via *n*_*e*_*/n*_*s*_ = *s*_*e*_*/s*_*s*_ = *s*_*e*_/(1− *x*_*v*_)

8 (i.e. the number of initial infections in a cluster)

9 systematic numerical evaluation is possible

10 Note that 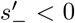 for *p*_*r*_*/p*_*i*_ > 1, consistent with the results in chapter 5 demonstrating the impossibility of an epidemic when *p*_*r*_*/p*_*i*_ > 1.

11 *n* being the total number of nodes/individuals in the population lattice

